# A shared MHC immunogenetic signal connects aging, rheumatoid arthritis, and herpes zoster through chronic high inflammatory burden–compensatory immune tolerance dysregulation

**DOI:** 10.64898/2026.04.28.26351835

**Authors:** Xin Yao, Manli Wan

**Author notes:** Correspondence: Xin Yao, MD, Department of Rehabilitation, Affiliated Yueqing Hospital of Wenzhou Medical University, No. 338, Qingyuan Road, Yueqing City, Zhejiang Province, China 325600.

## Abstract

Epidemiological studies connect aging and autoimmune diseases to increased herpes zoster (HZ) risk, yet their shared genetic basis remains unresolved. Here, we integrated large-scale genome-wide association studies (GWAS) of a multivariate aging latent factor (mvAge), rheumatoid arthritis (RA, representing autoimmunity), and HZ with multi-omics quantitative trait loci. Using linkage disequilibrium-aware colocalization and Mendelian randomization (MR), we identified a shared pleiotropic major histocompatibility complex (MHC) signal, tagged by *rs1800628*. Phenome-wide association studies (PheWAS) and network analyses indicated that the signal variation is associated with systemic immune remodeling, characterized by increased pro-inflammatory mediators, elevated T-cell regulation markers, and reduced lymphocyte counts. This pleiotropic genetic variation may alter the lifelong immune regulatory trajectory, accelerating aging and predisposing individuals to both autoimmunity and VZV reactivation. Based on these findings, we propose a life-course “chronic high inflammatory burden–compensatory immune tolerance dysregulation” conceptual model, providing a genetic basis for the epidemiological overlap of aging, autoimmunity, and HZ, and a hypothesis-generating framework for understanding and untangling systemic immune dysregulation prior to clinical disease onset.

---

Driven by a rapidly aging global population, the clinical burden of herpes zoster (HZ) is increasingly concentrated among the elderly and individuals with autoimmune diseases, particularly rheumatoid arthritis (RA)^1,2^. This susceptibility persists independently of immunosuppressive therapies, indicating intrinsic immune dysregulation^2^. This clinical overlap is underpinned by two interrelated phenomena: immunosenescence—the functional decline of cellular immunity—and “inflammaging”, a state of chronic, subclinical systemic inflammation^3–5^. Together, they induce a paradoxical immunological state that renders individuals vulnerable to both latent viral reactivation and autoimmunity^6^. Contemporary immunology indicates that immune regulatory trajectories are influenced by early-life immune development and tolerance mechanisms, and that baseline immune states can be conceptualized as an intrinsic “immunological set-point“^7,8^. Indeed, recent investigations identified pleiotropic MHC variants that modulate diverse autoimmune diseases and HZ susceptibility^9,10^. However, whether such immunogenetic signals extend beyond autoimmunity to influence aging-related immune remodeling and *varicella-zoster virus* (VZV) reactivation susceptibility remains elusive.

Disentangling these genetic signals is challenging, as the extensive linkage disequilibrium (LD) and complex haplotype structures of the MHC confound traditional fine-mapping methods^11^. To address these analytical challenges, we conducted a systems-level investigation (Fig. 1). Using RA as our primary discovery model for autoimmunity, we utilized large-scale genome-wide association study (GWAS) summary statistics of a multivariate aging genetic factor (mvAge, reflecting healthy aging), RA, and HZ^12–14^, by integrating bulk and cell-resolved single-cell expression quantitative trait loci (sc-eQTLs)^15^. We implemented a custom analytical pipeline integrating Sum of Single Effects (SuSiE)-based colocalization^16,17^ with a study-specific LD-aware signal deconvolution strategy. This approach accommodates multiple causal variants and facilitates the mapping of shared genetic associations within the complex MHC locus.

**Fig. 1.**
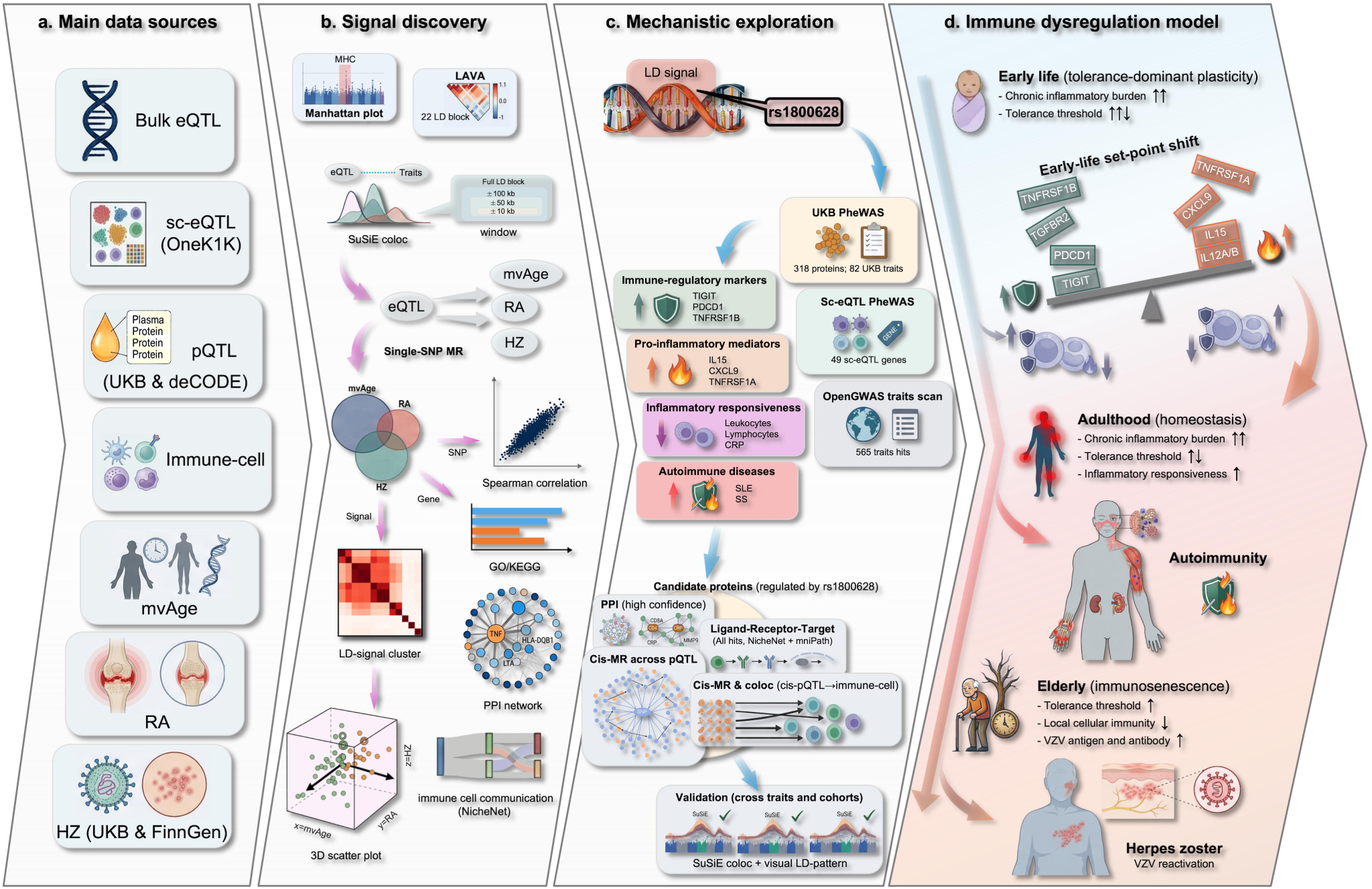
Study design and overall analytical framework. **a, Main data sources.** This panel summarizes the integration of large-scale GWAS summary statistics for a multivariate aging latent factor (mvAge), rheumatoid arthritis (RA), and herpes zoster (HZ), together with multi-omics quantitative trait locus datasets, including single-cell eQTL (sc-eQTL) and plasma protein QTL (pQTL) resources. **b, Signal discovery.** Assessment of shared local genetic architecture across the major histocompatibility complex (MHC) region using Local Analysis of [co]Variant Associations (LAVA). Identification of shared pleiotropic signals via a colocalization-constrained Mendelian randomization (MR) framework and a linkage disequilibrium (LD)-aware signal clustering strategy. A cross-trait alignment framework was used to evaluate regulatory concordance, supplemented by cross-trait functional enrichment, protein–protein interaction (PPI) networks, and NicheNet intercellular communication analyses to characterize shared biological pathways. **c, Mechanistic exploration.** Phenome-wide association study (PheWAS) centered on the representative shared MHC signal (tagged by *rs1800628*). Downstream analyses of PheWAS-associated plasma proteins via PPI and NicheNet frameworks resolved functional modules of immune regulation. This was further followed by cross-cohort *cis*-MR analyses to infer the causal hierarchy among these proteins, and subsequent *cis*-MR against extensive immune cell phenotypes to infer the downstream causal cascade. Additionally, cross-phenotype and cross-cohort Bayesian colocalization analyses were conducted to verify the pleiotropy of this shared immunogenetic signal set. **d, Immune dysregulation model.** The proposed life-course “chronic high inflammatory burden–compensatory immune tolerance dysregulation” model. This framework illustrates how the shared immunogenetic signal is associated with an early-life innate hyper-responsive state and a compensatory upregulation of adaptive immune tolerance, establishing an altered immunological set-point that co-regulates the aging trajectory, autoimmunity risk and HZ susceptibility.

Through these cross-trait analyses, we identified an MHC-resident signal (tagged by *rs1800628*) associated with widespread immune reprogramming. Subsequently, a phenome-wide association study (PheWAS) revealed that the signal is associated with an increase in pro-inflammatory mediators and decoy receptors (e.g., IL15, IL2RA), and a compensatory upregulation of T-cell regulation markers (e.g., TIGIT). By leveraging independent proteomic cohorts (UK Biobank and deCODE)^18,19^ and immune cell traits from SardiNIA study^20^, we inferred the causal hierarchy of these immune alterations. To conceptualize these molecular shifts, we propose a life-course “chronic high inflammatory burden–compensatory immune tolerance dysregulation” model (Fig. 1d) linking aging, autoimmune risk, and VZV reactivation. Our findings indicate that autoimmunity and VZV reactivation are linked outcomes along a lifelong immune regulatory trajectory, modulated by this shared MHC signal variation and the associated immune remodeling, providing a conceptual framework for understanding systemic immune dysregulation.

## Results

### Signal discovery

Genome-wide Manhattan plots for mvAge, RA, and HZ GWAS datasets highlighted prominent association peaks in the chromosome 6 MHC region (Fig. 2a)^11^. Local genetic correlation analysis using Local Analysis of [co]Variant Associations (LAVA)^21^ across the extended MHC region revealed spatially heterogeneous patterns of shared genetic architecture among mvAge, RA, and HZ. The distinct LD-defined sub-blocks exhibited divergent correlation directions and magnitudes. Multiple MHC segments demonstrated consistent negative local genetic correlations between mvAge and RA, and positive correlations between independent HZ GWAS datasets from different European populations (i.e., UK Biobank and FinnGen). In contrast, the local genetic correlations between mvAge and HZ, as well as between RA and HZ, were heterogeneous across different MHC segments. Furthermore, blocks showing correlations in the same direction tended to cluster spatially, suggesting non-random regional aggregation of local genetic effects within the MHC (Fig. 2b and Supplementary Table 3).

**Fig. 2.**
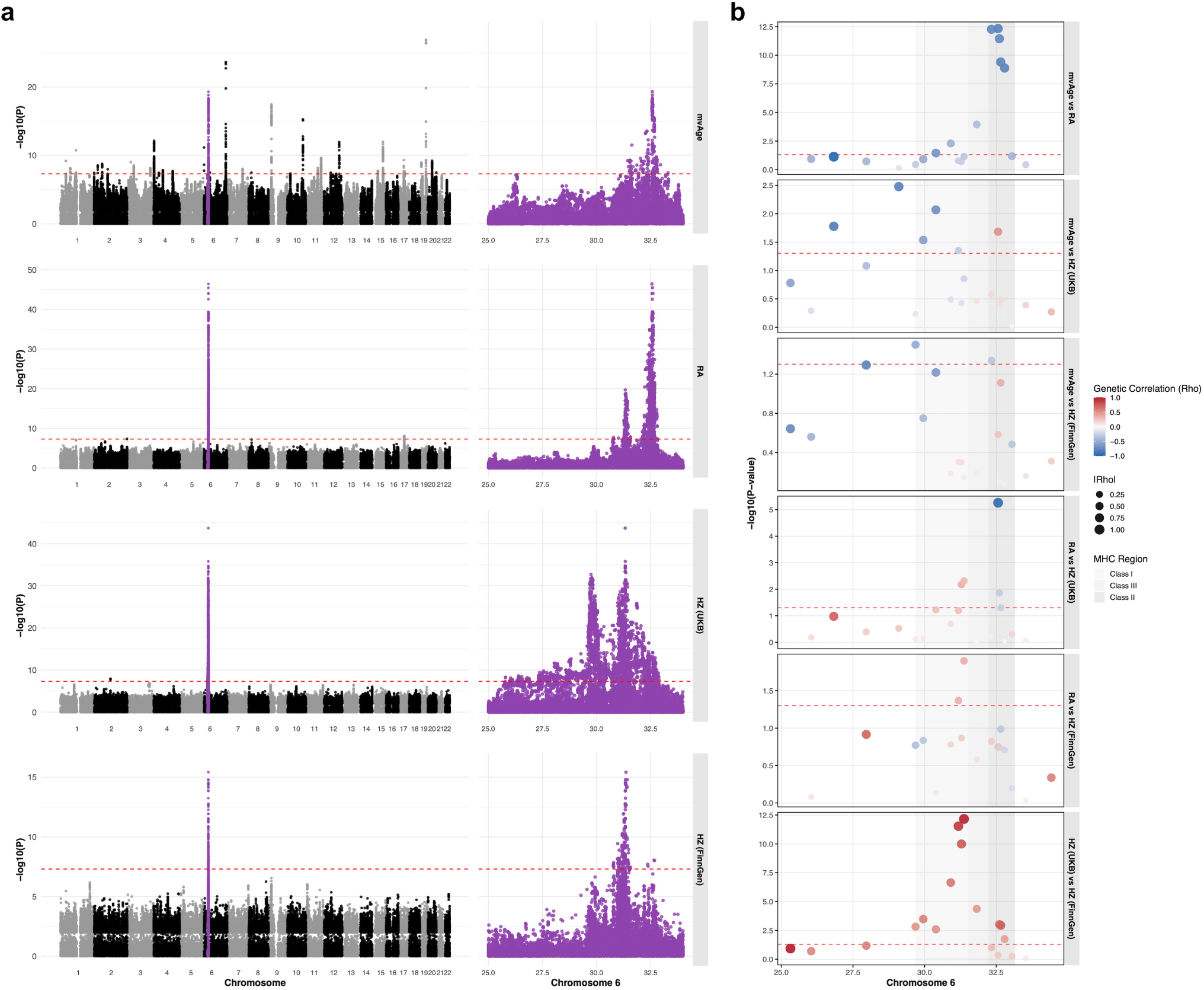
Regional-level discovery identifies the MHC as a shared genetic hotspot for mvAge, RA, and HZ. **a, Genome-wide Manhattan plots.** Association signals for mvAge (N = 1,958,774), RA (N_cases_ = 25,533, N_controls_ = 290,135), and HZ (UK Biobank, N_cases_ = 17,856, N_controls_ = 440,584; FinnGen, N_cases_ = 7,132, N_controls_ =480,316) are displayed. The y-axis represents the −log10(*P*) derived from two-sided association tests in the source GWAS summary statistics. The dashed line indicates the genome-wide significance threshold (*P* < 5 × 10^−8^). The chromosome 6 MHC region highlights prominent and shared association peaks across all three traits. **b, Local genetic correlation across the extended MHC region.** Bivariate local genetic correlations were estimated using LAVA, with P values derived from two-sided statistical tests evaluating whether the local genetic correlation significantly differs from zero. The region was partitioned into 22 LD-defined sub-blocks, revealing spatially heterogeneous and clustered patterns of shared genetic architecture among mvAge, RA, and HZ.

Using a colocalization-constrained Mendelian randomization (MR) framework, we identified 413 genes significantly associated with mvAge, 135 with RA, and 208 with UK Biobank (UKB) HZ genome-wide (false discovery rate [FDR] < 0.05; Fig. 3a, Supplementary Fig. 2 and Supplementary Tables 5–10). At the gene level, 58 mvAge–HZ and 54 RA–HZ overlaps were observed, with the majority localized to the MHC region—underscoring its prominent genetic contribution across these traits. Applying a strict shared-variant criterion (identical gene, immune cell type, and single nucleotide polymorphism [SNP]) yielded a set of consistent cell–gene–SNP triplets across pairwise trait comparisons (ranging from 36 to 51 shared triplets). We identified a core set of 10 triplets (comprising 6 unique genes and 8 variants) that were simultaneously shared across all three traits (Fig. 3b).

**Fig. 3.**
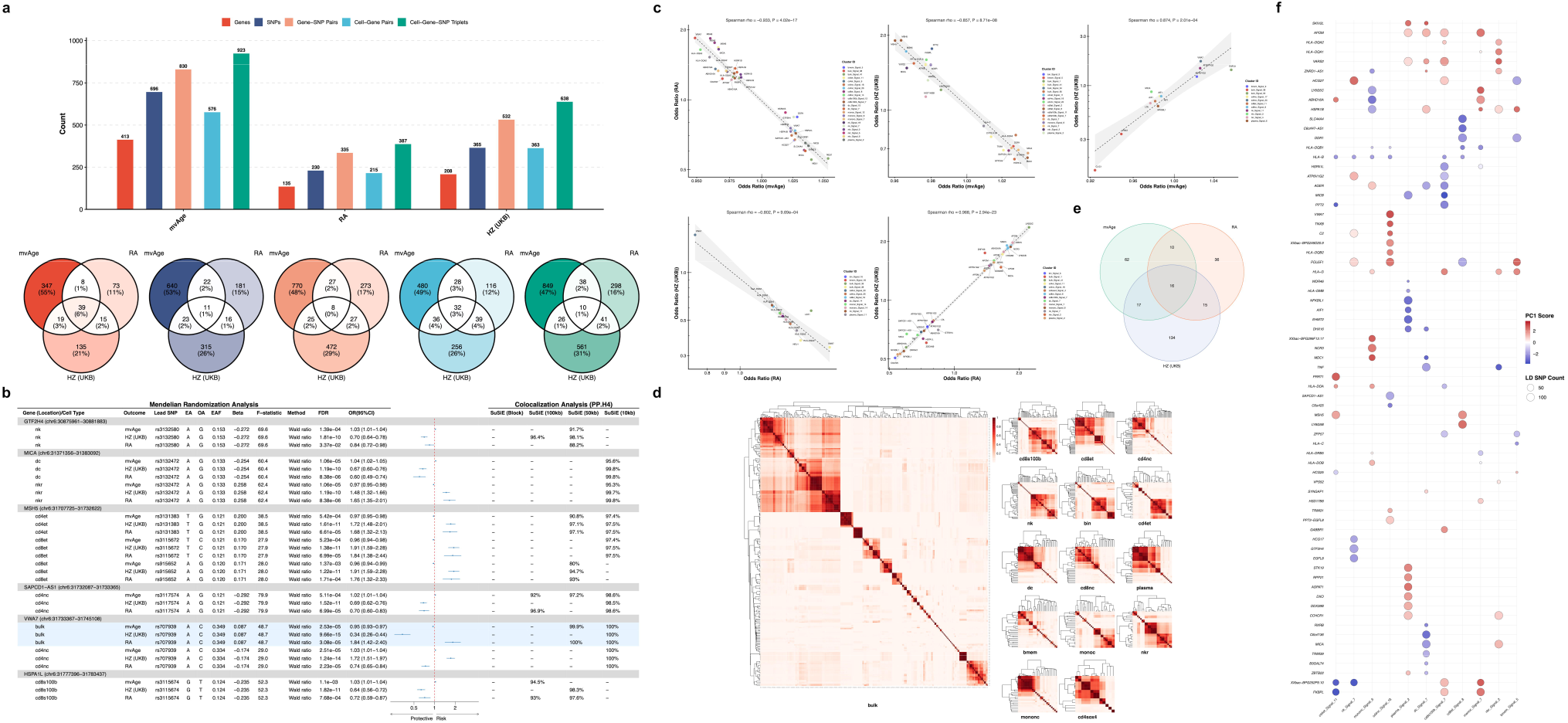
Signal-level discovery and cross-trait alignment of pleiotropic regulatory architecture. **a, Genome-wide gene overlap.** Venn diagram illustrating the overlap of genes significantly associated with mvAge, RA, and HZ, identified via a colocalization-constrained MR framework. The majority of shared genes localize to the MHC region. **b, Core shared cell–gene–SNP triplets.** Forest plots displaying the MR-derived odds ratios (ORs) for the 10 strictly shared triplets across all three traits. Center measures show ORs estimated using the Wald ratio MR method, and error bars represent 95% confidence intervals (CIs). With the exception of one discordant variant (*rs707939*), the remaining signals exhibit consistent cross-trait directional architecture. **c, Cross-trait regulatory concordance.** Spearman correlation analyses comparing the directions and magnitudes of MR effect estimates across pairwise trait comparisons. *P* values are derived from two-sided Spearman correlation tests. **d, e, LD-aware signal reconstitution.** Reconstitution of fragmented colocalized triplets into independent exposure signals using an LD-aware framework anchored by sc-eQTLs. This approach delineates local regulatory neighborhoods and groups exposure-side associations into distinct LD signal sets to provide a conservative representation of the shared genetic architecture. **f, Three-dimensional projection of cross-trait effects.** Projection of the LD signal-level variant sets onto a three-dimensional effect-size space and a fitted global principal component (PC) axis.

To evaluate cross-trait regulatory concordance, we compared the directions and magnitudes of MR-derived odds ratios across traits using Spearman correlation analysis. The 48 mvAge–RA triplets showed a consistent, strong negative correlation of effect estimates, while mvAge–HZ (36 triplets) and RA–HZ (51 triplets) displayed greater directional heterogeneity. When pooled, shared signals produced attenuated correlations owing to opposing directions of effect; stratification by effect direction, however, revealed strong within-subset concordance. These cross-trait correlation patterns were consistent with the results obtained from LAVA (Fig. 3c).

In the 10 triplets shared across mvAge, RA, and HZ, one variant (*rs707939*) displayed an isolated discordant effect pattern relative to the remaining signals (Fig. 3b). After excluding this outlier, the remaining triplets exhibited a consistent cross-trait architecture. Specifically, MR-derived effect estimates demonstrated a strong positive correlation between RA and HZ, indicating genetically predicted concordant effect directions across RA susceptibility and VZV reactivation susceptibility. In contrast, when healthy aging was considered, effect estimates showed strong inverse correlations with both RA and HZ.

To further characterize regulatory pleiotropy within the extended MHC region, we queried OneK1K sc-eQTL summary statistics^15^ for each prioritized cell–SNP pair and extracted all genes for which the SNP served as a cell-type-specific and significant eQTL (FDR < 0.05). This cell–SNP–anchored expansion captured local regulatory neighborhoods whereby a single variant modulates multiple adjacent genes, thereby delineating pleiotropic targets beyond the originally colocalized gene (Supplementary Fig. 3 and Supplementary Table 15).

To provide a conservative representation of shared genetic architecture amidst complex MHC LD, we applied an LD-aware framework to reconstitute fragmented colocalized triplets into independent exposure signals (Fig. 3d and e). This approach grouped the exposure-side associations into 290 LD signal sets (76 bulk-eQTL–derived and 214 cell-specific; Supplementary Table 16).

To focus on cell-resolved regulatory units, we excluded bulk-derived signal sets and an isolated CD4 naïve/central-memory eQTL cluster. This stringent filtering left 11 cell-specific eQTL signals that were shared across mvAge, RA, and HZ at the LD-signal level (Supplementary Table 16).

By projecting these LD signal-level variant sets onto a three-dimensional effect-size space (see Supplementary File 1), we visualized the harmonized SNP-level MR effects across all three traits. The projection of representative SNPs onto a fitted global principal component (PC) axis quantitatively supported a consistent cross-trait regulatory architecture, highlighting the putative contribution of each signal–gene unit to the shared pathogenic trajectory (Fig. 3f and Supplementary Table 17).

### Cross-trait functional and network analyses

Functional enrichment analyses (GO and KEGG) of the gene sets prioritized by colocalization-constrained MR (mvAge, n = 413; RA, n = 135; HZ, n = 208; Supplementary Table 18) revealed substantial cross-trait overlap (Supplementary Fig. 4, and Supplementary Tables 19 and 20). These shared functional signatures were dominated by antigen processing and presentation—specifically MHC class II–related processes and MHC protein complexes—as well as pro-inflammatory and antiviral signaling cascades, such as tumor necrosis factor (TNF), nuclear factor kappa B (NF-κB), and interferon pathways. Pathways governing leukocyte activation, adhesion, and cytokine–cytokine receptor interactions were also recurrently enriched, suggesting that the genetic modules underlying these three traits converge on coordinated antigen-presentation and cytokine-mediated immune circuits.

The protein–protein interaction (PPI) network analyses revealed that the three-way union network (comprising 636 proteins) integrated the corresponding protein products of all prioritized genes across mvAge, RA, and HZ^22^. The three-way shared core comprised 39 central proteins, which included multiple MHC/immune molecules and crucial inflammatory mediators (e.g., TNF, HLA-DQA1/2) that served as connector nodes mediating cross-trait molecular links (Supplementary Fig. 5, Supplementary Table 21).

Intercellular communication analyses via the genetics-anchored NicheNet framework further delineated these putative immune circuits^23^. The upstream analysis converged on a reproducible set of inflammatory and immune-interaction ligands, while the downstream analysis highlighted a compact interferon/NF-κB–centered transcriptional response module. Among the top 30 ranked results across all three traits, we identified a three-way intersection of 14 upstream ligands and 16 downstream targets (Supplementary Fig. 6, and Supplementary Tables 22 and 23).

### *Cis*-MR based on UKB pQTL

After FDR correction, *cis*-MR identified 223 proteins significantly associated with RA (Fig. 4a) and 153 significantly associated with HZ (FinnGen, Fig. 4b and Supplementary Table 25), with 22 proteins shared across both traits. Several shared signals showed concordant directions of effect across both diseases, including risk-associated proteins (e.g., LYVE1, NFU1), and protective proteins (e.g., HCG22, TNFRSF11A). By contrast, a subset of proteins (e.g., ACVRL1, ADAMTSL4) showed opposite directions of effect (Fig. 4c).

**Fig. 4.**
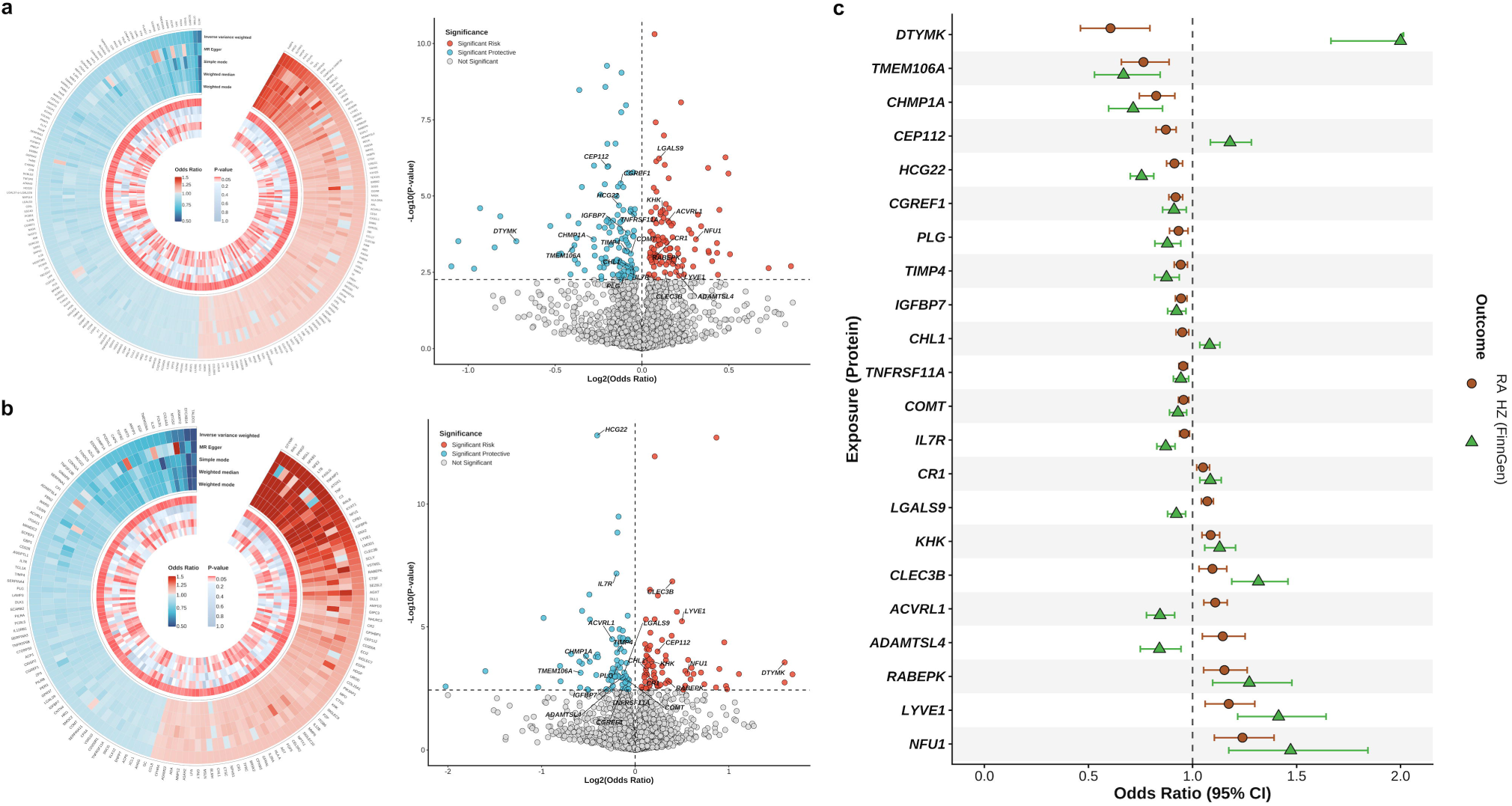
Identification of putative causal plasma proteins for RA and HZ via *cis*-MR. **a, b, Genetically predicted causal associations for RA and HZ.** Circular plots and volcano plots displaying the UK Biobank (UKB) plasma proteins (N = 47,745) significantly associated with RA (**a**, n = 223) and HZ in the FinnGen cohort (**b**, n = 153) following false discovery rate correction (FDR < 0.05). *P* values are derived from the inverse-variance weighted (IVW) and Wald ratio MR methods. **c, Cross-trait comparison of shared plasma proteins.** Forest plot comparing the MR-derived effect directions for the 22 plasma proteins shared between RA and HZ. Center measures show ORs from the IVW and Wald ratio MR methods, and error bars represent 95% CIs. The plot highlights proteins with concordant directions of effect across both diseases (e.g., LYVE1, NFU1 as risk-associated; HCG22, TNFRSF11A as protective) alongside a subset exhibiting opposing associations (e.g., ACVRL1, ADAMTSL4).

### PheWAS and downstream analyses

The PheWAS results (*P* < 5 × 10^−8^) for the representative variant *rs1800628* (G>A) of the shared MHC signal showed that this variant was significantly associated with 321 UKB plasma proteins (Fig. 5a and Supplementary Fig. 7a, and Supplementary Table 26), 82 UKB traits (Fig. 5b and Supplementary Table 27), and 49 sc-eQTL genes (Supplementary Fig. 7b and Supplementary Table 28). Subsequent screening across the OpenGWAS database^24^ identified 565 associated phenotypes, broadening the phenotypic spectrum (Supplementary Fig. 7c and d, and Supplementary Table 29). The associated traits included decreased counts of various blood cells (e.g., leukocytes, lymphocytes, monocytes) and alterations in the levels of plasma proteins. We observed the elevation of T-cell tolerance and exhaustion markers (e.g., TIGIT, PDCD1)^25^ alongside increased pro-inflammatory mediators (e.g., IL15, CXCL9)^26,27^ and the soluble decoy receptor IL2RA^28,29^. Furthermore, this variant was positively associated with multiple autoimmune diseases, including systemic lupus erythematosus (SLE), sicca syndrome (SS), and dermatopolymyositis (DM), as well as with elevated VZV glycoprotein E and I antibody levels^30^.

**Fig. 5.**
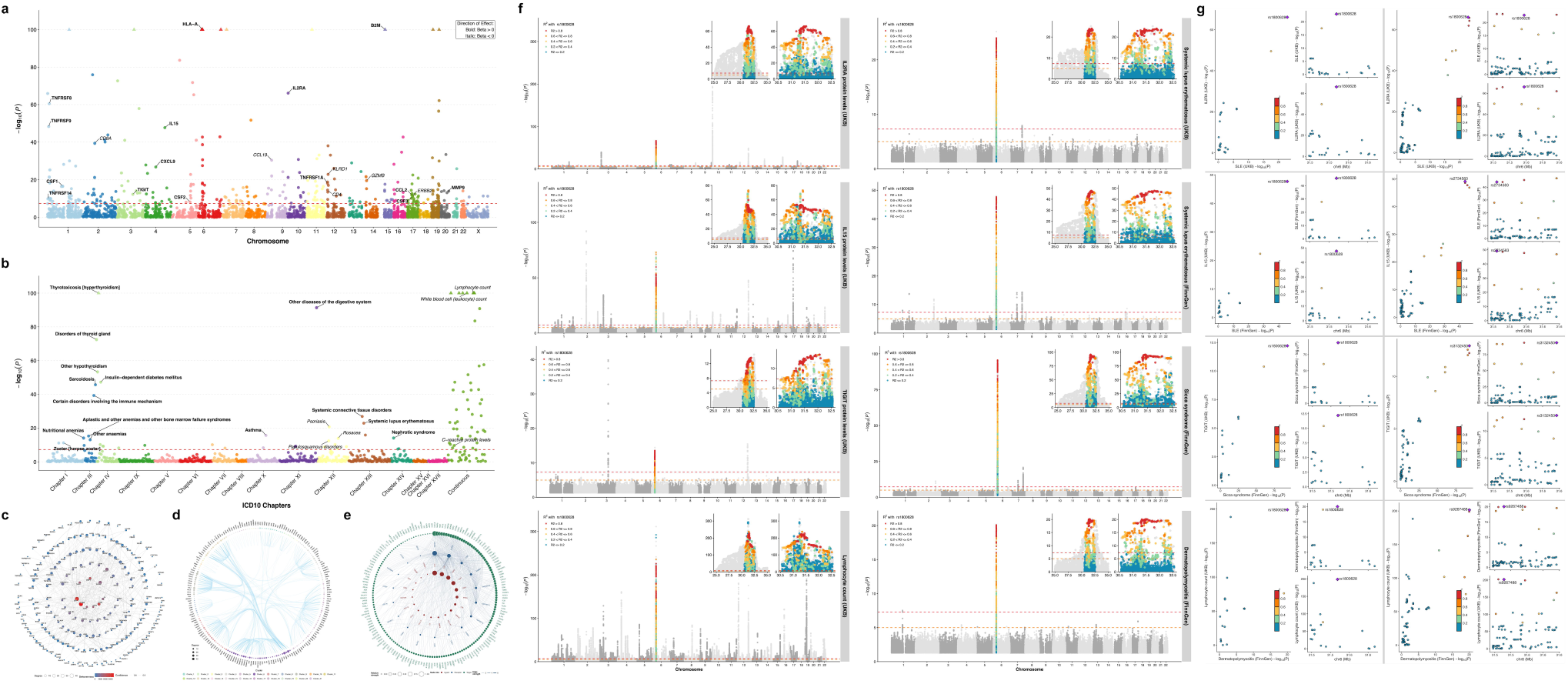
Phenome-wide characterization and cross-trait colocalization of the pleiotropic MHC signal tagged by rs1800628. **a, b, PheWAS screening.** Manhattan plots showing the significant associations of the *rs1800628* (G>A) variant with 321 UKB plasma proteins (**a**) and 82 UKB traits (**b**). The y-axis represents the −log10(*P*) derived from two-sided association tests in the source PheWAS/GWAS summary statistics. The dashed line indicates the genome-wide significance threshold (*P* < 5 × 10^−8^). The variant is linked to widespread immune perturbations, including elevated T-cell tolerance/exhaustion markers (e.g., TIGIT) and increased pro-inflammatory mediators (e.g., IL15, CXCL9). **c, d, Protein–protein interaction (PPI) network and hub topology.** Topological network diagram (**c**) and modularity clustering (**d**) of the 321 variant-associated plasma proteins. The network resolves distinct immune-related clusters, highlighting CD4, CD8A, MMP9, and C-reactive protein (CRP) as key central hubs that potentially delineate the attenuation of T-cell and acute-phase responses, alongside the activation of chronic inflammatory pathways (e.g., IL15, CXCL9). **e, Ligand–receptor intercellular communication.** NicheNet-based analysis delineating the intercellular signaling cascades among the signal-set-associated proteins, resolving complete ligand–receptor–target chains. **f, g, Cross-trait SuSiE-based colocalization and LD alignment.** Visual LD-pattern alignment (**f**) and Bayesian colocalization visualizations (**g**) across multiple UKB and FinnGen traits at the *rs1800628* locus. Panel **g** displays eight colocalization plots arranged in a 2×4 layout, contrasting the local genomic architectures within ±10 kb (left) and ±50 kb (right) windows. The posterior probability of colocalization (PP.H4) was calculated using the Sum of Single Effects (SuSiE) model. The high degree of spatial consistency among variants in strong LD provides strong evidence for a shared immunogenetic signal set mediating extensive pleiotropy across immune and autoimmune phenotypes.

To explore the functional connectivity of the 321 UKB candidate plasma proteins associated with the variant (*P* < 5 × 10^−8^), we conducted PPI network analysis. This analysis identified 23 hub proteins, such as CD4, IL15, C-reactive protein (CRP), and TIGIT, and revealed distinct functional clusters (Fig. 5c and d, and Supplementary Table 31). These specific protein alterations—suppressed CD4 and CRP levels alongside elevated IL15 and TIGIT—point towards a molecular signature characterized by the concurrent attenuation of baseline T-cell and acute-phase responses, coupled with the activation of chronic inflammatory and immune exhaustion networks^25,26^. A parallel ligand–receptor interaction analysis via NicheNet on these 321 proteins predicted 507 complete ligand–receptor–target chains, involving 19 ligands, 18 receptors, and 164 target genes (Fig. 5e, and Supplementary Table 32). To further reduce false positives and focus on robust *trans*-pQTL effects, we subsequently applied a more stringent association threshold of *P* < 1 × 10^−11^, retaining 195 UKB candidate plasma proteins. Repeated PPI and NicheNet analyses on this refined set similarly supported functional modules of immune regulation and pro-inflammatory mediators (Supplementary Fig. 8, and Supplementary Tables 33 and 34).

Although knowledge-based networks (STRING and NicheNet) highlighted functional connectivity among candidate proteins, they cannot resolve causal directionality. To infer the causal hierarchy underlying this immune remodeling, we performed a cross-cohort *cis*-MR analysis among the candidate proteins themselves. Among these 195 UKB candidates, 176 retained valid *cis*-pQTL instruments and were utilized for downstream causal inferences. The *cis*-MR analysis between the UKB cohort and the independent deCODE cohort^19^ identified 3,342 significant directed causal relationships (FDR < 0.05, Supplementary Fig. 9, Supplementary Table 35, and Supplementary File 2) across 28,710 non-homologous cross-protein evaluations. This cross-cohort causal mapping highlighted upstream regulators and downstream effectors within the immunoproteomic signature, providing a genetically inferred basis for the hierarchical propagation of the *rs1800628*-associated immune response.

Subsequently, we evaluated the impact of 176 candidate plasma proteins on the systemic immune cell landscape via *cis*-MR analyses against 731 immune cell GWAS traits^20^. We identified 7,966 significant causal associations, spanning 168 plasma proteins and 731 immune cell traits (Supplementary Table 36), demonstrating that genetically predicted alterations in *rs1800628*-associated plasma protein levels contribute to widespread disturbances to the immune system (e.g., elevated TIGIT and PDCD1 levels are linked to an increase in both the relative abundance and absolute counts of various regulatory T cell subsets; notably, the causal effects of TIGIT on specific subsets, such as CD25 on resting CD4 regulatory T cells, are further substantiated by strong genetic colocalization; Supplementary Fig. 10 and Supplementary Table 36).

For further validation of this cross-trait genetic pleiotropy, we performed local SuSiE-based colocalization analyses across eight traits of interest from UKB and FinnGen databases. We observed a high degree of spatial consistency in the association signals by characterizing the distribution of variants in strong LD with *rs1800628* across wider genomic windows. This complementary strategy—combining Bayesian colocalization with visual LD-pattern alignment—provided genetic evidence for this shared signal across traits (Fig. 5f and g).

### Sensitivity and robustness analyses

Effect sizes across 188 significant cell–gene–SNP triplets shared between mvAge and frailty index (FI) were inversely correlated (Spearman ρ = −0.988, *P* = 5.9 × 10^−152^; Supplementary Table 37), consistent with internal coherence. External validation using the independent FinnGen HZ GWAS further reinforced the robustness of our findings, demonstrating concordant effect directions for 88.9% (32 of 36) of mvAge–HZ shared triplets and 98.0% (50 of 51) of RA–HZ shared triplets relative to the primary UKB cohort (Supplementary Table 38).

Transcriptional analyses indicated that acute HZ infection did not broadly confound signals at GWAS-implicated loci. Of the 208 HZ risk genes identified by our eQTL-driven framework, 117 were quantifiable in peripheral blood bulk RNA-seq data^10^. Following FDR correction, no widespread transcriptional differences emerged between acute HZ cases and healthy controls. Only a single gene, *IGHGP*, showed significant upregulation during the acute phase, which was no longer detectable in convalescent samples (Supplementary Table 39). No significant differences in GWAS-implicated genes were observed between convalescent individuals and controls, supporting that the identified genetic associations primarily reflect baseline immunogenetic architecture rather than secondary effects of acute immune activation.

We evaluated whether the *rs1800628*-associated alterations in plasma protein levels identified in the UKB cohort were reproducible in the independent deCODE Icelandic cohort (Supplementary Table 30). Following strict allele alignment, 1,772 plasma proteins were successfully mapped between the two cohorts. Of these, 71.28% exhibited concordant effect directions. Stratification by significance thresholds demonstrated a positive relationship between association strength and directional concordance. Within the subset of 149 matched proteins that were highly significant in the primary UKB cohort (*P* < 1 × 10^−11^), the directional concordance rate was 81.88% (122/149). Further restriction to a core module of 13 proteins reaching high significance (*P* < 1 × 10^−11^) in both cohorts yielded a directional concordance rate of 92.31% (12/13, e.g., C2, Supplementary Fig. 11). These validation analyses indicate that the primary *rs1800628*-associated immunoproteomic signature is highly reproducible across the investigated European populations.

While the UKB and deCODE cohorts validate the cross-cohort consistency of this signature in adults, we sought to determine if this genetically driven immunological set-point is established early in life. Analyses of multiple early-life datasets indicated that the *rs1800628* variant is associated with persistent alterations in complement system protein levels from birth. Specifically, the variant is strongly associated with reduced neonatal circulating complement C4 levels (N = 68,768, beta = −0.55, *P* < 1 × 10^−300^)^31^, which persists into childhood and adolescence as decreased C4A protein levels (GCST90453157, N = 1,909, beta = −1.09, *P* = 6.54 × 10^−93^, Supplementary Table 40).

This protein-level deficiency is highly concordant with bulk eQTL data demonstrating down-regulated *C4A* gene expression (*P* = 1.06 × 10^−121^, Supplementary Table 28). Concurrently, the variant is associated with elevated levels of complement C2 during early development (*P* = 8.88 × 10^−11^, Supplementary Table 40)^32^—a protein that also demonstrated significance (*P* < 1 × 10^−11^) in both the UKB and deCODE adult cohorts (Supplementary Fig. 12). This early-life concordance provides population-level genetic support for our hypothesis that the *rs1800628*-tagged MHC signal shapes a lifelong immunological trajectory long before the clinical onset of autoimmunity or age-related viral reactivation.

Finally, extended pairwise local SuSiE-coloc analysis at the *rs1800628* locus supported signal sharing across 30 traits. Strong colocalization was consistently observed across all 435 comparisons within a 10 kb window and remained prevalent upon expansion to 50 kb (391 of 435 pairs). The broader 50 kb interval primarily redistributed the lead signal from *rs1800628* to variants in strong LD, whereas genuinely distinct signals accounted for less than 10% of the strong colocalization events (Supplementary Table 41).

## Discussion

By integrating large-scale GWAS summary statistics with multi-omics datasets, we identified a pleiotropic immune-regulatory signal—tagged by *rs1800628* within the MHC—that is jointly associated with the aging trajectory, RA, and VZV reactivation risk. The phenome-wide mapping indicated that this genetic signal extends beyond RA, associated with multiple autoimmune phenotypes. Subsequent network and *cis*-MR analyses implicate this shared signal in systemic immune remodeling. This transition from a specific disease overlap to a generalized mechanistic hypothesis highlights a shared genetic basis for the “paradox of immune aging“—where aging individuals exhibit dual vulnerability to opportunistic infections (e.g., VZV reactivation) and autoimmune activation^6^. Collectively, our findings indicate that the epidemiological link between autoimmunity and VZV reactivation is not merely a downstream consequence of extrinsic immunosuppression^1,2^, but may be rooted in a shared, genetically regulated immune trajectory that also contributes to the aging trajectory.

The Manhattan plots and LAVA indicate that the MHC and subregions represent the dominant association hotspot contributing to this shared susceptibility. The highly polymorphic *HLA* genes are critical for both viral antigen presentation and autoimmune susceptibility^11,33^. However, attributing this to specific SNPs is confounded by the extensive LD and pleiotropy inherent to this region. To resolve this, we employed an LD-aware, cross-trait colocalization framework. This approach bridges these traits by deconvoluting the macroscopic associations into distinct pleiotropic signal sets. By applying LD-aware clustering, we mitigated the risk of interpreting highly correlated ‘shadow signals’—which merely represent different localized projections of an extensive LD architecture—as multiple independent mechanisms. Rather than a single causal variant, this specific pleiotropic locus comprises multiple variants in strong LD that ultimately converge into unified, independent signal sets, each regulating distinct genes to collectively contribute to extensive pleiotropy. Our colocalization framework identified the *rs1800628*-tagged signal as a key nexus across the three traits. When the multidimensional effect sizes of this signal set are projected onto a shared PC axis—mathematically representing the primary axis of pleiotropic covariation—they converge into a consistent spatial trajectory. Interpreted through the lens of biological directionality, one pole of this axis corresponds to healthy aging and protection against RA and HZ, whereas the opposite pole is associated with accelerated aging alongside a heightened risk for both diseases. Functional enrichment and protein network analyses supported the involvement of shared pathways at different levels (e.g., antigen presentation and TNF-related inflammatory pathways).

To translate these genetic associations into biological mechanisms, we investigated the downstream functional consequences of this shared signal. PheWAS screening and early-life proteomic mapping highlighted its role as a pleiotropic regulator of numerous immune-related plasma proteins. A prominent innate immune feature associated with this signal variant is a persistent, longitudinal reduction in complement C4 (specifically C4A)—a deficiency classically linked to impaired clearance of immune complexes and an increased risk of systemic autoimmunity, as supported by recent large-scale neonatal and adult genetic studies^31^. Concurrently, the signal variant is associated with an elevation of pro-inflammatory mediators (e.g., IL15, CXCL9)^26,27^—hallmarks of a hyper-responsive innate immune state^3,34,35^—and the decoy receptor IL2RA^28,29^, alongside an increase in T-cell tolerance and exhaustion markers (e.g., PDCD1, TIGIT)^25^. These molecular shifts were accompanied by reduced peripheral leukocyte and lymphocyte counts, lower levels of circulating CD4 and CD8A proteins, and decreased CRP. By implementing a cross-cohort cis-MR between independent UKB and deCODE cohorts, we reconstructed the genetically inferred directional network among these candidate proteins. This cross-protein causal mapping provided genetic evidence that these concurrent molecular shifts are not merely parallel epiphenomena, but participate in a highly structured regulatory cascade, delineating distinct upstream regulators and downstream effectors. Utilizing these candidate plasma proteins as exposures against extensive immune cell phenotypes supported a broad downstream causal cascade. These analyses revealed divergent genetic associations with regulatory T cell (Treg) traits. While elevated immune checkpoints, such as PDCD1 and TIGIT, are genetically predicted to enhance Treg abundance, concurrently, elevations in pro-inflammatory mediators (e.g., IL15, CXCL9) and IL2RA have been reported in prior functional studies to potentially compromise Treg suppressive capacity^25–29^.

Based on these observations, we propose a unifying conceptual framework: the “chronic high inflammatory burden–compensatory immune tolerance dysregulation” model. The core feature of this model—the concurrent elevation of Treg/tolerance markers alongside an increased autoimmune risk—appears contradictory, given the established role of Tregs in limiting autoimmune responses. This phenomenon may be rooted within the critical window of early-life immune programming. During this highly plastic developmental phase, the immune system establishes a compartment of regulatory T cells that defines the lifelong immunological set-point for self-tolerance^7,8^. We hypothesize that the pleiotropy of this immunogenetic signal drives an early and persistent elevation of specific pro-inflammatory mediators. This sustained, genetically predicted chronic inflammatory burden functionally and epigenetically reprograms innate immune circuits, establishing an innate-like hyper-responsive state that mirrors trained immunity^35–37^, thereby shaping the baseline immunological set-point long before clinical pathology emerges. Recent experimental models provide support for this paradigm. On one hand, studies in autoimmune-prone mice have demonstrated that endogenous inflammatory environments can induce epigenetic reprogramming in hematopoietic stem cells, establishing a persistent trained immunity phenotype even in the absence of exogenous pathogens^37^. On the other hand, induction of trained immunity can persistently lower the activation threshold of innate immune cells, thereby accelerating and exacerbating the pathogenesis of autoimmune arthritis^38–40^.

To counteract this hyper-responsiveness, the immune system induces a compensatory upregulation of adaptive tolerance mechanisms, thereby establishing a new, albeit precarious, immunological set-point. Our cis-MR findings support this compensatory loop, providing genetic evidence that elevated TIGIT and PDCD1 plasma protein levels promote Treg expansion. This mechanism is consistent with *in vivo* models demonstrating that systemic administration of exogenous soluble TIGIT (TIGIT-Fc) directly triggers Treg expansion and restores systemic immune tolerance^41,42^. Simultaneously, elevated inflammatory mediators (e.g., IL15, CXCL9)^26,27^ and IL2RA—which acts as an IL-2 sink—can partially impair Treg function^28,29^. This complex interplay establishes a fragile chronic inflammation-tolerance equilibrium that maintains clinical homeostasis during early development while permitting the gradual accumulation of autoimmune potential.

However, this fragile compensatory state may not be indefinitely sustained. As individuals transition from this early, stressed immunological set-point toward adult immune homeostasis, the relative threshold of immune tolerance naturally recedes. This physiological recalibration renders the maturing immune system unable to adequately suppress the underlying hyper-responsive state. Combined with the cumulative pressure of environmental and microbial stimuli, the disruption of this equilibrium exposes the underlying inflammatory memory, precipitating the onset and progression of autoimmune diseases, such as RA. This dynamic is clinically supported by observations that blocking these compensatory checkpoints (e.g., via immune checkpoint inhibitors) can induce or exacerbate autoimmunity^43,44^. Later in life, age-related immunosenescence synergizes with this intrinsic genetic tendency toward immune exhaustion (marked by elevated TIGIT and PDCD1)^3,5^. This dual burden of natural senescence and lifelong compensatory tolerance impairs the local cellular surveillance required to prevent VZV reactivation, permitting higher viral antigen expression, elevated anti-VZV antibody responses, and ultimately, an increased risk of HZ^4,45,46^.

This genetic framework also contextualizes the clinical observation that the administration of anti-TNF therapy is associated with a further increased risk of HZ in different autoimmune diseases^2^. Conceptually, autoimmune pathogenesis has long been attributed to TNF-driven inflammatory cascades within specific local microenvironments (e.g., the synovial tissue in RA)^47,48^. This mechanistic paradigm underpins the clinical success of systemic anti-TNF agents, which have substantially advanced the management of these diseases and remain a cornerstone of targeted therapy^47,48^. Our colocalization-constrained single-SNP MR analysis between bulk eQTL and RA is consistent with this clinical consensus, supporting that genetically predicted elevation in systemic *TNF* expression is associated with an increased disease risk (Supplementary Table 7). However, the single-cell data introduce an additional shift: cell-type-specific *TNF* expression in key antiviral subsets—such as dendritic cells—actually acts as a protective factor (Supplementary Table 7). The identified pleiotropic genetic signal intrinsically down-regulates TNF-axis expression within these specific subsets (e.g., *TNF* expression in dendritic cells and *LTA* expression in *SOX4*-expressing CD4+ T cells). Consequently, we hypothesize that while systemic anti-TNF therapy effectively suppresses pathogenic tissue inflammation, it inadvertently compounds these pre-existing, genetically predicted local immune deficits in autoimmune diseases. This ‘dual hit’—where iatrogenic suppression overlaps with intrinsic genetic vulnerabilities in the same pathway—implies that the further elevated HZ risk observed in these patients is not solely a drug-induced side effect, but is partially rooted in their baseline immunogenetic architecture.

This study has several limitations. First, the computational nature of our findings necessitates *in vivo* experimental validation. While our findings are robust across multiple independent cohorts, computational inferences of protein interactions and intercellular communication should be interpreted with caution, as they act as proxies for, rather than direct measurements of, biological interactions. Second, while age-stratified cross-sectional datasets support early-life complement dysregulation, the absence of continuous, intra-individual longitudinal cohorts limits the direct observation of the lifelong immune programming trajectory. Third, the extensive linkage disequilibrium in the MHC region precludes the definitive statistical isolation of a single causal variant. This limitation requires future integration of high-resolution HLA typing and functional genomics to disentangle the relative contributions of classical HLA structural alleles versus non-coding regulatory variants within the identified pleiotropic signal sets. Finally, the lack of non-European ancestry data warrants caution regarding the generalizability of our immune dysregulation model across diverse populations.

By employing a cross-phenotype and cross-cohort strategy, we mapped the genetic landscape of the *rs1800628*-tagged signal set across eight initial immune traits and expanded this validation to an extensive array of 30 diverse autoimmune and immune-related phenotypes. This locus extends beyond the RA–HZ intersection, potentially acting as a shared genetic node broadly implicated in human immune regulation. Our discovery of this anchor aligns with independent genetic investigations. The *rs1800628* variant was recently implicated as a shared genetic anchor across diverse autoimmune diseases^9^. Parallel studies have identified this variant—alongside its strongly linked proxies (e.g., *rs3099844*, LD R^2^ = 0.938; *rs2734583*, LD R^2^ = 0.971)—as shared genetic risk factors not only for SLE and SS, but also for distinct immune-mediated traits including primary sclerosing cholangitis and severe drug-induced hypersensitivity^49–51^. However, while previous research characterized these associations as isolated or autoimmune-specific signals, our LD-aware signal deconvolution and phenome-wide mapping expand and unify these previous findings. Our findings highlight that this genetic anchor extends beyond linking multiple autoimmune diseases to shape a lifelong trajectory of immune remodeling. Within this framework, immunosenescence, autoimmunity and VZV reactivation emerge not as isolated clinical events, but as interconnected manifestations of a shared, dysregulated immunological set-point. Collectively, by mapping this genetically inferred trajectory of innate hyper-responsiveness and compensatory tolerance, our study provides a conceptual framework for understanding the early shift in the immunological set-point, and potential targets related to altered innate immune memory^52,53^ to prevent these interconnected immune-mediated disorders.

## Methods

### Ethics statement

This research complies with all relevant ethical regulations. All data used in this study were obtained from publicly available summary-level datasets. The original studies from which these data were derived obtained ethical approvals from their respective institutional review boards and informed consent from all participants. Therefore, no additional ethical approval or informed consent was required for this secondary data analysis.

### Data sources and phenotype definitions

To investigate the shared genetic architecture connecting aging, autoimmunity, and viral reactivation, we integrated large-scale GWAS summary statistics primarily from European-ancestry populations. All genetic coordinates were harmonized and lifted over, where necessary, to the GRCh37 (hg19) reference assembly.

Our core analyses leveraged a large-scale multivariate GWAS of aging-related traits (n = 1,958,774). This study utilized genomic structural equation modeling across five traits (healthspan, parental lifespan, exceptional longevity, frailty index, and epigenetic age acceleration) to derive a latent genetic factor, termed mvAge^12,54^. Biologically, mvAge captures genetically predicted healthy aging; thus, higher values indicate extended healthspan and reduced frailty. All cross-trait interpretations were based on this encoding (OR > 1: healthier aging; OR < 1: accelerated aging)^12^. Disease-specific GWAS datasets included rheumatoid arthritis (RA; 25,533 cases and 290,135 controls)^13^. To prevent sample overlap biases in multi-omics integrations, we leveraged two independent HZ datasets with distinct analytical roles. The UKB whole-genome sequencing cohort (17,856 cases and 440,584 controls)^14^ was employed as the primary dataset for genome-wide colocalization-constrained discoveries. For two-sample MR analyses—particularly those utilizing UKB-derived plasma pQTLs^18^ as exposures—we utilized the independent FinnGen study (Release 12; 7,132 cases and 480,316 controls)^55^ to ensure robust causal inference.

To map these macroscopic phenotypic associations to specific molecular and cellular regulatory networks, we integrated high-resolution multi-omics datasets. Transcriptomic anchors included single-cell *cis*-eQTLs across 14 immune cell types from the OneK1K cohort (n = 982)^15^ and bulk eQTLs^56^ downloaded from the OpenGWAS database^24^. For proteomic and cellular integration, we utilized plasma pQTLs for > 2,900 proteins measured in ∼47,745 UKB participants^18^ alongside GWAS summary statistics for 731 immune cell traits from 3,757 SardiNIA study participants^20^. To circumvent sample overlap bias in cross-protein causal network reconstructions and to ensure rigorous external validation of the adult immunoproteomic signature, we incorporated the independent deCODE Icelandic cohort. This dataset comprises GWAS summary statistics for 4,907 plasma proteins quantified via the SomaScan platform in 35,559 individuals^19^. Furthermore, to investigate whether the identified genetic associations are established during early development, we integrated two independent early-life GWAS datasets: a large-scale cohort assessing neonatal circulating complement C4 concentrations (iPSYCH, N = 68,768)^31^, and a recently published children and adolescent plasma proteome GWAS (HOLBAEK, N = 1,909)^32^.

Finally, to assess infection-induced transcriptional perturbations at GWAS-implicated loci, we analyzed peripheral blood bulk RNA-seq data from the Gene Expression Omnibus (GEO) dataset GSE242252, comprising 26 acute HZ samples, 23 follow-up samples collected one year after infection, and 31 healthy controls^10^.

All summary statistics used for this study are detailed in Supplementary Table 1.

### Local Analysis of [co]Variant Associations (LAVA)

To investigate the shared local genetic architecture among mvAge, RA, and HZ, we applied LAVA (v0.1.5)^21^ to estimate bivariate local genetic correlations across the extended MHC region. The region was partitioned into 22 LD blocks (GRCh37 coordinates spanning from 24,950,380 to 34,979,270; detailed in Supplementary Table 2) using the default LAVA partitioning algorithm^57^, to minimize LD between blocks. Local genetic correlations were computed utilizing the preprocessed UKB binary LD reference panel (v1.1) provided by LAVA, which estimates LD from 100,000 unrelated European-ancestry individuals. Given its design as an exploratory screening to observe local correlation trends rather than establish definitive causality, we reported nominal *P*-values without FDR correction. To ensure the integrity of the local genetic correlation estimates against the external LD reference panel, rigorous summary statistics preprocessing was applied. Specifically, variants with minor allele frequencies (MAF) < 0.05 or ambiguous palindromic alleles (A/T, C/G) were excluded. The remaining variants were strictly harmonized against the UKB LD reference panel; for alleles flipped relative to the reference, the effect size (Beta) and effect allele frequency (EAF) were correspondingly inverted, and non-matching variants were discarded.

### Colocalization-constrained MR analysis

To investigate the shared genetic architectures between eQTL datasets (18,549 bulk eQTLs and 46,361 sc-eQTLs) and GWAS traits, we performed fine-mapping and genetic colocalization utilizing the susieR and coloc R packages. We employed a comprehensive framework integrating two complementary approaches. First, we utilized SuSiE fine-mapping (susie_rss; maximum number of causal variants L = 10) to identify 95% credible sets^16^, accommodating multiple putative driving variants per locus. This was followed by coloc.susie to test for colocalization between eQTL and GWAS credible sets^17^. Second, we applied traditional colocalization (coloc.abf) to capture robust signals in simpler loci under the single causal variant assumption^58^. Default prior probabilities were applied across all tests (*p1* = 1 × 10^−4^, *p2* = 1 × 10^−4^, and *p12* = 1 × 10^−5^). Prior to fine-mapping, to ensure stable Bayesian estimation and valid LD matrix computation, GWAS and eQTL summary statistics were stringently quality-controlled. We restricted the analyses to common variants (MAF ≥ 0.05) with standard alleles (A, C, G, T). Within each evaluated genomic window, three-way allele harmonization was enforced among the eQTL dataset, the GWAS dataset, and the UKB LD European reference panel from Zenodo^59^. Alleles, effect sizes, and EAFs were dynamically aligned to the LD reference, and variants failing harmonization were excluded prior to executing the SuSiE iterative Bayesian stepwise selection (IBSS) algorithm.

Given the extreme LD complexity and dense gene clustering within the extended MHC region, traditional coloc.abf estimations are unreliable for resolving true shared etiology in this region. Consequently, we excluded the MHC from traditional coloc.abf evaluations, retaining exclusively SuSiE-based colocalization results for MHC loci. To address the high multicollinearity and dense LD inherent to the MHC region—which frequently limits the convergence of the IBSS algorithm of SuSiE when applied to macro-regions—we implemented a multi-window strategy. Analyses were conducted across four incrementally constrained genomic windows: the full local LD block (Supplementary Table 4), and localized extensions of ±100 kb, ±50 kb, and ±10 kb from eQTL gene boundaries, strictly bounded within their respective local LD blocks. A posterior probability of colocalization (PP.H4) > 0.8 in any evaluated window was utilized not as definitive proof of a single shared causal variant, but as a quantitative heuristic to capture the presence of a shared local association signal, thereby serving as a stringent genetic constraint to prioritize robust instrumental variables for subsequent causal inferences (e.g., Supplementary Fig. 1). While these localized windows facilitated algorithmic convergence, they inherently truncated the extended LD architecture. As a result, an extensive pleiotropic LD signal could be deconvoluted into multiple, highly correlated local colocalizations across adjacent eQTL genes— conceptually acting as ‘shadow signals’ that reflect different localized projections of the same underlying causal architecture. In our framework, which aims to identify pleiotropic ‘Signal Sets’ rather than isolate single causal variants, this dense LD and spatial consistency among proxy SNPs was interpreted as supporting evidence of a unified causal architecture. To mitigate statistical multicollinearity while preserving this unified architecture, these fragmented local signals were subsequently reconstituted into representative ‘Signal Sets’ via stringent LD-aware clustering (R² > 0.6). Finally, the spatial consistency of the shared immunogenetic signal set identified in our study across traits was further corroborated by macro-scale (±1 Mb) visual LD-pattern alignments.

MR effect estimates were derived using the Wald ratio and interpreted as genetically supported regulatory directions under colocalization constraints (PP.H4 > 0.8). Instruments were defined as the variant with the highest posterior inclusion probability in SuSiE or as the lead colocalized variant in coloc.abf. Instrument strength was evaluated using the F-statistic (*F* > 20), and Steiger filtering was applied to support the assumed exposure-to-outcome orientation^60^. Because these analyses relied on a single instrument, pleiotropy-robust sensitivity estimators were not applicable; instead, we relied on stringent colocalization constraints to mitigate bias driven by LD confounding or distinct underlying signals. MR analyses were performed using the TwoSampleMR package, with FDR < 0.05 considered significant^61^ (Supplementary Tables 5–14).

### Cross-trait genetic correlation and cell-type-specific pleiotropy extension

To evaluate the macroscopic consistency of genetic effects across aging, autoimmunity, and viral reactivation, we performed cross-trait correlation analyses anchored on the shared colocalized lead SNPs. For each overlapping locus, the harmonized MR effect sizes were aligned. Spearman’s rank correlation (cor.test function)^62^ was calculated to assess the monotonic relationship of regulatory effects across different phenotypic scales (e.g., mvAge versus RA or HZ). Correlation significance was evaluated exclusively for variant-trait pairs that survived stringent multiple testing (FDR < 0.05 in both corresponding analyses).

Furthermore, to dissect horizontal pleiotropy^63^ at the single-cell level, we extended the defined cell–SNP–gene architectures using the OneK1K sc-eQTL dataset^15^. For each colocalized lead SNP, we queried the full sc-eQTL summary statistics to identify all additional genes regulated by the exact same variant within the identical immune cell type. Expanded regulatory targets were filtered using an eQTL FDR threshold of < 0.05, and deduplicated to retain the single most significant association per cell–SNP–gene triad.

### LD-aware signal clustering and multidimensional spatial mapping

As described above, localized colocalization strategies can fragment extensive pleiotropic architectures into highly correlated local projections. To prevent redundant signal inflation from these fragmented proxies in cross-trait comparisons, we implemented an LD-aware clustering framework. Utilizing the UKB European reference panel^59^ and PLINK (v1.9)^64^, pairwise LD (R^2^) was calculated for all genome-wide significant lead SNPs within a 10 Mb window. SNPs exhibiting strong LD (R2 > 0.6) in the same cell-type were reconstituted into unified signal sets (Exposure Clusters).

To visually represent the shared genetic etiology across mvAge, RA, and HZ, we mapped these independent LD clusters into a three-dimensional spatial coordinate system. A feature matrix of scaled MR effect sizes was constructed, and dimensionality reduction was performed via Principal Component Analysis (PCA; prcomp function)^62^. This process was tailored to handle LD redundancy by explicitly selecting a single representative SNP to proxy each target gene within the cluster. Hierarchical clustering based on Euclidean distance was further applied to the matrix to order the spatial distribution. This multidimensional mapping aligned the complex pleiotropic architecture onto a unified principal component axis, polarizing the genes in the shared genetic signal: separating genes driving concordant protection (healthier aging and reduced disease risk) from those driving shared pathogenesis (accelerated aging and heightened disease risk).

### Cross-trait functional and network analyses

To functionally interpret the genes prioritized by our colocalization-constrained MR framework, we performed Gene Ontology (GO)^65^ and Kyoto Encyclopedia of Genes and Genomes (KEGG)^66^ pathway enrichment analyses utilizing the clusterProfiler R package^67^. Significant enrichment was defined using an FDR adjusted *P*-value < 0.05. To identify shared biological pathways potentially mediating the observed clinical links, we performed cross-trait intersection analyses on the significant enrichment terms using Venn diagrams, pinpointing conserved regulatory pathways overlapping among mvAge, RA, and HZ.

To contextualize the molecular mechanisms and functional connectivity of the trait-associated genes, we constructed PPI networks mapping the protein products of prioritized genes against the STRING database (v12.0)^22^. A medium-confidence threshold (minimum required interaction score ≥ 0.4) was applied to retain high-probability empirical and curated interactions. To map both unique and shared architectures, PPI networks were evaluated for individual traits, pairwise unions, and the three-trait union. Network topology was analyzed using the igraph R package^68^, calculating key centrality metrics including node degree and betweenness centrality to identify critical hub and bridge proteins. In the resulting network visualizations, nodes exhibiting high betweenness centrality within the union networks were spatially prioritized toward the core to highlight putative connector proteins mediating cross-trait molecular crosstalk.

To explore intercellular communication mechanisms underlying these trait-associated immune regulatory programs, we implemented a genetics-anchored framework utilizing the nichenetr R package^23^ alongside its updated prior-knowledge ligand-receptor-target regulatory matrices (NicheNet v2)^69,70^. MR/coloc-supported cell–gene pairs were designated as biologically anchored “cell-specific genes”. We conducted two complementary analyses: (i) an upstream analysis treating anchored genes as receiver-cell targets to identify potential regulatory ligands, and (ii) a downstream analysis treating the anchored protein products as sender-cell ligands to predict downstream regulatory impacts. The background ligand, receptor, and target universes were constrained to expressed genes specifically identified in the OneK1K peripheral blood immune-cell dataset. Finally, to decipher conserved intercellular regulatory motifs, we extracted the top 30 upstream regulatory ligands and downstream target predictions based on regulatory potential scores for each trait, enabling a cross-trait comparison of key immune communication hubs.

### *Cis*-MR based on UKB pQTL

To infer the causal effects of plasma proteins on the risk of RA and HZ, we conducted a two-sample MR analysis utilizing the UKB plasma proteome (pQTL) dataset^18^. To minimize potential horizontal pleiotropy typically associated with *trans*-pQTLs, IVs were restricted to *cis*-acting variants. A *cis*-window was precisely defined as ±250 kb flanking the transcription start and end sites of the respective protein-coding gene^71^.

Within these constrained genomic regions, candidate *cis*-IVs were selected based on genome-wide significance (*P* < 5 × 10^−8^), MAF > 0.01, and instrument strength (*F*-statistic > 10). To extract independent genetic signals and mitigate multicollinearity prior to estimation, localized LD clumping was performed (R^2^ < 0.3, window = 10,000 kb) leveraging the UKB European reference panel^59^ (Supplementary Table 24). We subsequently harmonized the datasets by removing ambiguous palindromic SNPs with intermediate allele frequencies. To satisfy the MR exclusion restriction assumption, we excluded any instrumental variant demonstrating a strong direct association with the outcome trait (*P* < 5 × 10^−8^).

For causal estimation, the Wald ratio method was employed for proteins instrumented by a single variant, whereas the inverse-variance weighted (IVW) method served as the primary analysis for proteins with multiple IVs^72^. Statistical significance for the primary MR estimates was determined utilizing an FDR adjusted *P*-value < 0.05^61^. To avoid sample overlap bias—given that the primary pQTL data originated from the UKB cohort—MR estimations for HZ were conducted using the independent FinnGen HZ dataset as the outcome. Subsequently, to dissect the shared plasma proteomic etiology, we conducted cross-trait intersection analyses on the significant *cis*-MR results, quantitatively comparing the overlapping protein targets to determine whether their causal effect directions were concordant (contributing to shared pathogenic risks) or antagonistic (exerting protective trade-offs) between RA and HZ.

### PheWAS and downstream analyses

In our study, the *rs1800628* (G>A) variant was associated with reduced *TNF* expression in dendritic cells and *LTA* expression in *SOX4*-expressing CD4+ T cells, and with an increased risk of RA and HZ, as supported by colocalization between sc-eQTLs and these traits. Given the central role of the TNF-related signal across multiple analyses, *rs1800628* was selected as a representative SNP for the identified MHC immune-regulatory signal. We performed a PheWAS centered on *rs1800628* to decode the systemic immunomodulatory footprint of the MHC signal. Summary statistics were queried across 2,931 UKB plasma protein datasets, 826 UKB diseases and traits, 921 genes in the OneK1K sc-eQTL dataset, and 10,561 European-ancestry GWAS datasets available in OpenGWAS^24^.

For downstream functional and network analyses, UKB candidate plasma proteins *trans*-regulated by *rs1800628* were stratified based on two distinct significance thresholds. First, for proteins associated with the variant at a standard GWAS threshold (*P* < 5 × 10^−8^), we constructed a PPI network utilizing the STRING database (v12.0)^22^ with a high-confidence threshold (minimum required interaction score ≥ 0.7). Topological features, including node degree and betweenness centrality, were computed via the igraph R package to identify functional clusters and hub proteins. In parallel, intercellular communication was modeled using the NicheNet prior-knowledge network. Full ligand–receptor–target communication circuits were reconstructed with the strict constraint that all three interaction nodes—ligands, receptors, and downstream targets—must be present within the identified candidate plasma protein set. To enhance biological validity, we queried the OmniPath API^73^ to retain direction-annotated receptor interactions (stimulation or inhibition), further filtering target predictions by a regulatory potential > 0.01.

Second, to minimize false positives and isolate robust signals for downstream causal inference, we applied a more stringent association threshold (*P* < 1 × 10^−11^), yielding 195 candidate plasma proteins. The PPI and NicheNet analyses were repeated on this refined subset utilizing identical parameters. To move beyond observational interaction networks and reconstruct a genetically inferred causal hierarchy among the candidate plasma proteins, we performed a cross-protein cis-MR analysis. To adhere to the two-sample MR assumption and eliminate sample overlap bias, the UKB pQTL dataset was utilized exclusively to derive cis-acting instrumental variables for the exposures, while the independent deCODE Icelandic cohort served as the outcome dataset. Following strict allele harmonization between the two cohorts and data availability filtering, 176 unique exposure proteins and 164 outcome proteins were retained. Causal effects across all possible protein pairs were estimated using the Wald ratio and IVW methods (excluding self-causal evaluations where the exposure and outcome represented the homologous protein), with significance established at FDR < 0.05.

Furthermore, to determine the causal downstream effects of these candidate plasma proteins on the systemic immune cell landscape, we evaluated these 195 proteins as exposures in a *cis*-MR framework against 731 immune cell GWAS traits. To maintain methodological consistency across the study, we utilized the exact same set of stringently filtered UKB pQTL IVs that were employed in the preceding *cis*-MR analyses of RA and HZ. Causal estimates were calculated using the IVW and Wald ratio methods, with significance established at FDR < 0.05. To exclude potential LD confounding, all FDR-significant protein–immune cell MR pairs were further subjected to colocalization analysis. Colocalization was assessed within a ±250 kb window flanking the target protein-coding genes utilizing both traditional colocalization (coloc.abf) and the SuSiE fine-mapping framework.

Finally, to verify the pleiotropy of the *rs1800628*-tagged signal, SuSiE-based colocalization analyses were conducted across eight traits of interest (including plasma proteins IL2RA, IL15, and TIGIT, alongside lymphocyte counts, SLE from UKB and FinnGen, SS, and DM). Given the extensive LD blocks in the MHC region, we applied a local window strategy by restricting the analyses to ±10 kb and ±50 kb regions centered on *rs1800628*. Visual evaluation of regional association architecture was performed using Manhattan plots spanning ±1 Mb around the index variant to assess the spatial consistency of LD-linked variants across traits.

### Sensitivity and robustness analyses

To evaluate the internal coherence and external validity of our integrative colocalization-constrained MR framework, we designed a set of sensitivity analyses. First, to assess internal coherence, we utilized the FI GWAS as a sensitivity trait. Because FI is one of the five constituent traits used to derive the latent mvAge factor—and was directionally flipped by the original authors to align with “healthier aging“^12^—we expected an antagonistic relationship. We extracted the shared cell–gene–SNP triplets between mvAge and FI, calculating the Spearman correlation of their respective MR effect sizes to quantitatively confirm this inverse directional consistency. Second, to confirm the cross-cohort robustness of our estimated effect directions, we performed external validation using an independent HZ GWAS from the FinnGen consortium^55^ for both the mvAge–UKB HZ and RA–UKB HZ shared triplets.

To evaluate whether acute HZ infection confounds transcriptional signals at GWAS-implicated loci (i.e., reverse causality via secondary immune activation), we analyzed peripheral blood bulk RNA-seq data from the GEO dataset GSE242252^10^. This dataset comprises healthy controls, acute HZ cases, and convalescent individuals approximately one year post-infection. We quantified the expression levels of GWAS-implicated HZ risk genes identified by our framework and tested for differential expression across these clinical stages using the Kruskal-Wallis test for global comparisons and Wilcoxon rank-sum tests for pairwise evaluations, followed by stringent FDR correction.

To validate the cross-cohort robustness of the *rs1800628*-associated plasma protein profile, we performed an external validation using the independent deCODE Icelandic cohort. Plasma proteins from the primary UKB cohort and the deCODE cohort were matched by their encoding gene symbols. Strict allele alignment was applied prior to comparison; effect estimates (beta values) from the deCODE dataset were inverted if the effect and reference alleles were flipped relative to the UKB dataset. Ambiguous variants were excluded. Directional concordance rates were then evaluated across three thresholds: (1) all mapped proteins, (2) proteins reaching high significance in the primary UKB cohort (*P* < 1 × 10^−11^), and (3) proteins reaching high significance in both cohorts (*P* < 1 × 10^−11^).

To investigate whether this genetically predicted immunoproteomic signature is established during early development—prior to the clinical onset of age-related and autoimmune diseases—we examined the association of the rs1800628 variant across two independent early-life datasets. Specifically, we evaluated neonatal circulating complement C4 concentrations (iPSYCH)^31^ alongside a pediatric and adolescent plasma proteome GWAS (HOLBAEK)^32^ to determine whether key complement proteins identified in adult cohorts (e.g., C4/C4A and C2) exhibited concordant genetic associations in early-life developmental stages.

Finally, to test the statistical robustness of the colocalization algorithm against varying local LD boundaries, we extended the pairwise SuSiE-coloc analysis at the critical *rs1800628* locus to an expanded panel of 30 immune and disease traits, generating 435 pairwise comparisons. We compared fine-mapping and colocalization patterns between ±10 kb and ±50 kb genomic windows to evaluate whether the shared genetic signals were driven by identical variants or tightly linked proxies.

### Statistical analyses

All statistical analyses, including multidimensional spatial mapping and hierarchical clustering, were performed using R statistical software version 4.5.1 (R Foundation for Statistical Computing). Unless otherwise specified, an FDR adjusted *P*-value < 0.05 was considered statistically significant to account for multiple testing.

### Use of Artificial Intelligence (AI) tools

During this study, the authors used Gemini-3.1 via the TRAE integrated development environment to assist with writing specific data processing and statistical analysis scripts, and to polish the English language. The authors emphasize that the conceptualization, study design, oversight, data interpretation, and derivation of all scientific conclusions were entirely driven by the human authors. Following the use of this AI tool, the authors reviewed, tested, and edited the generated code and text. The human authors take full and final responsibility for the scientific content, data integrity, and conclusions of this publication.

## Data availability

All GWAS summary statistics and multi-omics data supporting the findings of this study are publicly available. Detailed accession numbers, download URLs, and literature references for all datasets are comprehensively provided in Supplementary Table 1. The bulk RNA-seq dataset can be accessed in the NCBI GEO under accession number GSE242252. There are no restrictions on data access for any of the datasets used in this study. To facilitate the exploration of cross-trait genetic effects and complex causal networks, fully interactive 3D visualizations corresponding to the 3D scatter plot and the hierarchical edge-bundling network are hosted online and freely accessible via GitHub Pages at https://yaoxinyaoxinyaoxin.github.io/MHC-A2H/. For long-term archival and offline viewing, these interactive plots, along with the full summary results generated in this study, are provided on Zenodo (https://doi.org/10.5281/zenodo.20095908).

## Code availability

The custom analytical codes and pipelines used for data processing, MR, and colocalization analyses in this study are available in the GitHub repository (https://github.com/yaoxinyaoxinyaoxin/MHC-A2H). A persistent, archival version of this code is available on Zenodo (https://doi.org/10.5281/zenodo.20095908). All software tools, packages, and their specific versions utilized in this study are publicly available and detailed in the Methods section.

## Acknowledgements

We gratefully acknowledge all the participants and investigators of the primary cohorts whose dedication made this comprehensive multi-omics integration possible. We extend our profound gratitude to the foundational research teams who made their genome-wide association summary statistics publicly and unrestrictedly available. Specifically, we thank the authors and consortia responsible for curating the data derived from the UK Biobank (UKB), the deCODE genetics study, the FinnGen study, the Million Veteran Program (MVP), the iPSYCH consortium, the HOLBAEK study, and the TwinGene project. Furthermore, we acknowledge the eQTLGen Consortium, the OneK1K cohort investigators, and the SardiNIA project team for providing open access to their bulk eQTL, sc-eQTL, and immune cell trait datasets. Finally, we thank the authors of the foundational multivariate aging (mvAge) study and the acute HZ transcriptomic dataset (GSE242252), as well as the NHGRI-EBI GWAS Catalog and the MRC IEU OpenGWAS infrastructure for providing robust platforms that facilitate the unrestricted retrieval of these vital genetic resources.

## Author contributions

X.Y. designed the study, performed the formal data analyses, and was responsible for data visualization. X.Y. and M.W. jointly wrote the manuscript. X.Y. oversaw the overall project as the corresponding author. Both authors read and approved the final manuscript.

