## Supplementary Figure for "A shared MHC immunogenetic signal connects aging, rheumatoid arthritis, and herpes zoster through chronic high inflammatory burden–compensatory immune tolerance dysregulation"

**Supplementary Figure Legend**

**Supplementary Fig. 1 | Representative visualization of genetic colocalization across different genomic windows.** This figure shows the genetic colocalization between the single-cell expression quantitative trait loci (sc-eQTL) of *SAPCD1-AS1* in CD4+ naive and central memory T cells (CD4NC) cells (OneK1K cohort, N = 982) and three distinct traits: multivariate aging latent factor (mvAge), rheumatoid arthritis (RA), and herpes zoster (HZ from UK Biobank) (columns, from left to right). The analyses are shown across dynamically extended genomic windows from the specific gene boundaries: ±10 kb (top row), ±50 kb (middle row), and ±100 kb (bottom row). This multi-window approach was employed to minimize genomic window selection bias and evaluate the robustness of the colocalization signals (posterior probability of colocalization, PP.H4).

**Supplementary Fig. 2 | Genome-wide colocalization-constrained Mendelian randomization associations.** Visualizations of the colocalization-constrained MR analysis results across the genome. **a–c**, Circular plots based on OpenGWAS bulk eQTL data (eQTLGen consortium, N = 31,684) for mvAge (**a**), rheumatoid arthritis (**b**), and herpes zoster (**c**). **d–f**, Chord diagrams based on OneK1K sc-eQTL data (N = 982) for mvAge (**d**), rheumatoid arthritis (**e**), and herpes zoster (**f**). The purple background highlights the linkage disequilibrium (LD) blocks localized within the extended MHC region.

**Supplementary Fig. 3 | Cell–SNP-anchored expansion of local regulatory neighborhoods.** Delineation of pleiotropic targets within the extended MHC region based on OneK1K sc-eQTL summary statistics. The expansion captures local regulatory networks where individual pleiotropic signals modulate multiple adjacent genes in a cell-type-specific manner (false discovery rate < 0.05).

**Supplementary Fig. 4 | Cross-trait functional enrichment analysis of prioritized gene sets.** Gene Ontology (GO) and Kyoto Encyclopedia of Genes and Genomes (KEGG) enrichment analyses for genes prioritized by colocalization-constrained MR across mvAge, RA, and HZ. The results display shared functional signatures, prominently including antigen processing and presentation, and pro-inflammatory signaling cascades.

**Supplementary Fig. 5 | Protein–protein interaction networks of cross-trait intersections.** Topological protein–protein interaction (PPI) networks displaying the structural connectivity among proteins shared across different phenotype combinations. The networks correspond to the unions of mvAge & HZ (UKB), mvAge & RA, RA & HZ (UKB), and the three-way union of mvAge & RA & HZ (UKB).

**Supplementary Fig. 6 | Predicted intercellular communication networks inferred via NicheNet.** Computational predictions of upstream ligands and downstream targets based on the genetics-anchored NicheNet framework. The visualization illustrates the predicted ligand-target interaction profiles across mvAge, RA, and HZ.

**Supplementary Fig. 7 | Extended phenome-wide associations of the pleiotropic MHC signal set tagged by *rs1800628*.** Volcano plots summarizing the phenome-wide association study (PheWAS) results for this representative signal (*rs1800628*, G>A) across distinct datasets. The y-axis represents the −log10(*P*-value) derived from two-sided association tests in the source summary statistics. **a**, Associations with UKB plasma proteins (N = 47,745). **b**, Associations with sc-eQTL genes (OneK1K cohort, N = 982). **c**, **d**, Associations with an extended spectrum of phenotypes screened from the OpenGWAS database.

**Supplementary Fig. 8 | Functional network analyses under a stringent association threshold.** Repeated PPI and NicheNet intercellular communication analyses using a refined set of 195 candidate plasma proteins (association threshold $P < 1 \times 10^{-11}$). The results support the stability of the functional modules involved in immune regulation and pro-inflammatory mediation.

**Supplementary Fig. 9 | Cross-cohort causal network of candidate plasma proteins.** Hierarchical edge-bundling visualization of the genetically predicted directed causal relationships among the core candidate plasma proteins, reconstructed via cross-cohort *cis*-MR. To circumvent sample overlap bias, *cis*-acting instrumental variables were derived exclusively from the UKB plasma proteome cohort (exposures, N = 47,745), while the causal effects were estimated in the independent deCODE Icelandic cohort (outcomes, N = 35,559). Nodes represent individual plasma proteins, and the directed edges connecting them indicate significant causal effects determined by the inverse-variance weighted (IVW) or Wald ratio methods. 3,342 significant causal relationships surviving strict false discovery rate (FDR) correction (FDR < 0.05) across the non-homologous evaluated pairs (n = 28,710) are displayed. The color gradient of the edges represents the origin (source) of the causal flow, delineating distinct upstream regulators and downstream effectors within the immunoproteomic network. An interactive 3D version of this causal network, allowing for detailed exploration of individual or several protein-protein interactions, can be downloaded for offline viewing as Supplementary File 2, or accessed directly online at https://yaoxinyaoxinyaoxin.github.io/MHC-A2H/.

**Supplementary Fig. 10 | Genetic colocalization between TIGIT plasma protein levels and regulatory T cell traits.** Bayesian colocalization analyses demonstrating shared genetic signals at the TIGIT locus (tagged by *rs2654758*) between TIGIT plasma protein levels (N = 47,745) and specific regulatory T cell phenotypes (SardiNIA cohort, N =

1,244–3,669). **a**, Colocalization between TIGIT protein levels and *CD25* expression on resting CD4 regulatory T cells (posterior probability of colocalization [PP.H4] = 0.928). **b**, Colocalization between TIGIT protein levels and the proportion of CD4 regulatory T cells among total CD4+ T cells (PP.H4 = 0.854). These findings provide strong genetic support for the downstream causal effects of elevated TIGIT levels on regulatory T cell expansion and activation.

**Supplementary Fig. 11 | Cross-cohort validation of *rs1800628* effect sizes on the plasma proteome between the UK Biobank and deCODE cohorts.** Scatter plot comparing the causal effect sizes (betas) of the *rs1800628* variant on 1,772 shared plasma proteins between the UK Biobank (UKB; N = 47,745) and deCODE (N = 35,559) cohorts. The x-axis represents the effect size in the UKB cohort, and the y-axis represents the effect size in the deCODE cohort, with all alleles strictly aligned to the same effect allele. Colors indicate significance and directionality categories based on a highly stringent two-sided GWAS summary statistics threshold of $P < 1 \times 10^{-11}$: "Both Sig Concordant" (orange, N = 12), "UKB Sig Concordant" (yellow, N = 110), "Both Sig Discordant" (dark blue, N = 1), and "UKB Sig Discordant" (light blue, N = 26). The high proportion of directionally concordant signals across both cohorts supports the robustness and cross-population reproducibility of the variant's genetically predicted regulatory impact on the proteome.

**Supplementary Fig. 12 | Genetic associations of the *rs1800628*-tagged signal with circulating complement C2 and C4 levels across early development and adulthood.** Manhattan plots depicting the genome-wide association signals for plasma complement protein levels across four independent European cohorts. **a–c**, Associations for C2 protein levels in the UKB (adults, N = 47,745; **a**), deCODE (adults, N = 35,559; **b**), and HOLBAEK studies (children and adolescents, N = 1,909; **c**). **d**, Associations for C4A protein levels in the HOLBAEK study (N = 1,909). **e**, Associations for neonatal circulating C4 protein concentrations in the iPSYCH cohort (N = 68,768). Single nucleotide polymorphisms (SNPs) are colored based on their linkage disequilibrium (LD, $R^2$) with the index variant *rs1800628* (red dots, $R^2 > 0.8$), computed using the UKB European LD reference panel. The red and orange dashed lines denote the genome-wide significance threshold ($P = 5 \times 10^{-8}$) and the suggestive significance threshold ($P = 1 \times 10^{-5}$), respectively. The spatial consistency of the LD-structured association peaks across these developmental stages supports the hypothesis that the genetic associations with immune-related proteins linked to this MHC signal may be present early in life.

### Supplementary Figure 1

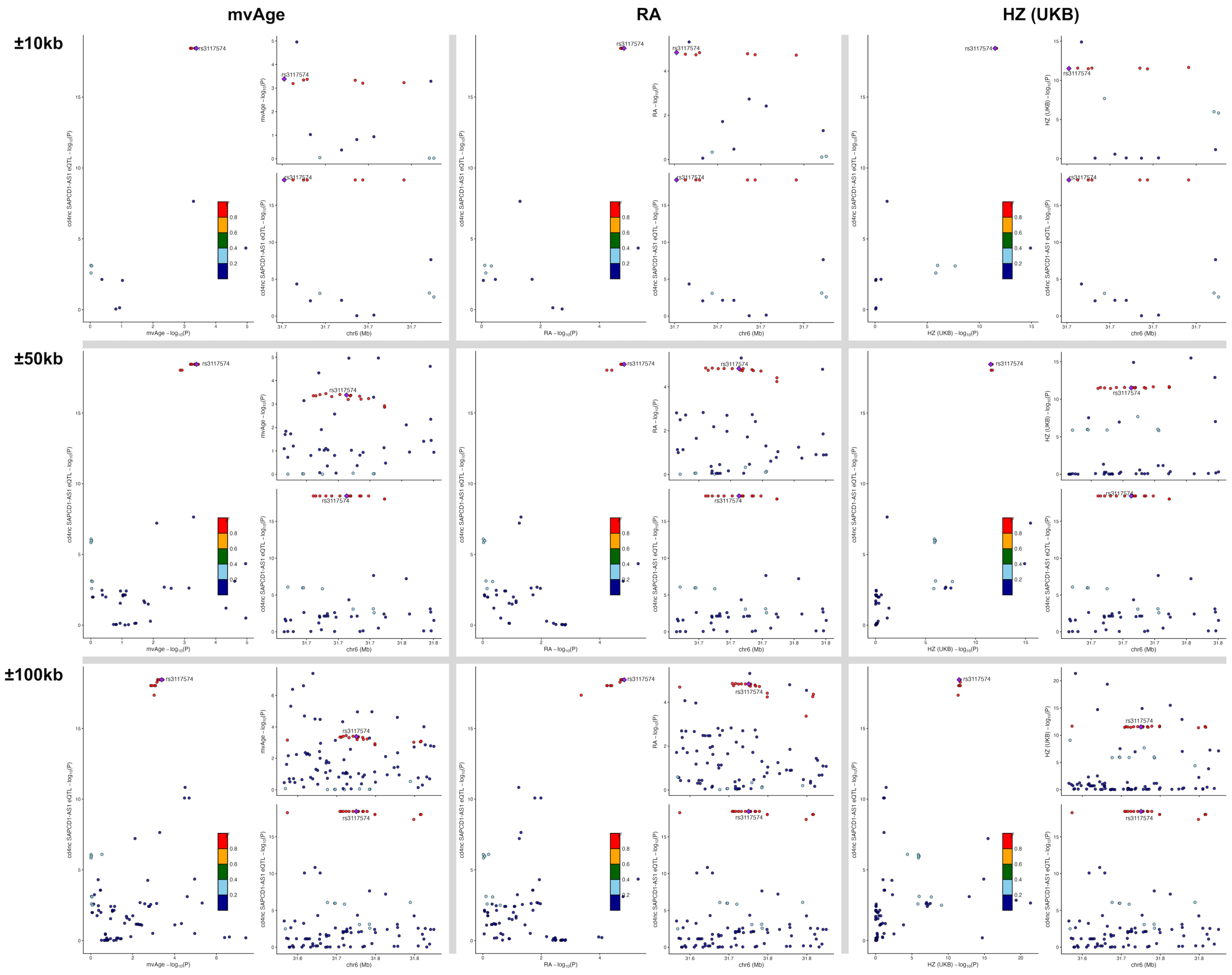

### Supplementary Figure 2

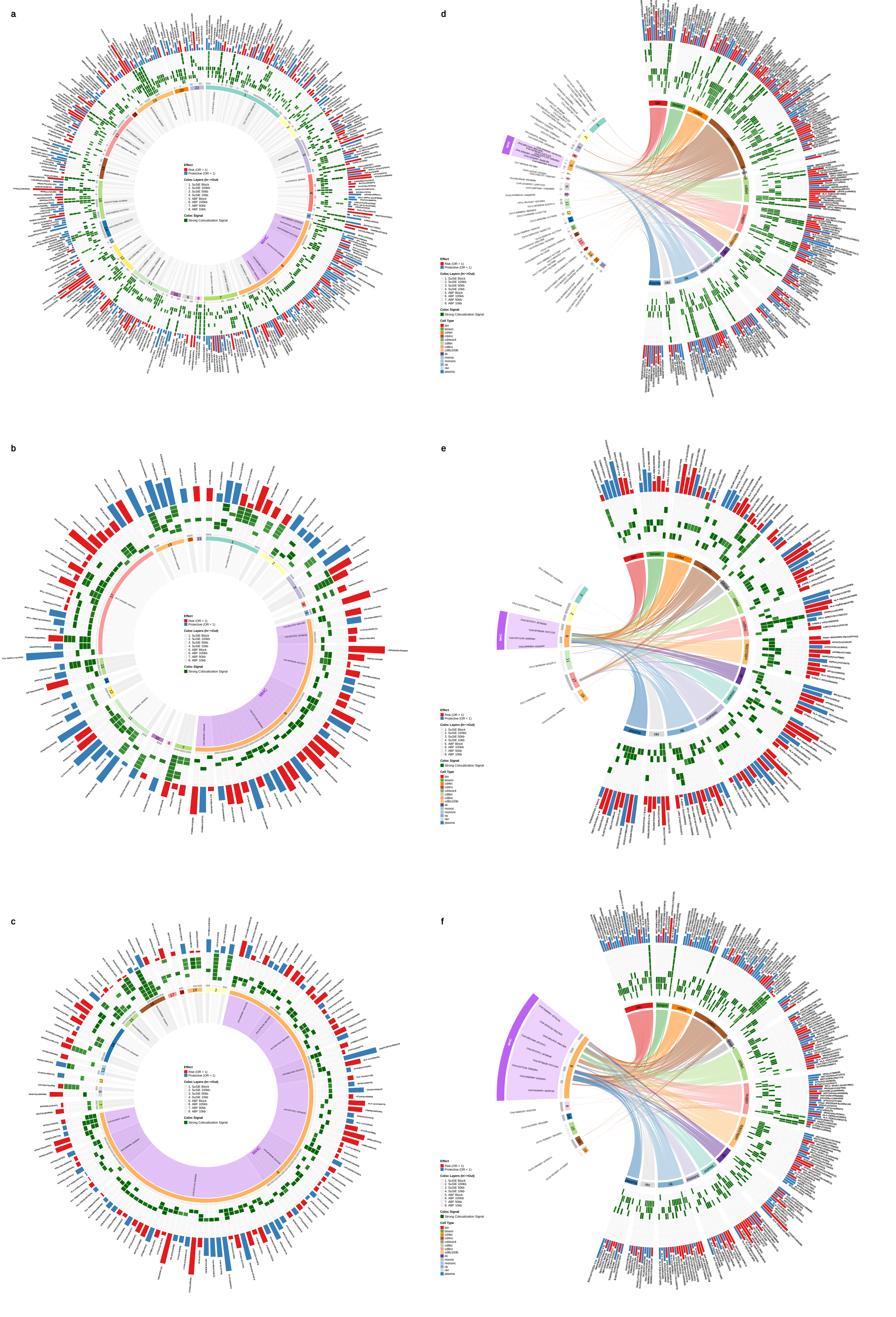

### Supplementary Figure 3

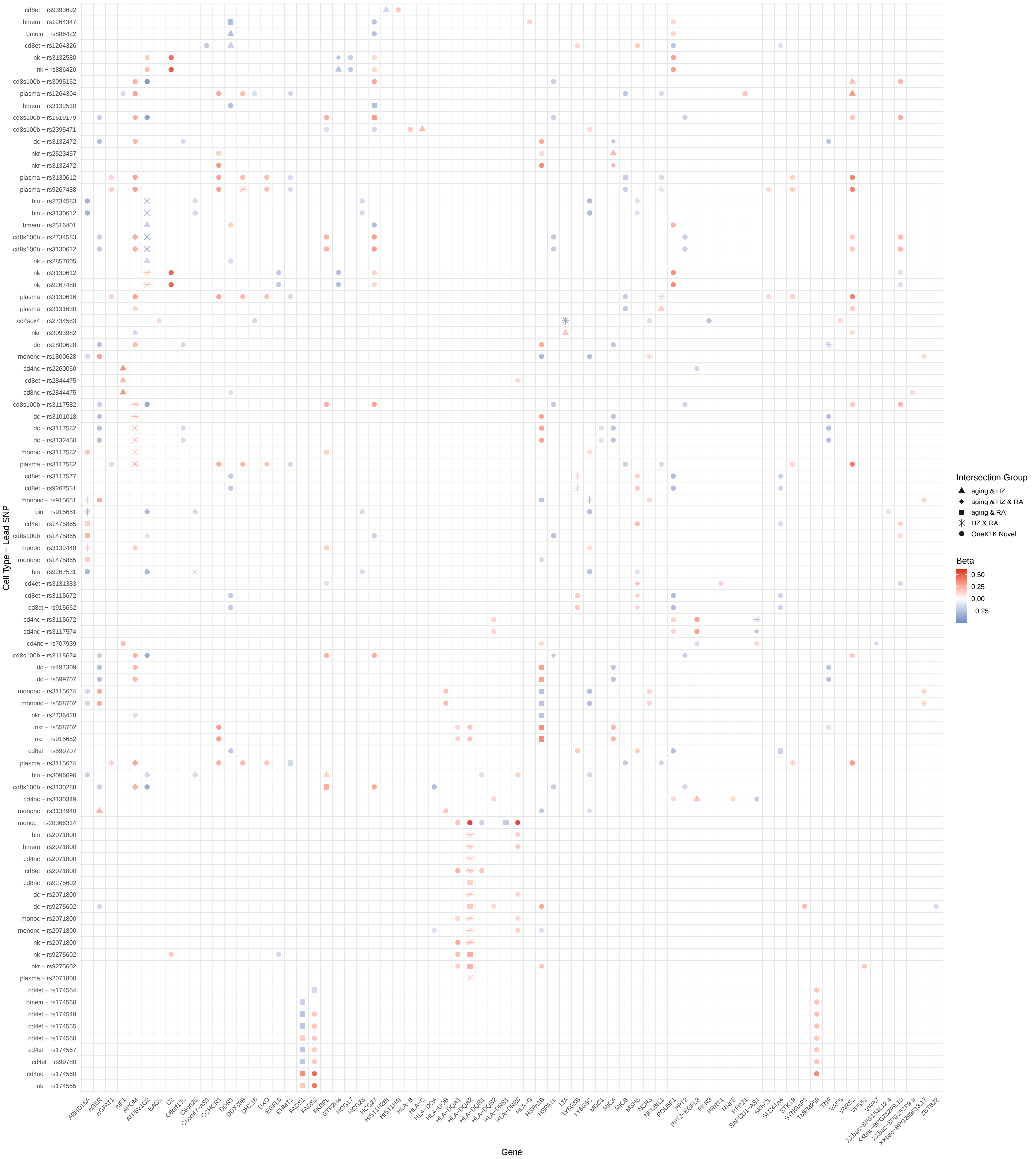

### Supplementary Figure 4

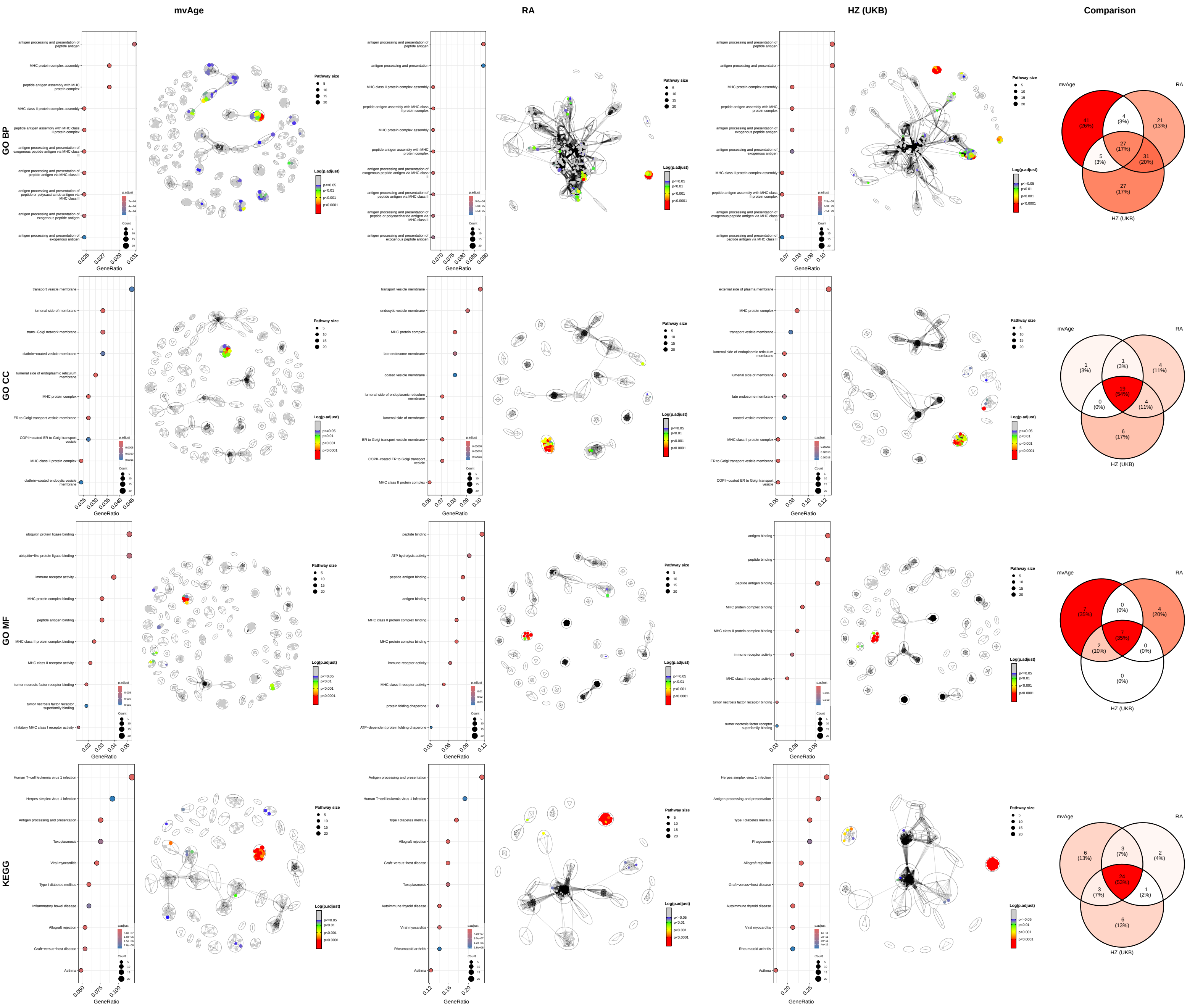

### Supplementary Figure 5

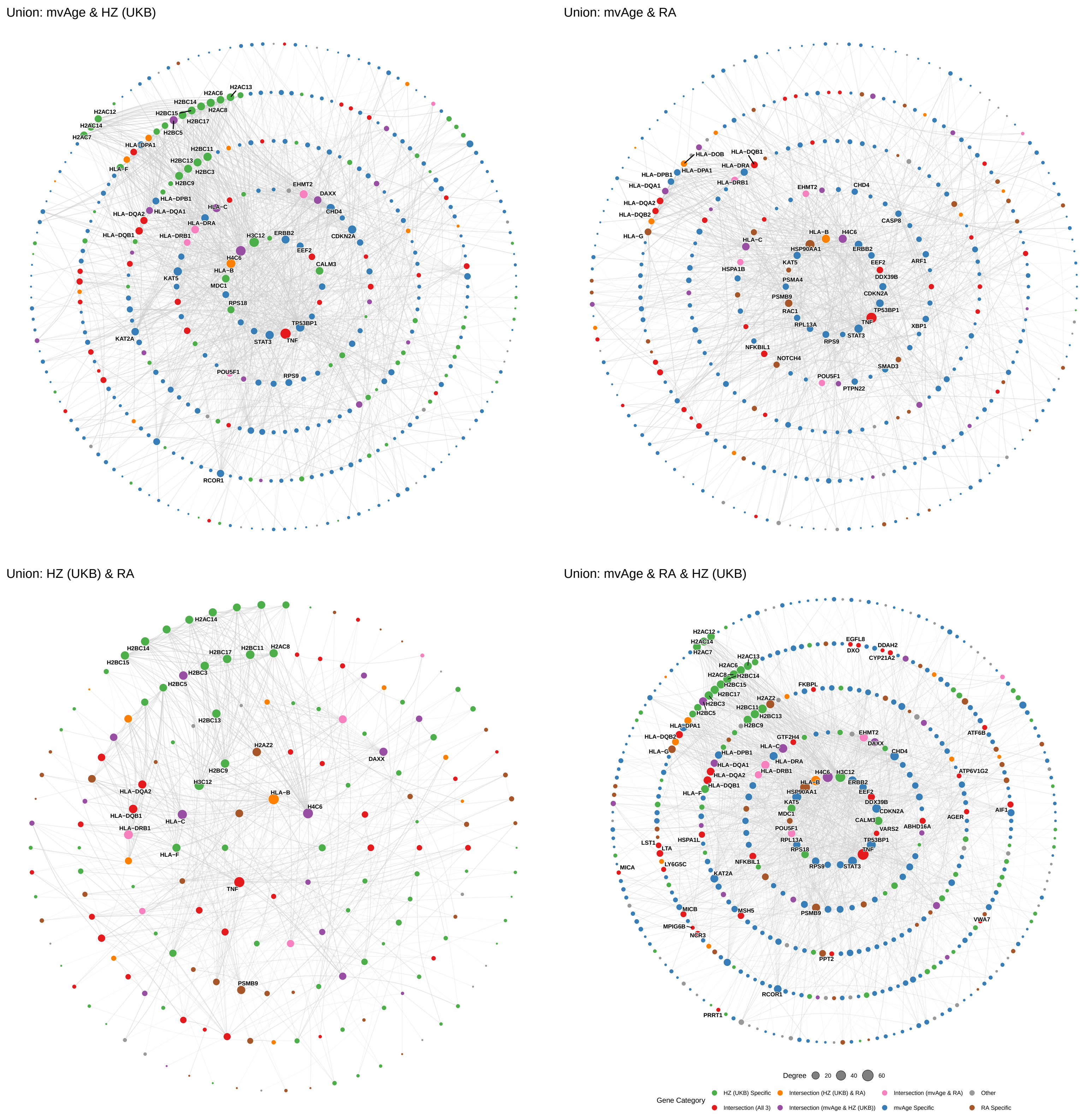

### Supplementary Figure 6

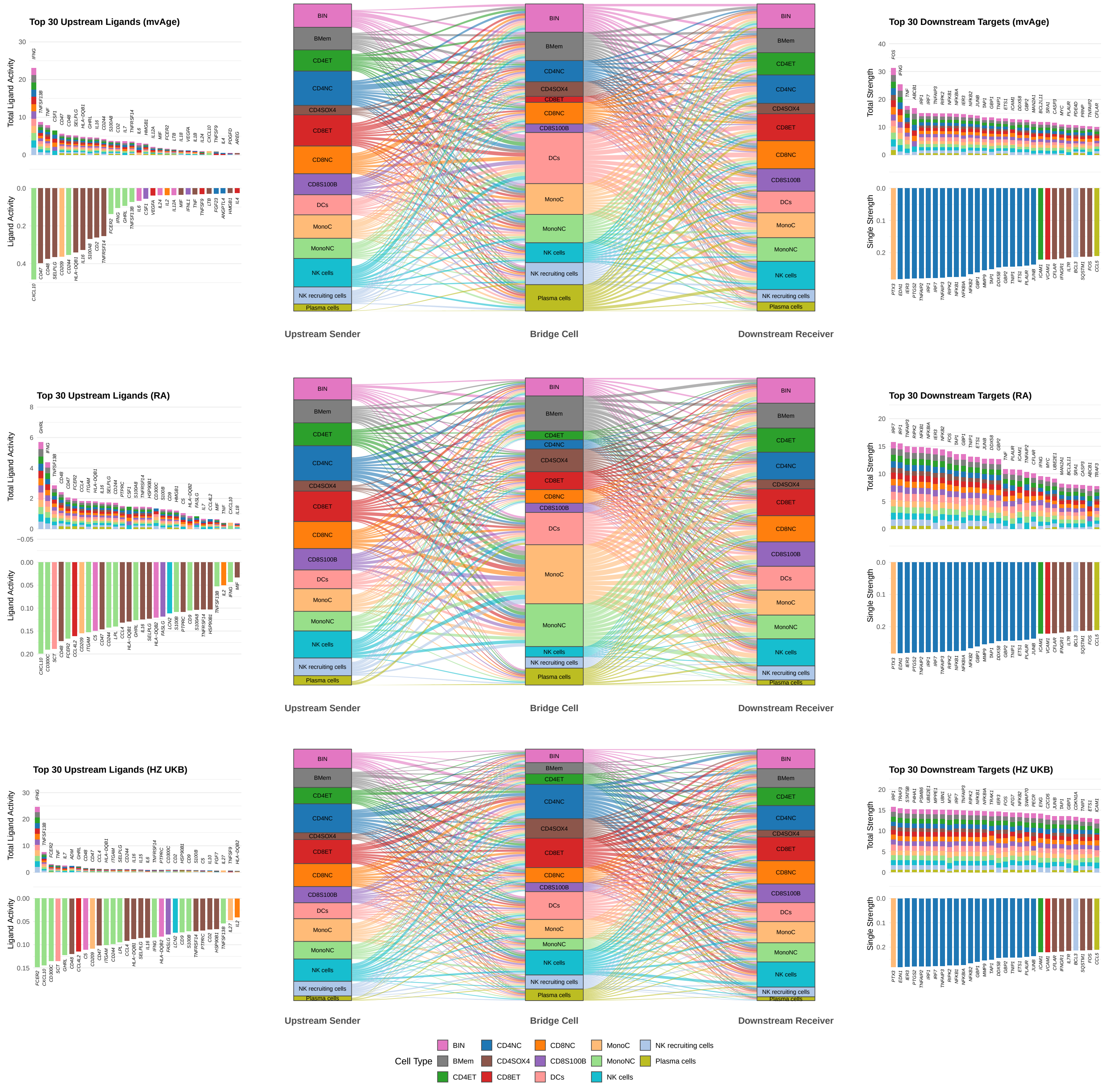

Supplementary Figure 7

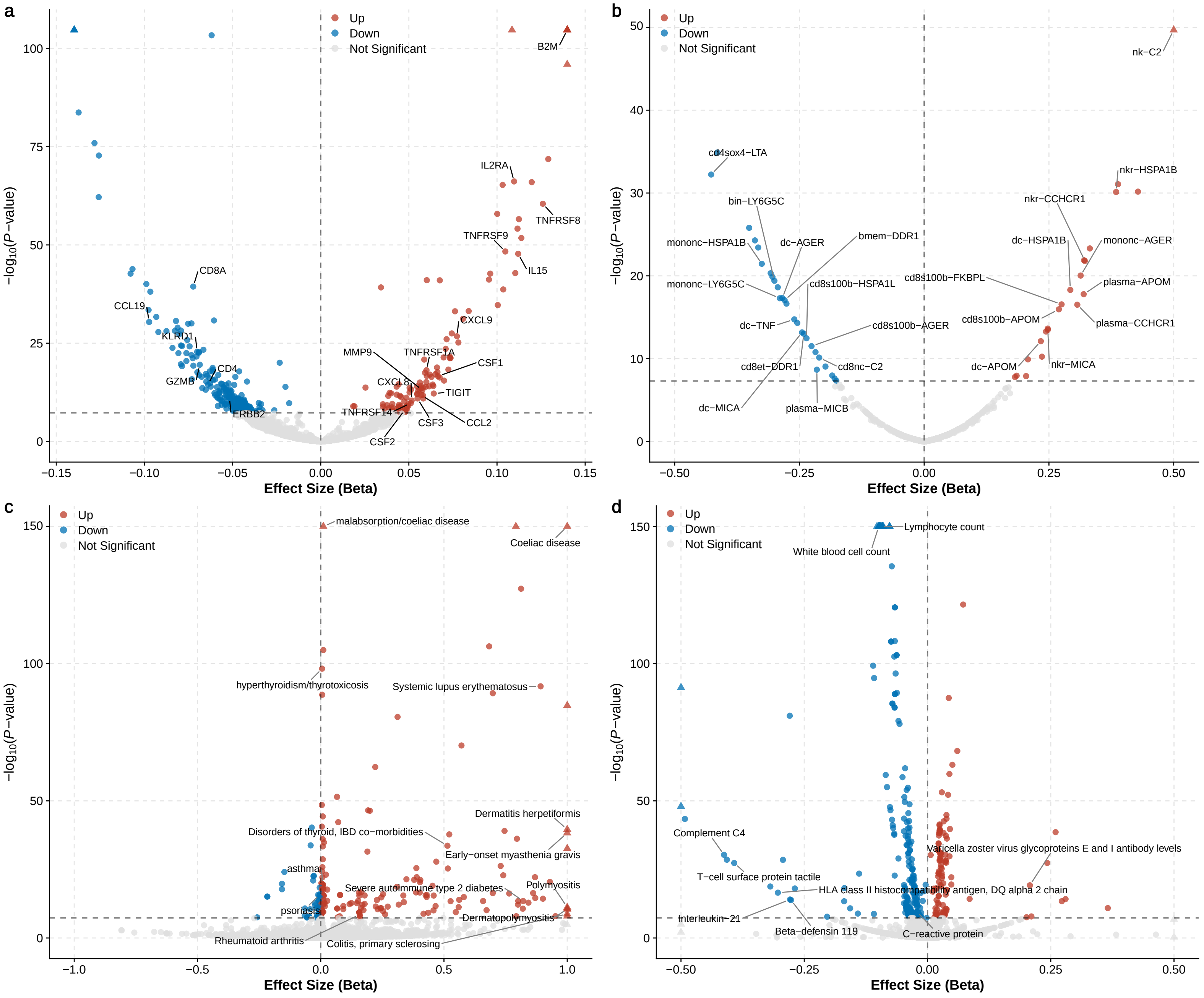

### Supplementary Figure 8

a

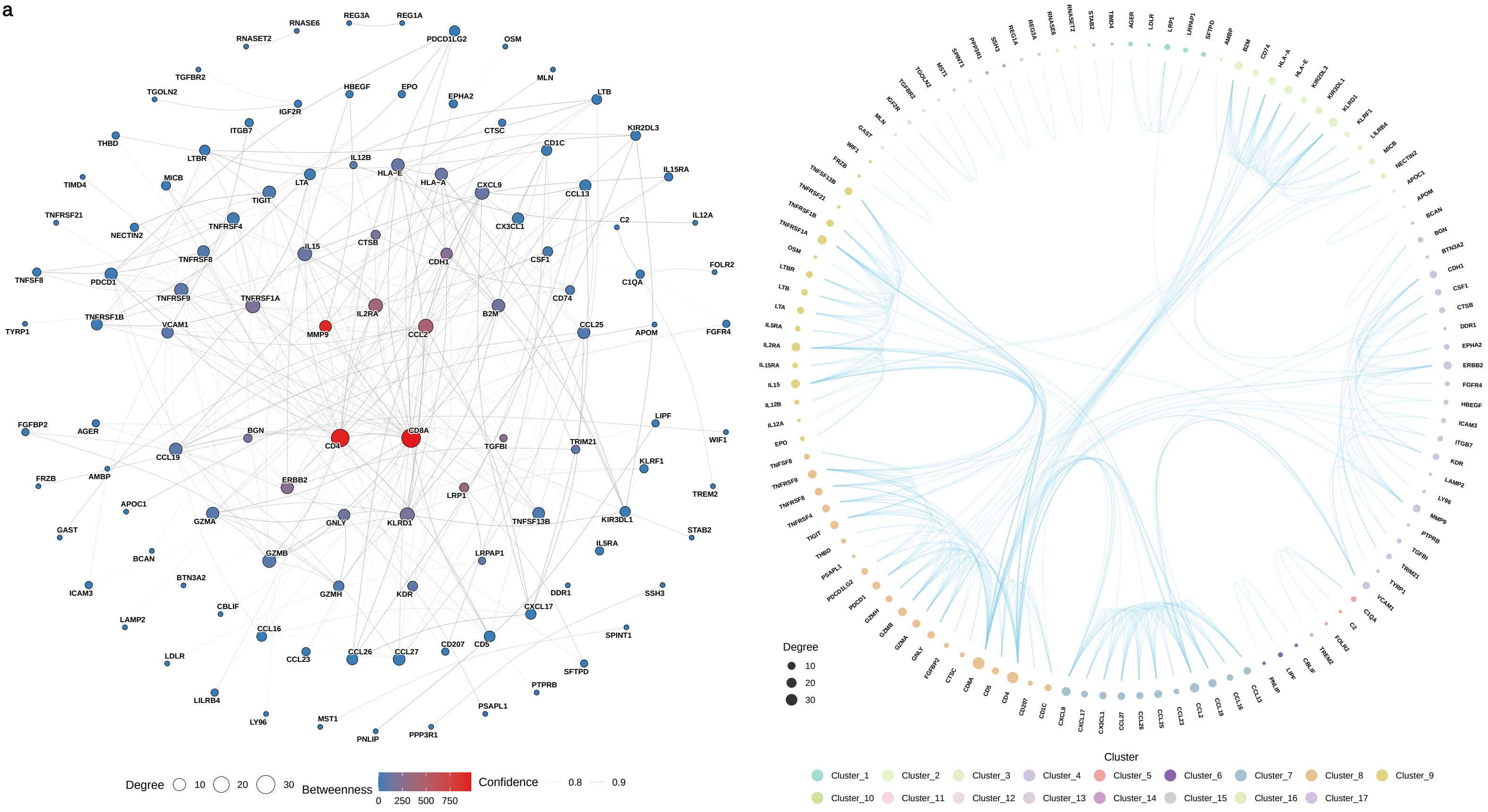

b

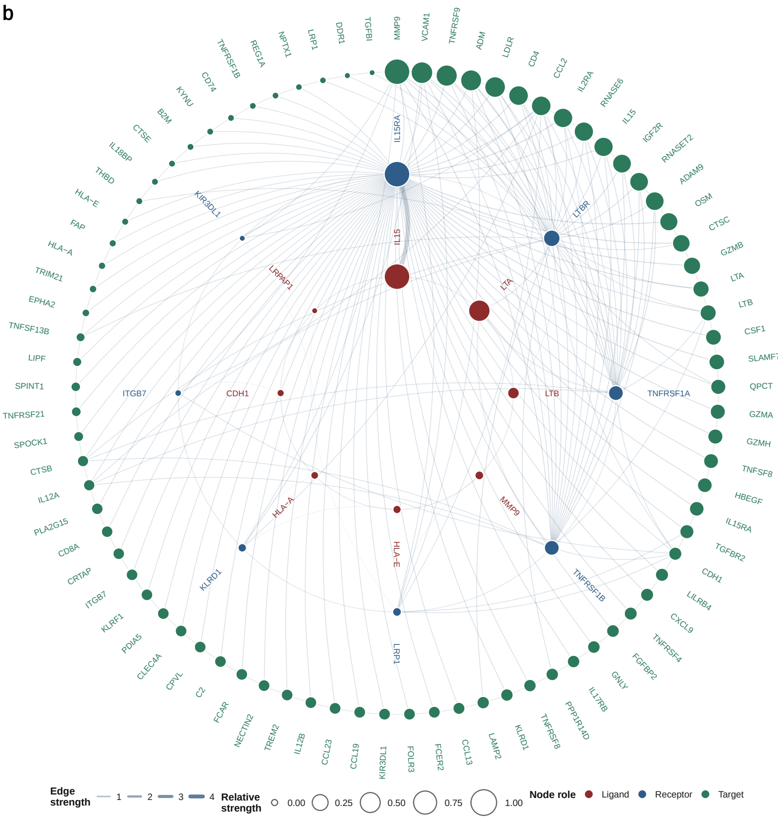

### Supplementary Figure 9

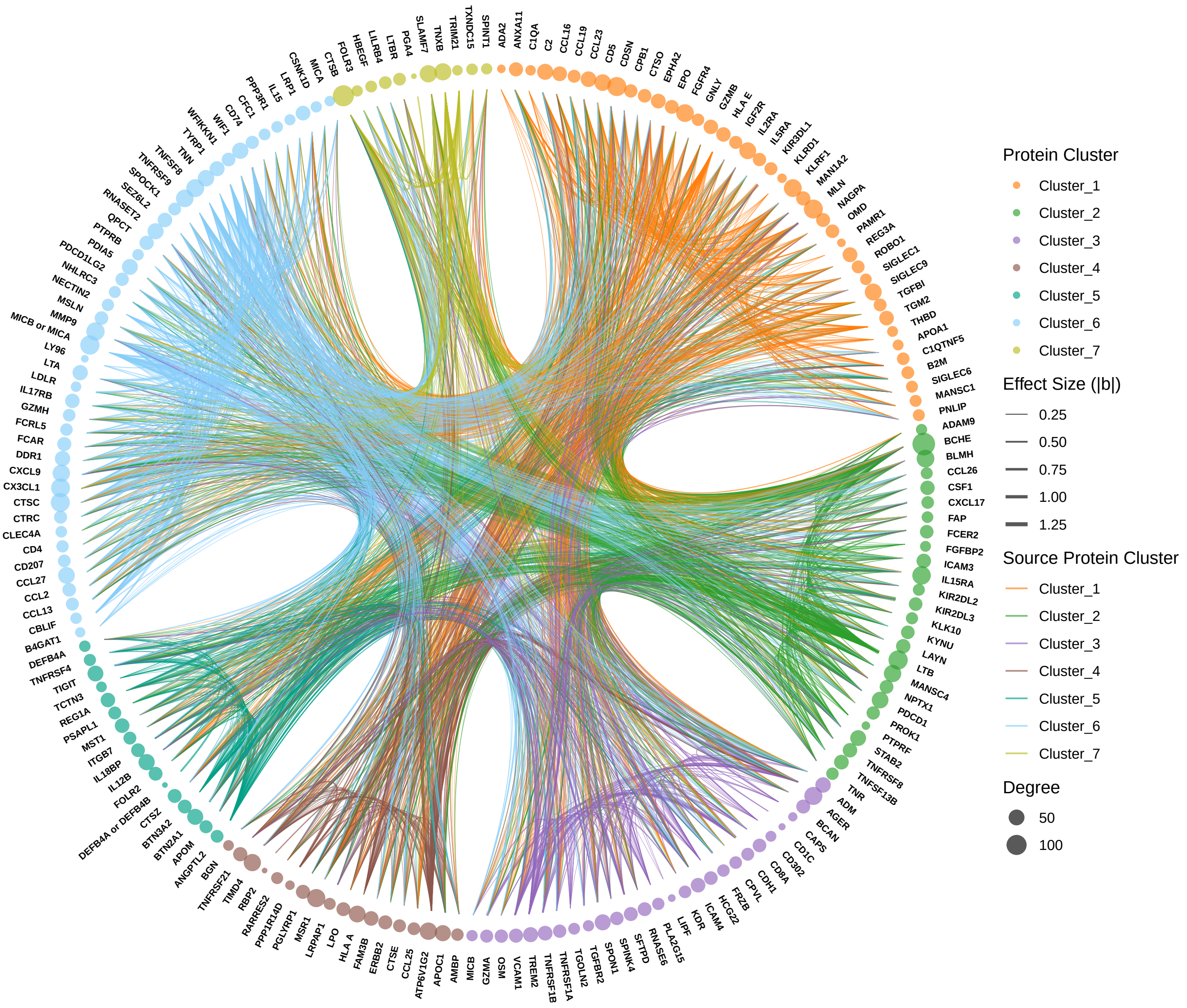

### Supplementary Figure 10

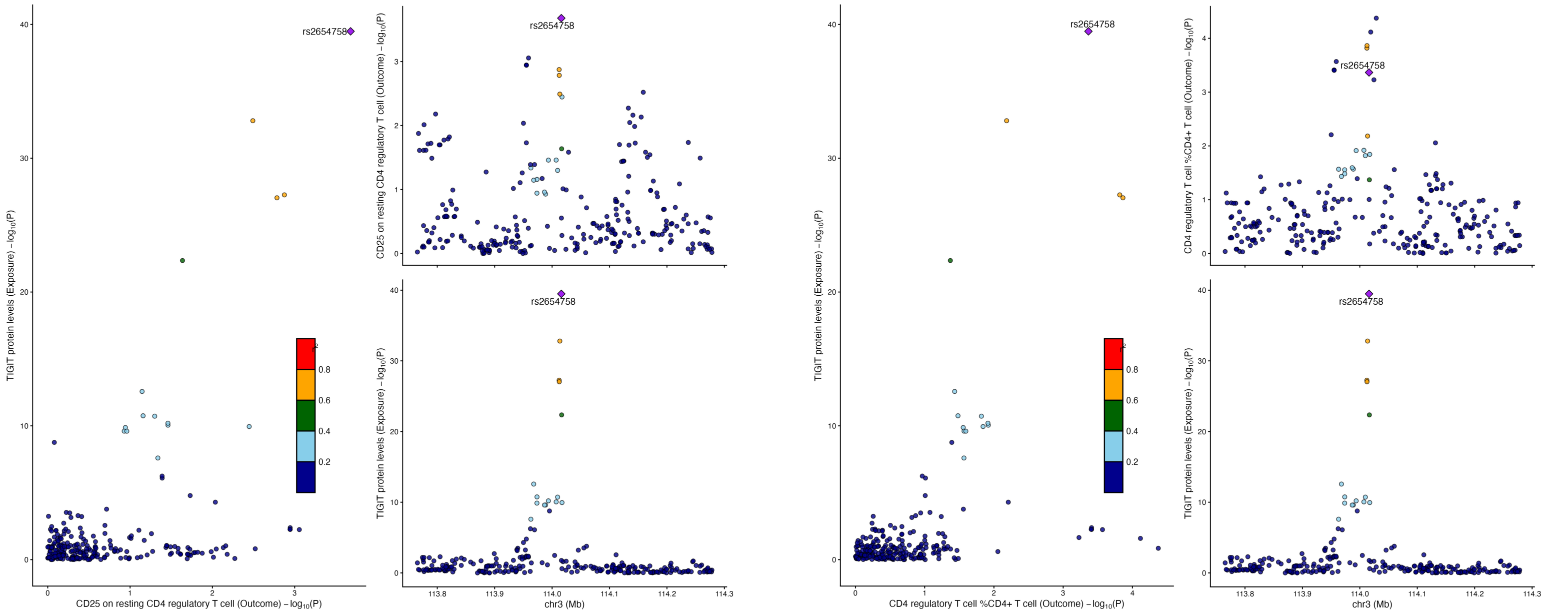

Supplementary Figure 11

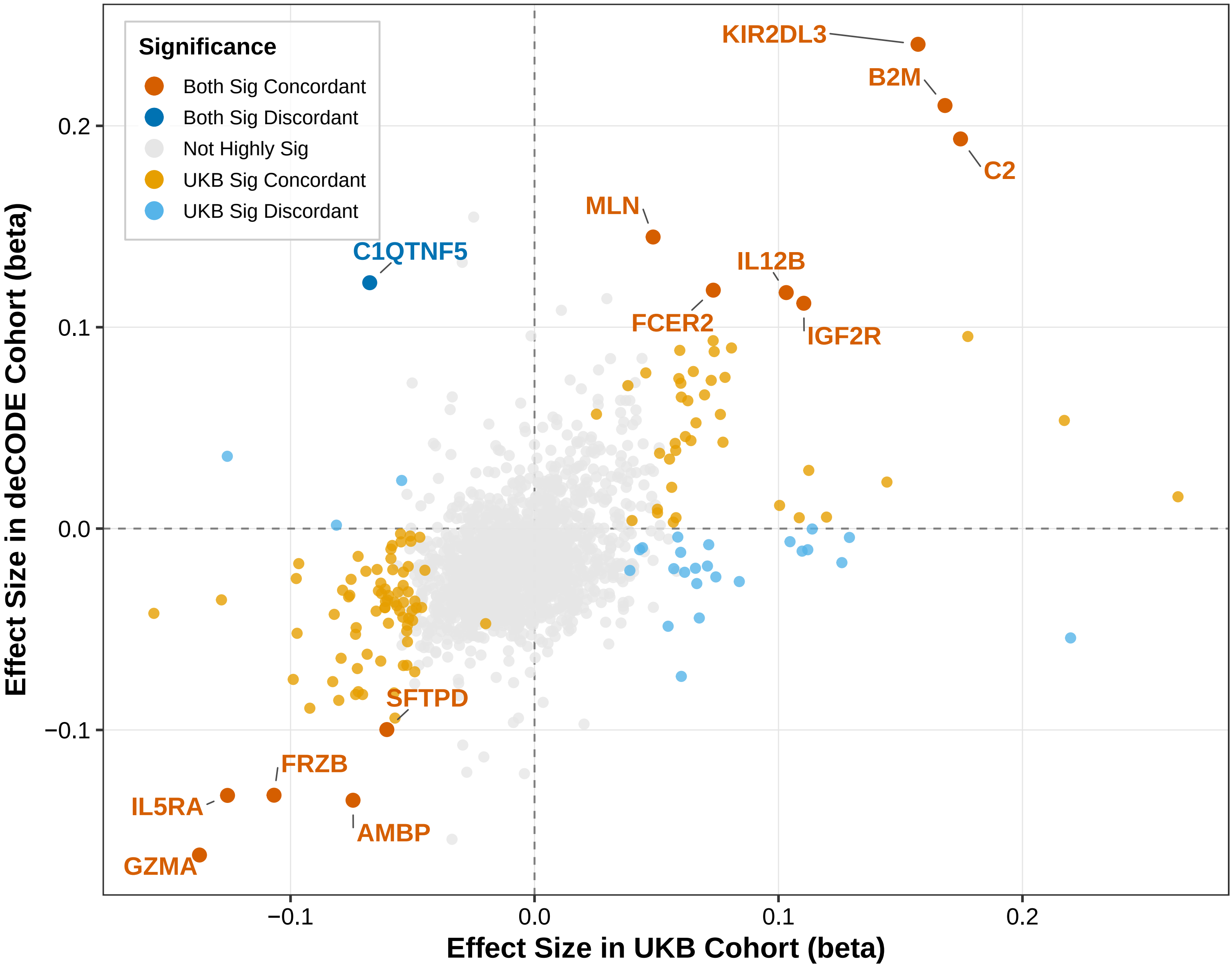

Supplementary Figure 12

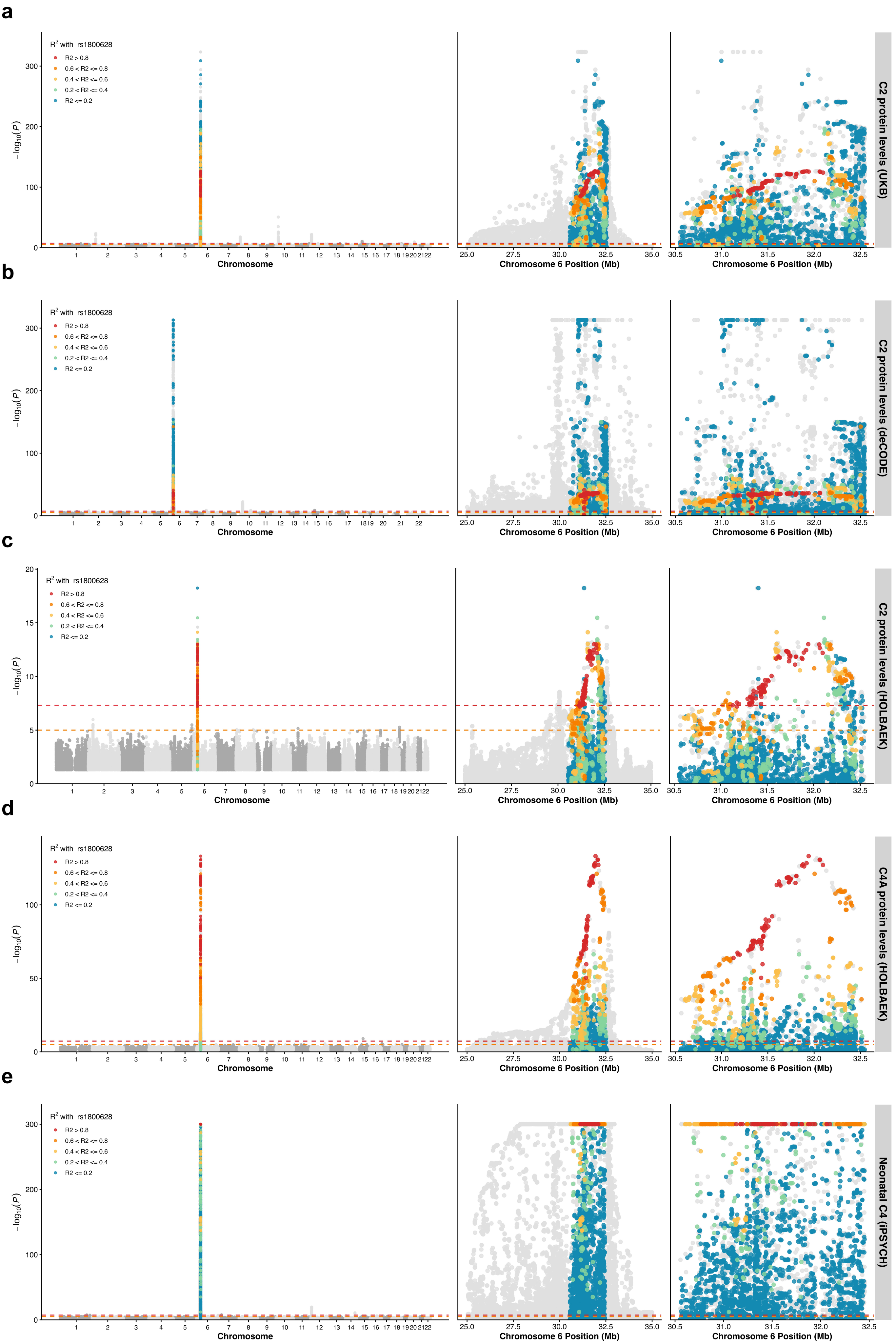
