## Supplementary File 1 for "A shared MHC immunogenetic signal connects aging, rheumatoid arthritis, and herpes zoster through chronic high inflammatory burden–compensatory immune tolerance dysregulation": plotly.min.js

```
/**
* plotly.js v2.32.0
* Copyright 2012-2024, Plotly, Inc.
* All rights reserved.
* Licensed under the MIT license
*/
/*! For license information please see plotly.min.js.LICENSE.txt */
!function(t,e){"object"==typeof exports&&"object"==typeof module?module.exports=e():"function"==typeof define&&define.amd?define([],e):"object"==typeof exports?exports.Plotly=e():t.Plotly=e()}(self,(function(){return function(){var t={79288:function(t,e,r){"use strict";var n=r(3400),i={"X,X div":'direction:ltr;font-family:"Open Sans",verdana,arial,sans-serif;margin:0;padding:0;',"X input,X button":'font-family:"Open Sans",verdana,arial,sans-serif;',"X input:focus,X button:focus":"outline:none;","X a":"text-decoration:none;","X a:hover":"text-decoration:none;","X .crisp":"shape-rendering:crispEdges;","X .user-select-none":"-webkit-user-select:none;-moz-user-select:none;-ms-user-select:none;-o-user-select:none;user-select:none;","X svg":"overflow:hidden;","X svg a":"fill:#447adb;","X svg a:hover":"fill:#3c6dc5;","X .main-svg":"position:absolute;top:0;left:0;pointer-events:none;","X .main-svg .draglayer":"pointer-events:all;","X .cursor-default":"cursor:default;","X .cursor-pointer":"cursor:pointer;","X .cursor-crosshair":"cursor:crosshair;","X .cursor-move":"cursor:move;","X .cursor-col-resize":"cursor:col-resize;","X .cursor-row-resize":"cursor:row-resize;","X .cursor-ns-resize":"cursor:ns-resize;","X .cursor-ew-resize":"cursor:ew-resize;","X .cursor-sw-resize":"cursor:sw-resize;","X .cursor-s-resize":"cursor:s-resize;","X .cursor-se-resize":"cursor:se-resize;","X .cursor-w-resize":"cursor:w-resize;","X .cursor-e-resize":"cursor:e-resize;","X .cursor-nw-resize":"cursor:nw-resize;","X .cursor-n-resize":"cursor:n-resize;","X .cursor-ne-resize":"cursor:ne-resize;","X .cursor-grab":"cursor:-webkit-grab;cursor:grab;","X .modebar":"position:absolute;top:2px;right:2px;","X .ease-bg":"-webkit-transition:background-color .3s ease 0s;-moz-transition:background-color .3s ease 0s;-ms-transition:background-color .3s ease 0s;-o-transition:background-color .3s ease 0s;transition:background-color .3s ease 0s;","X .modebar--hover>:not(.watermark)":"opacity:0;-webkit-transition:opacity .3s ease 0s;-moz-transition:opacity .3s ease 0s;-ms-transition:opacity .3s ease 0s;-o-transition:opacity .3s ease 0s;transition:opacity .3s ease 0s;","X:hover .modebar--hover .modebar-group":"opacity:1;","X .modebar-group":"float:left;display:inline-block;box-sizing:border-box;padding-left:8px;position:relative;vertical-align:middle;white-space:nowrap;","X .modebar-btn":"position:relative;font-size:16px;padding:3px 4px;height:22px;cursor:pointer;line-height:normal;box-sizing:border-box;","X .modebar-btn svg":"position:relative;top:2px;","X .modebar.vertical":"display:flex;flex-direction:column;flex-wrap:wrap;align-content:flex-end;max-height:100%;","X .modebar.vertical svg":"top:-1px;","X .modebar.vertical .modebar-group":"display:block;float:none;padding-left:0px;padding-bottom:8px;","X .modebar.vertical .modebar-group .modebar-btn":"display:block;text-align:center;","X [data-title]:before,X [data-title]:after":"position:absolute;-webkit-transform:translate3d(0, 0, 0);-moz-transform:translate3d(0, 0, 0);-ms-transform:translate3d(0, 0, 0);-o-transform:translate3d(0, 0, 0);transform:translate3d(0, 0, 0);display:none;opacity:0;z-index:1001;pointer-events:none;top:110%;right:50%;","X [data-title]:hover:before,X [data-title]:hover:after":"display:block;opacity:1;","X [data-title]:before":'content:"";position:absolute;background:rgba(0,0,0,0);border:6px solid rgba(0,0,0,0);z-index:1002;margin-top:-12px;border-bottom-color:#69738a;margin-right:-6px;',"X [data-title]:after":"content:attr(data-title);background:#69738a;color:#fff;padding:8px 10px;font-size:12px;line-height:12px;white-space:nowrap;margin-right:-18px;border-radius:2px;","X .vertical [data-title]:before,X .vertical [data-title]:after":"top:0%;right:200%;","X .vertical [data-title]:before":"border:6px solid rgba(0,0,0,0);border-left-color:#69738a;margin-top:8px;margin-right:-30px;",Y:'font-family:"Open Sans",verdana,arial,sans-serif;position:fixed;top:50px;right:20px;z-index:10000;font-size:10pt;max-width:180px;',"Y p":"margin:0;","Y .notifier-note":"min-width:180px;max-width:250px;border:1px solid #fff;z-index:3000;margin:0;background-color:#8c97af;background-color:rgba(140,151,175,.9);color:#fff;padding:10px;overflow-wrap:break-word;word-wrap:break-word;-ms-hyphens:auto;-webkit-hyphens:auto;hyphens:auto;","Y .notifier-close":"color:#fff;opacity:.8;float:right;padding:0 5px;background:none;border:none;font-size:20px;font-weight:bold;line-height:20px;","Y .notifier-close:hover":"color:#444;text-decoration:none;cursor:pointer;"};for(var a in i){var o=a.replace(/^,/," ,").replace(/X/g,".js-plotly-plot .plotly").replace(/Y/g,".plotly-notifier");n.addStyleRule(o,i[a])}},86712:function(t,e,r){"use strict";t.exports=r(84224)},37240:function(t,e,r){"use strict";t.exports=r(51132)},29744:function(t,e,r){"use strict";t.exports=r(94456)},29352:function(t,e,r){"use strict";t.exports=r(67244)},96144:function(t,e,r){"use strict";t.exports=r(97776)},53219:function(t,e,r){"use strict";t.exports=r(61712)},4624:function(t,e,r){"use strict";t.exports=r(95856)},54543:function(t,e,r){"use strict";t.exports=r(54272)},45e3:function(t,e,r){"use strict";t.exports=r(85404)},62300:function(t,e,r){"use strict";t.exports=r(26048)},6920:function(t,e,r){"use strict";t.exports=r(66240)},10264:function(t,e,r){"use strict";t.exports=r(40448)},32016:function(t,e,r){"use strict";t.exports=r(64884)},27528:function(t,e,r){"use strict";t.exports=r(15088)},75556:function(t,e,r){"use strict";t.exports=r(76744)},39204:function(t,e,r){"use strict";t.exports=r(94704)},73996:function(t,e,r){"use strict";t.exports=r(62396)},16489:function(t,e,r){"use strict";t.exports=r(32028)},5e3:function(t,e,r){"use strict";t.exports=r(81932)},77280:function(t,e,r){"use strict";t.exports=r(45536)},33992:function(t,e,r){"use strict";t.exports=r(42600)},17600:function(t,e,r){"use strict";t.exports=r(21536)},49116:function(t,e,r){"use strict";t.exports=r(65664)},46808:function(t,e,r){"use strict";t.exports=r(29044)},36168:function(t,e,r){"use strict";t.exports=r(48928)},13792:function(t,e,r){"use strict";var n=r(32016);n.register([r(37240),r(29352),r(5e3),r(33992),r(17600),r(49116),r(6920),r(67484),r(79440),r(39204),r(83096),r(36168),r(20260),r(63560),r(65832),r(46808),r(73996),r(48824),r(89904),r(25120),r(13752),r(4340),r(62300),r(29800),r(8363),r(54543),r(86636),r(42192),r(32140),r(77280),r(89296),r(56816),r(70192),r(45e3),r(27528),r(84764),r(3920),r(50248),r(4624),r(69967),r(10264),r(86152),r(53219),r(81604),r(63796),r(29744),r(89336),r(86712),r(75556),r(16489),r(97312),r(96144)]),t.exports=n},3920:function(t,e,r){"use strict";t.exports=r(43480)},25120:function(t,e,r){"use strict";t.exports=r(6296)},4340:function(t,e,r){"use strict";t.exports=r(7404)},86152:function(t,e,r){"use strict";t.exports=r(65456)},56816:function(t,e,r){"use strict";t.exports=r(22020)},89296:function(t,e,r){"use strict";t.exports=r(29928)},20260:function(t,e,r){"use strict";t.exports=r(75792)},32140:function(t,e,r){"use strict";t.exports=r(156)},84764:function(t,e,r){"use strict";t.exports=r(45499)},48824:function(t,e,r){"use strict";t.exports=r(3296)},69967:function(t,e,r){"use strict";t.exports=r(4184)},8363:function(t,e,r){"use strict";t.exports=r(36952)},86636:function(t,e,r){"use strict";t.exports=r(38983)},70192:function(t,e,r){"use strict";t.exports=r(11572)},81604:function(t,e,r){"use strict";t.exports=r(76924)},63796:function(t,e,r){"use strict";t.exports=r(62944)},89336:function(t,e,r){"use strict";t.exports=r(95443)},67484:function(t,e,r){"use strict";t.exports=r(34864)},97312:function(t,e,r){"use strict";t.exports=r(76272)},42192:function(t,e,r){"use strict";t.exports=r(97924)},29800:function(t,e,r){"use strict";t.exports=r(15436)},63560:function(t,e,r){"use strict";t.exports=r(5621)},89904:function(t,e,r){"use strict";t.exports=r(91304)},50248:function(t,e,r){"use strict";t.exports=r(41724)},65832:function(t,e,r){"use strict";t.exports=r(31991)},79440:function(t,e,r){"use strict";t.exports=r(22869)},13752:function(t,e,r){"use strict";t.exports=r(67776)},83096:function(t,e,r){"use strict";t.exports=r(95952)},72196:function(t){"use strict";t.exports=[{path:"",backoff:0},{path:"M-2.4,-3V3L0.6,0Z",backoff:.6},{path:"M-3.7,-2.5V2.5L1.3,0Z",backoff:1.3},{path:"M-4.45,-3L-1.65,-0.2V0.2L-4.45,3L1.55,0Z",backoff:1.55},{path:"M-2.2,-2.2L-0.2,-0.2V0.2L-2.2,2.2L-1.4,3L1.6,0L-1.4,-3Z",backoff:1.6},{path:"M-4.4,-2.1L-0.6,-0.2V0.2L-4.4,2.1L-4,3L2,0L-4,-3Z",backoff:2},{path:"M2,0A2,2 0 1,1 0,-2A2,2 0 0,1 2,0Z",backoff:0,noRotate:!0},{path:"M2,2V-2H-2V2Z",backoff:0,noRotate:!0}]},13916:function(t,e,r){"use strict";var n=r(72196),i=r(25376),a=r(33816),o=r(31780).templatedArray;r(36208),t.exports=o("annotation",{visible:{valType:"boolean",dflt:!0,editType:"calc+arraydraw"},text:{valType:"string",editType:"calc+arraydraw"},textangle:{valType:"angle",dflt:0,editType:"calc+arraydraw"},font:i({editType:"calc+arraydraw",colorEditType:"arraydraw"}),width:{valType:"number",min:1,dflt:null,editType:"calc+arraydraw"},height:{valType:"number",min:1,dflt:null,editType:"calc+arraydraw"},opacity:{valType:"number",min:0,max:1,dflt:1,editType:"arraydraw"},align:{valType:"enumerated",values:["left","center","right"],dflt:"center",editType:"arraydraw"},valign:{valType:"enumerated",values:["top","middle","bottom"],dflt:"middle",editType:"arraydraw"},bgcolor:{valType:"color",dflt:"rgba(0,0,0,0)",editType:"arraydraw"},bordercolor:{valType:"color",dflt:"rgba(0,0,0,0)",editType:"arraydraw"},borderpad:{valType:"number",min:0,dflt:1,editType:"calc+arraydraw"},borderwidth:{valType:"number",min:0,dflt:1,editType:"calc+arraydraw"},showarrow:{valType:"boolean",dflt:!0,editType:"calc+arraydraw"},arrowcolor:{valType:"color",editType:"arraydraw"},arrowhead:{valType:"integer",min:0,max:n.length,dflt:1,editType:"arraydraw"},startarrowhead:{valType:"integer",min:0,max:n.length,dflt:1,editType:"arraydraw"},arrowside:{valType:"flaglist",flags:["end","start"],extras:["none"],dflt:"end",editType:"arraydraw"},arrowsize:{valType:"number",min:.3,dflt:1,editType:"calc+arraydraw"},startarrowsize:{valType:"number",min:.3,dflt:1,editType:"calc+arraydraw"},arrowwidth:{valType:"number",min:.1,editType:"calc+arraydraw"},standoff:{valType:"number",min:0,dflt:0,editType:"calc+arraydraw"},startstandoff:{valType:"number",min:0,dflt:0,editType:"calc+arraydraw"},ax:{valType:"any",editType:"calc+arraydraw"},ay:{valType:"any",editType:"calc+arraydraw"},axref:{valType:"enumerated",dflt:"pixel",values:["pixel",a.idRegex.x.toString()],editType:"calc"},ayref:{valType:"enumerated",dflt:"pixel",values:["pixel",a.idRegex.y.toString()],editType:"calc"},xref:{valType:"enumerated",values:["paper",a.idRegex.x.toString()],editType:"calc"},x:{valType:"any",editType:"calc+arraydraw"},xanchor:{valType:"enumerated",values:["auto","left","center","right"],dflt:"auto",editType:"calc+arraydraw"},xshift:{valType:"number",dflt:0,editType:"calc+arraydraw"},yref:{valType:"enumerated",values:["paper",a.idRegex.y.toString()],editType:"calc"},y:{valType:"any",editType:"calc+arraydraw"},yanchor:{valType:"enumerated",values:["auto","top","middle","bottom"],dflt:"auto",editType:"calc+arraydraw"},yshift:{valType:"number",dflt:0,editType:"calc+arraydraw"},clicktoshow:{valType:"enumerated",values:[!1,"onoff","onout"],dflt:!1,editType:"arraydraw"},xclick:{valType:"any",editType:"arraydraw"},yclick:{valType:"any",editType:"arraydraw"},hovertext:{valType:"string",editType:"arraydraw"},hoverlabel:{bgcolor:{valType:"color",editType:"arraydraw"},bordercolor:{valType:"color",editType:"arraydraw"},font:i({editType:"arraydraw"}),editType:"arraydraw"},captureevents:{valType:"boolean",editType:"arraydraw"},editType:"calc",_deprecated:{ref:{valType:"string",editType:"calc"}}})},90272:function(t,e,r){"use strict";var n=r(3400),i=r(54460),a=r(23816).draw;function o(t){var e=t._fullLayout;n.filterVisible(e.annotations).forEach((function(e){var r=i.getFromId(t,e.xref),n=i.getFromId(t,e.yref),a=i.getRefType(e.xref),o=i.getRefType(e.yref);e._extremes={},"range"===a&&s(e,r),"range"===o&&s(e,n)}))}function s(t,e){var r,n=e._id,a=n.charAt(0),o=t[a],s=t["a"+a],l=t[a+"ref"],u=t["a"+a+"ref"],c=t["_"+a+"padplus"],f=t["_"+a+"padminus"],h={x:1,y:-1}[a]*t[a+"shift"],p=3*t.arrowsize*t.arrowwidth||0,d=p+h,v=p-h,g=3*t.startarrowsize*t.arrowwidth||0,y=g+h,m=g-h;if(u===l){var x=i.findExtremes(e,[e.r2c(o)],{ppadplus:d,ppadminus:v}),b=i.findExtremes(e,[e.r2c(s)],{ppadplus:Math.max(c,y),ppadminus:Math.max(f,m)});r={min:[x.min[0],b.min[0]],max:[x.max[0],b.max[0]]}}else y=s?y+s:y,m=s?m-s:m,r=i.findExtremes(e,[e.r2c(o)],{ppadplus:Math.max(c,d,y),ppadminus:Math.max(f,v,m)});t._extremes[n]=r}t.exports=function(t){var e=t._fullLayout;if(n.filterVisible(e.annotations).length&&t._fullData.length)return n.syncOrAsync([a,o],t)}},42300:function(t,e,r){"use strict";var n=r(3400),i=r(24040),a=r(31780).arrayEditor;function o(t,e){var r,n,i,a,o,l,u,c=t._fullLayout.annotations,f=[],h=[],p=[],d=(e||[]).length;for(r=0;r<c.length;r++)if(a=(i=c[r]).clicktoshow){for(n=0;n<d;n++)if(l=(o=e[n]).xaxis,u=o.yaxis,l._id===i.xref&&u._id===i.yref&&l.d2r(o.x)===s(i._xclick,l)&&u.d2r(o.y)===s(i._yclick,u)){(i.visible?"onout"===a?h:p:f).push(r);break}n===d&&i.visible&&"onout"===a&&h.push(r)}return{on:f,off:h,explicitOff:p}}function s(t,e){return"log"===e.type?e.l2r(t):e.d2r(t)}t.exports={hasClickToShow:function(t,e){var r=o(t,e);return r.on.length>0||r.explicitOff.length>0},onClick:function(t,e){var r,s,l=o(t,e),u=l.on,c=l.off.concat(l.explicitOff),f={},h=t._fullLayout.annotations;if(u.length||c.length){for(r=0;r<u.length;r++)(s=a(t.layout,"annotations",h[u[r]])).modifyItem("visible",!0),n.extendFlat(f,s.getUpdateObj());for(r=0;r<c.length;r++)(s=a(t.layout,"annotations",h[c[r]])).modifyItem("visible",!1),n.extendFlat(f,s.getUpdateObj());return i.call("update",t,{},f)}}}},87192:function(t,e,r){"use strict";var n=r(3400),i=r(76308);t.exports=function(t,e,r,a){a("opacity");var o=a("bgcolor"),s=a("bordercolor"),l=i.opacity(s);a("borderpad");var u=a("borderwidth"),c=a("showarrow");if(a("text",c?" ":r._dfltTitle.annotation),a("textangle"),n.coerceFont(a,"font",r.font),a("width"),a("align"),a("height")&&a("valign"),c){var f,h,p=a("arrowside");-1!==p.indexOf("end")&&(f=a("arrowhead"),h=a("arrowsize")),-1!==p.indexOf("start")&&(a("startarrowhead",f),a("startarrowsize",h)),a("arrowcolor",l?e.bordercolor:i.defaultLine),a("arrowwidth",2*(l&&u||1)),a("standoff"),a("startstandoff")}var d=a("hovertext"),v=r.hoverlabel||{};if(d){var g=a("hoverlabel.bgcolor",v.bgcolor||(i.opacity(o)?i.rgb(o):i.defaultLine)),y=a("hoverlabel.bordercolor",v.bordercolor||i.contrast(g)),m=n.extendFlat({},v.font);m.color||(m.color=y),n.coerceFont(a,"hoverlabel.font",m)}a("captureevents",!!d)}},26828:function(t,e,r){"use strict";var n=r(38248),i=r(36896);t.exports=function(t,e,r,a){e=e||{};var o="log"===r&&"linear"===e.type,s="linear"===r&&"log"===e.type;if(o||s)for(var l,u,c=t._fullLayout.annotations,f=e._id.charAt(0),h=0;h<c.length;h++)l=c[h],u="annotations["+h+"].",l[f+"ref"]===e._id&&p(f),l["a"+f+"ref"]===e._id&&p("a"+f);function p(t){var r=l[t],s=null;s=o?i(r,e.range):Math.pow(10,r),n(s)||(s=null),a(u+t,s)}}},45216:function(t,e,r){"use strict";var n=r(3400),i=r(54460),a=r(51272),o=r(87192),s=r(13916);function l(t,e,r){function a(r,i){return n.coerce(t,e,s,r,i)}var l=a("visible"),u=a("clicktoshow");if(l||u){o(t,e,r,a);for(var c=e.showarrow,f=["x","y"],h=[-10,-30],p={_fullLayout:r},d=0;d<2;d++){var v=f[d],g=i.coerceRef(t,e,p,v,"","paper");if("paper"!==g&&i.getFromId(p,g)._annIndices.push(e._index),i.coercePosition(e,p,a,g,v,.5),c){var y="a"+v,m=i.coerceRef(t,e,p,y,"pixel",["pixel","paper"]);"pixel"!==m&&m!==g&&(m=e[y]="pixel");var x="pixel"===m?h[d]:.4;i.coercePosition(e,p,a,m,y,x)}a(v+"anchor"),a(v+"shift")}if(n.noneOrAll(t,e,["x","y"]),c&&n.noneOrAll(t,e,["ax","ay"]),u){var b=a("xclick"),_=a("yclick");e._xclick=void 0===b?e.x:i.cleanPosition(b,p,e.xref),e._yclick=void 0===_?e.y:i.cleanPosition(_,p,e.yref)}}}t.exports=function(t,e){a(t,e,{name:"annotations",handleItemDefaults:l})}},23816:function(t,e,r){"use strict";var n=r(33428),i=r(24040),a=r(7316),o=r(3400),s=o.strTranslate,l=r(54460),u=r(76308),c=r(43616),f=r(93024),h=r(72736),p=r(93972),d=r(86476),v=r(31780).arrayEditor,g=r(33652);function y(t,e){var r=t._fullLayout.annotations[e]||{},n=l.getFromId(t,r.xref),i=l.getFromId(t,r.yref);n&&n.setScale(),i&&i.setScale(),x(t,r,e,!1,n,i)}function m(t,e,r,n,i){var a=i[r],o=i[r+"ref"],s=-1!==r.indexOf("y"),u="domain"===l.getRefType(o),c=s?n.h:n.w;return t?u?a+(s?-e:e)/t._length:t.p2r(t.r2p(a)+e):a+(s?-e:e)/c}function x(t,e,r,a,y,x){var b,_,w=t._fullLayout,T=t._fullLayout._size,k=t._context.edits;a?(b="annotation-"+a,_=a+".annotations"):(b="annotation",_="annotations");var A=v(t.layout,_,e),M=A.modifyBase,S=A.modifyItem,E=A.getUpdateObj;w._infolayer.selectAll("."+b+'[data-index="'+r+'"]').remove();var L="clip"+w._uid+"_ann"+r;if(e._input&&!1!==e.visible){var C={x:{},y:{}},P=+e.textangle||0,O=w._infolayer.append("g").classed(b,!0).attr("data-index",String(r)).style("opacity",e.opacity),I=O.append("g").classed("annotation-text-g",!0),z=k[e.showarrow?"annotationTail":"annotationPosition"],D=e.captureevents||k.annotationText||z,R=I.append("g").style("pointer-events",D?"all":null).call(p,"pointer").on("click",(function(){t._dragging=!1,t.emit("plotly_clickannotation",W(n.event))}));e.hovertext&&R.on("mouseover",(function(){var r=e.hoverlabel,n=r.font,i=this.getBoundingClientRect(),a=t.getBoundingClientRect();f.loneHover({x0:i.left-a.left,x1:i.right-a.left,y:(i.top+i.bottom)/2-a.top,text:e.hovertext,color:r.bgcolor,borderColor:r.bordercolor,fontFamily:n.family,fontSize:n.size,fontColor:n.color,fontWeight:n.weight,fontStyle:n.style,fontVariant:n.variant},{container:w._hoverlayer.node(),outerContainer:w._paper.node(),gd:t})})).on("mouseout",(function(){f.loneUnhover(w._hoverlayer.node())}));var F=e.borderwidth,B=e.borderpad,N=F+B,j=R.append("rect").attr("class","bg").style("stroke-width",F+"px").call(u.stroke,e.bordercolor).call(u.fill,e.bgcolor),U=e.width||e.height,V=w._topclips.selectAll("#"+L).data(U?[0]:[]);V.enter().append("clipPath").classed("annclip",!0).attr("id",L).append("rect"),V.exit().remove();var q=e.font,H=w._meta?o.templateString(e.text,w._meta):e.text,G=R.append("text").classed("annotation-text",!0).text(H);k.annotationText?G.call(h.makeEditable,{delegate:R,gd:t}).call(Y).on("edit",(function(r){e.text=r,this.call(Y),S("text",r),y&&y.autorange&&M(y._name+".autorange",!0),x&&x.autorange&&M(x._name+".autorange",!0),i.call("_guiRelayout",t,E())})):G.call(Y)}else n.selectAll("#"+L).remove();function W(t){var n={index:r,annotation:e._input,fullAnnotation:e,event:t};return a&&(n.subplotId=a),n}function Y(r){return r.call(c.font,q).attr({"text-anchor":{left:"start",right:"end"}[e.align]||"middle"}),h.convertToTspans(r,t,X),r}function X(){var r=G.selectAll("a");1===r.size()&&r.text()===G.text()&&R.insert("a",":first-child").attr({"xlink:xlink:href":r.attr("xlink:href"),"xlink:xlink:show":r.attr("xlink:show")}).style({cursor:"pointer"}).node().appendChild(j.node());var n=R.select(".annotation-text-math-group"),f=!n.empty(),v=c.bBox((f?n:G).node()),b=v.width,_=v.height,A=e.width||b,D=e.height||_,B=Math.round(A+2*N),q=Math.round(D+2*N);function H(t,e){return"auto"===e&&(e=t<1/3?"left":t>2/3?"right":"center"),{center:0,middle:0,left:.5,bottom:-.5,right:-.5,top:.5}[e]}for(var Y=!1,X=["x","y"],Z=0;Z<X.length;Z++){var K,J,$,Q,tt,et=X[Z],rt=e[et+"ref"]||et,nt=e["a"+et+"ref"],it={x:y,y:x}[et],at=(P+("x"===et?0:-90))*Math.PI/180,ot=B*Math.cos(at),st=q*Math.sin(at),lt=Math.abs(ot)+Math.abs(st),ut=e[et+"anchor"],ct=e[et+"shift"]*("x"===et?1:-1),ft=C[et],ht=l.getRefType(rt);if(it&&"domain"!==ht){var pt=it.r2fraction(e[et]);(pt<0||pt>1)&&(nt===rt?((pt=it.r2fraction(e["a"+et]))<0||pt>1)&&(Y=!0):Y=!0),K=it._offset+it.r2p(e[et]),Q=.5}else{var dt="domain"===ht;"x"===et?($=e[et],K=dt?it._offset+it._length*$:K=T.l+T.w*$):($=1-e[et],K=dt?it._offset+it._length*$:K=T.t+T.h*$),Q=e.showarrow?.5:$}if(e.showarrow){ft.head=K;var vt=e["a"+et];if(tt=ot*H(.5,e.xanchor)-st*H(.5,e.yanchor),nt===rt){var gt=l.getRefType(nt);"domain"===gt?("y"===et&&(vt=1-vt),ft.tail=it._offset+it._length*vt):"paper"===gt?"y"===et?(vt=1-vt,ft.tail=T.t+T.h*vt):ft.tail=T.l+T.w*vt:ft.tail=it._offset+it.r2p(vt),J=tt}else ft.tail=K+vt,J=tt+vt;ft.text=ft.tail+tt;var yt=w["x"===et?"width":"height"];if("paper"===rt&&(ft.head=o.constrain(ft.head,1,yt-1)),"pixel"===nt){var mt=-Math.max(ft.tail-3,ft.text),xt=Math.min(ft.tail+3,ft.text)-yt;mt>0?(ft.tail+=mt,ft.text+=mt):xt>0&&(ft.tail-=xt,ft.text-=xt)}ft.tail+=ct,ft.head+=ct}else J=tt=lt*H(Q,ut),ft.text=K+tt;ft.text+=ct,tt+=ct,J+=ct,e["_"+et+"padplus"]=lt/2+J,e["_"+et+"padminus"]=lt/2-J,e["_"+et+"size"]=lt,e["_"+et+"shift"]=tt}if(Y)R.remove();else{var bt=0,_t=0;if("left"!==e.align&&(bt=(A-b)*("center"===e.align?.5:1)),"top"!==e.valign&&(_t=(D-_)*("middle"===e.valign?.5:1)),f)n.select("svg").attr({x:N+bt-1,y:N+_t}).call(c.setClipUrl,U?L:null,t);else{var wt=N+_t-v.top,Tt=N+bt-v.left;G.call(h.positionText,Tt,wt).call(c.setClipUrl,U?L:null,t)}V.select("rect").call(c.setRect,N,N,A,D),j.call(c.setRect,F/2,F/2,B-F,q-F),R.call(c.setTranslate,Math.round(C.x.text-B/2),Math.round(C.y.text-q/2)),I.attr({transform:"rotate("+P+","+C.x.text+","+C.y.text+")"});var kt,At=function(r,n){O.selectAll(".annotation-arrow-g").remove();var l=C.x.head,f=C.y.head,h=C.x.tail+r,p=C.y.tail+n,v=C.x.text+r,b=C.y.text+n,_=o.rotationXYMatrix(P,v,b),w=o.apply2DTransform(_),A=o.apply2DTransform2(_),L=+j.attr("width"),z=+j.attr("height"),D=v-.5*L,F=D+L,B=b-.5*z,N=B+z,U=[[D,B,D,N],[D,N,F,N],[F,N,F,B],[F,B,D,B]].map(A);if(!U.reduce((function(t,e){return t^!!o.segmentsIntersect(l,f,l+1e6,f+1e6,e[0],e[1],e[2],e[3])}),!1)){U.forEach((function(t){var e=o.segmentsIntersect(h,p,l,f,t[0],t[1],t[2],t[3]);e&&(h=e.x,p=e.y)}));var V=e.arrowwidth,q=e.arrowcolor,H=e.arrowside,G=O.append("g").style({opacity:u.opacity(q)}).classed("annotation-arrow-g",!0),W=G.append("path").attr("d","M"+h+","+p+"L"+l+","+f).style("stroke-width",V+"px").call(u.stroke,u.rgb(q));if(g(W,H,e),k.annotationPosition&&W.node().parentNode&&!a){var Y=l,X=f;if(e.standoff){var Z=Math.sqrt(Math.pow(l-h,2)+Math.pow(f-p,2));Y+=e.standoff*(h-l)/Z,X+=e.standoff*(p-f)/Z}var K,J,$=G.append("path").classed("annotation-arrow",!0).classed("anndrag",!0).classed("cursor-move",!0).attr({d:"M3,3H-3V-3H3ZM0,0L"+(h-Y)+","+(p-X),transform:s(Y,X)}).style("stroke-width",V+6+"px").call(u.stroke,"rgba(0,0,0,0)").call(u.fill,"rgba(0,0,0,0)");d.init({element:$.node(),gd:t,prepFn:function(){var t=c.getTranslate(R);K=t.x,J=t.y,y&&y.autorange&&M(y._name+".autorange",!0),x&&x.autorange&&M(x._name+".autorange",!0)},moveFn:function(t,r){var n=w(K,J),i=n[0]+t,a=n[1]+r;R.call(c.setTranslate,i,a),S("x",m(y,t,"x",T,e)),S("y",m(x,r,"y",T,e)),e.axref===e.xref&&S("ax",m(y,t,"ax",T,e)),e.ayref===e.yref&&S("ay",m(x,r,"ay",T,e)),G.attr("transform",s(t,r)),I.attr({transform:"rotate("+P+","+i+","+a+")"})},doneFn:function(){i.call("_guiRelayout",t,E());var e=document.querySelector(".js-notes-box-panel");e&&e.redraw(e.selectedObj)}})}}};e.showarrow&&At(0,0),z&&d.init({element:R.node(),gd:t,prepFn:function(){kt=I.attr("transform")},moveFn:function(t,r){var n="pointer";if(e.showarrow)e.axref===e.xref?S("ax",m(y,t,"ax",T,e)):S("ax",e.ax+t),e.ayref===e.yref?S("ay",m(x,r,"ay",T.w,e)):S("ay",e.ay+r),At(t,r);else{if(a)return;var i,o;if(y)i=m(y,t,"x",T,e);else{var l=e._xsize/T.w,u=e.x+(e._xshift-e.xshift)/T.w-l/2;i=d.align(u+t/T.w,l,0,1,e.xanchor)}if(x)o=m(x,r,"y",T,e);else{var c=e._ysize/T.h,f=e.y-(e._yshift+e.yshift)/T.h-c/2;o=d.align(f-r/T.h,c,0,1,e.yanchor)}S("x",i),S("y",o),y&&x||(n=d.getCursor(y?.5:i,x?.5:o,e.xanchor,e.yanchor))}I.attr({transform:s(t,r)+kt}),p(R,n)},clickFn:function(r,n){e.captureevents&&t.emit("plotly_clickannotation",W(n))},doneFn:function(){p(R),i.call("_guiRelayout",t,E());var e=document.querySelector(".js-notes-box-panel");e&&e.redraw(e.selectedObj)}})}}}t.exports={draw:function(t){var e=t._fullLayout;e._infolayer.selectAll(".annotation").remove();for(var r=0;r<e.annotations.length;r++)e.annotations[r].visible&&y(t,r);return a.previousPromises(t)},drawOne:y,drawRaw:x}},33652:function(t,e,r){"use strict";var n=r(33428),i=r(76308),a=r(72196),o=r(3400),s=o.strScale,l=o.strRotate,u=o.strTranslate;t.exports=function(t,e,r){var o,c,f,h,p=t.node(),d=a[r.arrowhead||0],v=a[r.startarrowhead||0],g=(r.arrowwidth||1)*(r.arrowsize||1),y=(r.arrowwidth||1)*(r.startarrowsize||1),m=e.indexOf("start")>=0,x=e.indexOf("end")>=0,b=d.backoff*g+r.standoff,_=v.backoff*y+r.startstandoff;if("line"===p.nodeName){o={x:+t.attr("x1"),y:+t.attr("y1")},c={x:+t.attr("x2"),y:+t.attr("y2")};var w=o.x-c.x,T=o.y-c.y;if(h=(f=Math.atan2(T,w))+Math.PI,b&&_&&b+_>Math.sqrt(w*w+T*T))return void z();if(b){if(b*b>w*w+T*T)return void z();var k=b*Math.cos(f),A=b*Math.sin(f);c.x+=k,c.y+=A,t.attr({x2:c.x,y2:c.y})}if(_){if(_*_>w*w+T*T)return void z();var M=_*Math.cos(f),S=_*Math.sin(f);o.x-=M,o.y-=S,t.attr({x1:o.x,y1:o.y})}}else if("path"===p.nodeName){var E=p.getTotalLength(),L="";if(E<b+_)return void z();var C=p.getPointAtLength(0),P=p.getPointAtLength(.1);f=Math.atan2(C.y-P.y,C.x-P.x),o=p.getPointAtLength(Math.min(_,E)),L="0px,"+_+"px,";var O=p.getPointAtLength(E),I=p.getPointAtLength(E-.1);h=Math.atan2(O.y-I.y,O.x-I.x),c=p.getPointAtLength(Math.max(0,E-b)),L+=E-(L?_+b:b)+"px,"+E+"px",t.style("stroke-dasharray",L)}function z(){t.style("stroke-dasharray","0px,100px")}function D(e,a,o,c){e.path&&(e.noRotate&&(o=0),n.select(p.parentNode).append("path").attr({class:t.attr("class"),d:e.path,transform:u(a.x,a.y)+l(180*o/Math.PI)+s(c)}).style({fill:i.rgb(r.arrowcolor),"stroke-width":0}))}m&&D(v,o,f,y),x&&D(d,c,h,g)}},79180:function(t,e,r){"use strict";var n=r(23816),i=r(42300);t.exports={moduleType:"component",name:"annotations",layoutAttributes:r(13916),supplyLayoutDefaults:r(45216),includeBasePlot:r(36632)("annotations"),calcAutorange:r(90272),draw:n.draw,drawOne:n.drawOne,drawRaw:n.drawRaw,hasClickToShow:i.hasClickToShow,onClick:i.onClick,convertCoords:r(26828)}},45899:function(t,e,r){"use strict";var n=r(13916),i=r(67824).overrideAll,a=r(31780).templatedArray;t.exports=i(a("annotation",{visible:n.visible,x:{valType:"any"},y:{valType:"any"},z:{valType:"any"},ax:{valType:"number"},ay:{valType:"number"},xanchor:n.xanchor,xshift:n.xshift,yanchor:n.yanchor,yshift:n.yshift,text:n.text,textangle:n.textangle,font:n.font,width:n.width,height:n.height,opacity:n.opacity,align:n.align,valign:n.valign,bgcolor:n.bgcolor,bordercolor:n.bordercolor,borderpad:n.borderpad,borderwidth:n.borderwidth,showarrow:n.showarrow,arrowcolor:n.arrowcolor,arrowhead:n.arrowhead,startarrowhead:n.startarrowhead,arrowside:n.arrowside,arrowsize:n.arrowsize,startarrowsize:n.startarrowsize,arrowwidth:n.arrowwidth,standoff:n.standoff,startstandoff:n.startstandoff,hovertext:n.hovertext,hoverlabel:n.hoverlabel,captureevents:n.captureevents}),"calc","from-root")},42456:function(t,e,r){"use strict";var n=r(3400),i=r(54460);function a(t,e){var r=e.fullSceneLayout.domain,a=e.fullLayout._size,o={pdata:null,type:"linear",autorange:!1,range:[-1/0,1/0]};t._xa={},n.extendFlat(t._xa,o),i.setConvert(t._xa),t._xa._offset=a.l+r.x[0]*a.w,t._xa.l2p=function(){return.5*(1+t._pdata[0]/t._pdata[3])*a.w*(r.x[1]-r.x[0])},t._ya={},n.extendFlat(t._ya,o),i.setConvert(t._ya),t._ya._offset=a.t+(1-r.y[1])*a.h,t._ya.l2p=function(){return.5*(1-t._pdata[1]/t._pdata[3])*a.h*(r.y[1]-r.y[0])}}t.exports=function(t){for(var e=t.fullSceneLayout.annotations,r=0;r<e.length;r++)a(e[r],t);t.fullLayout._infolayer.selectAll(".annotation-"+t.id).remove()}},52808:function(t,e,r){"use strict";var n=r(3400),i=r(54460),a=r(51272),o=r(87192),s=r(45899);function l(t,e,r,a){function l(r,i){return n.coerce(t,e,s,r,i)}function u(t){var n=t+"axis",a={_fullLayout:{}};return a._fullLayout[n]=r[n],i.coercePosition(e,a,l,t,t,.5)}l("visible")&&(o(t,e,a.fullLayout,l),u("x"),u("y"),u("z"),n.noneOrAll(t,e,["x","y","z"]),e.xref="x",e.yref="y",e.zref="z",l("xanchor"),l("yanchor"),l("xshift"),l("yshift"),e.showarrow&&(e.axref="pixel",e.ayref="pixel",l("ax",-10),l("ay",-30),n.noneOrAll(t,e,["ax","ay"])))}t.exports=function(t,e,r){a(t,e,{name:"annotations",handleItemDefaults:l,fullLayout:r.fullLayout})}},71836:function(t,e,r){"use strict";var n=r(23816).drawRaw,i=r(94424),a=["x","y","z"];t.exports=function(t){for(var e=t.fullSceneLayout,r=t.dataScale,o=e.annotations,s=0;s<o.length;s++){for(var l=o[s],u=!1,c=0;c<3;c++){var f=a[c],h=l[f],p=e[f+"axis"].r2fraction(h);if(p<0||p>1){u=!0;break}}u?t.fullLayout._infolayer.select(".annotation-"+t.id+'[data-index="'+s+'"]').remove():(l._pdata=i(t.glplot.cameraParams,[e.xaxis.r2l(l.x)*r[0],e.yaxis.r2l(l.y)*r[1],e.zaxis.r2l(l.z)*r[2]]),n(t.graphDiv,l,s,t.id,l._xa,l._ya))}}},56864:function(t,e,r){"use strict";var n=r(24040),i=r(3400);t.exports={moduleType:"component",name:"annotations3d",schema:{subplots:{scene:{annotations:r(45899)}}},layoutAttributes:r(45899),handleDefaults:r(52808),includeBasePlot:function(t,e){var r=n.subplotsRegistry.gl3d;if(r)for(var a=r.attrRegex,o=Object.keys(t),s=0;s<o.length;s++){var l=o[s];a.test(l)&&(t[l].annotations||[]).length&&(i.pushUnique(e._basePlotModules,r),i.pushUnique(e._subplots.gl3d,l))}},convert:r(42456),draw:r(71836)}},54976:function(t,e,r){"use strict";t.exports=r(38700),r(15168),r(67020),r(89792),r(55668),r(65168),r(2084),r(26368),r(24747),r(65616),r(30632),r(73040),r(1104),r(51456),r(4592),r(45348)},97776:function(t,e,r){"use strict";var n=r(54976),i=r(3400),a=r(39032),o=a.EPOCHJD,s=a.ONEDAY,l={valType:"enumerated",values:i.sortObjectKeys(n.calendars),editType:"calc",dflt:"gregorian"},u=function(t,e,r,n){var a={};return a[r]=l,i.coerce(t,e,a,r,n)},c="##",f={d:{0:"dd","-":"d"},e:{0:"d","-":"d"},a:{0:"D","-":"D"},A:{0:"DD","-":"DD"},j:{0:"oo","-":"o"},W:{0:"ww","-":"w"},m:{0:"mm","-":"m"},b:{0:"M","-":"M"},B:{0:"MM","-":"MM"},y:{0:"yy","-":"yy"},Y:{0:"yyyy","-":"yyyy"},U:c,w:c,c:{0:"D M d %X yyyy","-":"D M d %X yyyy"},x:{0:"mm/dd/yyyy","-":"mm/dd/yyyy"}},h={};function p(t){var e=h[t];return e||(h[t]=n.instance(t))}function d(t){return i.extendFlat({},l,{description:t})}function v(t){return"Sets the calendar system to use with `"+t+"` date data."}var g={xcalendar:d(v("x"))},y=i.extendFlat({},g,{ycalendar:d(v("y"))}),m=i.extendFlat({},y,{zcalendar:d(v("z"))}),x=d(["Sets the calendar system to use for `range` and `tick0`","if this is a date axis. This does not set the calendar for","interpreting data on this axis, that's specified in the trace","or via the global `layout.calendar`"].join(" "));t.exports={moduleType:"component",name:"calendars",schema:{traces:{scatter:y,bar:y,box:y,heatmap:y,contour:y,histogram:y,histogram2d:y,histogram2dcontour:y,scatter3d:m,surface:m,mesh3d:m,scattergl:y,ohlc:g,candlestick:g},layout:{calendar:d(["Sets the default calendar system to use for interpreting and","displaying dates throughout the plot."].join(" "))},subplots:{xaxis:{calendar:x},yaxis:{calendar:x},scene:{xaxis:{calendar:x},yaxis:{calendar:x},zaxis:{calendar:x}},polar:{radialaxis:{calendar:x}}},transforms:{filter:{valuecalendar:d(["WARNING: All transforms are deprecated and may be removed from the API in next major version.","Sets the calendar system to use for `value`, if it is a date."].join(" ")),targetcalendar:d(["WARNING: All transforms are deprecated and may be removed from the API in next major version.","Sets the calendar system to use for `target`, if it is an","array of dates. If `target` is a string (eg *x*) we use the","corresponding trace attribute (eg `xcalendar`) if it exists,","even if `targetcalendar` is provided."].join(" "))}}},layoutAttributes:l,handleDefaults:u,handleTraceDefaults:function(t,e,r,n){for(var i=0;i<r.length;i++)u(t,e,r[i]+"calendar",n.calendar)},CANONICAL_SUNDAY:{chinese:"2000-01-02",coptic:"2000-01-03",discworld:"2000-01-03",ethiopian:"2000-01-05",hebrew:"5000-01-01",islamic:"1000-01-02",julian:"2000-01-03",mayan:"5000-01-01",nanakshahi:"1000-01-05",nepali:"2000-01-05",persian:"1000-01-01",jalali:"1000-01-01",taiwan:"1000-01-04",thai:"2000-01-04",ummalqura:"1400-01-06"},CANONICAL_TICK:{chinese:"2000-01-01",coptic:"2000-01-01",discworld:"2000-01-01",ethiopian:"2000-01-01",hebrew:"5000-01-01",islamic:"1000-01-01",julian:"2000-01-01",mayan:"5000-01-01",nanakshahi:"1000-01-01",nepali:"2000-01-01",persian:"1000-01-01",jalali:"1000-01-01",taiwan:"1000-01-01",thai:"2000-01-01",ummalqura:"1400-01-01"},DFLTRANGE:{chinese:["2000-01-01","2001-01-01"],coptic:["1700-01-01","1701-01-01"],discworld:["1800-01-01","1801-01-01"],ethiopian:["2000-01-01","2001-01-01"],hebrew:["5700-01-01","5701-01-01"],islamic:["1400-01-01","1401-01-01"],julian:["2000-01-01","2001-01-01"],mayan:["5200-01-01","5201-01-01"],nanakshahi:["0500-01-01","0501-01-01"],nepali:["2000-01-01","2001-01-01"],persian:["1400-01-01","1401-01-01"],jalali:["1400-01-01","1401-01-01"],taiwan:["0100-01-01","0101-01-01"],thai:["2500-01-01","2501-01-01"],ummalqura:["1400-01-01","1401-01-01"]},getCal:p,worldCalFmt:function(t,e,r){for(var n,i,a,l,u,h=Math.floor((e+.05)/s)+o,d=p(r).fromJD(h),v=0;-1!==(v=t.indexOf("%",v));)"0"===(n=t.charAt(v+1))||"-"===n||"_"===n?(a=3,i=t.charAt(v+2),"_"===n&&(n="-")):(i=n,n="0",a=2),(l=f[i])?(u=l===c?c:d.formatDate(l[n]),t=t.substr(0,v)+u+t.substr(v+a),v+=u.length):v+=a;return t}}},22548:function(t,e){"use strict";e.defaults=["#1f77b4","#ff7f0e","#2ca02c","#d62728","#9467bd","#8c564b","#e377c2","#7f7f7f","#bcbd22","#17becf"],e.defaultLine="#444",e.lightLine="#eee",e.background="#fff",e.borderLine="#BEC8D9",e.lightFraction=1e3/11},76308:function(t,e,r){"use strict";var n=r(49760),i=r(38248),a=r(38116).isTypedArray,o=t.exports={},s=r(22548);o.defaults=s.defaults;var l=o.defaultLine=s.defaultLine;o.lightLine=s.lightLine;var u=o.background=s.background;function c(t){if(i(t)||"string"!=typeof t)return t;var e=t.trim();if("rgb"!==e.substr(0,3))return t;var r=e.match(/^rgba?\s*\(([^()]*)\)$/);if(!r)return t;var n=r[1].trim().split(/\s*[\s,]\s*/),a="a"===e.charAt(3)&&4===n.length;if(!a&&3!==n.length)return t;for(var o=0;o<n.length;o++){if(!n[o].length)return t;if(n[o]=Number(n[o]),!(n[o]>=0))return t;if(3===o)n[o]>1&&(n[o]=1);else if(n[o]>=1)return t}var s=Math.round(255*n[0])+", "+Math.round(255*n[1])+", "+Math.round(255*n[2]);return a?"rgba("+s+", "+n[3]+")":"rgb("+s+")"}o.tinyRGB=function(t){var e=t.toRgb();return"rgb("+Math.round(e.r)+", "+Math.round(e.g)+", "+Math.round(e.b)+")"},o.rgb=function(t){return o.tinyRGB(n(t))},o.opacity=function(t){return t?n(t).getAlpha():0},o.addOpacity=function(t,e){var r=n(t).toRgb();return"rgba("+Math.round(r.r)+", "+Math.round(r.g)+", "+Math.round(r.b)+", "+e+")"},o.combine=function(t,e){var r=n(t).toRgb();if(1===r.a)return n(t).toRgbString();var i=n(e||u).toRgb(),a=1===i.a?i:{r:255*(1-i.a)+i.r*i.a,g:255*(1-i.a)+i.g*i.a,b:255*(1-i.a)+i.b*i.a},o={r:a.r*(1-r.a)+r.r*r.a,g:a.g*(1-r.a)+r.g*r.a,b:a.b*(1-r.a)+r.b*r.a};return n(o).toRgbString()},o.interpolate=function(t,e,r){var i=n(t).toRgb(),a=n(e).toRgb(),o={r:r*i.r+(1-r)*a.r,g:r*i.g+(1-r)*a.g,b:r*i.b+(1-r)*a.b};return n(o).toRgbString()},o.contrast=function(t,e,r){var i=n(t);return 1!==i.getAlpha()&&(i=n(o.combine(t,u))),(i.isDark()?e?i.lighten(e):u:r?i.darken(r):l).toString()},o.stroke=function(t,e){var r=n(e);t.style({stroke:o.tinyRGB(r),"stroke-opacity":r.getAlpha()})},o.fill=function(t,e){var r=n(e);t.style({fill:o.tinyRGB(r),"fill-opacity":r.getAlpha()})},o.clean=function(t){if(t&&"object"==typeof t){var e,r,n,i,s=Object.keys(t);for(e=0;e<s.length;e++)if(i=t[n=s[e]],"color"===n.substr(n.length-5))if(Array.isArray(i))for(r=0;r<i.length;r++)i[r]=c(i[r]);else t[n]=c(i);else if("colorscale"===n.substr(n.length-10)&&Array.isArray(i))for(r=0;r<i.length;r++)Array.isArray(i[r])&&(i[r][1]=c(i[r][1]));else if(Array.isArray(i)){var l=i[0];if(!Array.isArray(l)&&l&&"object"==typeof l)for(r=0;r<i.length;r++)o.clean(i[r])}else i&&"object"==typeof i&&!a(i)&&o.clean(i)}}},42996:function(t,e,r){"use strict";var n=r(94724),i=r(25376),a=r(92880).extendFlat,o=r(67824).overrideAll;t.exports=o({orientation:{valType:"enumerated",values:["h","v"],dflt:"v"},thicknessmode:{valType:"enumerated",values:["fraction","pixels"],dflt:"pixels"},thickness:{valType:"number",min:0,dflt:30},lenmode:{valType:"enumerated",values:["fraction","pixels"],dflt:"fraction"},len:{valType:"number",min:0,dflt:1},x:{valType:"number"},xref:{valType:"enumerated",dflt:"paper",values:["container","paper"],editType:"layoutstyle"},xanchor:{valType:"enumerated",values:["left","center","right"]},xpad:{valType:"number",min:0,dflt:10},y:{valType:"number"},yref:{valType:"enumerated",dflt:"paper",values:["container","paper"],editType:"layoutstyle"},yanchor:{valType:"enumerated",values:["top","middle","bottom"]},ypad:{valType:"number",min:0,dflt:10},outlinecolor:n.linecolor,outlinewidth:n.linewidth,bordercolor:n.linecolor,borderwidth:{valType:"number",min:0,dflt:0},bgcolor:{valType:"color",dflt:"rgba(0,0,0,0)"},tickmode:n.minor.tickmode,nticks:n.nticks,tick0:n.tick0,dtick:n.dtick,tickvals:n.tickvals,ticktext:n.ticktext,ticks:a({},n.ticks,{dflt:""}),ticklabeloverflow:a({},n.ticklabeloverflow,{}),ticklabelposition:{valType:"enumerated",values:["outside","inside","outside top","inside top","outside left","inside left","outside right","inside right","outside bottom","inside bottom"],dflt:"outside"},ticklen:n.ticklen,tickwidth:n.tickwidth,tickcolor:n.tickcolor,ticklabelstep:n.ticklabelstep,showticklabels:n.showticklabels,labelalias:n.labelalias,tickfont:i({}),tickangle:n.tickangle,tickformat:n.tickformat,tickformatstops:n.tickformatstops,tickprefix:n.tickprefix,showtickprefix:n.showtickprefix,ticksuffix:n.ticksuffix,showticksuffix:n.showticksuffix,separatethousands:n.separatethousands,exponentformat:n.exponentformat,minexponent:n.minexponent,showexponent:n.showexponent,title:{text:{valType:"string"},font:i({}),side:{valType:"enumerated",values:["right","top","bottom"]}},_deprecated:{title:{valType:"string"},titlefont:i({}),titleside:{valType:"enumerated",values:["right","top","bottom"],dflt:"top"}}},"colorbars","from-root")},63964:function(t){"use strict";t.exports={cn:{colorbar:"colorbar",cbbg:"cbbg",cbfill:"cbfill",cbfills:"cbfills",cbline:"cbline",cblines:"cblines",cbaxis:"cbaxis",cbtitleunshift:"cbtitleunshift",cbtitle:"cbtitle",cboutline:"cboutline",crisp:"crisp",jsPlaceholder:"js-placeholder"}}},64013:function(t,e,r){"use strict";var n=r(3400),i=r(31780),a=r(26332),o=r(25404),s=r(95936),l=r(42568),u=r(42996);t.exports=function(t,e,r){var c=i.newContainer(e,"colorbar"),f=t.colorbar||{};function h(t,e){return n.coerce(f,c,u,t,e)}var p=r.margin||{t:0,b:0,l:0,r:0},d=r.width-p.l-p.r,v=r.height-p.t-p.b,g="v"===h("orientation"),y=h("thicknessmode");h("thickness","fraction"===y?30/(g?d:v):30);var m=h("lenmode");h("len","fraction"===m?1:g?v:d);var x,b,_,w="paper"===h("yref"),T="paper"===h("xref"),k="left";g?(_="middle",k=T?"left":"right",x=T?1.02:1,b=.5):(_=w?"bottom":"top",k="center",x=.5,b=w?1.02:1),n.coerce(f,c,{x:{valType:"number",min:T?-2:0,max:T?3:1,dflt:x}},"x"),n.coerce(f,c,{y:{valType:"number",min:w?-2:0,max:w?3:1,dflt:b}},"y"),h("xanchor",k),h("xpad"),h("yanchor",_),h("ypad"),n.noneOrAll(f,c,["x","y"]),h("outlinecolor"),h("outlinewidth"),h("bordercolor"),h("borderwidth"),h("bgcolor");var A=n.coerce(f,c,{ticklabelposition:{valType:"enumerated",dflt:"outside",values:g?["outside","inside","outside top","inside top","outside bottom","inside bottom"]:["outside","inside","outside left","inside left","outside right","inside right"]}},"ticklabelposition");h("ticklabeloverflow",-1!==A.indexOf("inside")?"hide past domain":"hide past div"),a(f,c,h,"linear");var M=r.font,S={noAutotickangles:!0,outerTicks:!1,font:M};-1!==A.indexOf("inside")&&(S.bgColor="black"),l(f,c,h,"linear",S),s(f,c,h,"linear",S),o(f,c,h,"linear",S),h("title.text",r._dfltTitle.colorbar);var E=c.showticklabels?c.tickfont:M,L=n.extendFlat({},E,{weight:M.weight,style:M.style,variant:M.variant,color:M.color,size:n.bigFont(E.size)});n.coerceFont(h,"title.font",L),h("title.side",g?"top":"right")}},37848:function(t,e,r){"use strict";var n=r(33428),i=r(49760),a=r(7316),o=r(24040),s=r(54460),l=r(86476),u=r(3400),c=u.strTranslate,f=r(92880).extendFlat,h=r(93972),p=r(43616),d=r(76308),v=r(81668),g=r(72736),y=r(94288).flipScale,m=r(28336),x=r(37668),b=r(94724),_=r(84284),w=_.LINE_SPACING,T=_.FROM_TL,k=_.FROM_BR,A=r(63964).cn;t.exports={draw:function(t){var e=t._fullLayout._infolayer.selectAll("g."+A.colorbar).data(function(t){var e,r,n,i,a=t._fullLayout,o=t.calcdata,s=[];function l(t){return f(t,{_fillcolor:null,_line:{color:null,width:null,dash:null},_levels:{start:null,end:null,size:null},_filllevels:null,_fillgradient:null,_zrange:null})}function u(){"function"==typeof i.calc?i.calc(t,n,e):(e._fillgradient=r.reversescale?y(r.colorscale):r.colorscale,e._zrange=[r[i.min],r[i.max]])}for(var c=0;c<o.length;c++){var h=o[c];if((n=h[0].trace)._module){var p=n._module.colorbar;if(!0===n.visible&&p)for(var d=Array.isArray(p),v=d?p:[p],g=0;g<v.length;g++){var m=(i=v[g]).container;(r=m?n[m]:n)&&r.showscale&&((e=l(r.colorbar))._id="cb"+n.uid+(d&&m?"-"+m:""),e._traceIndex=n.index,e._propPrefix=(m?m+".":"")+"colorbar.",e._meta=n._meta,u(),s.push(e))}}}for(var x in a._colorAxes)if((r=a[x]).showscale){var b=a._colorAxes[x];(e=l(r.colorbar))._id="cb"+x,e._propPrefix=x+".colorbar.",e._meta=a._meta,i={min:"cmin",max:"cmax"},"heatmap"!==b[0]&&(n=b[1],i.calc=n._module.colorbar.calc),u(),s.push(e)}return s}(t),(function(t){return t._id}));e.enter().append("g").attr("class",(function(t){return t._id})).classed(A.colorbar,!0),e.each((function(e){var r=n.select(this);u.ensureSingle(r,"rect",A.cbbg),u.ensureSingle(r,"g",A.cbfills),u.ensureSingle(r,"g",A.cblines),u.ensureSingle(r,"g",A.cbaxis,(function(t){t.classed(A.crisp,!0)})),u.ensureSingle(r,"g",A.cbtitleunshift,(function(t){t.append("g").classed(A.cbtitle,!0)})),u.ensureSingle(r,"rect",A.cboutline);var y=function(t,e,r){var o="v"===e.orientation,l=e.len,h=e.lenmode,y=e.thickness,_=e.thicknessmode,M=e.outlinewidth,S=e.borderwidth,E=e.bgcolor,L=e.xanchor,C=e.yanchor,P=e.xpad,O=e.ypad,I=e.x,z=o?e.y:1-e.y,D="paper"===e.yref,R="paper"===e.xref,F=r._fullLayout,B=F._size,N=e._fillcolor,j=e._line,U=e.title,V=U.side,q=e._zrange||n.extent(("function"==typeof N?N:j.color).domain()),H="function"==typeof j.color?j.color:function(){return j.color},G="function"==typeof N?N:function(){return N},W=e._levels,Y=function(t,e,r){var n,i,a=e._levels,o=[],s=[],l=a.end+a.size/100,u=a.size,c=1.001*r[0]-.001*r[1],f=1.001*r[1]-.001*r[0];for(i=0;i<1e5&&(n=a.start+i*u,!(u>0?n>=l:n<=l));i++)n>c&&n<f&&o.push(n);if(e._fillgradient)s=[0];else if("function"==typeof e._fillcolor){var h=e._filllevels;if(h)for(l=h.end+h.size/100,u=h.size,i=0;i<1e5&&(n=h.start+i*u,!(u>0?n>=l:n<=l));i++)n>r[0]&&n<r[1]&&s.push(n);else(s=o.map((function(t){return t-a.size/2}))).push(s[s.length-1]+a.size)}else e._fillcolor&&"string"==typeof e._fillcolor&&(s=[0]);return a.size<0&&(o.reverse(),s.reverse()),{line:o,fill:s}}(0,e,q),X=Y.fill,Z=Y.line,K=Math.round(y*("fraction"===_?o?B.w:B.h:1)),J=K/(o?B.w:B.h),$=Math.round(l*("fraction"===h?o?B.h:B.w:1)),Q=$/(o?B.h:B.w),tt=R?B.w:r._fullLayout.width,et=D?B.h:r._fullLayout.height,rt=Math.round(o?I*tt+P:z*et+O),nt={center:.5,right:1}[L]||0,it={top:1,middle:.5}[C]||0,at=o?I-nt*J:z-it*J,ot=o?z-it*Q:I-nt*Q,st=Math.round(o?et*(1-ot):tt*ot);e._lenFrac=Q,e._thickFrac=J,e._uFrac=at,e._vFrac=ot;var lt=e._axis=function(t,e,r){var n=t._fullLayout,i="v"===e.orientation,a={type:"linear",range:r,tickmode:e.tickmode,nticks:e.nticks,tick0:e.tick0,dtick:e.dtick,tickvals:e.tickvals,ticktext:e.ticktext,ticks:e.ticks,ticklen:e.ticklen,tickwidth:e.tickwidth,tickcolor:e.tickcolor,showticklabels:e.showticklabels,labelalias:e.labelalias,ticklabelposition:e.ticklabelposition,ticklabeloverflow:e.ticklabeloverflow,ticklabelstep:e.ticklabelstep,tickfont:e.tickfont,tickangle:e.tickangle,tickformat:e.tickformat,exponentformat:e.exponentformat,minexponent:e.minexponent,separatethousands:e.separatethousands,showexponent:e.showexponent,showtickprefix:e.showtickprefix,tickprefix:e.tickprefix,showticksuffix:e.showticksuffix,ticksuffix:e.ticksuffix,title:e.title,showline:!0,anchor:"free",side:i?"right":"bottom",position:1},o=i?"y":"x",s={type:"linear",_id:o+e._id},l={letter:o,font:n.font,noAutotickangles:"y"===o,noHover:!0,noTickson:!0,noTicklabelmode:!0,noInsideRange:!0,calendar:n.calendar};function c(t,e){return u.coerce(a,s,b,t,e)}return m(a,s,c,l,n),x(a,s,c,l),s}(r,e,q);lt.position=J+(o?I+P/B.w:z+O/B.h);var ut=-1!==["top","bottom"].indexOf(V);if(o&&ut&&(lt.title.side=V,lt.titlex=I+P/B.w,lt.titley=ot+("top"===U.side?Q-O/B.h:O/B.h)),o||ut||(lt.title.side=V,lt.titley=z+O/B.h,lt.titlex=ot+P/B.w),j.color&&"auto"===e.tickmode){lt.tickmode="linear",lt.tick0=W.start;var ct=W.size,ft=u.constrain($/50,4,15)+1,ht=(q[1]-q[0])/((e.nticks||ft)*ct);if(ht>1){var pt=Math.pow(10,Math.floor(Math.log(ht)/Math.LN10));ct*=pt*u.roundUp(ht/pt,[2,5,10]),(Math.abs(W.start)/W.size+1e-6)%1<2e-6&&(lt.tick0=0)}lt.dtick=ct}lt.domain=o?[ot+O/B.h,ot+Q-O/B.h]:[ot+P/B.w,ot+Q-P/B.w],lt.setScale(),t.attr("transform",c(Math.round(B.l),Math.round(B.t)));var dt,vt=t.select("."+A.cbtitleunshift).attr("transform",c(-Math.round(B.l),-Math.round(B.t))),gt=lt.ticklabelposition,yt=lt.title.font.size,mt=t.select("."+A.cbaxis),xt=0,bt=0;function _t(n,i){var a={propContainer:lt,propName:e._propPrefix+"title",traceIndex:e._traceIndex,_meta:e._meta,placeholder:F._dfltTitle.colorbar,containerGroup:t.select("."+A.cbtitle)},o="h"===n.charAt(0)?n.substr(1):"h"+n;t.selectAll("."+o+",."+o+"-math-group").remove(),v.draw(r,n,f(a,i||{}))}return u.syncOrAsync([a.previousPromises,function(){var t,e;(o&&ut||!o&&!ut)&&("top"===V&&(t=P+B.l+tt*I,e=O+B.t+et*(1-ot-Q)+3+.75*yt),"bottom"===V&&(t=P+B.l+tt*I,e=O+B.t+et*(1-ot)-3-.25*yt),"right"===V&&(e=O+B.t+et*z+3+.75*yt,t=P+B.l+tt*ot),_t(lt._id+"title",{attributes:{x:t,y:e,"text-anchor":o?"start":"middle"}}))},function(){if(!o&&!ut||o&&ut){var a,l=t.select("."+A.cbtitle),f=l.select("text"),h=[-M/2,M/2],d=l.select(".h"+lt._id+"title-math-group").node(),v=15.6;if(f.node()&&(v=parseInt(f.node().style.fontSize,10)*w),d?(a=p.bBox(d),bt=a.width,(xt=a.height)>v&&(h[1]-=(xt-v)/2)):f.node()&&!f.classed(A.jsPlaceholder)&&(a=p.bBox(f.node()),bt=a.width,xt=a.height),o){if(xt){if(xt+=5,"top"===V)lt.domain[1]-=xt/B.h,h[1]*=-1;else{lt.domain[0]+=xt/B.h;var y=g.lineCount(f);h[1]+=(1-y)*v}l.attr("transform",c(h[0],h[1])),lt.setScale()}}else bt&&("right"===V&&(lt.domain[0]+=(bt+yt/2)/B.w),l.attr("transform",c(h[0],h[1])),lt.setScale())}t.selectAll("."+A.cbfills+",."+A.cblines).attr("transform",o?c(0,Math.round(B.h*(1-lt.domain[1]))):c(Math.round(B.w*lt.domain[0]),0)),mt.attr("transform",o?c(0,Math.round(-B.t)):c(Math.round(-B.l),0));var m=t.select("."+A.cbfills).selectAll("rect."+A.cbfill).attr("style","").data(X);m.enter().append("rect").classed(A.cbfill,!0).attr("style",""),m.exit().remove();var x=q.map(lt.c2p).map(Math.round).sort((function(t,e){return t-e}));m.each((function(t,a){var s=[0===a?q[0]:(X[a]+X[a-1])/2,a===X.length-1?q[1]:(X[a]+X[a+1])/2].map(lt.c2p).map(Math.round);o&&(s[1]=u.constrain(s[1]+(s[1]>s[0])?1:-1,x[0],x[1]));var l=n.select(this).attr(o?"x":"y",rt).attr(o?"y":"x",n.min(s)).attr(o?"width":"height",Math.max(K,2)).attr(o?"height":"width",Math.max(n.max(s)-n.min(s),2));if(e._fillgradient)p.gradient(l,r,e._id,o?"vertical":"horizontalreversed",e._fillgradient,"fill");else{var c=G(t).replace("e-","");l.attr("fill",i(c).toHexString())}}));var b=t.select("."+A.cblines).selectAll("path."+A.cbline).data(j.color&&j.width?Z:[]);b.enter().append("path").classed(A.cbline,!0),b.exit().remove(),b.each((function(t){var e=rt,r=Math.round(lt.c2p(t))+j.width/2%1;n.select(this).attr("d","M"+(o?e+","+r:r+","+e)+(o?"h":"v")+K).call(p.lineGroupStyle,j.width,H(t),j.dash)})),mt.selectAll("g."+lt._id+"tick,path").remove();var _=rt+K+(M||0)/2-("outside"===e.ticks?1:0),T=s.calcTicks(lt),k=s.getTickSigns(lt)[2];return s.drawTicks(r,lt,{vals:"inside"===lt.ticks?s.clipEnds(lt,T):T,layer:mt,path:s.makeTickPath(lt,_,k),transFn:s.makeTransTickFn(lt)}),s.drawLabels(r,lt,{vals:T,layer:mt,transFn:s.makeTransTickLabelFn(lt),labelFns:s.makeLabelFns(lt,_)})},function(){if(o&&!ut||!o&&ut){var t,i,a=lt.position||0,s=lt._offset+lt._length/2;if("right"===V)i=s,t=B.l+tt*a+10+yt*(lt.showticklabels?1:.5);else if(t=s,"bottom"===V&&(i=B.t+et*a+10+(-1===gt.indexOf("inside")?lt.tickfont.size:0)+("intside"!==lt.ticks&&e.ticklen||0)),"top"===V){var l=U.text.split("<br>").length;i=B.t+et*a+10-K-w*yt*l}_t((o?"h":"v")+lt._id+"title",{avoid:{selection:n.select(r).selectAll("g."+lt._id+"tick"),side:V,offsetTop:o?0:B.t,offsetLeft:o?B.l:0,maxShift:o?F.width:F.height},attributes:{x:t,y:i,"text-anchor":"middle"},transform:{rotate:o?-90:0,offset:0}})}},a.previousPromises,function(){var n,s=K+M/2;-1===gt.indexOf("inside")&&(n=p.bBox(mt.node()),s+=o?n.width:n.height),dt=vt.select("text");var u=0,f=o&&"top"===V,v=!o&&"right"===V,g=0;if(dt.node()&&!dt.classed(A.jsPlaceholder)){var m,x=vt.select(".h"+lt._id+"title-math-group").node();x&&(o&&ut||!o&&!ut)?(u=(n=p.bBox(x)).width,m=n.height):(u=(n=p.bBox(vt.node())).right-B.l-(o?rt:st),m=n.bottom-B.t-(o?st:rt),o||"top"!==V||(s+=n.height,g=n.height)),v&&(dt.attr("transform",c(u/2+yt/2,0)),u*=2),s=Math.max(s,o?u:m)}var b=2*(o?P:O)+s+S+M/2,w=0;!o&&U.text&&"bottom"===C&&z<=0&&(b+=w=b/2,g+=w),F._hColorbarMoveTitle=w,F._hColorbarMoveCBTitle=g;var N=S+M,j=(o?rt:st)-N/2-(o?P:0),q=(o?st:rt)-(o?$:O+g-w);t.select("."+A.cbbg).attr("x",j).attr("y",q).attr(o?"width":"height",Math.max(b-w,2)).attr(o?"height":"width",Math.max($+N,2)).call(d.fill,E).call(d.stroke,e.bordercolor).style("stroke-width",S);var H=v?Math.max(u-10,0):0;t.selectAll("."+A.cboutline).attr("x",(o?rt:st+P)+H).attr("y",(o?st+O-$:rt)+(f?xt:0)).attr(o?"width":"height",Math.max(K,2)).attr(o?"height":"width",Math.max($-(o?2*O+xt:2*P+H),2)).call(d.stroke,e.outlinecolor).style({fill:"none","stroke-width":M});var G=o?nt*b:0,W=o?0:(1-it)*b-g;if(G=R?B.l-G:-G,W=D?B.t-W:-W,t.attr("transform",c(G,W)),!o&&(S||i(E).getAlpha()&&!i.equals(F.paper_bgcolor,E))){var Y=mt.selectAll("text"),X=Y[0].length,Z=t.select("."+A.cbbg).node(),J=p.bBox(Z),Q=p.getTranslate(t);Y.each((function(t,e){var r=X-1;if(0===e||e===r){var n,i=p.bBox(this),a=p.getTranslate(this);if(e===r){var o=i.right+a.x;(n=J.right+Q.x+st-S-2+I-o)>0&&(n=0)}else if(0===e){var s=i.left+a.x;(n=J.left+Q.x+st+S+2-s)<0&&(n=0)}n&&(X<3?this.setAttribute("transform","translate("+n+",0) "+this.getAttribute("transform")):this.setAttribute("visibility","hidden"))}}))}var tt={},et=T[L],at=k[L],ot=T[C],ct=k[C],ft=b-K;o?("pixels"===h?(tt.y=z,tt.t=$*ot,tt.b=$*ct):(tt.t=tt.b=0,tt.yt=z+l*ot,tt.yb=z-l*ct),"pixels"===_?(tt.x=I,tt.l=b*et,tt.r=b*at):(tt.l=ft*et,tt.r=ft*at,tt.xl=I-y*et,tt.xr=I+y*at)):("pixels"===h?(tt.x=I,tt.l=$*et,tt.r=$*at):(tt.l=tt.r=0,tt.xl=I+l*et,tt.xr=I-l*at),"pixels"===_?(tt.y=1-z,tt.t=b*ot,tt.b=b*ct):(tt.t=ft*ot,tt.b=ft*ct,tt.yt=z-y*ot,tt.yb=z+y*ct));var ht=e.y<.5?"b":"t",pt=e.x<.5?"l":"r";r._fullLayout._reservedMargin[e._id]={};var bt={r:F.width-j-G,l:j+tt.r,b:F.height-q-W,t:q+tt.b};R&&D?a.autoMargin(r,e._id,tt):R?r._fullLayout._reservedMargin[e._id][ht]=bt[ht]:D||o?r._fullLayout._reservedMargin[e._id][pt]=bt[pt]:r._fullLayout._reservedMargin[e._id][ht]=bt[ht]}],r)}(r,e,t);y&&y.then&&(t._promises||[]).push(y),t._context.edits.colorbarPosition&&function(t,e,r){var n,i,a,s="v"===e.orientation,u=r._fullLayout._size;l.init({element:t.node(),gd:r,prepFn:function(){n=t.attr("transform"),h(t)},moveFn:function(r,o){t.attr("transform",n+c(r,o)),i=l.align((s?e._uFrac:e._vFrac)+r/u.w,s?e._thickFrac:e._lenFrac,0,1,e.xanchor),a=l.align((s?e._vFrac:1-e._uFrac)-o/u.h,s?e._lenFrac:e._thickFrac,0,1,e.yanchor);var f=l.getCursor(i,a,e.xanchor,e.yanchor);h(t,f)},doneFn:function(){if(h(t),void 0!==i&&void 0!==a){var n={};n[e._propPrefix+"x"]=i,n[e._propPrefix+"y"]=a,void 0!==e._traceIndex?o.call("_guiRestyle",r,n,e._traceIndex):o.call("_guiRelayout",r,n)}}})}(r,e,t)})),e.exit().each((function(e){a.autoMargin(t,e._id)})).remove(),e.order()}}},90553:function(t,e,r){"use strict";var n=r(3400);t.exports=function(t){return n.isPlainObject(t.colorbar)}},55080:function(t,e,r){"use strict";t.exports={moduleType:"component",name:"colorbar",attributes:r(42996),supplyDefaults:r(64013),draw:r(37848).draw,hasColorbar:r(90553)}},49084:function(t,e,r){"use strict";var n=r(42996),i=r(53756).counter,a=r(95376),o=r(88304).scales;function s(t){return"`"+t+"`"}a(o),t.exports=function(t,e){t=t||"";var r,a=(e=e||{}).cLetter||"c",l=("onlyIfNumerical"in e?e.onlyIfNumerical:Boolean(t),"noScale"in e?e.noScale:"marker.line"===t),u="showScaleDflt"in e?e.showScaleDflt:"z"===a,c="string"==typeof e.colorscaleDflt?o[e.colorscaleDflt]:null,f=e.editTypeOverride||"",h=t?t+".":"";"colorAttr"in e?(r=e.colorAttr,e.colorAttr):s(h+(r={z:"z",c:"color"}[a]));var p=a+"auto",d=a+"min",v=a+"max",g=a+"mid",y=(s(h+p),s(h+d),s(h+v),{});y[d]=y[v]=void 0;var m={};m[p]=!1;var x={};return"color"===r&&(x.color={valType:"color",arrayOk:!0,editType:f||"style"},e.anim&&(x.color.anim=!0)),x[p]={valType:"boolean",dflt:!0,editType:"calc",impliedEdits:y},x[d]={valType:"number",dflt:null,editType:f||"plot",impliedEdits:m},x[v]={valType:"number",dflt:null,editType:f||"plot",impliedEdits:m},x[g]={valType:"number",dflt:null,editType:"calc",impliedEdits:y},x.colorscale={valType:"colorscale",editType:"calc",dflt:c,impliedEdits:{autocolorscale:!1}},x.autocolorscale={valType:"boolean",dflt:!1!==e.autoColorDflt,editType:"calc",impliedEdits:{colorscale:void 0}},x.reversescale={valType:"boolean",dflt:!1,editType:"plot"},l||(x.showscale={valType:"boolean",dflt:u,editType:"calc"},x.colorbar=n),e.noColorAxis||(x.coloraxis={valType:"subplotid",regex:i("coloraxis"),dflt:null,editType:"calc"}),x}},47128:function(t,e,r){"use strict";var n=r(38248),i=r(3400),a=r(94288).extractOpts;t.exports=function(t,e,r){var o,s=t._fullLayout,l=r.vals,u=r.containerStr,c=u?i.nestedProperty(e,u).get():e,f=a(c),h=!1!==f.auto,p=f.min,d=f.max,v=f.mid,g=function(){return i.aggNums(Math.min,null,l)},y=function(){return i.aggNums(Math.max,null,l)};void 0===p?p=g():h&&(p=c._colorAx&&n(p)?Math.min(p,g()):g()),void 0===d?d=y():h&&(d=c._colorAx&&n(d)?Math.max(d,y()):y()),h&&void 0!==v&&(d-v>v-p?p=v-(d-v):d-v<v-p&&(d=v+(v-p))),p===d&&(p-=.5,d+=.5),f._sync("min",p),f._sync("max",d),f.autocolorscale&&(o=p*d<0?s.colorscale.diverging:p>=0?s.colorscale.sequential:s.colorscale.sequentialminus,f._sync("colorscale",o))}},95504:function(t,e,r){"use strict";var n=r(3400),i=r(94288).hasColorscale,a=r(94288).extractOpts;t.exports=function(t,e){function r(t,e){var r=t["_"+e];void 0!==r&&(t[e]=r)}function o(t,i){var o=i.container?n.nestedProperty(t,i.container).get():t;if(o)if(o.coloraxis)o._colorAx=e[o.coloraxis];else{var s=a(o),l=s.auto;(l||void 0===s.min)&&r(o,i.min),(l||void 0===s.max)&&r(o,i.max),s.autocolorscale&&r(o,"colorscale")}}for(var s=0;s<t.length;s++){var l=t[s],u=l._module.colorbar;if(u)if(Array.isArray(u))for(var c=0;c<u.length;c++)o(l,u[c]);else o(l,u);i(l,"marker.line")&&o(l,{container:"marker.line",min:"cmin",max:"cmax"})}for(var f in e._colorAxes)o(e[f],{min:"cmin",max:"cmax"})}},27260:function(t,e,r){"use strict";var n=r(38248),i=r(3400),a=r(90553),o=r(64013),s=r(88304).isValid,l=r(24040).traceIs;function u(t,e){var r=e.slice(0,e.length-1);return e?i.nestedProperty(t,r).get()||{}:t}t.exports=function t(e,r,c,f,h){var p=h.prefix,d=h.cLetter,v="_module"in r,g=u(e,p),y=u(r,p),m=u(r._template||{},p)||{},x=function(){return delete e.coloraxis,delete r.coloraxis,t(e,r,c,f,h)};if(v){var b=c._colorAxes||{},_=f(p+"coloraxis");if(_){var w=l(r,"contour")&&i.nestedProperty(r,"contours.coloring").get()||"heatmap",T=b[_];return void(T?(T[2].push(x),T[0]!==w&&(T[0]=!1,i.warn(["Ignoring coloraxis:",_,"setting","as it is linked to incompatible colorscales."].join(" ")))):b[_]=[w,r,[x]])}}var k=g[d+"min"],A=g[d+"max"],M=n(k)&&n(A)&&k<A;f(p+d+"auto",!M)?f(p+d+"mid"):(f(p+d+"min"),f(p+d+"max"));var S,E,L=g.colorscale,C=m.colorscale;void 0!==L&&(S=!s(L)),void 0!==C&&(S=!s(C)),f(p+"autocolorscale",S),f(p+"colorscale"),f(p+"reversescale"),"marker.line."!==p&&(p&&v&&(E=a(g)),f(p+"showscale",E)&&(p&&m&&(y._template=m),o(g,y,c)))}},94288:function(t,e,r){"use strict";var n=r(33428),i=r(49760),a=r(38248),o=r(3400),s=r(76308),l=r(88304).isValid,u=["showscale","autocolorscale","colorscale","reversescale","colorbar"],c=["min","max","mid","auto"];function f(t){var e,r,n,i=t._colorAx,a=i||t,o={};for(r=0;r<u.length;r++)o[n=u[r]]=a[n];if(i)for(e="c",r=0;r<c.length;r++)o[n=c[r]]=a["c"+n];else{var s;for(r=0;r<c.length;r++)((s="c"+(n=c[r]))in a||(s="z"+n)in a)&&(o[n]=a[s]);e=s.charAt(0)}return o._sync=function(t,r){var n=-1!==c.indexOf(t)?e+t:t;a[n]=a["_"+n]=r},o}function h(t){for(var e=f(t),r=e.min,n=e.max,i=e.reversescale?p(e.colorscale):e.colorscale,a=i.length,o=new Array(a),s=new Array(a),l=0;l<a;l++){var u=i[l];o[l]=r+u[0]*(n-r),s[l]=u[1]}return{domain:o,range:s}}function p(t){for(var e=t.length,r=new Array(e),n=e-1,i=0;n>=0;n--,i++){var a=t[n];r[i]=[1-a[0],a[1]]}return r}function d(t,e){e=e||{};for(var r=t.domain,o=t.range,l=o.length,u=new Array(l),c=0;c<l;c++){var f=i(o[c]).toRgb();u[c]=[f.r,f.g,f.b,f.a]}var h,p=n.scale.linear().domain(r).range(u).clamp(!0),d=e.noNumericCheck,g=e.returnArray;return(h=d&&g?p:d?function(t){return v(p(t))}:g?function(t){return a(t)?p(t):i(t).isValid()?t:s.defaultLine}:function(t){return a(t)?v(p(t)):i(t).isValid()?t:s.defaultLine}).domain=p.domain,h.range=function(){return o},h}function v(t){var e={r:t[0],g:t[1],b:t[2],a:t[3]};return i(e).toRgbString()}t.exports={hasColorscale:function(t,e,r){var n=e?o.nestedProperty(t,e).get()||{}:t,i=n[r||"color"];i&&i._inputArray&&(i=i._inputArray);var s=!1;if(o.isArrayOrTypedArray(i))for(var u=0;u<i.length;u++)if(a(i[u])){s=!0;break}return o.isPlainObject(n)&&(s||!0===n.showscale||a(n.cmin)&&a(n.cmax)||l(n.colorscale)||o.isPlainObject(n.colorbar))},extractOpts:f,extractScale:h,flipScale:p,makeColorScaleFunc:d,makeColorScaleFuncFromTrace:function(t,e){return d(h(t),e)}}},8932:function(t,e,r){"use strict";var n=r(88304),i=r(94288);t.exports={moduleType:"component",name:"colorscale",attributes:r(49084),layoutAttributes:r(92332),supplyLayoutDefaults:r(51608),handleDefaults:r(27260),crossTraceDefaults:r(95504),calc:r(47128),scales:n.scales,defaultScale:n.defaultScale,getScale:n.get,isValidScale:n.isValid,hasColorscale:i.hasColorscale,extractOpts:i.extractOpts,extractScale:i.extractScale,flipScale:i.flipScale,makeColorScaleFunc:i.makeColorScaleFunc,makeColorScaleFuncFromTrace:i.makeColorScaleFuncFromTrace}},92332:function(t,e,r){"use strict";var n=r(92880).extendFlat,i=r(49084),a=r(88304).scales;t.exports={editType:"calc",colorscale:{editType:"calc",sequential:{valType:"colorscale",dflt:a.Reds,editType:"calc"},sequentialminus:{valType:"colorscale",dflt:a.Blues,editType:"calc"},diverging:{valType:"colorscale",dflt:a.RdBu,editType:"calc"}},coloraxis:n({_isSubplotObj:!0,editType:"calc"},i("",{colorAttr:"corresponding trace color array(s)",noColorAxis:!0,showScaleDflt:!0}))}},51608:function(t,e,r){"use strict";var n=r(3400),i=r(31780),a=r(92332),o=r(27260);t.exports=function(t,e){function r(r,i){return n.coerce(t,e,a,r,i)}r("colorscale.sequential"),r("colorscale.sequentialminus"),r("colorscale.diverging");var s,l,u=e._colorAxes;function c(t,e){return n.coerce(s,l,a.coloraxis,t,e)}for(var f in u){var h=u[f];if(h[0])s=t[f]||{},(l=i.newContainer(e,f,"coloraxis"))._name=f,o(s,l,e,c,{prefix:"",cLetter:"c"});else{for(var p=0;p<h[2].length;p++)h[2][p]();delete e._colorAxes[f]}}}},88304:function(t,e,r){"use strict";var n=r(49760),i={Greys:[[0,"rgb(0,0,0)"],[1,"rgb(255,255,255)"]],YlGnBu:[[0,"rgb(8,29,88)"],[.125,"rgb(37,52,148)"],[.25,"rgb(34,94,168)"],[.375,"rgb(29,145,192)"],[.5,"rgb(65,182,196)"],[.625,"rgb(127,205,187)"],[.75,"rgb(199,233,180)"],[.875,"rgb(237,248,217)"],[1,"rgb(255,255,217)"]],Greens:[[0,"rgb(0,68,27)"],[.125,"rgb(0,109,44)"],[.25,"rgb(35,139,69)"],[.375,"rgb(65,171,93)"],[.5,"rgb(116,196,118)"],[.625,"rgb(161,217,155)"],[.75,"rgb(199,233,192)"],[.875,"rgb(229,245,224)"],[1,"rgb(247,252,245)"]],YlOrRd:[[0,"rgb(128,0,38)"],[.125,"rgb(189,0,38)"],[.25,"rgb(227,26,28)"],[.375,"rgb(252,78,42)"],[.5,"rgb(253,141,60)"],[.625,"rgb(254,178,76)"],[.75,"rgb(254,217,118)"],[.875,"rgb(255,237,160)"],[1,"rgb(255,255,204)"]],Bluered:[[0,"rgb(0,0,255)"],[1,"rgb(255,0,0)"]],RdBu:[[0,"rgb(5,10,172)"],[.35,"rgb(106,137,247)"],[.5,"rgb(190,190,190)"],[.6,"rgb(220,170,132)"],[.7,"rgb(230,145,90)"],[1,"rgb(178,10,28)"]],Reds:[[0,"rgb(220,220,220)"],[.2,"rgb(245,195,157)"],[.4,"rgb(245,160,105)"],[1,"rgb(178,10,28)"]],Blues:[[0,"rgb(5,10,172)"],[.35,"rgb(40,60,190)"],[.5,"rgb(70,100,245)"],[.6,"rgb(90,120,245)"],[.7,"rgb(106,137,247)"],[1,"rgb(220,220,220)"]],Picnic:[[0,"rgb(0,0,255)"],[.1,"rgb(51,153,255)"],[.2,"rgb(102,204,255)"],[.3,"rgb(153,204,255)"],[.4,"rgb(204,204,255)"],[.5,"rgb(255,255,255)"],[.6,"rgb(255,204,255)"],[.7,"rgb(255,153,255)"],[.8,"rgb(255,102,204)"],[.9,"rgb(255,102,102)"],[1,"rgb(255,0,0)"]],Rainbow:[[0,"rgb(150,0,90)"],[.125,"rgb(0,0,200)"],[.25,"rgb(0,25,255)"],[.375,"rgb(0,152,255)"],[.5,"rgb(44,255,150)"],[.625,"rgb(151,255,0)"],[.75,"rgb(255,234,0)"],[.875,"rgb(255,111,0)"],[1,"rgb(255,0,0)"]],Portland:[[0,"rgb(12,51,131)"],[.25,"rgb(10,136,186)"],[.5,"rgb(242,211,56)"],[.75,"rgb(242,143,56)"],[1,"rgb(217,30,30)"]],Jet:[[0,"rgb(0,0,131)"],[.125,"rgb(0,60,170)"],[.375,"rgb(5,255,255)"],[.625,"rgb(255,255,0)"],[.875,"rgb(250,0,0)"],[1,"rgb(128,0,0)"]],Hot:[[0,"rgb(0,0,0)"],[.3,"rgb(230,0,0)"],[.6,"rgb(255,210,0)"],[1,"rgb(255,255,255)"]],Blackbody:[[0,"rgb(0,0,0)"],[.2,"rgb(230,0,0)"],[.4,"rgb(230,210,0)"],[.7,"rgb(255,255,255)"],[1,"rgb(160,200,255)"]],Earth:[[0,"rgb(0,0,130)"],[.1,"rgb(0,180,180)"],[.2,"rgb(40,210,40)"],[.4,"rgb(230,230,50)"],[.6,"rgb(120,70,20)"],[1,"rgb(255,255,255)"]],Electric:[[0,"rgb(0,0,0)"],[.15,"rgb(30,0,100)"],[.4,"rgb(120,0,100)"],[.6,"rgb(160,90,0)"],[.8,"rgb(230,200,0)"],[1,"rgb(255,250,220)"]],Viridis:[[0,"#440154"],[.06274509803921569,"#48186a"],[.12549019607843137,"#472d7b"],[.18823529411764706,"#424086"],[.25098039215686274,"#3b528b"],[.3137254901960784,"#33638d"],[.3764705882352941,"#2c728e"],[.4392156862745098,"#26828e"],[.5019607843137255,"#21918c"],[.5647058823529412,"#1fa088"],[.6274509803921569,"#28ae80"],[.6901960784313725,"#3fbc73"],[.7529411764705882,"#5ec962"],[.8156862745098039,"#84d44b"],[.8784313725490196,"#addc30"],[.9411764705882353,"#d8e219"],[1,"#fde725"]],Cividis:[[0,"rgb(0,32,76)"],[.058824,"rgb(0,42,102)"],[.117647,"rgb(0,52,110)"],[.176471,"rgb(39,63,108)"],[.235294,"rgb(60,74,107)"],[.294118,"rgb(76,85,107)"],[.352941,"rgb(91,95,109)"],[.411765,"rgb(104,106,112)"],[.470588,"rgb(117,117,117)"],[.529412,"rgb(131,129,120)"],[.588235,"rgb(146,140,120)"],[.647059,"rgb(161,152,118)"],[.705882,"rgb(176,165,114)"],[.764706,"rgb(192,177,109)"],[.823529,"rgb(209,191,102)"],[.882353,"rgb(225,204,92)"],[.941176,"rgb(243,219,79)"],[1,"rgb(255,233,69)"]]},a=i.RdBu;function o(t){var e=0;if(!Array.isArray(t)||t.length<2)return!1;if(!t[0]||!t[t.length-1])return!1;if(0!=+t[0][0]||1!=+t[t.length-1][0])return!1;for(var r=0;r<t.length;r++){var i=t[r];if(2!==i.length||+i[0]<e||!n(i[1]).isValid())return!1;e=+i[0]}return!0}t.exports={scales:i,defaultScale:a,get:function(t,e){if(e||(e=a),!t)return e;function r(){try{t=i[t]||JSON.parse(t)}catch(r){t=e}}return"string"==typeof t&&(r(),"string"==typeof t&&r()),o(t)?t:e},isValid:function(t){return void 0!==i[t]||o(t)}}},78316:function(t){"use strict";t.exports=function(t,e,r,n,i){var a=(t-r)/(n-r),o=a+e/(n-r),s=(a+o)/2;return"left"===i||"bottom"===i?a:"center"===i||"middle"===i?s:"right"===i||"top"===i?o:a<2/3-s?a:o>4/3-s?o:s}},67416:function(t,e,r){"use strict";var n=r(3400),i=[["sw-resize","s-resize","se-resize"],["w-resize","move","e-resize"],["nw-resize","n-resize","ne-resize"]];t.exports=function(t,e,r,a){return t="left"===r?0:"center"===r?1:"right"===r?2:n.constrain(Math.floor(3*t),0,2),e="bottom"===a?0:"middle"===a?1:"top"===a?2:n.constrain(Math.floor(3*e),0,2),i[e][t]}},72760:function(t,e){"use strict";e.selectMode=function(t){return"lasso"===t||"select"===t},e.drawMode=function(t){return"drawclosedpath"===t||"drawopenpath"===t||"drawline"===t||"drawrect"===t||"drawcircle"===t},e.openMode=function(t){return"drawline"===t||"drawopenpath"===t},e.rectMode=function(t){return"select"===t||"drawline"===t||"drawrect"===t||"drawcircle"===t},e.freeMode=function(t){return"lasso"===t||"drawclosedpath"===t||"drawopenpath"===t},e.selectingOrDrawing=function(t){return e.freeMode(t)||e.rectMode(t)}},86476:function(t,e,r){"use strict";var n=r(29128),i=r(52264),a=r(89184),o=r(3400).removeElement,s=r(33816),l=t.exports={};l.align=r(78316),l.getCursor=r(67416);var u=r(2616);function c(){var t=document.createElement("div");t.className="dragcover";var e=t.style;return e.position="fixed",e.left=0,e.right=0,e.top=0,e.bottom=0,e.zIndex=999999999,e.background="none",document.body.appendChild(t),t}function f(t){return n(t.changedTouches?t.changedTouches[0]:t,document.body)}l.unhover=u.wrapped,l.unhoverRaw=u.raw,l.init=function(t){var e,r,n,u,h,p,d,v,g=t.gd,y=1,m=g._context.doubleClickDelay,x=t.element;g._mouseDownTime||(g._mouseDownTime=0),x.style.pointerEvents="all",x.onmousedown=_,a?(x._ontouchstart&&x.removeEventListener("touchstart",x._ontouchstart),x._ontouchstart=_,x.addEventListener("touchstart",_,{passive:!1})):x.ontouchstart=_;var b=t.clampFn||function(t,e,r){return Math.abs(t)<r&&(t=0),Math.abs(e)<r&&(e=0),[t,e]};function _(a){g._dragged=!1,g._dragging=!0;var o=f(a);e=o[0],r=o[1],d=a.target,p=a,v=2===a.buttons||a.ctrlKey,void 0===a.clientX&&void 0===a.clientY&&(a.clientX=e,a.clientY=r),(n=(new Date).getTime())-g._mouseDownTime<m?y+=1:(y=1,g._mouseDownTime=n),t.prepFn&&t.prepFn(a,e,r),i&&!v?(h=c()).style.cursor=window.getComputedStyle(x).cursor:i||(h=document,u=window.getComputedStyle(document.documentElement).cursor,document.documentElement.style.cursor=window.getComputedStyle(x).cursor),document.addEventListener("mouseup",T),document.addEventListener("touchend",T),!1!==t.dragmode&&(a.preventDefault(),document.addEventListener("mousemove",w),document.addEventListener("touchmove",w,{passive:!1}))}function w(n){n.preventDefault();var i=f(n),a=t.minDrag||s.MINDRAG,o=b(i[0]-e,i[1]-r,a),u=o[0],c=o[1];(u||c)&&(g._dragged=!0,l.unhover(g,n)),g._dragged&&t.moveFn&&!v&&(g._dragdata={element:x,dx:u,dy:c},t.moveFn(u,c))}function T(e){if(delete g._dragdata,!1!==t.dragmode&&(e.preventDefault(),document.removeEventListener("mousemove",w),document.removeEventListener("touchmove",w)),document.removeEventListener("mouseup",T),document.removeEventListener("touchend",T),i?o(h):u&&(h.documentElement.style.cursor=u,u=null),g._dragging){if(g._dragging=!1,(new Date).getTime()-g._mouseDownTime>m&&(y=Math.max(y-1,1)),g._dragged)t.doneFn&&t.doneFn();else if(t.clickFn&&t.clickFn(y,p),!v){var r;try{r=new MouseEvent("click",e)}catch(t){var n=f(e);(r=document.createEvent("MouseEvents")).initMouseEvent("click",e.bubbles,e.cancelable,e.view,e.detail,e.screenX,e.screenY,n[0],n[1],e.ctrlKey,e.altKey,e.shiftKey,e.metaKey,e.button,e.relatedTarget)}d.dispatchEvent(r)}g._dragging=!1,g._dragged=!1}else g._dragged=!1}},l.coverSlip=c},2616:function(t,e,r){"use strict";var n=r(95924),i=r(91200),a=r(52200).getGraphDiv,o=r(92456),s=t.exports={};s.wrapped=function(t,e,r){(t=a(t))._fullLayout&&i.clear(t._fullLayout._uid+o.HOVERID),s.raw(t,e,r)},s.raw=function(t,e){var r=t._fullLayout,i=t._hoverdata;e||(e={}),e.target&&!t._dragged&&!1===n.triggerHandler(t,"plotly_beforehover",e)||(r._hoverlayer.selectAll("g").remove(),r._hoverlayer.selectAll("line").remove(),r._hoverlayer.selectAll("circle").remove(),t._hoverdata=void 0,e.target&&i&&t.emit("plotly_unhover",{event:e,points:i}))}},98192:function(t,e){"use strict";e.u={valType:"string",values:["solid","dot","dash","longdash","dashdot","longdashdot"],dflt:"solid",editType:"style"},e.c={shape:{valType:"enumerated",values:["","/","\\","x","-","|","+","."],dflt:"",arrayOk:!0,editType:"style"},fillmode:{valType:"enumerated",values:["replace","overlay"],dflt:"replace",editType:"style"},bgcolor:{valType:"color",arrayOk:!0,editType:"style"},fgcolor:{valType:"color",arrayOk:!0,editType:"style"},fgopacity:{valType:"number",editType:"style",min:0,max:1},size:{valType:"number",min:0,dflt:8,arrayOk:!0,editType:"style"},solidity:{valType:"number",min:0,max:1,dflt:.3,arrayOk:!0,editType:"style"},editType:"style"}},43616:function(t,e,r){"use strict";var n=r(33428),i=r(3400),a=i.numberFormat,o=r(38248),s=r(49760),l=r(24040),u=r(76308),c=r(8932),f=i.strTranslate,h=r(72736),p=r(9616),d=r(84284).LINE_SPACING,v=r(13448).DESELECTDIM,g=r(43028),y=r(7152),m=r(10624).appendArrayPointValue,x=t.exports={};function b(t,e,r,n){var i=e.fillpattern,a=e.fillgradient,o=i&&x.getPatternAttr(i.shape,0,"");if(o){var s=x.getPatternAttr(i.bgcolor,0,null),l=x.getPatternAttr(i.fgcolor,0,null),c=i.fgopacity,f=x.getPatternAttr(i.size,0,8),h=x.getPatternAttr(i.solidity,0,.3),p=e.uid;x.pattern(t,"point",r,p,o,f,h,void 0,i.fillmode,s,l,c)}else if(a&&"none"!==a.type){var d,v,g=a.type,y="scatterfill-"+e.uid;n&&(y="legendfill-"+e.uid),n||void 0===a.start&&void 0===a.stop?("horizontal"===g&&(g+="reversed"),t.call(x.gradient,r,y,g,a.colorscale,"fill")):("horizontal"===g?(d={x:a.start,y:0},v={x:a.stop,y:0}):"vertical"===g&&(d={x:0,y:a.start},v={x:0,y:a.stop}),d.x=e._xA.c2p(void 0===d.x?e._extremes.x.min[0].val:d.x,!0),d.y=e._yA.c2p(void 0===d.y?e._extremes.y.min[0].val:d.y,!0),v.x=e._xA.c2p(void 0===v.x?e._extremes.x.max[0].val:v.x,!0),v.y=e._yA.c2p(void 0===v.y?e._extremes.y.max[0].val:v.y,!0),t.call(M,r,y,"linear",a.colorscale,"fill",d,v,!0,!1))}else e.fillcolor&&t.call(u.fill,e.fillcolor)}x.font=function(t,e,r,n,a,o,s){i.isPlainObject(e)&&(s=e.variant,o=e.style,a=e.weight,n=e.color,r=e.size,e=e.family),e&&t.style("font-family",e),r+1&&t.style("font-size",r+"px"),n&&t.call(u.fill,n),a&&t.style("font-weight",a),o&&t.style("font-style",o),s&&t.style("font-variant",s)},x.setPosition=function(t,e,r){t.attr("x",e).attr("y",r)},x.setSize=function(t,e,r){t.attr("width",e).attr("height",r)},x.setRect=function(t,e,r,n,i){t.call(x.setPosition,e,r).call(x.setSize,n,i)},x.translatePoint=function(t,e,r,n){var i=r.c2p(t.x),a=n.c2p(t.y);return!!(o(i)&&o(a)&&e.node())&&("text"===e.node().nodeName?e.attr("x",i).attr("y",a):e.attr("transform",f(i,a)),!0)},x.translatePoints=function(t,e,r){t.each((function(t){var i=n.select(this);x.translatePoint(t,i,e,r)}))},x.hideOutsideRangePoint=function(t,e,r,n,i,a){e.attr("display",r.isPtWithinRange(t,i)&&n.isPtWithinRange(t,a)?null:"none")},x.hideOutsideRangePoints=function(t,e){if(e._hasClipOnAxisFalse){var r=e.xaxis,i=e.yaxis;t.each((function(e){var a=e[0].trace,o=a.xcalendar,s=a.ycalendar,u=l.traceIs(a,"bar-like")?".bartext":".point,.textpoint";t.selectAll(u).each((function(t){x.hideOutsideRangePoint(t,n.select(this),r,i,o,s)}))}))}},x.crispRound=function(t,e,r){return e&&o(e)?t._context.staticPlot?e:e<1?1:Math.round(e):r||0},x.singleLineStyle=function(t,e,r,n,i){e.style("fill","none");var a=(((t||[])[0]||{}).trace||{}).line||{},o=r||a.width||0,s=i||a.dash||"";u.stroke(e,n||a.color),x.dashLine(e,s,o)},x.lineGroupStyle=function(t,e,r,i){t.style("fill","none").each((function(t){var a=(((t||[])[0]||{}).trace||{}).line||{},o=e||a.width||0,s=i||a.dash||"";n.select(this).call(u.stroke,r||a.color).call(x.dashLine,s,o)}))},x.dashLine=function(t,e,r){r=+r||0,e=x.dashStyle(e,r),t.style({"stroke-dasharray":e,"stroke-width":r+"px"})},x.dashStyle=function(t,e){e=+e||1;var r=Math.max(e,3);return"solid"===t?t="":"dot"===t?t=r+"px,"+r+"px":"dash"===t?t=3*r+"px,"+3*r+"px":"longdash"===t?t=5*r+"px,"+5*r+"px":"dashdot"===t?t=3*r+"px,"+r+"px,"+r+"px,"+r+"px":"longdashdot"===t&&(t=5*r+"px,"+2*r+"px,"+r+"px,"+2*r+"px"),t},x.singleFillStyle=function(t,e){var r=n.select(t.node());b(t,((r.data()[0]||[])[0]||{}).trace||{},e,!1)},x.fillGroupStyle=function(t,e,r){t.style("stroke-width",0).each((function(t){var i=n.select(this);t[0].trace&&b(i,t[0].trace,e,r)}))};var _=r(71984);x.symbolNames=[],x.symbolFuncs=[],x.symbolBackOffs=[],x.symbolNeedLines={},x.symbolNoDot={},x.symbolNoFill={},x.symbolList=[],Object.keys(_).forEach((function(t){var e=_[t],r=e.n;x.symbolList.push(r,String(r),t,r+100,String(r+100),t+"-open"),x.symbolNames[r]=t,x.symbolFuncs[r]=e.f,x.symbolBackOffs[r]=e.backoff||0,e.needLine&&(x.symbolNeedLines[r]=!0),e.noDot?x.symbolNoDot[r]=!0:x.symbolList.push(r+200,String(r+200),t+"-dot",r+300,String(r+300),t+"-open-dot"),e.noFill&&(x.symbolNoFill[r]=!0)}));var w=x.symbolNames.length;function T(t,e,r,n){var i=t%100;return x.symbolFuncs[i](e,r,n)+(t>=200?"M0,0.5L0.5,0L0,-0.5L-0.5,0Z":"")}x.symbolNumber=function(t){if(o(t))t=+t;else if("string"==typeof t){var e=0;t.indexOf("-open")>0&&(e=100,t=t.replace("-open","")),t.indexOf("-dot")>0&&(e+=200,t=t.replace("-dot","")),(t=x.symbolNames.indexOf(t))>=0&&(t+=e)}return t%100>=w||t>=400?0:Math.floor(Math.max(t,0))};var k=a("~f"),A={radial:{type:"radial"},radialreversed:{type:"radial",reversed:!0},horizontal:{type:"linear",start:{x:1,y:0},stop:{x:0,y:0}},horizontalreversed:{type:"linear",start:{x:1,y:0},stop:{x:0,y:0},reversed:!0},vertical:{type:"linear",start:{x:0,y:1},stop:{x:0,y:0}},verticalreversed:{type:"linear",start:{x:0,y:1},stop:{x:0,y:0},reversed:!0}};function M(t,e,r,a,o,l,c,f,h,p){var d,v=o.length;"linear"===a?d={node:"linearGradient",attrs:{x1:c.x,y1:c.y,x2:f.x,y2:f.y,gradientUnits:h?"userSpaceOnUse":"objectBoundingBox"},reversed:p}:"radial"===a&&(d={node:"radialGradient",reversed:p});for(var g=new Array(v),y=0;y<v;y++)d.reversed?g[v-1-y]=[k(100*(1-o[y][0])),o[y][1]]:g[y]=[k(100*o[y][0]),o[y][1]];var m=e._fullLayout,x="g"+m._uid+"-"+r,b=m._defs.select(".gradients").selectAll("#"+x).data([a+g.join(";")],i.identity);b.exit().remove(),b.enter().append(d.node).each((function(){var t=n.select(this);d.attrs&&t.attr(d.attrs),t.attr("id",x);var e=t.selectAll("stop").data(g);e.exit().remove(),e.enter().append("stop"),e.each((function(t){var e=s(t[1]);n.select(this).attr({offset:t[0]+"%","stop-color":u.tinyRGB(e),"stop-opacity":e.getAlpha()})}))})),t.style(l,U(x,e)).style(l+"-opacity",null),t.classed("gradient_filled",!0)}x.gradient=function(t,e,r,n,i,a){var o=A[n];return M(t,e,r,o.type,i,a,o.start,o.stop,!1,o.reversed)},x.pattern=function(t,e,r,a,o,l,c,f,h,p,d,v){var g="legend"===e;f&&("overlay"===h?(p=f,d=u.contrast(p)):(p=void 0,d=f));var y,m,x,b,_,w,T,k,A,M=r._fullLayout,S="p"+M._uid+"-"+a,E={},L=s(d),C=u.tinyRGB(L),P=v*L.getAlpha();switch(o){case"/":y=l*Math.sqrt(2),m=l*Math.sqrt(2),w="path",E={d:x="M-"+y/4+","+m/4+"l"+y/2+",-"+m/2+"M0,"+m+"L"+y+",0M"+y/4*3+","+m/4*5+"l"+y/2+",-"+m/2,opacity:P,stroke:C,"stroke-width":(b=c*l)+"px"};break;case"\\":y=l*Math.sqrt(2),m=l*Math.sqrt(2),w="path",E={d:x="M"+y/4*3+",-"+m/4+"l"+y/2+","+m/2+"M0,0L"+y+","+m+"M-"+y/4+","+m/4*3+"l"+y/2+","+m/2,opacity:P,stroke:C,"stroke-width":(b=c*l)+"px"};break;case"x":y=l*Math.sqrt(2),m=l*Math.sqrt(2),x="M-"+y/4+","+m/4+"l"+y/2+",-"+m/2+"M0,"+m+"L"+y+",0M"+y/4*3+","+m/4*5+"l"+y/2+",-"+m/2+"M"+y/4*3+",-"+m/4+"l"+y/2+","+m/2+"M0,0L"+y+","+m+"M-"+y/4+","+m/4*3+"l"+y/2+","+m/2,b=l-l*Math.sqrt(1-c),w="path",E={d:x,opacity:P,stroke:C,"stroke-width":b+"px"};break;case"|":w="path",w="path",E={d:x="M"+(y=l)/2+",0L"+y/2+","+(m=l),opacity:P,stroke:C,"stroke-width":(b=c*l)+"px"};break;case"-":w="path",w="path",E={d:x="M0,"+(m=l)/2+"L"+(y=l)+","+m/2,opacity:P,stroke:C,"stroke-width":(b=c*l)+"px"};break;case"+":w="path",x="M"+(y=l)/2+",0L"+y/2+","+(m=l)+"M0,"+m/2+"L"+y+","+m/2,b=l-l*Math.sqrt(1-c),w="path",E={d:x,opacity:P,stroke:C,"stroke-width":b+"px"};break;case".":y=l,m=l,c<Math.PI/4?_=Math.sqrt(c*l*l/Math.PI):(T=c,k=Math.PI/4,1,_=(A=l/2)+(l/Math.sqrt(2)-A)*(T-k)/(1-k)),w="circle",E={cx:y/2,cy:m/2,r:_,opacity:P,fill:C}}var O=[o||"noSh",p||"noBg",d||"noFg",l,c].join(";"),I=M._defs.select(".patterns").selectAll("#"+S).data([O],i.identity);I.exit().remove(),I.enter().append("pattern").each((function(){var t=n.select(this);if(t.attr({id:S,width:y+"px",height:m+"px",patternUnits:"userSpaceOnUse",patternTransform:g?"scale(0.8)":""}),p){var e=s(p),r=u.tinyRGB(e),i=e.getAlpha(),a=t.selectAll("rect").data([0]);a.exit().remove(),a.enter().append("rect").attr({width:y+"px",height:m+"px",fill:r,"fill-opacity":i})}var o=t.selectAll(w).data([0]);o.exit().remove(),o.enter().append(w).attr(E)})),t.style("fill",U(S,r)).style("fill-opacity",null),t.classed("pattern_filled",!0)},x.initGradients=function(t){var e=t._fullLayout;i.ensureSingle(e._defs,"g","gradients").selectAll("linearGradient,radialGradient").remove(),n.select(t).selectAll(".gradient_filled").classed("gradient_filled",!1)},x.initPatterns=function(t){var e=t._fullLayout;i.ensureSingle(e._defs,"g","patterns").selectAll("pattern").remove(),n.select(t).selectAll(".pattern_filled").classed("pattern_filled",!1)},x.getPatternAttr=function(t,e,r){return t&&i.isArrayOrTypedArray(t)?e<t.length?t[e]:r:t},x.pointStyle=function(t,e,r,i){if(t.size()){var a=x.makePointStyleFns(e);t.each((function(t){x.singlePointStyle(t,n.select(this),e,a,r,i)}))}},x.singlePointStyle=function(t,e,r,n,a,o){var s=r.marker,l=s.line;if(o&&o.i>=0&&void 0===t.i&&(t.i=o.i),e.style("opacity",n.selectedOpacityFn?n.selectedOpacityFn(t):void 0===t.mo?s.opacity:t.mo),n.ms2mrc){var c;c="various"===t.ms||"various"===s.size?3:n.ms2mrc(t.ms),t.mrc=c,n.selectedSizeFn&&(c=t.mrc=n.selectedSizeFn(t));var f=x.symbolNumber(t.mx||s.symbol)||0;t.om=f%200>=100;var h=et(t,r),p=H(t,r);e.attr("d",T(f,c,h,p))}var d,v,g,y=!1;if(t.so)g=l.outlierwidth,v=l.outliercolor,d=s.outliercolor;else{var m=(l||{}).width;g=(t.mlw+1||m+1||(t.trace?(t.trace.marker.line||{}).width:0)+1)-1||0,v="mlc"in t?t.mlcc=n.lineScale(t.mlc):i.isArrayOrTypedArray(l.color)?u.defaultLine:l.color,i.isArrayOrTypedArray(s.color)&&(d=u.defaultLine,y=!0),d="mc"in t?t.mcc=n.markerScale(t.mc):s.color||s.colors||"rgba(0,0,0,0)",n.selectedColorFn&&(d=n.selectedColorFn(t))}if(t.om)e.call(u.stroke,d).style({"stroke-width":(g||1)+"px",fill:"none"});else{e.style("stroke-width",(t.isBlank?0:g)+"px");var b=s.gradient,_=t.mgt;_?y=!0:_=b&&b.type,i.isArrayOrTypedArray(_)&&(_=_[0],A[_]||(_=0));var w=s.pattern,k=w&&x.getPatternAttr(w.shape,t.i,"");if(_&&"none"!==_){var M=t.mgc;M?y=!0:M=b.color;var S=r.uid;y&&(S+="-"+t.i),x.gradient(e,a,S,_,[[0,M],[1,d]],"fill")}else if(k){var E=!1,L=w.fgcolor;!L&&o&&o.color&&(L=o.color,E=!0);var C=x.getPatternAttr(L,t.i,o&&o.color||null),P=x.getPatternAttr(w.bgcolor,t.i,null),O=w.fgopacity,I=x.getPatternAttr(w.size,t.i,8),z=x.getPatternAttr(w.solidity,t.i,.3);E=E||t.mcc||i.isArrayOrTypedArray(w.shape)||i.isArrayOrTypedArray(w.bgcolor)||i.isArrayOrTypedArray(w.fgcolor)||i.isArrayOrTypedArray(w.size)||i.isArrayOrTypedArray(w.solidity);var D=r.uid;E&&(D+="-"+t.i),x.pattern(e,"point",a,D,k,I,z,t.mcc,w.fillmode,P,C,O)}else i.isArrayOrTypedArray(d)?u.fill(e,d[t.i]):u.fill(e,d);g&&u.stroke(e,v)}},x.makePointStyleFns=function(t){var e={},r=t.marker;return e.markerScale=x.tryColorscale(r,""),e.lineScale=x.tryColorscale(r,"line"),l.traceIs(t,"symbols")&&(e.ms2mrc=g.isBubble(t)?y(t):function(){return(r.size||6)/2}),t.selectedpoints&&i.extendFlat(e,x.makeSelectedPointStyleFns(t)),e},x.makeSelectedPointStyleFns=function(t){var e={},r=t.selected||{},n=t.unselected||{},a=t.marker||{},o=r.marker||{},s=n.marker||{},u=a.opacity,c=o.opacity,f=s.opacity,h=void 0!==c,p=void 0!==f;(i.isArrayOrTypedArray(u)||h||p)&&(e.selectedOpacityFn=function(t){var e=void 0===t.mo?a.opacity:t.mo;return t.selected?h?c:e:p?f:v*e});var d=a.color,g=o.color,y=s.color;(g||y)&&(e.selectedColorFn=function(t){var e=t.mcc||d;return t.selected?g||e:y||e});var m=a.size,x=o.size,b=s.size,_=void 0!==x,w=void 0!==b;return l.traceIs(t,"symbols")&&(_||w)&&(e.selectedSizeFn=function(t){var e=t.mrc||m/2;return t.selected?_?x/2:e:w?b/2:e}),e},x.makeSelectedTextStyleFns=function(t){var e={},r=t.selected||{},n=t.unselected||{},i=t.textfont||{},a=r.textfont||{},o=n.textfont||{},s=i.color,l=a.color,c=o.color;return e.selectedTextColorFn=function(t){var e=t.tc||s;return t.selected?l||e:c||(l?e:u.addOpacity(e,v))},e},x.selectedPointStyle=function(t,e){if(t.size()&&e.selectedpoints){var r=x.makeSelectedPointStyleFns(e),i=e.marker||{},a=[];r.selectedOpacityFn&&a.push((function(t,e){t.style("opacity",r.selectedOpacityFn(e))})),r.selectedColorFn&&a.push((function(t,e){u.fill(t,r.selectedColorFn(e))})),r.selectedSizeFn&&a.push((function(t,n){var a=n.mx||i.symbol||0,o=r.selectedSizeFn(n);t.attr("d",T(x.symbolNumber(a),o,et(n,e),H(n,e))),n.mrc2=o})),a.length&&t.each((function(t){for(var e=n.select(this),r=0;r<a.length;r++)a[r](e,t)}))}},x.tryColorscale=function(t,e){var r=e?i.nestedProperty(t,e).get():t;if(r){var n=r.color;if((r.colorscale||r._colorAx)&&i.isArrayOrTypedArray(n))return c.makeColorScaleFuncFromTrace(r)}return i.identity};var S,E,L={start:1,end:-1,middle:0,bottom:1,top:-1};function C(t,e,r,i,a){var o=n.select(t.node().parentNode),s=-1!==e.indexOf("top")?"top":-1!==e.indexOf("bottom")?"bottom":"middle",l=-1!==e.indexOf("left")?"end":-1!==e.indexOf("right")?"start":"middle",u=i?i/.8+1:0,c=(h.lineCount(t)-1)*d+1,p=L[l]*u,v=.75*r+L[s]*u+(L[s]-1)*c*r/2;t.attr("text-anchor",l),a||o.attr("transform",f(p,v))}function P(t,e){var r=t.ts||e.textfont.size;return o(r)&&r>0?r:0}function O(t,e,r){return r&&(t=B(t)),e?z(t[1]):I(t[0])}function I(t){var e=n.round(t,2);return S=e,e}function z(t){var e=n.round(t,2);return E=e,e}function D(t,e,r,n){var i=t[0]-e[0],a=t[1]-e[1],o=r[0]-e[0],s=r[1]-e[1],l=Math.pow(i*i+a*a,.25),u=Math.pow(o*o+s*s,.25),c=(u*u*i-l*l*o)*n,f=(u*u*a-l*l*s)*n,h=3*u*(l+u),p=3*l*(l+u);return[[I(e[0]+(h&&c/h)),z(e[1]+(h&&f/h))],[I(e[0]-(p&&c/p)),z(e[1]-(p&&f/p))]]}x.textPointStyle=function(t,e,r){if(t.size()){var a;if(e.selectedpoints){var o=x.makeSelectedTextStyleFns(e);a=o.selectedTextColorFn}var s=e.texttemplate,l=r._fullLayout;t.each((function(t){var o=n.select(this),u=s?i.extractOption(t,e,"txt","texttemplate"):i.extractOption(t,e,"tx","text");if(u||0===u){if(s){var c=e._module.formatLabels,f=c?c(t,e,l):{},p={};m(p,e,t.i);var d=e._meta||{};u=i.texttemplateString(u,f,l._d3locale,p,t,d)}var v=t.tp||e.textposition,g=P(t,e),y=a?a(t):t.tc||e.textfont.color;o.call(x.font,{family:t.tf||e.textfont.family,weight:t.tw||e.textfont.weight,style:t.ty||e.textfont.style,variant:t.tv||e.textfont.variant,size:g,color:y}).text(u).call(h.convertToTspans,r).call(C,v,g,t.mrc)}else o.remove()}))}},x.selectedTextStyle=function(t,e){if(t.size()&&e.selectedpoints){var r=x.makeSelectedTextStyleFns(e);t.each((function(t){var i=n.select(this),a=r.selectedTextColorFn(t),o=t.tp||e.textposition,s=P(t,e);u.fill(i,a);var c=l.traceIs(e,"bar-like");C(i,o,s,t.mrc2||t.mrc,c)}))}},x.smoothopen=function(t,e){if(t.length<3)return"M"+t.join("L");var r,n="M"+t[0],i=[];for(r=1;r<t.length-1;r++)i.push(D(t[r-1],t[r],t[r+1],e));for(n+="Q"+i[0][0]+" "+t[1],r=2;r<t.length-1;r++)n+="C"+i[r-2][1]+" "+i[r-1][0]+" "+t[r];return n+"Q"+i[t.length-3][1]+" "+t[t.length-1]},x.smoothclosed=function(t,e){if(t.length<3)return"M"+t.join("L")+"Z";var r,n="M"+t[0],i=t.length-1,a=[D(t[i],t[0],t[1],e)];for(r=1;r<i;r++)a.push(D(t[r-1],t[r],t[r+1],e));for(a.push(D(t[i-1],t[i],t[0],e)),r=1;r<=i;r++)n+="C"+a[r-1][1]+" "+a[r][0]+" "+t[r];return n+"C"+a[i][1]+" "+a[0][0]+" "+t[0]+"Z"};var R={hv:function(t,e,r){return"H"+I(e[0])+"V"+O(e,1,r)},vh:function(t,e,r){return"V"+z(e[1])+"H"+O(e,0,r)},hvh:function(t,e,r){return"H"+I((t[0]+e[0])/2)+"V"+z(e[1])+"H"+O(e,0,r)},vhv:function(t,e,r){return"V"+z((t[1]+e[1])/2)+"H"+I(e[0])+"V"+O(e,1,r)}},F=function(t,e,r){return"L"+O(e,0,r)+","+O(e,1,r)};function B(t,e){var r=t.backoff,n=t.trace,a=t.d,o=t.i;if(r&&n&&n.marker&&n.marker.angle%360==0&&n.line&&"spline"!==n.line.shape){var s=i.isArrayOrTypedArray(r),l=t,u=e?e[0]:S||0,c=e?e[1]:E||0,f=l[0],h=l[1],p=f-u,d=h-c,v=Math.atan2(d,p),g=s?r[o]:r;if("auto"===g){var y=l.i;"scatter"===n.type&&y--;var m=l.marker,b=m.symbol;i.isArrayOrTypedArray(b)&&(b=b[y]);var _=m.size;i.isArrayOrTypedArray(_)&&(_=_[y]),g=m?x.symbolBackOffs[x.symbolNumber(b)]*_:0,g+=x.getMarkerStandoff(a[y],n)||0}var w=f-g*Math.cos(v),T=h-g*Math.sin(v);(w<=f&&w>=u||w>=f&&w<=u)&&(T<=h&&T>=c||T>=h&&T<=c)&&(t=[w,T])}return t}x.steps=function(t){var e=R[t]||F;return function(t){for(var r="M"+I(t[0][0])+","+z(t[0][1]),n=t.length,i=1;i<n;i++)r+=e(t[i-1],t[i],i===n-1);return r}},x.applyBackoff=B,x.makeTester=function(){var t=i.ensureSingleById(n.select("body"),"svg","js-plotly-tester",(function(t){t.attr(p.svgAttrs).style({position:"absolute",left:"-10000px",top:"-10000px",width:"9000px",height:"9000px","z-index":"1"})})),e=i.ensureSingle(t,"path","js-reference-point",(function(t){t.attr("d","M0,0H1V1H0Z").style({"stroke-width":0,fill:"black"})}));x.tester=t,x.testref=e},x.savedBBoxes={};var N=0;function j(t){var e=t.getAttribute("data-unformatted");if(null!==e)return e+t.getAttribute("data-math")+t.getAttribute("text-anchor")+t.getAttribute("style")}function U(t,e){if(!t)return null;var r=e._context,n=r._exportedPlot?"":r._baseUrl||"";return n?"url('"+n+"#"+t+"')":"url(#"+t+")"}x.bBox=function(t,e,r){var a,o,s;if(r||(r=j(t)),r){if(a=x.savedBBoxes[r])return i.extendFlat({},a)}else if(1===t.childNodes.length){var l=t.childNodes[0];if(r=j(l)){var u=+l.getAttribute("x")||0,c=+l.getAttribute("y")||0,f=l.getAttribute("transform");if(!f){var p=x.bBox(l,!1,r);return u&&(p.left+=u,p.right+=u),c&&(p.top+=c,p.bottom+=c),p}if(r+="~"+u+"~"+c+"~"+f,a=x.savedBBoxes[r])return i.extendFlat({},a)}}e?o=t:(s=x.tester.node(),o=t.cloneNode(!0),s.appendChild(o)),n.select(o).attr("transform",null).call(h.positionText,0,0);var d=o.getBoundingClientRect(),v=x.testref.node().getBoundingClientRect();e||s.removeChild(o);var g={height:d.height,width:d.width,left:d.left-v.left,top:d.top-v.top,right:d.right-v.left,bottom:d.bottom-v.top};return N>=1e4&&(x.savedBBoxes={},N=0),r&&(x.savedBBoxes[r]=g),N++,i.extendFlat({},g)},x.setClipUrl=function(t,e,r){t.attr("clip-path",U(e,r))},x.getTranslate=function(t){var e=(t[t.attr?"attr":"getAttribute"]("transform")||"").replace(/.*\btranslate\((-?\d*\.?\d*)[^-\d]*(-?\d*\.?\d*)[^\d].*/,(function(t,e,r){return[e,r].join(" ")})).split(" ");return{x:+e[0]||0,y:+e[1]||0}},x.setTranslate=function(t,e,r){var n=t.attr?"attr":"getAttribute",i=t.attr?"attr":"setAttribute",a=t[n]("transform")||"";return e=e||0,r=r||0,a=a.replace(/(\btranslate\(.*?\);?)/,"").trim(),a=(a+=f(e,r)).trim(),t[i]("transform",a),a},x.getScale=function(t){var e=(t[t.attr?"attr":"getAttribute"]("transform")||"").replace(/.*\bscale\((\d*\.?\d*)[^\d]*(\d*\.?\d*)[^\d].*/,(function(t,e,r){return[e,r].join(" ")})).split(" ");return{x:+e[0]||1,y:+e[1]||1}},x.setScale=function(t,e,r){var n=t.attr?"attr":"getAttribute",i=t.attr?"attr":"setAttribute",a=t[n]("transform")||"";return e=e||1,r=r||1,a=a.replace(/(\bscale\(.*?\);?)/,"").trim(),a=(a+="scale("+e+","+r+")").trim(),t[i]("transform",a),a};var V=/\s*sc.*/;x.setPointGroupScale=function(t,e,r){if(e=e||1,r=r||1,t){var n=1===e&&1===r?"":"scale("+e+","+r+")";t.each((function(){var t=(this.getAttribute("transform")||"").replace(V,"");t=(t+=n).trim(),this.setAttribute("transform",t)}))}};var q=/translate\([^)]*\)\s*$/;function H(t,e){var r;return t&&(r=t.mf),void 0===r&&(r=e.marker&&e.marker.standoff||0),e._geo||e._xA?r:-r}x.setTextPointsScale=function(t,e,r){t&&t.each((function(){var t,i=n.select(this),a=i.select("text");if(a.node()){var o=parseFloat(a.attr("x")||0),s=parseFloat(a.attr("y")||0),l=(i.attr("transform")||"").match(q);t=1===e&&1===r?[]:[f(o,s),"scale("+e+","+r+")",f(-o,-s)],l&&t.push(l),i.attr("transform",t.join(""))}}))},x.getMarkerStandoff=H;var G,W,Y,X,Z,K,J=Math.atan2,$=Math.cos,Q=Math.sin;function tt(t,e){var r=e[0],n=e[1];return[r*$(t)-n*Q(t),r*Q(t)+n*$(t)]}function et(t,e){var r,n,a=t.ma;void 0===a&&((a=e.marker.angle)&&!i.isArrayOrTypedArray(a)||(a=0));var s=e.marker.angleref;if("previous"===s||"north"===s){if(e._geo){var l=e._geo.project(t.lonlat);r=l[0],n=l[1]}else{var u=e._xA,c=e._yA;if(!u||!c)return 90;r=u.c2p(t.x),n=c.c2p(t.y)}if(e._geo){var f,h=t.lonlat[0],p=t.lonlat[1],d=e._geo.project([h,p+1e-5]),v=e._geo.project([h+1e-5,p]),g=J(v[1]-n,v[0]-r),y=J(d[1]-n,d[0]-r);if("north"===s)f=a/180*Math.PI;else if("previous"===s){var m=h/180*Math.PI,x=p/180*Math.PI,b=G/180*Math.PI,_=W/180*Math.PI,w=b-m,T=$(_)*Q(w),k=Q(_)*$(x)-$(_)*Q(x)*$(w);f=-J(T,k)-Math.PI,G=h,W=p}var A=tt(g,[$(f),0]),M=tt(y,[Q(f),0]);a=J(A[1]+M[1],A[0]+M[0])/Math.PI*180,"previous"!==s||K===e.uid&&t.i===Z+1||(a=null)}if("previous"===s&&!e._geo)if(K===e.uid&&t.i===Z+1&&o(r)&&o(n)){var S=r-Y,E=n-X,L=e.line&&e.line.shape||"",C=L.slice(L.length-1);"h"===C&&(E=0),"v"===C&&(S=0),a+=J(E,S)/Math.PI*180+90}else a=null}return Y=r,X=n,Z=t.i,K=e.uid,a}x.getMarkerAngle=et},71984:function(t,e,r){"use strict";var n,i,a,o,s=r(21984),l=r(33428).round,u="M0,0Z",c=Math.sqrt(2),f=Math.sqrt(3),h=Math.PI,p=Math.cos,d=Math.sin;function v(t){return null===t}function g(t,e,r){if(!(t&&t%360!=0||e))return r;if(a===t&&o===e&&n===r)return i;function l(t,r){var n=p(t),i=d(t),a=r[0],o=r[1]+(e||0);return[a*n-o*i,a*i+o*n]}a=t,o=e,n=r;for(var u=t/180*h,c=0,f=0,v=s(r),g="",y=0;y<v.length;y++){var m=v[y],x=m[0],b=c,_=f;if("M"===x||"L"===x)c=+m[1],f=+m[2];else if("m"===x||"l"===x)c+=+m[1],f+=+m[2];else if("H"===x)c=+m[1];else if("h"===x)c+=+m[1];else if("V"===x)f=+m[1];else if("v"===x)f+=+m[1];else if("A"===x){c=+m[1],f=+m[2];var w=l(u,[+m[6],+m[7]]);m[6]=w[0],m[7]=w[1],m[3]=+m[3]+t}"H"!==x&&"V"!==x||(x="L"),"h"!==x&&"v"!==x||(x="l"),"m"!==x&&"l"!==x||(c-=b,f-=_);var T=l(u,[c,f]);"H"!==x&&"V"!==x||(x="L"),"M"!==x&&"L"!==x&&"m"!==x&&"l"!==x||(m[1]=T[0],m[2]=T[1]),m[0]=x,g+=m[0]+m.slice(1).join(",")}return i=g,g}t.exports={circle:{n:0,f:function(t,e,r){if(v(e))return u;var n=l(t,2),i="M"+n+",0A"+n+","+n+" 0 1,1 0,-"+n+"A"+n+","+n+" 0 0,1 "+n+",0Z";return r?g(e,r,i):i}},square:{n:1,f:function(t,e,r){if(v(e))return u;var n=l(t,2);return g(e,r,"M"+n+","+n+"H-"+n+"V-"+n+"H"+n+"Z")}},diamond:{n:2,f:function(t,e,r){if(v(e))return u;var n=l(1.3*t,2);return g(e,r,"M"+n+",0L0,"+n+"L-"+n+",0L0,-"+n+"Z")}},cross:{n:3,f:function(t,e,r){if(v(e))return u;var n=l(.4*t,2),i=l(1.2*t,2);return g(e,r,"M"+i+","+n+"H"+n+"V"+i+"H-"+n+"V"+n+"H-"+i+"V-"+n+"H-"+n+"V-"+i+"H"+n+"V-"+n+"H"+i+"Z")}},x:{n:4,f:function(t,e,r){if(v(e))return u;var n=l(.8*t/c,2),i="l"+n+","+n,a="l"+n+",-"+n,o="l-"+n+",-"+n,s="l-"+n+","+n;return g(e,r,"M0,"+n+i+a+o+a+o+s+o+s+i+s+i+"Z")}},"triangle-up":{n:5,f:function(t,e,r){if(v(e))return u;var n=l(2*t/f,2);return g(e,r,"M-"+n+","+l(t/2,2)+"H"+n+"L0,-"+l(t,2)+"Z")}},"triangle-down":{n:6,f:function(t,e,r){if(v(e))return u;var n=l(2*t/f,2);return g(e,r,"M-"+n+",-"+l(t/2,2)+"H"+n+"L0,"+l(t,2)+"Z")}},"triangle-left":{n:7,f:function(t,e,r){if(v(e))return u;var n=l(2*t/f,2);return g(e,r,"M"+l(t/2,2)+",-"+n+"V"+n+"L-"+l(t,2)+",0Z")}},"triangle-right":{n:8,f:function(t,e,r){if(v(e))return u;var n=l(2*t/f,2);return g(e,r,"M-"+l(t/2,2)+",-"+n+"V"+n+"L"+l(t,2)+",0Z")}},"triangle-ne":{n:9,f:function(t,e,r){if(v(e))return u;var n=l(.6*t,2),i=l(1.2*t,2);return g(e,r,"M-"+i+",-"+n+"H"+n+"V"+i+"Z")}},"triangle-se":{n:10,f:function(t,e,r){if(v(e))return u;var n=l(.6*t,2),i=l(1.2*t,2);return g(e,r,"M"+n+",-"+i+"V"+n+"H-"+i+"Z")}},"triangle-sw":{n:11,f:function(t,e,r){if(v(e))return u;var n=l(.6*t,2),i=l(1.2*t,2);return g(e,r,"M"+i+","+n+"H-"+n+"V-"+i+"Z")}},"triangle-nw":{n:12,f:function(t,e,r){if(v(e))return u;var n=l(.6*t,2),i=l(1.2*t,2);return g(e,r,"M-"+n+","+i+"V-"+n+"H"+i+"Z")}},pentagon:{n:13,f:function(t,e,r){if(v(e))return u;var n=l(.951*t,2),i=l(.588*t,2),a=l(-t,2),o=l(-.309*t,2);return g(e,r,"M"+n+","+o+"L"+i+","+l(.809*t,2)+"H-"+i+"L-"+n+","+o+"L0,"+a+"Z")}},hexagon:{n:14,f:function(t,e,r){if(v(e))return u;var n=l(t,2),i=l(t/2,2),a=l(t*f/2,2);return g(e,r,"M"+a+",-"+i+"V"+i+"L0,"+n+"L-"+a+","+i+"V-"+i+"L0,-"+n+"Z")}},hexagon2:{n:15,f:function(t,e,r){if(v(e))return u;var n=l(t,2),i=l(t/2,2),a=l(t*f/2,2);return g(e,r,"M-"+i+","+a+"H"+i+"L"+n+",0L"+i+",-"+a+"H-"+i+"L-"+n+",0Z")}},octagon:{n:16,f:function(t,e,r){if(v(e))return u;var n=l(.924*t,2),i=l(.383*t,2);return g(e,r,"M-"+i+",-"+n+"H"+i+"L"+n+",-"+i+"V"+i+"L"+i+","+n+"H-"+i+"L-"+n+","+i+"V-"+i+"Z")}},star:{n:17,f:function(t,e,r){if(v(e))return u;var n=1.4*t,i=l(.225*n,2),a=l(.951*n,2),o=l(.363*n,2),s=l(.588*n,2),c=l(-n,2),f=l(-.309*n,2),h=l(.118*n,2),p=l(.809*n,2);return g(e,r,"M"+i+","+f+"H"+a+"L"+o+","+h+"L"+s+","+p+"L0,"+l(.382*n,2)+"L-"+s+","+p+"L-"+o+","+h+"L-"+a+","+f+"H-"+i+"L0,"+c+"Z")}},hexagram:{n:18,f:function(t,e,r){if(v(e))return u;var n=l(.66*t,2),i=l(.38*t,2),a=l(.76*t,2);return g(e,r,"M-"+a+",0l-"+i+",-"+n+"h"+a+"l"+i+",-"+n+"l"+i+","+n+"h"+a+"l-"+i+","+n+"l"+i+","+n+"h-"+a+"l-"+i+","+n+"l-"+i+",-"+n+"h-"+a+"Z")}},"star-triangle-up":{n:19,f:function(t,e,r){if(v(e))return u;var n=l(t*f*.8,2),i=l(.8*t,2),a=l(1.6*t,2),o=l(4*t,2),s="A "+o+","+o+" 0 0 1 ";return g(e,r,"M-"+n+","+i+s+n+","+i+s+"0,-"+a+s+"-"+n+","+i+"Z")}},"star-triangle-down":{n:20,f:function(t,e,r){if(v(e))return u;var n=l(t*f*.8,2),i=l(.8*t,2),a=l(1.6*t,2),o=l(4*t,2),s="A "+o+","+o+" 0 0 1 ";return g(e,r,"M"+n+",-"+i+s+"-"+n+",-"+i+s+"0,"+a+s+n+",-"+i+"Z")}},"star-square":{n:21,f:function(t,e,r){if(v(e))return u;var n=l(1.1*t,2),i=l(2*t,2),a="A "+i+","+i+" 0 0 1 ";return g(e,r,"M-"+n+",-"+n+a+"-"+n+","+n+a+n+","+n+a+n+",-"+n+a+"-"+n+",-"+n+"Z")}},"star-diamond":{n:22,f:function(t,e,r){if(v(e))return u;var n=l(1.4*t,2),i=l(1.9*t,2),a="A "+i+","+i+" 0 0 1 ";return g(e,r,"M-"+n+",0"+a+"0,"+n+a+n+",0"+a+"0,-"+n+a+"-"+n+",0Z")}},"diamond-tall":{n:23,f:function(t,e,r){if(v(e))return u;var n=l(.7*t,2),i=l(1.4*t,2);return g(e,r,"M0,"+i+"L"+n+",0L0,-"+i+"L-"+n+",0Z")}},"diamond-wide":{n:24,f:function(t,e,r){if(v(e))return u;var n=l(1.4*t,2),i=l(.7*t,2);return g(e,r,"M0,"+i+"L"+n+",0L0,-"+i+"L-"+n+",0Z")}},hourglass:{n:25,f:function(t,e,r){if(v(e))return u;var n=l(t,2);return g(e,r,"M"+n+","+n+"H-"+n+"L"+n+",-"+n+"H-"+n+"Z")},noDot:!0},bowtie:{n:26,f:function(t,e,r){if(v(e))return u;var n=l(t,2);return g(e,r,"M"+n+","+n+"V-"+n+"L-"+n+","+n+"V-"+n+"Z")},noDot:!0},"circle-cross":{n:27,f:function(t,e,r){if(v(e))return u;var n=l(t,2);return g(e,r,"M0,"+n+"V-"+n+"M"+n+",0H-"+n+"M"+n+",0A"+n+","+n+" 0 1,1 0,-"+n+"A"+n+","+n+" 0 0,1 "+n+",0Z")},needLine:!0,noDot:!0},"circle-x":{n:28,f:function(t,e,r){if(v(e))return u;var n=l(t,2),i=l(t/c,2);return g(e,r,"M"+i+","+i+"L-"+i+",-"+i+"M"+i+",-"+i+"L-"+i+","+i+"M"+n+",0A"+n+","+n+" 0 1,1 0,-"+n+"A"+n+","+n+" 0 0,1 "+n+",0Z")},needLine:!0,noDot:!0},"square-cross":{n:29,f:function(t,e,r){if(v(e))return u;var n=l(t,2);return g(e,r,"M0,"+n+"V-"+n+"M"+n+",0H-"+n+"M"+n+","+n+"H-"+n+"V-"+n+"H"+n+"Z")},needLine:!0,noDot:!0},"square-x":{n:30,f:function(t,e,r){if(v(e))return u;var n=l(t,2);return g(e,r,"M"+n+","+n+"L-"+n+",-"+n+"M"+n+",-"+n+"L-"+n+","+n+"M"+n+","+n+"H-"+n+"V-"+n+"H"+n+"Z")},needLine:!0,noDot:!0},"diamond-cross":{n:31,f:function(t,e,r){if(v(e))return u;var n=l(1.3*t,2);return g(e,r,"M"+n+",0L0,"+n+"L-"+n+",0L0,-"+n+"ZM0,-"+n+"V"+n+"M-"+n+",0H"+n)},needLine:!0,noDot:!0},"diamond-x":{n:32,f:function(t,e,r){if(v(e))return u;var n=l(1.3*t,2),i=l(.65*t,2);return g(e,r,"M"+n+",0L0,"+n+"L-"+n+",0L0,-"+n+"ZM-"+i+",-"+i+"L"+i+","+i+"M-"+i+","+i+"L"+i+",-"+i)},needLine:!0,noDot:!0},"cross-thin":{n:33,f:function(t,e,r){if(v(e))return u;var n=l(1.4*t,2);return g(e,r,"M0,"+n+"V-"+n+"M"+n+",0H-"+n)},needLine:!0,noDot:!0,noFill:!0},"x-thin":{n:34,f:function(t,e,r){if(v(e))return u;var n=l(t,2);return g(e,r,"M"+n+","+n+"L-"+n+",-"+n+"M"+n+",-"+n+"L-"+n+","+n)},needLine:!0,noDot:!0,noFill:!0},asterisk:{n:35,f:function(t,e,r){if(v(e))return u;var n=l(1.2*t,2),i=l(.85*t,2);return g(e,r,"M0,"+n+"V-"+n+"M"+n+",0H-"+n+"M"+i+","+i+"L-"+i+",-"+i+"M"+i+",-"+i+"L-"+i+","+i)},needLine:!0,noDot:!0,noFill:!0},hash:{n:36,f:function(t,e,r){if(v(e))return u;var n=l(t/2,2),i=l(t,2);return g(e,r,"M"+n+","+i+"V-"+i+"M"+(n-i)+",-"+i+"V"+i+"M"+i+","+n+"H-"+i+"M-"+i+","+(n-i)+"H"+i)},needLine:!0,noFill:!0},"y-up":{n:37,f:function(t,e,r){if(v(e))return u;var n=l(1.2*t,2),i=l(1.6*t,2),a=l(.8*t,2);return g(e,r,"M-"+n+","+a+"L0,0M"+n+","+a+"L0,0M0,-"+i+"L0,0")},needLine:!0,noDot:!0,noFill:!0},"y-down":{n:38,f:function(t,e,r){if(v(e))return u;var n=l(1.2*t,2),i=l(1.6*t,2),a=l(.8*t,2);return g(e,r,"M-"+n+",-"+a+"L0,0M"+n+",-"+a+"L0,0M0,"+i+"L0,0")},needLine:!0,noDot:!0,noFill:!0},"y-left":{n:39,f:function(t,e,r){if(v(e))return u;var n=l(1.2*t,2),i=l(1.6*t,2),a=l(.8*t,2);return g(e,r,"M"+a+","+n+"L0,0M"+a+",-"+n+"L0,0M-"+i+",0L0,0")},needLine:!0,noDot:!0,noFill:!0},"y-right":{n:40,f:function(t,e,r){if(v(e))return u;var n=l(1.2*t,2),i=l(1.6*t,2),a=l(.8*t,2);return g(e,r,"M-"+a+","+n+"L0,0M-"+a+",-"+n+"L0,0M"+i+",0L0,0")},needLine:!0,noDot:!0,noFill:!0},"line-ew":{n:41,f:function(t,e,r){if(v(e))return u;var n=l(1.4*t,2);return g(e,r,"M"+n+",0H-"+n)},needLine:!0,noDot:!0,noFill:!0},"line-ns":{n:42,f:function(t,e,r){if(v(e))return u;var n=l(1.4*t,2);return g(e,r,"M0,"+n+"V-"+n)},needLine:!0,noDot:!0,noFill:!0},"line-ne":{n:43,f:function(t,e,r){if(v(e))return u;var n=l(t,2);return g(e,r,"M"+n+",-"+n+"L-"+n+","+n)},needLine:!0,noDot:!0,noFill:!0},"line-nw":{n:44,f:function(t,e,r){if(v(e))return u;var n=l(t,2);return g(e,r,"M"+n+","+n+"L-"+n+",-"+n)},needLine:!0,noDot:!0,noFill:!0},"arrow-up":{n:45,f:function(t,e,r){if(v(e))return u;var n=l(t,2);return g(e,r,"M0,0L-"+n+","+l(2*t,2)+"H"+n+"Z")},backoff:1,noDot:!0},"arrow-down":{n:46,f:function(t,e,r){if(v(e))return u;var n=l(t,2);return g(e,r,"M0,0L-"+n+",-"+l(2*t,2)+"H"+n+"Z")},noDot:!0},"arrow-left":{n:47,f:function(t,e,r){if(v(e))return u;var n=l(2*t,2),i=l(t,2);return g(e,r,"M0,0L"+n+",-"+i+"V"+i+"Z")},noDot:!0},"arrow-right":{n:48,f:function(t,e,r){if(v(e))return u;var n=l(2*t,2),i=l(t,2);return g(e,r,"M0,0L-"+n+",-"+i+"V"+i+"Z")},noDot:!0},"arrow-bar-up":{n:49,f:function(t,e,r){if(v(e))return u;var n=l(t,2);return g(e,r,"M-"+n+",0H"+n+"M0,0L-"+n+","+l(2*t,2)+"H"+n+"Z")},backoff:1,needLine:!0,noDot:!0},"arrow-bar-down":{n:50,f:function(t,e,r){if(v(e))return u;var n=l(t,2);return g(e,r,"M-"+n+",0H"+n+"M0,0L-"+n+",-"+l(2*t,2)+"H"+n+"Z")},needLine:!0,noDot:!0},"arrow-bar-left":{n:51,f:function(t,e,r){if(v(e))return u;var n=l(2*t,2),i=l(t,2);return g(e,r,"M0,-"+i+"V"+i+"M0,0L"+n+",-"+i+"V"+i+"Z")},needLine:!0,noDot:!0},"arrow-bar-right":{n:52,f:function(t,e,r){if(v(e))return u;var n=l(2*t,2),i=l(t,2);return g(e,r,"M0,-"+i+"V"+i+"M0,0L-"+n+",-"+i+"V"+i+"Z")},needLine:!0,noDot:!0},arrow:{n:53,f:function(t,e,r){if(v(e))return u;var n=h/2.5,i=2*t*p(n),a=2*t*d(n);return g(e,r,"M0,0L"+-i+","+a+"L"+i+","+a+"Z")},backoff:.9,noDot:!0},"arrow-wide":{n:54,f:function(t,e,r){if(v(e))return u;var n=h/4,i=2*t*p(n),a=2*t*d(n);return g(e,r,"M0,0L"+-i+","+a+"A "+2*t+","+2*t+" 0 0 1 "+i+","+a+"Z")},backoff:.4,noDot:!0}}},97644:function(t){"use strict";t.exports={visible:{valType:"boolean",editType:"calc"},type:{valType:"enumerated",values:["percent","constant","sqrt","data"],editType:"calc"},symmetric:{valType:"boolean",editType:"calc"},array:{valType:"data_array",editType:"calc"},arrayminus:{valType:"data_array",editType:"calc"},value:{valType:"number",min:0,dflt:10,editType:"calc"},valueminus:{valType:"number",min:0,dflt:10,editType:"calc"},traceref:{valType:"integer",min:0,dflt:0,editType:"style"},tracerefminus:{valType:"integer",min:0,dflt:0,editType:"style"},copy_ystyle:{valType:"boolean",editType:"plot"},copy_zstyle:{valType:"boolean",editType:"style"},color:{valType:"color",editType:"style"},thickness:{valType:"number",min:0,dflt:2,editType:"style"},width:{valType:"number",min:0,editType:"plot"},editType:"calc",_deprecated:{opacity:{valType:"number",editType:"style"}}}},14880:function(t,e,r){"use strict";var n=r(38248),i=r(24040),a=r(54460),o=r(3400),s=r(93792);function l(t,e,r,i){var l=e["error_"+i]||{},u=[];if(l.visible&&-1!==["linear","log"].indexOf(r.type)){for(var c=s(l),f=0;f<t.length;f++){var h=t[f],p=h.i;if(void 0===p)p=f;else if(null===p)continue;var d=h[i];if(n(r.c2l(d))){var v=c(d,p);if(n(v[0])&&n(v[1])){var g=h[i+"s"]=d-v[0],y=h[i+"h"]=d+v[1];u.push(g,y)}}}var m=r._id,x=e._extremes[m],b=a.findExtremes(r,u,o.extendFlat({tozero:x.opts.tozero},{padded:!0}));x.min=x.min.concat(b.min),x.max=x.max.concat(b.max)}}t.exports=function(t){for(var e=t.calcdata,r=0;r<e.length;r++){var n=e[r],o=n[0].trace;if(!0===o.visible&&i.traceIs(o,"errorBarsOK")){var s=a.getFromId(t,o.xaxis),u=a.getFromId(t,o.yaxis);l(n,o,s,"x"),l(n,o,u,"y")}}}},93792:function(t){"use strict";function e(t,e){return"percent"===t?function(t){return Math.abs(t*e/100)}:"constant"===t?function(){return Math.abs(e)}:"sqrt"===t?function(t){return Math.sqrt(Math.abs(t))}:void 0}t.exports=function(t){var r=t.type,n=t.symmetric;if("data"===r){var i=t.array||[];if(n)return function(t,e){var r=+i[e];return[r,r]};var a=t.arrayminus||[];return function(t,e){var r=+i[e],n=+a[e];return isNaN(r)&&isNaN(n)?[NaN,NaN]:[n||0,r||0]}}var o=e(r,t.value),s=e(r,t.valueminus);return n||void 0===t.valueminus?function(t){var e=o(t);return[e,e]}:function(t){return[s(t),o(t)]}}},65200:function(t,e,r){"use strict";var n=r(38248),i=r(24040),a=r(3400),o=r(31780),s=r(97644);t.exports=function(t,e,r,l){var u="error_"+l.axis,c=o.newContainer(e,u),f=t[u]||{};function h(t,e){return a.coerce(f,c,s,t,e)}if(!1!==h("visible",void 0!==f.array||void 0!==f.value||"sqrt"===f.type)){var p=h("type","array"in f?"data":"percent"),d=!0;"sqrt"!==p&&(d=h("symmetric",!(("data"===p?"arrayminus":"valueminus")in f))),"data"===p?(h("array"),h("traceref"),d||(h("arrayminus"),h("tracerefminus"))):"percent"!==p&&"constant"!==p||(h("value"),d||h("valueminus"));var v="copy_"+l.inherit+"style";l.inherit&&(e["error_"+l.inherit]||{}).visible&&h(v,!(f.color||n(f.thickness)||n(f.width))),l.inherit&&c[v]||(h("color",r),h("thickness"),h("width",i.traceIs(e,"gl3d")?0:4))}}},64968:function(t,e,r){"use strict";var n=r(3400),i=r(67824).overrideAll,a=r(97644),o={error_x:n.extendFlat({},a),error_y:n.extendFlat({},a)};delete o.error_x.copy_zstyle,delete o.error_y.copy_zstyle,delete o.error_y.copy_ystyle;var s={error_x:n.extendFlat({},a),error_y:n.extendFlat({},a),error_z:n.extendFlat({},a)};delete s.error_x.copy_ystyle,delete s.error_y.copy_ystyle,delete s.error_z.copy_ystyle,delete s.error_z.copy_zstyle,t.exports={moduleType:"component",name:"errorbars",schema:{traces:{scatter:o,bar:o,histogram:o,scatter3d:i(s,"calc","nested"),scattergl:i(o,"calc","nested")}},supplyDefaults:r(65200),calc:r(14880),makeComputeError:r(93792),plot:r(78512),style:r(92036),hoverInfo:function(t,e,r){(e.error_y||{}).visible&&(r.yerr=t.yh-t.y,e.error_y.symmetric||(r.yerrneg=t.y-t.ys)),(e.error_x||{}).visible&&(r.xerr=t.xh-t.x,e.error_x.symmetric||(r.xerrneg=t.x-t.xs))}}},78512:function(t,e,r){"use strict";var n=r(33428),i=r(38248),a=r(43616),o=r(43028);t.exports=function(t,e,r,s){var l=r.xaxis,u=r.yaxis,c=s&&s.duration>0,f=t._context.staticPlot;e.each((function(e){var h,p=e[0].trace,d=p.error_x||{},v=p.error_y||{};p.ids&&(h=function(t){return t.id});var g=o.hasMarkers(p)&&p.marker.maxdisplayed>0;v.visible||d.visible||(e=[]);var y=n.select(this).selectAll("g.errorbar").data(e,h);if(y.exit().remove(),e.length){d.visible||y.selectAll("path.xerror").remove(),v.visible||y.selectAll("path.yerror").remove(),y.style("opacity",1);var m=y.enter().append("g").classed("errorbar",!0);c&&m.style("opacity",0).transition().duration(s.duration).style("opacity",1),a.setClipUrl(y,r.layerClipId,t),y.each((function(t){var e=n.select(this),r=function(t,e,r){var n={x:e.c2p(t.x),y:r.c2p(t.y)};return void 0!==t.yh&&(n.yh=r.c2p(t.yh),n.ys=r.c2p(t.ys),i(n.ys)||(n.noYS=!0,n.ys=r.c2p(t.ys,!0))),void 0!==t.xh&&(n.xh=e.c2p(t.xh),n.xs=e.c2p(t.xs),i(n.xs)||(n.noXS=!0,n.xs=e.c2p(t.xs,!0))),n}(t,l,u);if(!g||t.vis){var a,o=e.select("path.yerror");if(v.visible&&i(r.x)&&i(r.yh)&&i(r.ys)){var h=v.width;a="M"+(r.x-h)+","+r.yh+"h"+2*h+"m-"+h+",0V"+r.ys,r.noYS||(a+="m-"+h+",0h"+2*h),o.size()?c&&(o=o.transition().duration(s.duration).ease(s.easing)):o=e.append("path").style("vector-effect",f?"none":"non-scaling-stroke").classed("yerror",!0),o.attr("d",a)}else o.remove();var p=e.select("path.xerror");if(d.visible&&i(r.y)&&i(r.xh)&&i(r.xs)){var y=(d.copy_ystyle?v:d).width;a="M"+r.xh+","+(r.y-y)+"v"+2*y+"m0,-"+y+"H"+r.xs,r.noXS||(a+="m0,-"+y+"v"+2*y),p.size()?c&&(p=p.transition().duration(s.duration).ease(s.easing)):p=e.append("path").style("vector-effect",f?"none":"non-scaling-stroke").classed("xerror",!0),p.attr("d",a)}else p.remove()}}))}}))}},92036:function(t,e,r){"use strict";var n=r(33428),i=r(76308);t.exports=function(t){t.each((function(t){var e=t[0].trace,r=e.error_y||{},a=e.error_x||{},o=n.select(this);o.selectAll("path.yerror").style("stroke-width",r.thickness+"px").call(i.stroke,r.color),a.copy_ystyle&&(a=r),o.selectAll("path.xerror").style("stroke-width",a.thickness+"px").call(i.stroke,a.color)}))}},55756:function(t,e,r){"use strict";var n=r(25376),i=r(65460).hoverlabel,a=r(92880).extendFlat;t.exports={hoverlabel:{bgcolor:a({},i.bgcolor,{arrayOk:!0}),bordercolor:a({},i.bordercolor,{arrayOk:!0}),font:n({arrayOk:!0,editType:"none"}),align:a({},i.align,{arrayOk:!0}),namelength:a({},i.namelength,{arrayOk:!0}),editType:"none"}}},55056:function(t,e,r){"use strict";var n=r(3400),i=r(24040);function a(t,e,r,i){i=i||n.identity,Array.isArray(t)&&(e[0][r]=i(t))}t.exports=function(t){var e=t.calcdata,r=t._fullLayout;function o(t){return function(e){return n.coerceHoverinfo({hoverinfo:e},{_module:t._module},r)}}for(var s=0;s<e.length;s++){var l=e[s],u=l[0].trace;if(!i.traceIs(u,"pie-like")){var c=i.traceIs(u,"2dMap")?a:n.fillArray;c(u.hoverinfo,l,"hi",o(u)),u.hovertemplate&&c(u.hovertemplate,l,"ht"),u.hoverlabel&&(c(u.hoverlabel.bgcolor,l,"hbg"),c(u.hoverlabel.bordercolor,l,"hbc"),c(u.hoverlabel.font.size,l,"hts"),c(u.hoverlabel.font.color,l,"htc"),c(u.hoverlabel.font.family,l,"htf"),c(u.hoverlabel.font.weight,l,"htw"),c(u.hoverlabel.font.style,l,"hty"),c(u.hoverlabel.font.variant,l,"htv"),c(u.hoverlabel.namelength,l,"hnl"),c(u.hoverlabel.align,l,"hta"))}}}},62376:function(t,e,r){"use strict";var n=r(24040),i=r(83292).hover;t.exports=function(t,e,r){var a=n.getComponentMethod("annotations","onClick")(t,t._hoverdata);function o(){t.emit("plotly_click",{points:t._hoverdata,event:e})}void 0!==r&&i(t,e,r,!0),t._hoverdata&&e&&e.target&&(a&&a.then?a.then(o):o(),e.stopImmediatePropagation&&e.stopImmediatePropagation())}},92456:function(t){"use strict";t.exports={YANGLE:60,HOVERARROWSIZE:6,HOVERTEXTPAD:3,HOVERFONTSIZE:13,HOVERFONT:"Arial, sans-serif",HOVERMINTIME:50,HOVERID:"-hover"}},95448:function(t,e,r){"use strict";var n=r(3400),i=r(55756),a=r(16132);t.exports=function(t,e,r,o){var s=n.extendFlat({},o.hoverlabel);e.hovertemplate&&(s.namelength=-1),a(t,e,(function(r,a){return n.coerce(t,e,i,r,a)}),s)}},10624:function(t,e,r){"use strict";var n=r(3400);e.getSubplot=function(t){return t.subplot||t.xaxis+t.yaxis||t.geo},e.isTraceInSubplots=function(t,r){if("splom"===t.type){for(var n=t.xaxes||[],i=t.yaxes||[],a=0;a<n.length;a++)for(var o=0;o<i.length;o++)if(-1!==r.indexOf(n[a]+i[o]))return!0;return!1}return-1!==r.indexOf(e.getSubplot(t))},e.flat=function(t,e){for(var r=new Array(t.length),n=0;n<t.length;n++)r[n]=e;return r},e.p2c=function(t,e){for(var r=new Array(t.length),n=0;n<t.length;n++)r[n]=t[n].p2c(e);return r},e.getDistanceFunction=function(t,r,n,i){return"closest"===t?i||e.quadrature(r,n):"x"===t.charAt(0)?r:n},e.getClosest=function(t,e,r){if(!1!==r.index)r.index>=0&&r.index<t.length?r.distance=0:r.index=!1;else for(var n=0;n<t.length;n++){var i=e(t[n]);i<=r.distance&&(r.index=n,r.distance=i)}return r},e.inbox=function(t,e,r){return t*e<0||0===t?r:1/0},e.quadrature=function(t,e){return function(r){var n=t(r),i=e(r);return Math.sqrt(n*n+i*i)}},e.makeEventData=function(t,r,n){var i="index"in t?t.index:t.pointNumber,a={data:r._input,fullData:r,curveNumber:r.index,pointNumber:i};if(r._indexToPoints){var o=r._indexToPoints[i];1===o.length?a.pointIndex=o[0]:a.pointIndices=o}else a.pointIndex=i;return r._module.eventData?a=r._module.eventData(a,t,r,n,i):("xVal"in t?a.x=t.xVal:"x"in t&&(a.x=t.x),"yVal"in t?a.y=t.yVal:"y"in t&&(a.y=t.y),t.xa&&(a.xaxis=t.xa),t.ya&&(a.yaxis=t.ya),void 0!==t.zLabelVal&&(a.z=t.zLabelVal)),e.appendArrayPointValue(a,r,i),a},e.appendArrayPointValue=function(t,e,r){var i=e._arrayAttrs;if(i)for(var s=0;s<i.length;s++){var l=i[s],u=a(l);if(void 0===t[u]){var c=o(n.nestedProperty(e,l).get(),r);void 0!==c&&(t[u]=c)}}},e.appendArrayMultiPointValues=function(t,e,r){var i=e._arrayAttrs;if(i)for(var s=0;s<i.length;s++){var l=i[s],u=a(l);if(void 0===t[u]){for(var c=n.nestedProperty(e,l).get(),f=new Array(r.length),h=0;h<r.length;h++)f[h]=o(c,r[h]);t[u]=f}}};var i={ids:"id",locations:"location",labels:"label",values:"value","marker.colors":"color",parents:"parent"};function a(t){return i[t]||t}function o(t,e){return Array.isArray(e)?Array.isArray(t)&&Array.isArray(t[e[0]])?t[e[0]][e[1]]:void 0:t[e]}var s={x:!0,y:!0},l={"x unified":!0,"y unified":!0};e.isUnifiedHover=function(t){return"string"==typeof t&&!!l[t]},e.isXYhover=function(t){return"string"==typeof t&&!!s[t]}},83292:function(t,e,r){"use strict";var n=r(33428),i=r(38248),a=r(49760),o=r(3400),s=o.pushUnique,l=o.strTranslate,u=o.strRotate,c=r(95924),f=r(72736),h=r(72213),p=r(43616),d=r(76308),v=r(86476),g=r(54460),y=r(24040),m=r(10624),x=r(92456),b=r(77864),_=r(31140),w=x.YANGLE,T=Math.PI*w/180,k=1/Math.sin(T),A=Math.cos(T),M=Math.sin(T),S=x.HOVERARROWSIZE,E=x.HOVERTEXTPAD,L={box:!0,ohlc:!0,violin:!0,candlestick:!0},C={scatter:!0,scattergl:!0,splom:!0};function P(t,e){return t.distance-e.distance}function O(t){return[t.trace.index,t.index,t.x0,t.y0,t.name,t.attr,t.xa?t.xa._id:"",t.ya?t.ya._id:""].join(",")}e.hover=function(t,e,r,a){t=o.getGraphDiv(t);var l=e.target;o.throttle(t._fullLayout._uid+x.HOVERID,x.HOVERMINTIME,(function(){!function(t,e,r,a,l){r||(r="xy");var u,f,p,x=Array.isArray(r)?r:[r],b=t._fullLayout,_=b.hoversubplots,w=b._plots||[],T=w[r],A=b._has("cartesian"),M=e.hovermode||b.hovermode,E="x"===(M||"").charAt(0),I="y"===(M||"").charAt(0);if(A&&(E||I)&&"axis"===_)for(var D=x.length,U=0;U<D;U++)if(w[u=x[U]]){f=g.getFromId(t,u,"x"),p=g.getFromId(t,u,"y");var G=(E?f:p)._subplotsWith;if(G&&G.length)for(var W=0;W<G.length;W++)s(x,G[W])}if(T&&"single"!==_){var Y=T.overlays.map((function(t){return t.id}));x=x.concat(Y)}for(var X=x.length,Z=new Array(X),K=new Array(X),J=!1,$=0;$<X;$++)if(w[u=x[$]])J=!0,Z[$]=w[u].xaxis,K[$]=w[u].yaxis;else{if(!b[u]||!b[u]._subplot)return void o.warn("Unrecognized subplot: "+u);var Q=b[u]._subplot;Z[$]=Q.xaxis,K[$]=Q.yaxis}if(M&&!J&&(M="closest"),-1===["x","y","closest","x unified","y unified"].indexOf(M)||!t.calcdata||t.querySelector(".zoombox")||t._dragging)return v.unhoverRaw(t,e);var tt=b.hoverdistance;-1===tt&&(tt=1/0);var et=b.spikedistance;-1===et&&(et=1/0);var rt,nt,it,at,ot,st,lt,ut,ct,ft,ht,pt,dt,vt=[],gt=[],yt={hLinePoint:null,vLinePoint:null},mt=!1;if(Array.isArray(e))for(M="array",it=0;it<e.length;it++)(ot=t.calcdata[e[it].curveNumber||0])&&(st=ot[0].trace,"skip"!==ot[0].trace.hoverinfo&&(gt.push(ot),"h"===st.orientation&&(mt=!0)));else{var xt,bt,_t=t.calcdata.slice();for(_t.sort((function(t,e){return(t[0].trace.zorder||0)-(e[0].trace.zorder||0)})),at=0;at<_t.length;at++)ot=_t[at],"skip"!==(st=ot[0].trace).hoverinfo&&m.isTraceInSubplots(st,x)&&(gt.push(ot),"h"===st.orientation&&(mt=!0));if(l){if(!1===c.triggerHandler(t,"plotly_beforehover",e))return;var wt=l.getBoundingClientRect();xt=e.clientX-wt.left,bt=e.clientY-wt.top,b._calcInverseTransform(t);var Tt=o.apply3DTransform(b._invTransform)(xt,bt);if(xt=Tt[0],bt=Tt[1],xt<0||xt>Z[0]._length||bt<0||bt>K[0]._length)return v.unhoverRaw(t,e)}else xt="xpx"in e?e.xpx:Z[0]._length/2,bt="ypx"in e?e.ypx:K[0]._length/2;if(e.pointerX=xt+Z[0]._offset,e.pointerY=bt+K[0]._offset,rt="xval"in e?m.flat(x,e.xval):m.p2c(Z,xt),nt="yval"in e?m.flat(x,e.yval):m.p2c(K,bt),!i(rt[0])||!i(nt[0]))return o.warn("Fx.hover failed",e,t),v.unhoverRaw(t,e)}var kt=1/0;function At(r,n){for(at=0;at<gt.length;at++)if((ot=gt[at])&&ot[0]&&ot[0].trace&&!0===(st=ot[0].trace).visible&&0!==st._length&&-1===["carpet","contourcarpet"].indexOf(st._module.name)){if(ct=M,m.isUnifiedHover(ct)&&(ct=ct.charAt(0)),"splom"===st.type?lt=x[ut=0]:(lt=m.getSubplot(st),ut=x.indexOf(lt)),pt={cd:ot,trace:st,xa:Z[ut],ya:K[ut],maxHoverDistance:tt,maxSpikeDistance:et,index:!1,distance:Math.min(kt,tt),spikeDistance:1/0,xSpike:void 0,ySpike:void 0,color:d.defaultLine,name:st.name,x0:void 0,x1:void 0,y0:void 0,y1:void 0,xLabelVal:void 0,yLabelVal:void 0,zLabelVal:void 0,text:void 0},b[lt]&&(pt.subplot=b[lt]._subplot),b._splomScenes&&b._splomScenes[st.uid]&&(pt.scene=b._splomScenes[st.uid]),"array"===ct){var a=e[at];"pointNumber"in a?(pt.index=a.pointNumber,ct="closest"):(ct="","xval"in a&&(ft=a.xval,ct="x"),"yval"in a&&(ht=a.yval,ct=ct?"closest":"y"))}else void 0!==r&&void 0!==n?(ft=r,ht=n):(ft=rt[ut],ht=nt[ut]);if(dt=vt.length,0!==tt)if(st._module&&st._module.hoverPoints){var s=st._module.hoverPoints(pt,ft,ht,ct,{finiteRange:!0,hoverLayer:b._hoverlayer,hoversubplots:_,gd:t});if(s)for(var l,u=0;u<s.length;u++)l=s[u],i(l.x0)&&i(l.y0)&&vt.push(B(l,M))}else o.log("Unrecognized trace type in hover:",st);if("closest"===M&&vt.length>dt&&(vt.splice(0,dt),kt=vt[0].distance),A&&0!==et&&0===vt.length){pt.distance=et,pt.index=!1;var c=st._module.hoverPoints(pt,ft,ht,"closest",{hoverLayer:b._hoverlayer});if(c&&(c=c.filter((function(t){return t.spikeDistance<=et}))),c&&c.length){var f,h=c.filter((function(t){return t.xa.showspikes&&"hovered data"!==t.xa.spikesnap}));if(h.length){var p=h[0];i(p.x0)&&i(p.y0)&&(f=St(p),(!yt.vLinePoint||yt.vLinePoint.spikeDistance>f.spikeDistance)&&(yt.vLinePoint=f))}var v=c.filter((function(t){return t.ya.showspikes&&"hovered data"!==t.ya.spikesnap}));if(v.length){var g=v[0];i(g.x0)&&i(g.y0)&&(f=St(g),(!yt.hLinePoint||yt.hLinePoint.spikeDistance>f.spikeDistance)&&(yt.hLinePoint=f))}}}}}function Mt(t,e,r){for(var n,i=null,a=1/0,o=0;o<t.length;o++)f&&f._id!==t[o].xa._id||p&&p._id!==t[o].ya._id||(n=t[o].spikeDistance,r&&0===o&&(n=-1/0),n<=a&&n<=e&&(i=t[o],a=n));return i}function St(t){return t?{xa:t.xa,ya:t.ya,x:void 0!==t.xSpike?t.xSpike:(t.x0+t.x1)/2,y:void 0!==t.ySpike?t.ySpike:(t.y0+t.y1)/2,distance:t.distance,spikeDistance:t.spikeDistance,curveNumber:t.trace.index,color:t.color,pointNumber:t.index}:null}At();var Et={fullLayout:b,container:b._hoverlayer,event:e},Lt=t._spikepoints,Ct={vLinePoint:yt.vLinePoint,hLinePoint:yt.hLinePoint};t._spikepoints=Ct;var Pt=function(){var t=vt.filter((function(t){return f&&f._id===t.xa._id&&p&&p._id===t.ya._id})),e=vt.filter((function(t){return!(f&&f._id===t.xa._id&&p&&p._id===t.ya._id)}));t.sort(P),e.sort(P),vt=function(t,e){for(var r=e.charAt(0),n=[],i=[],a=[],o=0;o<t.length;o++){var s=t[o];y.traceIs(s.trace,"bar-like")||y.traceIs(s.trace,"box-violin")?a.push(s):s.trace[r+"period"]?i.push(s):n.push(s)}return n.concat(i).concat(a)}(vt=t.concat(e),M)};Pt();var Ot=M.charAt(0),It=("x"===Ot||"y"===Ot)&&vt[0]&&C[vt[0].trace.type];if(A&&0!==et&&0!==vt.length){var zt=Mt(vt.filter((function(t){return t.ya.showspikes})),et,It);yt.hLinePoint=St(zt);var Dt=Mt(vt.filter((function(t){return t.xa.showspikes})),et,It);yt.vLinePoint=St(Dt)}if(0===vt.length){var Rt=v.unhoverRaw(t,e);return!A||null===yt.hLinePoint&&null===yt.vLinePoint||j(Lt)&&N(t,yt,Et),Rt}if(A&&j(Lt)&&N(t,yt,Et),m.isXYhover(ct)&&0!==vt[0].length&&"splom"!==vt[0].trace.type){var Ft=vt[0],Bt=(vt=L[Ft.trace.type]?vt.filter((function(t){return t.trace.index===Ft.trace.index})):[Ft]).length;At(V("x",Ft,b),V("y",Ft,b));var Nt,jt=[],Ut={},Vt=0,qt=function(t){var e=L[t.trace.type]?O(t):t.trace.index;if(Ut[e]){var r=Ut[e]-1,n=jt[r];r>0&&Math.abs(t.distance)<Math.abs(n.distance)&&(jt[r]=t)}else Vt++,Ut[e]=Vt,jt.push(t)};for(Nt=0;Nt<Bt;Nt++)qt(vt[Nt]);for(Nt=vt.length-1;Nt>Bt-1;Nt--)qt(vt[Nt]);vt=jt,Pt()}var Ht=t._hoverdata,Gt=[],Wt=q(t),Yt=H(t);for(it=0;it<vt.length;it++){var Xt=vt[it],Zt=m.makeEventData(Xt,Xt.trace,Xt.cd);if(!1!==Xt.hovertemplate){var Kt=!1;Xt.cd[Xt.index]&&Xt.cd[Xt.index].ht&&(Kt=Xt.cd[Xt.index].ht),Xt.hovertemplate=Kt||Xt.trace.hovertemplate||!1}if(Xt.xa&&Xt.ya){var Jt=Xt.x0+Xt.xa._offset,$t=Xt.x1+Xt.xa._offset,Qt=Xt.y0+Xt.ya._offset,te=Xt.y1+Xt.ya._offset,ee=Math.min(Jt,$t),re=Math.max(Jt,$t),ne=Math.min(Qt,te),ie=Math.max(Qt,te);Zt.bbox={x0:ee+Yt,x1:re+Yt,y0:ne+Wt,y1:ie+Wt}}Xt.eventData=[Zt],Gt.push(Zt)}t._hoverdata=Gt;var ae="y"===M&&(gt.length>1||vt.length>1)||"closest"===M&&mt&&vt.length>1,oe=d.combine(b.plot_bgcolor||d.background,b.paper_bgcolor),se=z(vt,{gd:t,hovermode:M,rotateLabels:ae,bgColor:oe,container:b._hoverlayer,outerContainer:b._paper.node(),commonLabelOpts:b.hoverlabel,hoverdistance:b.hoverdistance}),le=se.hoverLabels;if(m.isUnifiedHover(M)||(function(t,e,r,n){var i,a,o,s,l,u,c,f=e?"xa":"ya",h=e?"ya":"xa",p=0,d=1,v=t.size(),g=new Array(v),y=0,m=n.minX,x=n.maxX,b=n.minY,_=n.maxY,w=function(t){return t*r._invScaleX},T=function(t){return t*r._invScaleY};function A(t){var e=t[0],r=t[t.length-1];if(a=e.pmin-e.pos-e.dp+e.size,o=r.pos+r.dp+r.size-e.pmax,a>.01){for(l=t.length-1;l>=0;l--)t[l].dp+=a;i=!1}if(!(o<.01)){if(a<-.01){for(l=t.length-1;l>=0;l--)t[l].dp-=o;i=!1}if(i){var n=0;for(s=0;s<t.length;s++)(u=t[s]).pos+u.dp+u.size>e.pmax&&n++;for(s=t.length-1;s>=0&&!(n<=0);s--)(u=t[s]).pos>e.pmax-1&&(u.del=!0,n--);for(s=0;s<t.length&&!(n<=0);s++)if((u=t[s]).pos<e.pmin+1)for(u.del=!0,n--,o=2*u.size,l=t.length-1;l>=0;l--)t[l].dp-=o;for(s=t.length-1;s>=0&&!(n<=0);s--)(u=t[s]).pos+u.dp+u.size>e.pmax&&(u.del=!0,n--)}}}for(t.each((function(t){var n=t[f],i=t[h],a="x"===n._id.charAt(0),o=n.range;0===y&&o&&o[0]>o[1]!==a&&(d=-1);var s=0,l=a?r.width:r.height;if("x"===r.hovermode||"y"===r.hovermode){var u,c,p=R(t,e),v=t.anchor,A="end"===v?-1:1;if("middle"===v)c=(u=t.crossPos+(a?T(p.y-t.by/2):w(t.bx/2+t.tx2width/2)))+(a?T(t.by):w(t.bx));else if(a)c=(u=t.crossPos+T(S+p.y)-T(t.by/2-S))+T(t.by);else{var M=w(A*S+p.x),E=M+w(A*t.bx);u=t.crossPos+Math.min(M,E),c=t.crossPos+Math.max(M,E)}a?void 0!==b&&void 0!==_&&Math.min(c,_)-Math.max(u,b)>1&&("left"===i.side?(s=i._mainLinePosition,l=r.width):l=i._mainLinePosition):void 0!==m&&void 0!==x&&Math.min(c,x)-Math.max(u,m)>1&&("top"===i.side?(s=i._mainLinePosition,l=r.height):l=i._mainLinePosition)}g[y++]=[{datum:t,traceIndex:t.trace.index,dp:0,pos:t.pos,posref:t.posref,size:t.by*(a?k:1)/2,pmin:s,pmax:l}]})),g.sort((function(t,e){return t[0].posref-e[0].posref||d*(e[0].traceIndex-t[0].traceIndex)}));!i&&p<=v;){for(p++,i=!0,s=0;s<g.length-1;){var M=g[s],E=g[s+1],L=M[M.length-1],C=E[0];if((a=L.pos+L.dp+L.size-C.pos-C.dp+C.size)>.01&&L.pmin===C.pmin&&L.pmax===C.pmax){for(l=E.length-1;l>=0;l--)E[l].dp+=a;for(M.push.apply(M,E),g.splice(s+1,1),c=0,l=M.length-1;l>=0;l--)c+=M[l].dp;for(o=c/M.length,l=M.length-1;l>=0;l--)M[l].dp-=o;i=!1}else s++}g.forEach(A)}for(s=g.length-1;s>=0;s--){var P=g[s];for(l=P.length-1;l>=0;l--){var O=P[l],I=O.datum;I.offset=O.dp,I.del=O.del}}}(le,ae,b,se.commonLabelBoundingBox),F(le,ae,b._invScaleX,b._invScaleY)),l&&l.tagName){var ue=y.getComponentMethod("annotations","hasClickToShow")(t,Gt);h(n.select(l),ue?"pointer":"")}l&&!a&&function(t,e,r){if(!r||r.length!==t._hoverdata.length)return!0;for(var n=r.length-1;n>=0;n--){var i=r[n],a=t._hoverdata[n];if(i.curveNumber!==a.curveNumber||String(i.pointNumber)!==String(a.pointNumber)||String(i.pointNumbers)!==String(a.pointNumbers))return!0}return!1}(t,0,Ht)&&(Ht&&t.emit("plotly_unhover",{event:e,points:Ht}),t.emit("plotly_hover",{event:e,points:t._hoverdata,xaxes:Z,yaxes:K,xvals:rt,yvals:nt}))}(t,e,r,a,l)}))},e.loneHover=function(t,e){var r=!0;Array.isArray(t)||(r=!1,t=[t]);var i=e.gd,a=q(i),o=H(i),s=z(t.map((function(t){var r=t._x0||t.x0||t.x||0,n=t._x1||t.x1||t.x||0,s=t._y0||t.y0||t.y||0,l=t._y1||t.y1||t.y||0,u=t.eventData;if(u){var c=Math.min(r,n),f=Math.max(r,n),h=Math.min(s,l),p=Math.max(s,l),v=t.trace;if(y.traceIs(v,"gl3d")){var g=i._fullLayout[v.scene]._scene.container,m=g.offsetLeft,x=g.offsetTop;c+=m,f+=m,h+=x,p+=x}u.bbox={x0:c+o,x1:f+o,y0:h+a,y1:p+a},e.inOut_bbox&&e.inOut_bbox.push(u.bbox)}else u=!1;return{color:t.color||d.defaultLine,x0:t.x0||t.x||0,x1:t.x1||t.x||0,y0:t.y0||t.y||0,y1:t.y1||t.y||0,xLabel:t.xLabel,yLabel:t.yLabel,zLabel:t.zLabel,text:t.text,name:t.name,idealAlign:t.idealAlign,borderColor:t.borderColor,fontFamily:t.fontFamily,fontSize:t.fontSize,fontColor:t.fontColor,fontWeight:t.fontWeight,fontStyle:t.fontStyle,fontVariant:t.fontVariant,nameLength:t.nameLength,textAlign:t.textAlign,trace:t.trace||{index:0,hoverinfo:""},xa:{_offset:0},ya:{_offset:0},index:0,hovertemplate:t.hovertemplate||!1,hovertemplateLabels:t.hovertemplateLabels||!1,eventData:u}})),{gd:i,hovermode:"closest",rotateLabels:!1,bgColor:e.bgColor||d.background,container:n.select(e.container),outerContainer:e.outerContainer||e.container}).hoverLabels,l=0,u=0;return s.sort((function(t,e){return t.y0-e.y0})).each((function(t,r){var n=t.y0-t.by/2;t.offset=n-5<l?l-n+5:0,l=n+t.by+t.offset,r===e.anchorIndex&&(u=t.offset)})).each((function(t){t.offset-=u})),F(s,!1,i._fullLayout._invScaleX,i._fullLayout._invScaleY),r?s:s.node()};var I=/<extra>([\s\S]*)<\/extra>/;function z(t,e){var r=e.gd,i=r._fullLayout,a=e.hovermode,s=e.rotateLabels,c=e.bgColor,h=e.container,v=e.outerContainer,g=e.commonLabelOpts||{};if(0===t.length)return[[]];var T=e.fontFamily||x.HOVERFONT,k=e.fontSize||x.HOVERFONTSIZE,A=e.fontWeight||i.font.weight,M=e.fontStyle||i.font.style,L=e.fontVariant||i.font.variant,C=t[0],P=C.xa,I=C.ya,z=a.charAt(0),R=z+"Label",F=C[R];if(void 0===F&&"multicategory"===P.type)for(var B=0;B<t.length&&void 0===(F=t[B][R]);B++);var N=G(r,v),j=N.top,U=N.width,V=N.height,q=void 0!==F&&C.distance<=e.hoverdistance&&("x"===a||"y"===a);if(q){var H,W,Y=!0;for(H=0;H<t.length;H++)if(Y&&void 0===t[H].zLabel&&(Y=!1),W=t[H].hoverinfo||t[H].trace.hoverinfo){var X=Array.isArray(W)?W:W.split("+");if(-1===X.indexOf("all")&&-1===X.indexOf(a)){q=!1;break}}Y&&(q=!1)}var Z=h.selectAll("g.axistext").data(q?[0]:[]);Z.enter().append("g").classed("axistext",!0),Z.exit().remove();var K={minX:0,maxX:0,minY:0,maxY:0};if(Z.each((function(){var t=n.select(this),e=o.ensureSingle(t,"path","",(function(t){t.style({"stroke-width":"1px"})})),s=o.ensureSingle(t,"text","",(function(t){t.attr("data-notex",1)})),u=g.bgcolor||d.defaultLine,c=g.bordercolor||d.contrast(u),h=d.contrast(u),v={weight:g.font.weight||A,style:g.font.style||M,variant:g.font.variant||L,family:g.font.family||T,size:g.font.size||k,color:g.font.color||h};e.style({fill:u,stroke:c}),s.text(F).call(p.font,v).call(f.positionText,0,0).call(f.convertToTspans,r),t.attr("transform","");var y,m,x=G(r,s.node());if("x"===a){var b="top"===P.side?"-":"";s.attr("text-anchor","middle").call(f.positionText,0,"top"===P.side?j-x.bottom-S-E:j-x.top+S+E),y=P._offset+(C.x0+C.x1)/2,m=I._offset+("top"===P.side?0:I._length);var _=x.width/2+E,w=y;y<_?w=_:y>i.width-_&&(w=i.width-_),e.attr("d","M"+(y-w)+",0L"+(y-w+S)+","+b+S+"H"+_+"v"+b+(2*E+x.height)+"H"+-_+"V"+b+S+"H"+(y-w-S)+"Z"),y=w,K.minX=y-_,K.maxX=y+_,"top"===P.side?(K.minY=m-(2*E+x.height),K.maxY=m-E):(K.minY=m+E,K.maxY=m+(2*E+x.height))}else{var O,z,D;"right"===I.side?(O="start",z=1,D="",y=P._offset+P._length):(O="end",z=-1,D="-",y=P._offset),m=I._offset+(C.y0+C.y1)/2,s.attr("text-anchor",O),e.attr("d","M0,0L"+D+S+","+S+"V"+(E+x.height/2)+"h"+D+(2*E+x.width)+"V-"+(E+x.height/2)+"H"+D+S+"V-"+S+"Z"),K.minY=m-(E+x.height/2),K.maxY=m+(E+x.height/2),"right"===I.side?(K.minX=y+S,K.maxX=y+S+(2*E+x.width)):(K.minX=y-S-(2*E+x.width),K.maxX=y-S);var R,B=x.height/2,N=j-x.top-B,U="clip"+i._uid+"commonlabel"+I._id;if(y<x.width+2*E+S){R="M-"+(S+E)+"-"+B+"h-"+(x.width-E)+"V"+B+"h"+(x.width-E)+"Z";var V=x.width-y+E;f.positionText(s,V,N),"end"===O&&s.selectAll("tspan").each((function(){var t=n.select(this),e=p.tester.append("text").text(t.text()).call(p.font,v),i=G(r,e.node());Math.round(i.width)<Math.round(x.width)&&t.attr("x",V-i.width),e.remove()}))}else f.positionText(s,z*(E+S),N),R=null;var q=i._topclips.selectAll("#"+U).data(R?[0]:[]);q.enter().append("clipPath").attr("id",U).append("path"),q.exit().remove(),q.select("path").attr("d",R),p.setClipUrl(s,R?U:null,r)}t.attr("transform",l(y,m))})),m.isUnifiedHover(a)){h.selectAll("g.hovertext").remove();var J=t.filter((function(t){return"none"!==t.hoverinfo}));if(0===J.length)return[];var $=i.hoverlabel,Q=$.font,tt={showlegend:!0,legend:{title:{text:F,font:Q},font:Q,bgcolor:$.bgcolor,bordercolor:$.bordercolor,borderwidth:1,tracegroupgap:7,traceorder:i.legend?i.legend.traceorder:void 0,orientation:"v"}},et={font:Q};b(tt,et,r._fullData);var rt=et.legend;rt.entries=[];for(var nt=0;nt<J.length;nt++){var it=J[nt];if("none"!==it.hoverinfo){var at=D(it,!0,a,i,F),ot=at[0],st=at[1];it.name=st,it.text=""!==st?st+" : "+ot:ot;var lt=it.cd[it.index];lt&&(lt.mc&&(it.mc=lt.mc),lt.mcc&&(it.mc=lt.mcc),lt.mlc&&(it.mlc=lt.mlc),lt.mlcc&&(it.mlc=lt.mlcc),lt.mlw&&(it.mlw=lt.mlw),lt.mrc&&(it.mrc=lt.mrc),lt.dir&&(it.dir=lt.dir)),it._distinct=!0,rt.entries.push([it])}}rt.entries.sort((function(t,e){return t[0].trace.index-e[0].trace.index})),rt.layer=h,rt._inHover=!0,rt._groupTitleFont=$.grouptitlefont,_(r,rt);var ut,ct,ft,ht,pt=h.select("g.legend"),dt=G(r,pt.node()),vt=dt.width+2*E,gt=dt.height+2*E,yt=J[0],mt=(yt.x0+yt.x1)/2,xt=(yt.y0+yt.y1)/2,bt=!(y.traceIs(yt.trace,"bar-like")||y.traceIs(yt.trace,"box-violin"));"y"===z?bt?(ct=xt-E,ut=xt+E):(ct=Math.min.apply(null,J.map((function(t){return Math.min(t.y0,t.y1)}))),ut=Math.max.apply(null,J.map((function(t){return Math.max(t.y0,t.y1)})))):ct=ut=o.mean(J.map((function(t){return(t.y0+t.y1)/2})))-gt/2,"x"===z?bt?(ft=mt+E,ht=mt-E):(ft=Math.max.apply(null,J.map((function(t){return Math.max(t.x0,t.x1)}))),ht=Math.min.apply(null,J.map((function(t){return Math.min(t.x0,t.x1)})))):ft=ht=o.mean(J.map((function(t){return(t.x0+t.x1)/2})))-vt/2;var _t,wt,Tt=P._offset,kt=I._offset;return ht+=Tt-vt,ct+=kt-gt,_t=(ft+=Tt)+vt<U&&ft>=0?ft:ht+vt<U&&ht>=0?ht:Tt+vt<U?Tt:ft-mt<mt-ht+vt?U-vt:0,_t+=E,wt=(ut+=kt)+gt<V&&ut>=0?ut:ct+gt<V&&ct>=0?ct:kt+gt<V?kt:ut-xt<xt-ct+gt?V-gt:0,wt+=E,pt.attr("transform",l(_t-1,wt-1)),pt}var At=h.selectAll("g.hovertext").data(t,(function(t){return O(t)}));return At.enter().append("g").classed("hovertext",!0).each((function(){var t=n.select(this);t.append("rect").call(d.fill,d.addOpacity(c,.8)),t.append("text").classed("name",!0),t.append("path").style("stroke-width","1px"),t.append("text").classed("nums",!0).call(p.font,{weight:A,style:M,variant:L,family:T,size:k})})),At.exit().remove(),At.each((function(t){var e=n.select(this).attr("transform",""),o=t.color;Array.isArray(o)&&(o=o[t.eventData[0].pointNumber]);var h=t.bgcolor||o,v=d.combine(d.opacity(h)?h:d.defaultLine,c),g=d.combine(d.opacity(o)?o:d.defaultLine,c),y=t.borderColor||d.contrast(v),m=D(t,q,a,i,F,e),x=m[0],b=m[1],_=e.select("text.nums").call(p.font,{family:t.fontFamily||T,size:t.fontSize||k,color:t.fontColor||y,weight:t.fontWeight||A,style:t.fontStyle||M,variant:t.fontVariant||L}).text(x).attr("data-notex",1).call(f.positionText,0,0).call(f.convertToTspans,r),C=e.select("text.name"),P=0,O=0;if(b&&b!==x){C.call(p.font,{family:t.fontFamily||T,size:t.fontSize||k,color:g,weight:t.fontWeight||A,style:t.fontStyle||M,variant:t.fontVariant||L}).text(b).attr("data-notex",1).call(f.positionText,0,0).call(f.convertToTspans,r);var I=G(r,C.node());P=I.width+2*E,O=I.height+2*E}else C.remove(),e.select("rect").remove();e.select("path").style({fill:v,stroke:y});var z=t.xa._offset+(t.x0+t.x1)/2,R=t.ya._offset+(t.y0+t.y1)/2,B=Math.abs(t.x1-t.x0),N=Math.abs(t.y1-t.y0),H=G(r,_.node()),W=H.width/i._invScaleX,Y=H.height/i._invScaleY;t.ty0=(j-H.top)/i._invScaleY,t.bx=W+2*E,t.by=Math.max(Y+2*E,O),t.anchor="start",t.txwidth=W,t.tx2width=P,t.offset=0;var X,Z,K=(W+S+E+P)*i._invScaleX;if(s)t.pos=z,X=R+N/2+K<=V,Z=R-N/2-K>=0,"top"!==t.idealAlign&&X||!Z?X?(R+=N/2,t.anchor="start"):t.anchor="middle":(R-=N/2,t.anchor="end"),t.crossPos=R;else{if(t.pos=R,X=z+B/2+K<=U,Z=z-B/2-K>=0,"left"!==t.idealAlign&&X||!Z)if(X)z+=B/2,t.anchor="start";else{t.anchor="middle";var J=K/2,$=z+J-U,Q=z-J;$>0&&(z-=$),Q<0&&(z+=-Q)}else z-=B/2,t.anchor="end";t.crossPos=z}_.attr("text-anchor",t.anchor),P&&C.attr("text-anchor",t.anchor),e.attr("transform",l(z,R)+(s?u(w):""))})),{hoverLabels:At,commonLabelBoundingBox:K}}function D(t,e,r,n,i,a){var s="",l="";void 0!==t.nameOverride&&(t.name=t.nameOverride),t.name&&(t.trace._meta&&(t.name=o.templateString(t.name,t.trace._meta)),s=U(t.name,t.nameLength));var u=r.charAt(0),c="x"===u?"y":"x";void 0!==t.zLabel?(void 0!==t.xLabel&&(l+="x: "+t.xLabel+"<br>"),void 0!==t.yLabel&&(l+="y: "+t.yLabel+"<br>"),"choropleth"!==t.trace.type&&"choroplethmapbox"!==t.trace.type&&(l+=(l?"z: ":"")+t.zLabel)):e&&t[u+"Label"]===i?l=t[c+"Label"]||"":void 0===t.xLabel?void 0!==t.yLabel&&"scattercarpet"!==t.trace.type&&(l=t.yLabel):l=void 0===t.yLabel?t.xLabel:"("+t.xLabel+", "+t.yLabel+")",!t.text&&0!==t.text||Array.isArray(t.text)||(l+=(l?"<br>":"")+t.text),void 0!==t.extraText&&(l+=(l?"<br>":"")+t.extraText),a&&""===l&&!t.hovertemplate&&(""===s&&a.remove(),l=s);var f=t.hovertemplate||!1;if(f){var h=t.hovertemplateLabels||t;t[u+"Label"]!==i&&(h[u+"other"]=h[u+"Val"],h[u+"otherLabel"]=h[u+"Label"]),l=(l=o.hovertemplateString(f,h,n._d3locale,t.eventData[0]||{},t.trace._meta)).replace(I,(function(e,r){return s=U(r,t.nameLength),""}))}return[l,s]}function R(t,e){var r=0,n=t.offset;return e&&(n*=-M,r=t.offset*A),{x:r,y:n}}function F(t,e,r,i){var a=function(t){return t*r},o=function(t){return t*i};t.each((function(t){var r=n.select(this);if(t.del)return r.remove();var i,s,l,u,c=r.select("text.nums"),h=t.anchor,d="end"===h?-1:1,v=(u=(l=(s={start:1,end:-1,middle:0}[(i=t).anchor])*(S+E))+s*(i.txwidth+E),"middle"===i.anchor&&(l-=i.tx2width/2,u+=i.txwidth/2+E),{alignShift:s,textShiftX:l,text2ShiftX:u}),g=R(t,e),y=g.x,m=g.y,x="middle"===h;r.select("path").attr("d",x?"M-"+a(t.bx/2+t.tx2width/2)+","+o(m-t.by/2)+"h"+a(t.bx)+"v"+o(t.by)+"h-"+a(t.bx)+"Z":"M0,0L"+a(d*S+y)+","+o(S+m)+"v"+o(t.by/2-S)+"h"+a(d*t.bx)+"v-"+o(t.by)+"H"+a(d*S+y)+"V"+o(m-S)+"Z");var b=y+v.textShiftX,_=m+t.ty0-t.by/2+E,w=t.textAlign||"auto";"auto"!==w&&("left"===w&&"start"!==h?(c.attr("text-anchor","start"),b=x?-t.bx/2-t.tx2width/2+E:-t.bx-E):"right"===w&&"end"!==h&&(c.attr("text-anchor","end"),b=x?t.bx/2-t.tx2width/2-E:t.bx+E)),c.call(f.positionText,a(b),o(_)),t.tx2width&&(r.select("text.name").call(f.positionText,a(v.text2ShiftX+v.alignShift*E+y),o(m+t.ty0-t.by/2+E)),r.select("rect").call(p.setRect,a(v.text2ShiftX+(v.alignShift-1)*t.tx2width/2+y),o(m-t.by/2-1),a(t.tx2width),o(t.by+2)))}))}function B(t,e){var r=t.index,n=t.trace||{},a=t.cd[0],s=t.cd[r]||{};function l(t){return t||i(t)&&0===t}var u=Array.isArray(r)?function(t,e){var i=o.castOption(a,r,t);return l(i)?i:o.extractOption({},n,"",e)}:function(t,e){return o.extractOption(s,n,t,e)};function c(e,r,n){var i=u(r,n);l(i)&&(t[e]=i)}if(c("hoverinfo","hi","hoverinfo"),c("bgcolor","hbg","hoverlabel.bgcolor"),c("borderColor","hbc","hoverlabel.bordercolor"),c("fontFamily","htf","hoverlabel.font.family"),c("fontSize","hts","hoverlabel.font.size"),c("fontColor","htc","hoverlabel.font.color"),c("fontWeight","htw","hoverlabel.font.weight"),c("fontStyle","hty","hoverlabel.font.style"),c("fontVariant","htv","hoverlabel.font.variant"),c("nameLength","hnl","hoverlabel.namelength"),c("textAlign","hta","hoverlabel.align"),t.posref="y"===e||"closest"===e&&"h"===n.orientation?t.xa._offset+(t.x0+t.x1)/2:t.ya._offset+(t.y0+t.y1)/2,t.x0=o.constrain(t.x0,0,t.xa._length),t.x1=o.constrain(t.x1,0,t.xa._length),t.y0=o.constrain(t.y0,0,t.ya._length),t.y1=o.constrain(t.y1,0,t.ya._length),void 0!==t.xLabelVal&&(t.xLabel="xLabel"in t?t.xLabel:g.hoverLabelText(t.xa,t.xLabelVal,n.xhoverformat),t.xVal=t.xa.c2d(t.xLabelVal)),void 0!==t.yLabelVal&&(t.yLabel="yLabel"in t?t.yLabel:g.hoverLabelText(t.ya,t.yLabelVal,n.yhoverformat),t.yVal=t.ya.c2d(t.yLabelVal)),void 0!==t.zLabelVal&&void 0===t.zLabel&&(t.zLabel=String(t.zLabelVal)),!(isNaN(t.xerr)||"log"===t.xa.type&&t.xerr<=0)){var f=g.tickText(t.xa,t.xa.c2l(t.xerr),"hover").text;void 0!==t.xerrneg?t.xLabel+=" +"+f+" / -"+g.tickText(t.xa,t.xa.c2l(t.xerrneg),"hover").text:t.xLabel+=" ± "+f,"x"===e&&(t.distance+=1)}if(!(isNaN(t.yerr)||"log"===t.ya.type&&t.yerr<=0)){var h=g.tickText(t.ya,t.ya.c2l(t.yerr),"hover").text;void 0!==t.yerrneg?t.yLabel+=" +"+h+" / -"+g.tickText(t.ya,t.ya.c2l(t.yerrneg),"hover").text:t.yLabel+=" ± "+h,"y"===e&&(t.distance+=1)}var p=t.hoverinfo||t.trace.hoverinfo;return p&&"all"!==p&&(-1===(p=Array.isArray(p)?p:p.split("+")).indexOf("x")&&(t.xLabel=void 0),-1===p.indexOf("y")&&(t.yLabel=void 0),-1===p.indexOf("z")&&(t.zLabel=void 0),-1===p.indexOf("text")&&(t.text=void 0),-1===p.indexOf("name")&&(t.name=void 0)),t}function N(t,e,r){var n,i,o=r.container,s=r.fullLayout,l=s._size,u=r.event,c=!!e.hLinePoint,f=!!e.vLinePoint;if(o.selectAll(".spikeline").remove(),f||c){var h=d.combine(s.plot_bgcolor,s.paper_bgcolor);if(c){var v,y,m=e.hLinePoint;n=m&&m.xa,"cursor"===(i=m&&m.ya).spikesnap?(v=u.pointerX,y=u.pointerY):(v=n._offset+m.x,y=i._offset+m.y);var x,b,_=a.readability(m.color,h)<1.5?d.contrast(h):m.color,w=i.spikemode,T=i.spikethickness,k=i.spikecolor||_,A=g.getPxPosition(t,i);if(-1!==w.indexOf("toaxis")||-1!==w.indexOf("across")){if(-1!==w.indexOf("toaxis")&&(x=A,b=v),-1!==w.indexOf("across")){var M=i._counterDomainMin,S=i._counterDomainMax;"free"===i.anchor&&(M=Math.min(M,i.position),S=Math.max(S,i.position)),x=l.l+M*l.w,b=l.l+S*l.w}o.insert("line",":first-child").attr({x1:x,x2:b,y1:y,y2:y,"stroke-width":T,stroke:k,"stroke-dasharray":p.dashStyle(i.spikedash,T)}).classed("spikeline",!0).classed("crisp",!0),o.insert("line",":first-child").attr({x1:x,x2:b,y1:y,y2:y,"stroke-width":T+2,stroke:h}).classed("spikeline",!0).classed("crisp",!0)}-1!==w.indexOf("marker")&&o.insert("circle",":first-child").attr({cx:A+("right"!==i.side?T:-T),cy:y,r:T,fill:k}).classed("spikeline",!0)}if(f){var E,L,C=e.vLinePoint;n=C&&C.xa,i=C&&C.ya,"cursor"===n.spikesnap?(E=u.pointerX,L=u.pointerY):(E=n._offset+C.x,L=i._offset+C.y);var P,O,I=a.readability(C.color,h)<1.5?d.contrast(h):C.color,z=n.spikemode,D=n.spikethickness,R=n.spikecolor||I,F=g.getPxPosition(t,n);if(-1!==z.indexOf("toaxis")||-1!==z.indexOf("across")){if(-1!==z.indexOf("toaxis")&&(P=F,O=L),-1!==z.indexOf("across")){var B=n._counterDomainMin,N=n._counterDomainMax;"free"===n.anchor&&(B=Math.min(B,n.position),N=Math.max(N,n.position)),P=l.t+(1-N)*l.h,O=l.t+(1-B)*l.h}o.insert("line",":first-child").attr({x1:E,x2:E,y1:P,y2:O,"stroke-width":D,stroke:R,"stroke-dasharray":p.dashStyle(n.spikedash,D)}).classed("spikeline",!0).classed("crisp",!0),o.insert("line",":first-child").attr({x1:E,x2:E,y1:P,y2:O,"stroke-width":D+2,stroke:h}).classed("spikeline",!0).classed("crisp",!0)}-1!==z.indexOf("marker")&&o.insert("circle",":first-child").attr({cx:E,cy:F-("top"!==n.side?D:-D),r:D,fill:R}).classed("spikeline",!0)}}}function j(t,e){return!e||e.vLinePoint!==t._spikepoints.vLinePoint||e.hLinePoint!==t._spikepoints.hLinePoint}function U(t,e){return f.plainText(t||"",{len:e,allowedTags:["br","sub","sup","b","i","em"]})}function V(t,e,r){var n=e[t+"a"],i=e[t+"Val"],a=e.cd[0];if("category"===n.type||"multicategory"===n.type)i=n._categoriesMap[i];else if("date"===n.type){var o=e.trace[t+"periodalignment"];if(o){var s=e.cd[e.index],l=s[t+"Start"];void 0===l&&(l=s[t]);var u=s[t+"End"];void 0===u&&(u=s[t]);var c=u-l;"end"===o?i+=c:"middle"===o&&(i+=c/2)}i=n.d2c(i)}return a&&a.t&&a.t.posLetter===n._id&&("group"!==r.boxmode&&"group"!==r.violinmode||(i+=a.t.dPos)),i}function q(t){return t.offsetTop+t.clientTop}function H(t){return t.offsetLeft+t.clientLeft}function G(t,e){var r=t._fullLayout,n=e.getBoundingClientRect(),i=n.left,a=n.top,s=i+n.width,l=a+n.height,u=o.apply3DTransform(r._invTransform)(i,a),c=o.apply3DTransform(r._invTransform)(s,l),f=u[0],h=u[1],p=c[0],d=c[1];return{x:f,y:h,width:p-f,height:d-h,top:Math.min(h,d),left:Math.min(f,p),right:Math.max(f,p),bottom:Math.max(h,d)}}},16132:function(t,e,r){"use strict";var n=r(3400),i=r(76308),a=r(10624).isUnifiedHover;t.exports=function(t,e,r,o){o=o||{};var s=e.legend;function l(t){o.font[t]||(o.font[t]=s?e.legend.font[t]:e.font[t])}e&&a(e.hovermode)&&(o.font||(o.font={}),l("size"),l("family"),l("color"),l("weight"),l("style"),l("variant"),s?(o.bgcolor||(o.bgcolor=i.combine(e.legend.bgcolor,e.paper_bgcolor)),o.bordercolor||(o.bordercolor=e.legend.bordercolor)):o.bgcolor||(o.bgcolor=e.paper_bgcolor)),r("hoverlabel.bgcolor",o.bgcolor),r("hoverlabel.bordercolor",o.bordercolor),r("hoverlabel.namelength",o.namelength),n.coerceFont(r,"hoverlabel.font",o.font),r("hoverlabel.align",o.align)}},41008:function(t,e,r){"use strict";var n=r(3400),i=r(65460);t.exports=function(t,e){function r(r,a){return void 0!==e[r]?e[r]:n.coerce(t,e,i,r,a)}return r("clickmode"),r("hoversubplots"),r("hovermode")}},93024:function(t,e,r){"use strict";var n=r(33428),i=r(3400),a=r(86476),o=r(10624),s=r(65460),l=r(83292);t.exports={moduleType:"component",name:"fx",constants:r(92456),schema:{layout:s},attributes:r(55756),layoutAttributes:s,supplyLayoutGlobalDefaults:r(81976),supplyDefaults:r(95448),supplyLayoutDefaults:r(88336),calc:r(55056),getDistanceFunction:o.getDistanceFunction,getClosest:o.getClosest,inbox:o.inbox,quadrature:o.quadrature,appendArrayPointValue:o.appendArrayPointValue,castHoverOption:function(t,e,r){return i.castOption(t,e,"hoverlabel."+r)},castHoverinfo:function(t,e,r){return i.castOption(t,r,"hoverinfo",(function(r){return i.coerceHoverinfo({hoverinfo:r},{_module:t._module},e)}))},hover:l.hover,unhover:a.unhover,loneHover:l.loneHover,loneUnhover:function(t){var e=i.isD3Selection(t)?t:n.select(t);e.selectAll("g.hovertext").remove(),e.selectAll(".spikeline").remove()},click:r(62376)}},65460:function(t,e,r){"use strict";var n=r(92456),i=r(25376),a=i({editType:"none"});a.family.dflt=n.HOVERFONT,a.size.dflt=n.HOVERFONTSIZE,t.exports={clickmode:{valType:"flaglist",flags:["event","select"],dflt:"event",editType:"plot",extras:["none"]},dragmode:{valType:"enumerated",values:["zoom","pan","select","lasso","drawclosedpath","drawopenpath","drawline","drawrect","drawcircle","orbit","turntable",!1],dflt:"zoom",editType:"modebar"},hovermode:{valType:"enumerated",values:["x","y","closest",!1,"x unified","y unified"],dflt:"closest",editType:"modebar"},hoversubplots:{valType:"enumerated",values:["single","overlaying","axis"],dflt:"overlaying",editType:"none"},hoverdistance:{valType:"integer",min:-1,dflt:20,editType:"none"},spikedistance:{valType:"integer",min:-1,dflt:-1,editType:"none"},hoverlabel:{bgcolor:{valType:"color",editType:"none"},bordercolor:{valType:"color",editType:"none"},font:a,grouptitlefont:i({editType:"none"}),align:{valType:"enumerated",values:["left","right","auto"],dflt:"auto",editType:"none"},namelength:{valType:"integer",min:-1,dflt:15,editType:"none"},editType:"none"},selectdirection:{valType:"enumerated",values:["h","v","d","any"],dflt:"any",editType:"none"}}},88336:function(t,e,r){"use strict";var n=r(3400),i=r(65460),a=r(41008),o=r(16132);t.exports=function(t,e){function r(r,a){return n.coerce(t,e,i,r,a)}a(t,e)&&(r("hoverdistance"),r("spikedistance")),"select"===r("dragmode")&&r("selectdirection");var s=e._has("mapbox"),l=e._has("geo"),u=e._basePlotModules.length;"zoom"===e.dragmode&&((s||l)&&1===u||s&&l&&2===u)&&(e.dragmode="pan"),o(t,e,r),n.coerceFont(r,"hoverlabel.grouptitlefont",e.hoverlabel.font)}},81976:function(t,e,r){"use strict";var n=r(3400),i=r(16132),a=r(65460);t.exports=function(t,e){i(t,e,(function(r,i){return n.coerce(t,e,a,r,i)}))}},12704:function(t,e,r){"use strict";var n=r(3400),i=r(53756).counter,a=r(86968).u,o=r(33816).idRegex,s=r(31780),l={rows:{valType:"integer",min:1,editType:"plot"},roworder:{valType:"enumerated",values:["top to bottom","bottom to top"],dflt:"top to bottom",editType:"plot"},columns:{valType:"integer",min:1,editType:"plot"},subplots:{valType:"info_array",freeLength:!0,dimensions:2,items:{valType:"enumerated",values:[i("xy").toString(),""],editType:"plot"},editType:"plot"},xaxes:{valType:"info_array",freeLength:!0,items:{valType:"enumerated",values:[o.x.toString(),""],editType:"plot"},editType:"plot"},yaxes:{valType:"info_array",freeLength:!0,items:{valType:"enumerated",values:[o.y.toString(),""],editType:"plot"},editType:"plot"},pattern:{valType:"enumerated",values:["independent","coupled"],dflt:"coupled",editType:"plot"},xgap:{valType:"number",min:0,max:1,editType:"plot"},ygap:{valType:"number",min:0,max:1,editType:"plot"},domain:a({name:"grid",editType:"plot",noGridCell:!0},{}),xside:{valType:"enumerated",values:["bottom","bottom plot","top plot","top"],dflt:"bottom plot",editType:"plot"},yside:{valType:"enumerated",values:["left","left plot","right plot","right"],dflt:"left plot",editType:"plot"},editType:"plot"};function u(t,e,r){var n=e[r+"axes"],i=Object.keys((t._splomAxes||{})[r]||{});return Array.isArray(n)?n:i.length?i:void 0}function c(t,e,r,n,i,a){var o=e(t+"gap",r),s=e("domain."+t);e(t+"side",n);for(var l=new Array(i),u=s[0],c=(s[1]-u)/(i-o),f=c*(1-o),h=0;h<i;h++){var p=u+c*h;l[a?i-1-h:h]=[p,p+f]}return l}function f(t,e,r,n,i){var a,o=new Array(r);function s(t,r){-1!==e.indexOf(r)&&void 0===n[r]?(o[t]=r,n[r]=t):o[t]=""}if(Array.isArray(t))for(a=0;a<r;a++)s(a,t[a]);else for(s(0,i),a=1;a<r;a++)s(a,i+(a+1));return o}t.exports={moduleType:"component",name:"grid",schema:{layout:{grid:l}},layoutAttributes:l,sizeDefaults:function(t,e){var r=t.grid||{},i=u(e,r,"x"),a=u(e,r,"y");if(t.grid||i||a){var o,f,h=Array.isArray(r.subplots)&&Array.isArray(r.subplots[0]),p=Array.isArray(i),d=Array.isArray(a),v=p&&i!==r.xaxes&&d&&a!==r.yaxes;h?(o=r.subplots.length,f=r.subplots[0].length):(d&&(o=a.length),p&&(f=i.length));var g=s.newContainer(e,"grid"),y=k("rows",o),m=k("columns",f);if(y*m>1){h||p||d||"independent"===k("pattern")&&(h=!0),g._hasSubplotGrid=h;var x,b,_="top to bottom"===k("roworder"),w=h?.2:.1,T=h?.3:.1;v&&e._splomGridDflt&&(x=e._splomGridDflt.xside,b=e._splomGridDflt.yside),g._domains={x:c("x",k,w,x,m),y:c("y",k,T,b,y,_)}}else delete e.grid}function k(t,e){return n.coerce(r,g,l,t,e)}},contentDefaults:function(t,e){var r=e.grid;if(r&&r._domains){var n,i,a,o,s,l,c,h=t.grid||{},p=e._subplots,d=r._hasSubplotGrid,v=r.rows,g=r.columns,y="independent"===r.pattern,m=r._axisMap={};if(d){var x=h.subplots||[];l=r.subplots=new Array(v);var b=1;for(n=0;n<v;n++){var _=l[n]=new Array(g),w=x[n]||[];for(i=0;i<g;i++)if(y?(s=1===b?"xy":"x"+b+"y"+b,b++):s=w[i],_[i]="",-1!==p.cartesian.indexOf(s)){if(c=s.indexOf("y"),a=s.slice(0,c),o=s.slice(c),void 0!==m[a]&&m[a]!==i||void 0!==m[o]&&m[o]!==n)continue;_[i]=s,m[a]=i,m[o]=n}}}else{var T=u(e,h,"x"),k=u(e,h,"y");r.xaxes=f(T,p.xaxis,g,m,"x"),r.yaxes=f(k,p.yaxis,v,m,"y")}var A=r._anchors={},M="top to bottom"===r.roworder;for(var S in m){var E,L,C,P=S.charAt(0),O=r[P+"side"];if(O.length<8)A[S]="free";else if("x"===P){if("t"===O.charAt(0)===M?(E=0,L=1,C=v):(E=v-1,L=-1,C=-1),d){var I=m[S];for(n=E;n!==C;n+=L)if((s=l[n][I])&&(c=s.indexOf("y"),s.slice(0,c)===S)){A[S]=s.slice(c);break}}else for(n=E;n!==C;n+=L)if(o=r.yaxes[n],-1!==p.cartesian.indexOf(S+o)){A[S]=o;break}}else if("l"===O.charAt(0)?(E=0,L=1,C=g):(E=g-1,L=-1,C=-1),d){var z=m[S];for(n=E;n!==C;n+=L)if((s=l[z][n])&&(c=s.indexOf("y"),s.slice(c)===S)){A[S]=s.slice(0,c);break}}else for(n=E;n!==C;n+=L)if(a=r.xaxes[n],-1!==p.cartesian.indexOf(a+S)){A[S]=a;break}}}}}},65760:function(t,e,r){"use strict";var n=r(33816),i=r(31780).templatedArray;r(36208),t.exports=i("image",{visible:{valType:"boolean",dflt:!0,editType:"arraydraw"},source:{valType:"string",editType:"arraydraw"},layer:{valType:"enumerated",values:["below","above"],dflt:"above",editType:"arraydraw"},sizex:{valType:"number",dflt:0,editType:"arraydraw"},sizey:{valType:"number",dflt:0,editType:"arraydraw"},sizing:{valType:"enumerated",values:["fill","contain","stretch"],dflt:"contain",editType:"arraydraw"},opacity:{valType:"number",min:0,max:1,dflt:1,editType:"arraydraw"},x:{valType:"any",dflt:0,editType:"arraydraw"},y:{valType:"any",dflt:0,editType:"arraydraw"},xanchor:{valType:"enumerated",values:["left","center","right"],dflt:"left",editType:"arraydraw"},yanchor:{valType:"enumerated",values:["top","middle","bottom"],dflt:"top",editType:"arraydraw"},xref:{valType:"enumerated",values:["paper",n.idRegex.x.toString()],dflt:"paper",editType:"arraydraw"},yref:{valType:"enumerated",values:["paper",n.idRegex.y.toString()],dflt:"paper",editType:"arraydraw"},editType:"arraydraw"})},63556:function(t,e,r){"use strict";var n=r(38248),i=r(36896);t.exports=function(t,e,r,a){e=e||{};var o="log"===r&&"linear"===e.type,s="linear"===r&&"log"===e.type;if(o||s)for(var l,u,c=t._fullLayout.images,f=e._id.charAt(0),h=0;h<c.length;h++)if(u="images["+h+"].",(l=c[h])[f+"ref"]===e._id){var p=l[f],d=l["size"+f],v=null,g=null;if(o){v=i(p,e.range);var y=d/Math.pow(10,v)/2;g=2*Math.log(y+Math.sqrt(1+y*y))/Math.LN10}else g=(v=Math.pow(10,p))*(Math.pow(10,d/2)-Math.pow(10,-d/2));n(v)?n(g)||(g=null):(v=null,g=null),a(u+f,v),a(u+"size"+f,g)}}},25024:function(t,e,r){"use strict";var n=r(3400),i=r(54460),a=r(51272),o=r(65760);function s(t,e,r){function a(r,i){return n.coerce(t,e,o,r,i)}var s=a("source");if(!a("visible",!!s))return e;a("layer"),a("xanchor"),a("yanchor"),a("sizex"),a("sizey"),a("sizing"),a("opacity");for(var l={_fullLayout:r},u=["x","y"],c=0;c<2;c++){var f=u[c],h=i.coerceRef(t,e,l,f,"paper",void 0);"paper"!==h&&i.getFromId(l,h)._imgIndices.push(e._index),i.coercePosition(e,l,a,h,f,0)}return e}t.exports=function(t,e){a(t,e,{name:"images",handleItemDefaults:s})}},60963:function(t,e,r){"use strict";var n=r(33428),i=r(43616),a=r(54460),o=r(79811),s=r(9616);t.exports=function(t){var e,r,l=t._fullLayout,u=[],c={},f=[];for(r=0;r<l.images.length;r++){var h=l.images[r];if(h.visible)if("below"===h.layer&&"paper"!==h.xref&&"paper"!==h.yref){e=o.ref2id(h.xref)+o.ref2id(h.yref);var p=l._plots[e];if(!p){f.push(h);continue}p.mainplot&&(e=p.mainplot.id),c[e]||(c[e]=[]),c[e].push(h)}else"above"===h.layer?u.push(h):f.push(h)}var d={left:{sizing:"xMin",offset:0},center:{sizing:"xMid",offset:-.5},right:{sizing:"xMax",offset:-1}},v={top:{sizing:"YMin",offset:0},middle:{sizing:"YMid",offset:-.5},bottom:{sizing:"YMax",offset:-1}};function g(e){var r=n.select(this);if(this._imgSrc!==e.source)if(r.attr("xmlns",s.svg),e.source&&"data:"===e.source.slice(0,5))r.attr("xlink:href",e.source),this._imgSrc=e.source;else{var i=new Promise(function(t){var n=new Image;function i(){r.remove(),t()}this.img=n,n.setAttribute("crossOrigin","anonymous"),n.onerror=i,n.onload=function(){var e=document.createElement("canvas");e.width=this.width,e.height=this.height,e.getContext("2d",{willReadFrequently:!0}).drawImage(this,0,0);var n=e.toDataURL("image/png");r.attr("xlink:href",n),t()},r.on("error",i),n.src=e.source,this._imgSrc=e.source}.bind(this));t._promises.push(i)}}function y(e){var r,o,s=n.select(this),u=a.getFromId(t,e.xref),c=a.getFromId(t,e.yref),f="domain"===a.getRefType(e.xref),h="domain"===a.getRefType(e.yref),p=l._size;r=void 0!==u?"string"==typeof e.xref&&f?u._length*e.sizex:Math.abs(u.l2p(e.sizex)-u.l2p(0)):e.sizex*p.w,o=void 0!==c?"string"==typeof e.yref&&h?c._length*e.sizey:Math.abs(c.l2p(e.sizey)-c.l2p(0)):e.sizey*p.h;var g,y,m=r*d[e.xanchor].offset,x=o*v[e.yanchor].offset,b=d[e.xanchor].sizing+v[e.yanchor].sizing;switch(g=void 0!==u?"string"==typeof e.xref&&f?u._length*e.x+u._offset:u.r2p(e.x)+u._offset:e.x*p.w+p.l,g+=m,y=void 0!==c?"string"==typeof e.yref&&h?c._length*(1-e.y)+c._offset:c.r2p(e.y)+c._offset:p.h-e.y*p.h+p.t,y+=x,e.sizing){case"fill":b+=" slice";break;case"stretch":b="none"}s.attr({x:g,y:y,width:r,height:o,preserveAspectRatio:b,opacity:e.opacity});var _=(u&&"domain"!==a.getRefType(e.xref)?u._id:"")+(c&&"domain"!==a.getRefType(e.yref)?c._id:"");i.setClipUrl(s,_?"clip"+l._uid+_:null,t)}var m=l._imageLowerLayer.selectAll("image").data(f),x=l._imageUpperLayer.selectAll("image").data(u);m.enter().append("image"),x.enter().append("image"),m.exit().remove(),x.exit().remove(),m.each((function(t){g.bind(this)(t),y.bind(this)(t)})),x.each((function(t){g.bind(this)(t),y.bind(this)(t)}));var b=Object.keys(l._plots);for(r=0;r<b.length;r++){e=b[r];var _=l._plots[e];if(_.imagelayer){var w=_.imagelayer.selectAll("image").data(c[e]||[]);w.enter().append("image"),w.exit().remove(),w.each((function(t){g.bind(this)(t),y.bind(this)(t)}))}}}},7402:function(t,e,r){"use strict";t.exports={moduleType:"component",name:"images",layoutAttributes:r(65760),supplyLayoutDefaults:r(25024),includeBasePlot:r(36632)("images"),draw:r(60963),convertCoords:r(63556)}},3800:function(t,e,r){"use strict";var n=r(25376),i=r(22548);t.exports={_isSubplotObj:!0,visible:{valType:"boolean",dflt:!0,editType:"legend"},bgcolor:{valType:"color",editType:"legend"},bordercolor:{valType:"color",dflt:i.defaultLine,editType:"legend"},borderwidth:{valType:"number",min:0,dflt:0,editType:"legend"},font:n({editType:"legend"}),grouptitlefont:n({editType:"legend"}),orientation:{valType:"enumerated",values:["v","h"],dflt:"v",editType:"legend"},traceorder:{valType:"flaglist",flags:["reversed","grouped"],extras:["normal"],editType:"legend"},tracegroupgap:{valType:"number",min:0,dflt:10,editType:"legend"},entrywidth:{valType:"number",min:0,editType:"legend"},entrywidthmode:{valType:"enumerated",values:["fraction","pixels"],dflt:"pixels",editType:"legend"},indentation:{valType:"number",min:-15,dflt:0,editType:"legend"},itemsizing:{valType:"enumerated",values:["trace","constant"],dflt:"trace",editType:"legend"},itemwidth:{valType:"number",min:30,dflt:30,editType:"legend"},itemclick:{valType:"enumerated",values:["toggle","toggleothers",!1],dflt:"toggle",editType:"legend"},itemdoubleclick:{valType:"enumerated",values:["toggle","toggleothers",!1],dflt:"toggleothers",editType:"legend"},groupclick:{valType:"enumerated",values:["toggleitem","togglegroup"],dflt:"togglegroup",editType:"legend"},x:{valType:"number",editType:"legend"},xref:{valType:"enumerated",dflt:"paper",values:["container","paper"],editType:"layoutstyle"},xanchor:{valType:"enumerated",values:["auto","left","center","right"],dflt:"left",editType:"legend"},y:{valType:"number",editType:"legend"},yref:{valType:"enumerated",dflt:"paper",values:["container","paper"],editType:"layoutstyle"},yanchor:{valType:"enumerated",values:["auto","top","middle","bottom"],editType:"legend"},uirevision:{valType:"any",editType:"none"},valign:{valType:"enumerated",values:["top","middle","bottom"],dflt:"middle",editType:"legend"},title:{text:{valType:"string",dflt:"",editType:"legend"},font:n({editType:"legend"}),side:{valType:"enumerated",values:["top","left","top left","top center","top right"],editType:"legend"},editType:"legend"},editType:"legend"}},65196:function(t){"use strict";t.exports={scrollBarWidth:6,scrollBarMinHeight:20,scrollBarColor:"#808BA4",scrollBarMargin:4,scrollBarEnterAttrs:{rx:20,ry:3,width:0,height:0},titlePad:2,itemGap:5}},77864:function(t,e,r){"use strict";var n=r(24040),i=r(3400),a=r(31780),o=r(45464),s=r(3800),l=r(64859),u=r(42451);function c(t,e,r,c){var f=e[t]||{},h=a.newContainer(r,t);function p(t,e){return i.coerce(f,h,s,t,e)}var d=i.coerceFont(p,"font",r.font);if(p("bgcolor",r.paper_bgcolor),p("bordercolor"),p("visible")){for(var v,g=function(t,e){var r=v._input,n=v;return i.coerce(r,n,o,t,e)},y=r.font||{},m=i.coerceFont(p,"grouptitlefont",i.extendFlat({},y,{size:Math.round(1.1*y.size)})),x=0,b=!1,_="normal",w=(r.shapes||[]).filter((function(t){return t.showlegend})),T=c.concat(w).filter((function(e){return t===(e.legend||"legend")})),k=0;k<T.length;k++)if((v=T[k]).visible){var A=v._isShape;(v.showlegend||v._dfltShowLegend&&!(v._module&&v._module.attributes&&v._module.attributes.showlegend&&!1===v._module.attributes.showlegend.dflt))&&(x++,v.showlegend&&(b=!0,(!A&&n.traceIs(v,"pie-like")||!0===v._input.showlegend)&&x++),i.coerceFont(g,"legendgrouptitle.font",m)),(!A&&n.traceIs(v,"bar")&&"stack"===r.barmode||-1!==["tonextx","tonexty"].indexOf(v.fill))&&(_=u.isGrouped({traceorder:_})?"grouped+reversed":"reversed"),void 0!==v.legendgroup&&""!==v.legendgroup&&(_=u.isReversed({traceorder:_})?"reversed+grouped":"grouped")}var M=i.coerce(e,r,l,"showlegend",b&&x>("legend"===t?1:0));if(!1===M&&(r[t]=void 0),(!1!==M||f.uirevision)&&(p("uirevision",r.uirevision),!1!==M)){p("borderwidth");var S,E,L,C="h"===p("orientation"),P="paper"===p("yref"),O="paper"===p("xref"),I="left";if(C?(S=0,n.getComponentMethod("rangeslider","isVisible")(e.xaxis)?P?(E=1.1,L="bottom"):(E=1,L="top"):P?(E=-.1,L="top"):(E=0,L="bottom")):(E=1,L="auto",O?S=1.02:(S=1,I="right")),i.coerce(f,h,{x:{valType:"number",editType:"legend",min:O?-2:0,max:O?3:1,dflt:S}},"x"),i.coerce(f,h,{y:{valType:"number",editType:"legend",min:P?-2:0,max:P?3:1,dflt:E}},"y"),p("traceorder",_),u.isGrouped(r[t])&&p("tracegroupgap"),p("entrywidth"),p("entrywidthmode"),p("indentation"),p("itemsizing"),p("itemwidth"),p("itemclick"),p("itemdoubleclick"),p("groupclick"),p("xanchor",I),p("yanchor",L),p("valign"),i.noneOrAll(f,h,["x","y"]),p("title.text")){p("title.side",C?"left":"top");var z=i.extendFlat({},d,{size:i.bigFont(d.size)});i.coerceFont(p,"title.font",z)}}}}t.exports=function(t,e,r){var n,a=r.slice(),o=e.shapes;if(o)for(n=0;n<o.length;n++){var s=o[n];if(s.showlegend){var l={_input:s._input,visible:s.visible,showlegend:s.showlegend,legend:s.legend};a.push(l)}}var u=["legend"];for(n=0;n<a.length;n++)i.pushUnique(u,a[n].legend);for(e._legends=[],n=0;n<u.length;n++){var f=u[n];c(f,t,e,a),e[f]&&e[f].visible&&(e[f]._id=f),e._legends.push(f)}}},31140:function(t,e,r){"use strict";var n=r(33428),i=r(3400),a=r(7316),o=r(24040),s=r(95924),l=r(86476),u=r(43616),c=r(76308),f=r(72736),h=r(33048),p=r(65196),d=r(84284),v=d.LINE_SPACING,g=d.FROM_TL,y=d.FROM_BR,m=r(35456),x=r(2012),b=r(42451),_=1,w=/^legend[0-9]*$/;function T(t,e){var r,s,h=e||{},d=t._fullLayout,w=O(h),T=h._inHover;if(T?(s=h.layer,r="hover"):(s=d._infolayer,r=w),s){var S;if(r+=d._uid,t._legendMouseDownTime||(t._legendMouseDownTime=0),T){if(!h.entries)return;S=m(h.entries,h)}else{for(var I=(t.calcdata||[]).slice(),z=d.shapes,D=0;D<z.length;D++){var R=z[D];if(R.showlegend){var F={_isShape:!0,_fullInput:R,index:R._index,name:R.name||R.label.text||"shape "+R._index,legend:R.legend,legendgroup:R.legendgroup,legendgrouptitle:R.legendgrouptitle,legendrank:R.legendrank,legendwidth:R.legendwidth,showlegend:R.showlegend,visible:R.visible,opacity:R.opacity,mode:"line"===R.type?"lines":"markers",line:R.line,marker:{line:R.line,color:R.fillcolor,size:12,symbol:"rect"===R.type?"square":"circle"===R.type?"circle":"hexagon2"}};I.push([{trace:F}])}}S=d.showlegend&&m(I,h,d._legends.length>1)}var B=d.hiddenlabels||[];if(!(T||d.showlegend&&S.length))return s.selectAll("."+w).remove(),d._topdefs.select("#"+r).remove(),a.autoMargin(t,w);var N=i.ensureSingle(s,"g",w,(function(t){T||t.attr("pointer-events","all")})),j=i.ensureSingleById(d._topdefs,"clipPath",r,(function(t){t.append("rect")})),U=i.ensureSingle(N,"rect","bg",(function(t){t.attr("shape-rendering","crispEdges")}));U.call(c.stroke,h.bordercolor).call(c.fill,h.bgcolor).style("stroke-width",h.borderwidth+"px");var V,q=i.ensureSingle(N,"g","scrollbox"),H=h.title;h._titleWidth=0,h._titleHeight=0,H.text?((V=i.ensureSingle(q,"text",w+"titletext")).attr("text-anchor","start").call(u.font,H.font).text(H.text),L(V,q,t,h,_)):q.selectAll("."+w+"titletext").remove();var G=i.ensureSingle(N,"rect","scrollbar",(function(t){t.attr(p.scrollBarEnterAttrs).call(c.fill,p.scrollBarColor)})),W=q.selectAll("g.groups").data(S);W.enter().append("g").attr("class","groups"),W.exit().remove();var Y=W.selectAll("g.traces").data(i.identity);Y.enter().append("g").attr("class","traces"),Y.exit().remove(),Y.style("opacity",(function(t){var e=t[0].trace;return o.traceIs(e,"pie-like")?-1!==B.indexOf(t[0].label)?.5:1:"legendonly"===e.visible?.5:1})).each((function(){n.select(this).call(M,t,h)})).call(x,t,h).each((function(){T||n.select(this).call(E,t,w)})),i.syncOrAsync([a.previousPromises,function(){return function(t,e,r,i){var a=t._fullLayout,o=O(i);i||(i=a[o]);var s=a._size,l=b.isVertical(i),c=b.isGrouped(i),f="fraction"===i.entrywidthmode,h=i.borderwidth,d=2*h,v=p.itemGap,g=i.indentation+i.itemwidth+2*v,y=2*(h+v),m=P(i),x=i.y<0||0===i.y&&"top"===m,_=i.y>1||1===i.y&&"bottom"===m,w=i.tracegroupgap,T={};i._maxHeight=Math.max(x||_?a.height/2:s.h,30);var A=0;i._width=0,i._height=0;var M=function(t){var e=0,r=0,n=t.title.side;return n&&(-1!==n.indexOf("left")&&(e=t._titleWidth),-1!==n.indexOf("top")&&(r=t._titleHeight)),[e,r]}(i);if(l)r.each((function(t){var e=t[0].height;u.setTranslate(this,h+M[0],h+M[1]+i._height+e/2+v),i._height+=e,i._width=Math.max(i._width,t[0].width)})),A=g+i._width,i._width+=v+g+d,i._height+=y,c&&(e.each((function(t,e){u.setTranslate(this,0,e*i.tracegroupgap)})),i._height+=(i._lgroupsLength-1)*i.tracegroupgap);else{var S=C(i),E=i.x<0||0===i.x&&"right"===S,L=i.x>1||1===i.x&&"left"===S,I=_||x,z=a.width/2;i._maxWidth=Math.max(E?I&&"left"===S?s.l+s.w:z:L?I&&"right"===S?s.r+s.w:z:s.w,2*g);var D=0,R=0;r.each((function(t){var e=k(t,i,g);D=Math.max(D,e),R+=e})),A=null;var F=0;if(c){var B=0,N=0,j=0;e.each((function(){var t=0,e=0;n.select(this).selectAll("g.traces").each((function(r){var n=k(r,i,g),a=r[0].height;u.setTranslate(this,M[0],M[1]+h+v+a/2+e),e+=a,t=Math.max(t,n),T[r[0].trace.legendgroup]=t}));var r=t+v;N>0&&r+h+N>i._maxWidth?(F=Math.max(F,N),N=0,j+=B+w,B=e):B=Math.max(B,e),u.setTranslate(this,N,j),N+=r})),i._width=Math.max(F,N)+h,i._height=j+B+y}else{var U=r.size(),V=R+d+(U-1)*v<i._maxWidth,q=0,H=0,G=0,W=0;r.each((function(t){var e=t[0].height,r=k(t,i,g),n=V?r:D;f||(n+=v),n+h+H-v>=i._maxWidth&&(F=Math.max(F,W),H=0,G+=q,i._height+=q,q=0),u.setTranslate(this,M[0]+h+H,M[1]+h+G+e/2+v),W=H+r+v,H+=n,q=Math.max(q,e)})),V?(i._width=H+d,i._height=q+y):(i._width=Math.max(F,W)+d,i._height+=q+y)}}i._width=Math.ceil(Math.max(i._width+M[0],i._titleWidth+2*(h+p.titlePad))),i._height=Math.ceil(Math.max(i._height+M[1],i._titleHeight+2*(h+p.itemGap))),i._effHeight=Math.min(i._height,i._maxHeight);var Y=t._context.edits,X=Y.legendText||Y.legendPosition;r.each((function(t){var e=n.select(this).select("."+o+"toggle"),r=t[0].height,a=t[0].trace.legendgroup,s=k(t,i,g);c&&""!==a&&(s=T[a]);var h=X?g:A||s;l||f||(h+=v/2),u.setRect(e,0,-r/2,h,r)}))}(t,W,Y,h)},function(){var e,c,m,x,b=d._size,_=h.borderwidth,k="paper"===h.xref,M="paper"===h.yref;if(H.text&&function(t,e,r){if("top center"===e.title.side||"top right"===e.title.side){var n=e.title.font.size*v,i=0,a=t.node(),o=u.bBox(a).width;"top center"===e.title.side?i=.5*(e._width-2*r-2*p.titlePad-o):"top right"===e.title.side&&(i=e._width-2*r-2*p.titlePad-o),f.positionText(t,r+p.titlePad+i,r+n)}}(V,h,_),!T){var S,E;S=k?b.l+b.w*h.x-g[C(h)]*h._width:d.width*h.x-g[C(h)]*h._width,E=M?b.t+b.h*(1-h.y)-g[P(h)]*h._effHeight:d.height*(1-h.y)-g[P(h)]*h._effHeight;var L=function(t,e,r,n){var i=t._fullLayout,o=i[e],s=C(o),l=P(o),u="paper"===o.xref,c="paper"===o.yref;t._fullLayout._reservedMargin[e]={};var f=o.y<.5?"b":"t",h=o.x<.5?"l":"r",p={r:i.width-r,l:r+o._width,b:i.height-n,t:n+o._effHeight};if(u&&c)return a.autoMargin(t,e,{x:o.x,y:o.y,l:o._width*g[s],r:o._width*y[s],b:o._effHeight*y[l],t:o._effHeight*g[l]});u?t._fullLayout._reservedMargin[e][f]=p[f]:c||"v"===o.orientation?t._fullLayout._reservedMargin[e][h]=p[h]:t._fullLayout._reservedMargin[e][f]=p[f]}(t,w,S,E);if(L)return;if(d.margin.autoexpand){var O=S,I=E;S=k?i.constrain(S,0,d.width-h._width):O,E=M?i.constrain(E,0,d.height-h._effHeight):I,S!==O&&i.log("Constrain "+w+".x to make legend fit inside graph"),E!==I&&i.log("Constrain "+w+".y to make legend fit inside graph")}u.setTranslate(N,S,E)}if(G.on(".drag",null),N.on("wheel",null),T||h._height<=h._maxHeight||t._context.staticPlot){var z=h._effHeight;T&&(z=h._height),U.attr({width:h._width-_,height:z-_,x:_/2,y:_/2}),u.setTranslate(q,0,0),j.select("rect").attr({width:h._width-2*_,height:z-2*_,x:_,y:_}),u.setClipUrl(q,r,t),u.setRect(G,0,0,0,0),delete h._scrollY}else{var D,R,F,B=Math.max(p.scrollBarMinHeight,h._effHeight*h._effHeight/h._height),W=h._effHeight-B-2*p.scrollBarMargin,Y=h._height-h._effHeight,X=W/Y,Z=Math.min(h._scrollY||0,Y);U.attr({width:h._width-2*_+p.scrollBarWidth+p.scrollBarMargin,height:h._effHeight-_,x:_/2,y:_/2}),j.select("rect").attr({width:h._width-2*_+p.scrollBarWidth+p.scrollBarMargin,height:h._effHeight-2*_,x:_,y:_+Z}),u.setClipUrl(q,r,t),$(Z,B,X),N.on("wheel",(function(){$(Z=i.constrain(h._scrollY+n.event.deltaY/W*Y,0,Y),B,X),0!==Z&&Z!==Y&&n.event.preventDefault()}));var K=n.behavior.drag().on("dragstart",(function(){var t=n.event.sourceEvent;D="touchstart"===t.type?t.changedTouches[0].clientY:t.clientY,F=Z})).on("drag",(function(){var t=n.event.sourceEvent;2===t.buttons||t.ctrlKey||(R="touchmove"===t.type?t.changedTouches[0].clientY:t.clientY,Z=function(t,e,r){var n=(r-e)/X+t;return i.constrain(n,0,Y)}(F,D,R),$(Z,B,X))}));G.call(K);var J=n.behavior.drag().on("dragstart",(function(){var t=n.event.sourceEvent;"touchstart"===t.type&&(D=t.changedTouches[0].clientY,F=Z)})).on("drag",(function(){var t=n.event.sourceEvent;"touchmove"===t.type&&(R=t.changedTouches[0].clientY,Z=function(t,e,r){var n=(e-r)/X+t;return i.constrain(n,0,Y)}(F,D,R),$(Z,B,X))}));q.call(J)}function $(e,r,n){h._scrollY=t._fullLayout[w]._scrollY=e,u.setTranslate(q,0,-e),u.setRect(G,h._width,p.scrollBarMargin+e*n,p.scrollBarWidth,r),j.select("rect").attr("y",_+e)}t._context.edits.legendPosition&&(N.classed("cursor-move",!0),l.init({element:N.node(),gd:t,prepFn:function(){var t=u.getTranslate(N);m=t.x,x=t.y},moveFn:function(t,r){var n=m+t,i=x+r;u.setTranslate(N,n,i),e=l.align(n,h._width,b.l,b.l+b.w,h.xanchor),c=l.align(i+h._height,-h._height,b.t+b.h,b.t,h.yanchor)},doneFn:function(){if(void 0!==e&&void 0!==c){var r={};r[w+".x"]=e,r[w+".y"]=c,o.call("_guiRelayout",t,r)}},clickFn:function(e,r){var n=s.selectAll("g.traces").filter((function(){var t=this.getBoundingClientRect();return r.clientX>=t.left&&r.clientX<=t.right&&r.clientY>=t.top&&r.clientY<=t.bottom}));n.size()>0&&A(t,N,n,e,r)}}))}],t)}}function k(t,e,r){var n=t[0],i=n.width,a=e.entrywidthmode,o=n.trace.legendwidth||e.entrywidth;return"fraction"===a?e._maxWidth*o:r+(o||i)}function A(t,e,r,n,i){var a=r.data()[0][0].trace,l={event:i,node:r.node(),curveNumber:a.index,expandedIndex:a._expandedIndex,data:t.data,layout:t.layout,frames:t._transitionData._frames,config:t._context,fullData:t._fullData,fullLayout:t._fullLayout};a._group&&(l.group=a._group),o.traceIs(a,"pie-like")&&(l.label=r.datum()[0].label);var u=s.triggerHandler(t,"plotly_legendclick",l);if(1===n){if(!1===u)return;e._clickTimeout=setTimeout((function(){t._fullLayout&&h(r,t,n)}),t._context.doubleClickDelay)}else 2===n&&(e._clickTimeout&&clearTimeout(e._clickTimeout),t._legendMouseDownTime=0,!1!==s.triggerHandler(t,"plotly_legenddoubleclick",l)&&!1!==u&&h(r,t,n))}function M(t,e,r){var n,a,s=O(r),l=t.data()[0][0],c=l.trace,h=o.traceIs(c,"pie-like"),d=!r._inHover&&e._context.edits.legendText&&!h,v=r._maxNameLength;l.groupTitle?(n=l.groupTitle.text,a=l.groupTitle.font):(a=r.font,r.entries?n=l.text:(n=h?l.label:c.name,c._meta&&(n=i.templateString(n,c._meta))));var g=i.ensureSingle(t,"text",s+"text");g.attr("text-anchor","start").call(u.font,a).text(d?S(n,v):n);var y=r.indentation+r.itemwidth+2*p.itemGap;f.positionText(g,y,0),d?g.call(f.makeEditable,{gd:e,text:n}).call(L,t,e,r).on("edit",(function(n){this.text(S(n,v)).call(L,t,e,r);var a=l.trace._fullInput||{},s={};if(o.hasTransform(a,"groupby")){var u=o.getTransformIndices(a,"groupby"),f=u[u.length-1],h=i.keyedContainer(a,"transforms["+f+"].styles","target","value.name");h.set(l.trace._group,n),s=h.constructUpdate()}else s.name=n;return a._isShape?o.call("_guiRelayout",e,"shapes["+c.index+"].name",s.name):o.call("_guiRestyle",e,s,c.index)})):L(g,t,e,r)}function S(t,e){var r=Math.max(4,e);if(t&&t.trim().length>=r/2)return t;for(var n=r-(t=t||"").length;n>0;n--)t+=" ";return t}function E(t,e,r){var a,o=e._context.doubleClickDelay,s=1,l=i.ensureSingle(t,"rect",r+"toggle",(function(t){e._context.staticPlot||t.style("cursor","pointer").attr("pointer-events","all"),t.call(c.fill,"rgba(0,0,0,0)")}));e._context.staticPlot||(l.on("mousedown",(function(){(a=(new Date).getTime())-e._legendMouseDownTime<o?s+=1:(s=1,e._legendMouseDownTime=a)})),l.on("mouseup",(function(){if(!e._dragged&&!e._editing){var i=e._fullLayout[r];(new Date).getTime()-e._legendMouseDownTime>o&&(s=Math.max(s-1,1)),A(e,i,t,s,n.event)}})))}function L(t,e,r,n,i){n._inHover&&t.attr("data-notex",!0),f.convertToTspans(t,r,(function(){!function(t,e,r,n){var i=t.data()[0][0];if(r._inHover||!i||i.trace.showlegend){var a=t.select("g[class*=math-group]"),o=a.node(),s=O(r);r||(r=e._fullLayout[s]);var l,c,h=r.borderwidth,d=(n===_?r.title.font:i.groupTitle?i.groupTitle.font:r.font).size*v;if(o){var g=u.bBox(o);l=g.height,c=g.width,n===_?u.setTranslate(a,h,h+.75*l):u.setTranslate(a,0,.25*l)}else{var y="."+s+(n===_?"title":"")+"text",m=t.select(y),x=f.lineCount(m),b=m.node();if(l=d*x,c=b?u.bBox(b).width:0,n===_)"left"===r.title.side&&(c+=2*p.itemGap),f.positionText(m,h+p.titlePad,h+d);else{var w=2*p.itemGap+r.indentation+r.itemwidth;i.groupTitle&&(w=p.itemGap,c-=r.indentation+r.itemwidth),f.positionText(m,w,-d*((x-1)/2-.3))}}n===_?(r._titleWidth=c,r._titleHeight=l):(i.lineHeight=d,i.height=Math.max(l,16)+3,i.width=c)}else t.remove()}(e,r,n,i)}))}function C(t){return i.isRightAnchor(t)?"right":i.isCenterAnchor(t)?"center":"left"}function P(t){return i.isBottomAnchor(t)?"bottom":i.isMiddleAnchor(t)?"middle":"top"}function O(t){return t._id||"legend"}t.exports=function(t,e){if(e)T(t,e);else{var r=t._fullLayout,i=r._legends;r._infolayer.selectAll('[class^="legend"]').each((function(){var t=n.select(this),e=t.attr("class").split(" ")[0];e.match(w)&&-1===i.indexOf(e)&&t.remove()}));for(var a=0;a<i.length;a++){var o=i[a];T(t,t._fullLayout[o])}}}},35456:function(t,e,r){"use strict";var n=r(24040),i=r(42451);t.exports=function(t,e,r){var a,o,s=e._inHover,l=i.isGrouped(e),u=i.isReversed(e),c={},f=[],h=!1,p={},d=0,v=0;function g(t,n,a){if(!1!==e.visible&&(!r||t===e._id))if(""!==n&&i.isGrouped(e))-1===f.indexOf(n)?(f.push(n),h=!0,c[n]=[a]):c[n].push(a);else{var o="~~i"+d;f.push(o),c[o]=[a],d++}}for(a=0;a<t.length;a++){var y=t[a],m=y[0],x=m.trace,b=x.legend,_=x.legendgroup;if(s||x.visible&&x.showlegend)if(n.traceIs(x,"pie-like"))for(p[_]||(p[_]={}),o=0;o<y.length;o++){var w=y[o].label;p[_][w]||(g(b,_,{label:w,color:y[o].color,i:y[o].i,trace:x,pts:y[o].pts}),p[_][w]=!0,v=Math.max(v,(w||"").length))}else g(b,_,m),v=Math.max(v,(x.name||"").length)}if(!f.length)return[];var T=!h||!l,k=[];for(a=0;a<f.length;a++){var A=c[f[a]];T?k.push(A[0]):k.push(A)}for(T&&(k=[k]),a=0;a<k.length;a++){var M=1/0;for(o=0;o<k[a].length;o++){var S=k[a][o].trace.legendrank;M>S&&(M=S)}k[a][0]._groupMinRank=M,k[a][0]._preGroupSort=a}var E=function(t,e){return t.trace.legendrank-e.trace.legendrank||t._preSort-e._preSort};for(k.forEach((function(t,e){t[0]._preGroupSort=e})),k.sort((function(t,e){return t[0]._groupMinRank-e[0]._groupMinRank||t[0]._preGroupSort-e[0]._preGroupSort})),a=0;a<k.length;a++){k[a].forEach((function(t,e){t._preSort=e})),k[a].sort(E);var L=k[a][0].trace,C=null;for(o=0;o<k[a].length;o++){var P=k[a][o].trace.legendgrouptitle;if(P&&P.text){C=P,s&&(P.font=e._groupTitleFont);break}}if(u&&k[a].reverse(),C){var O=!1;for(o=0;o<k[a].length;o++)if(n.traceIs(k[a][o].trace,"pie-like")){O=!0;break}k[a].unshift({i:-1,groupTitle:C,noClick:O,trace:{showlegend:L.showlegend,legendgroup:L.legendgroup,visible:"toggleitem"===e.groupclick||L.visible}})}for(o=0;o<k[a].length;o++)k[a][o]=[k[a][o]]}return e._lgroupsLength=k.length,e._maxNameLength=v,k}},33048:function(t,e,r){"use strict";var n=r(24040),i=r(3400),a=i.pushUnique,o=!0;t.exports=function(t,e,r){var s=e._fullLayout;if(!e._dragged&&!e._editing){var l,u=s.legend.itemclick,c=s.legend.itemdoubleclick,f=s.legend.groupclick;if(1===r&&"toggle"===u&&"toggleothers"===c&&o&&e.data&&e._context.showTips?(i.notifier(i._(e,"Double-click on legend to isolate one trace"),"long"),o=!1):o=!1,1===r?l=u:2===r&&(l=c),l){var h="togglegroup"===f,p=s.hiddenlabels?s.hiddenlabels.slice():[],d=t.data()[0][0];if(!d.groupTitle||!d.noClick){var v=e._fullData,g=(s.shapes||[]).filter((function(t){return t.showlegend})),y=v.concat(g),m=d.trace;m._isShape&&(m=m._fullInput);var x,b,_,w,T,k=m.legendgroup,A={},M=[],S=[],E=[],L=(s.shapes||[]).map((function(t){return t._input})),C=!1,P=m.legend,O=m._fullInput;if(O&&O._isShape||!n.traceIs(m,"pie-like")){var I,z=k&&k.length,D=[];if(z)for(x=0;x<y.length;x++)(I=y[x]).visible&&I.legendgroup===k&&D.push(x);if("toggle"===l){var R;switch(m.visible){case!0:R="legendonly";break;case!1:R=!1;break;case"legendonly":R=!0}if(z)if(h)for(x=0;x<y.length;x++){var F=y[x];!1!==F.visible&&F.legendgroup===k&&tt(F,R)}else tt(m,R);else tt(m,R)}else if("toggleothers"===l){var B,N,j,U,V=!0;for(x=0;x<y.length;x++)if(B=(U=y[x])===m,N=!0!==U.showlegend,!(B||N||z&&U.legendgroup===k||U.legend!==P||!0!==U.visible||n.traceIs(U,"notLegendIsolatable"))){V=!1;break}for(x=0;x<y.length;x++)if(!1!==(U=y[x]).visible&&U.legend===P&&!n.traceIs(U,"notLegendIsolatable"))switch(m.visible){case"legendonly":tt(U,!0);break;case!0:j=!!V||"legendonly",B=U===m,N=!0!==U.showlegend&&!U.legendgroup,tt(U,!!(B||z&&U.legendgroup===k||N)||j)}}for(x=0;x<S.length;x++)if(_=S[x]){var q=_.constructUpdate(),H=Object.keys(q);for(b=0;b<H.length;b++)w=H[b],(A[w]=A[w]||[])[E[x]]=q[w]}for(T=Object.keys(A),x=0;x<T.length;x++)for(w=T[x],b=0;b<M.length;b++)A[w].hasOwnProperty(b)||(A[w][b]=void 0);C?n.call("_guiUpdate",e,A,{shapes:L},M):n.call("_guiRestyle",e,A,M)}else{var G=d.label,W=p.indexOf(G);if("toggle"===l)-1===W?p.push(G):p.splice(W,1);else if("toggleothers"===l){var Y=-1!==W,X=[];for(x=0;x<e.calcdata.length;x++){var Z=e.calcdata[x];for(b=0;b<Z.length;b++){var K=Z[b].label;P===Z[0].trace.legend&&G!==K&&(-1===p.indexOf(K)&&(Y=!0),a(p,K),X.push(K))}}if(!Y)for(var J=0;J<X.length;J++){var $=p.indexOf(X[J]);-1!==$&&p.splice($,1)}}n.call("_guiRelayout",e,"hiddenlabels",p)}}}}function Q(t,e){var r=M.indexOf(t),n=A.visible;return n||(n=A.visible=[]),-1===M.indexOf(t)&&(M.push(t),r=M.length-1),n[r]=e,r}function tt(t,e){if(!d.groupTitle||h){var r,a=t._fullInput||t,o=a._isShape,s=a.index;if(void 0===s&&(s=a._index),n.hasTransform(a,"groupby")){var l=S[s];if(!l){var u=n.getTransformIndices(a,"groupby"),c=u[u.length-1];l=i.keyedContainer(a,"transforms["+c+"].styles","target","value.visible"),S[s]=l}var f=l.get(t._group);void 0===f&&(f=!0),!1!==f&&l.set(t._group,e),E[s]=Q(s,!1!==a.visible)}else{var p=!1!==a.visible&&e;o?(r=p,L[s].visible=r,C=!0):Q(s,p)}}}}},42451:function(t,e){"use strict";e.isGrouped=function(t){return-1!==(t.traceorder||"").indexOf("grouped")},e.isVertical=function(t){return"h"!==t.orientation},e.isReversed=function(t){return-1!==(t.traceorder||"").indexOf("reversed")}},2780:function(t,e,r){"use strict";t.exports={moduleType:"component",name:"legend",layoutAttributes:r(3800),supplyLayoutDefaults:r(77864),draw:r(31140),style:r(2012)}},2012:function(t,e,r){"use strict";var n=r(33428),i=r(24040),a=r(3400),o=a.strTranslate,s=r(43616),l=r(76308),u=r(94288).extractOpts,c=r(43028),f=r(10528),h=r(69656).castOption,p=r(65196);function d(t,e){return(e?"radial":"horizontal")+(t?"":"reversed")}function v(t){var e=t[0].trace,r=e.contours,n=c.hasLines(e),i=c.hasMarkers(e),a=e.visible&&e.fill&&"none"!==e.fill,o=!1,s=!1;if(r){var l=r.coloring;"lines"===l?o=!0:n="none"===l||"heatmap"===l||r.showlines,"constraint"===r.type?a="="!==r._operation:"fill"!==l&&"heatmap"!==l||(s=!0)}return{showMarker:i,showLine:n,showFill:a,showGradientLine:o,showGradientFill:s,anyLine:n||o,anyFill:a||s}}function g(t,e,r){return t&&a.isArrayOrTypedArray(t)?e:t>r?r:t}t.exports=function(t,e,r){var y=e._fullLayout;r||(r=y.legend);var m="constant"===r.itemsizing,x=r.itemwidth,b=(x+2*p.itemGap)/2,_=o(b,0),w=function(t,e,r,n){var i;if(t+1)i=t;else{if(!(e&&e.width>0))return 0;i=e.width}return m?n:Math.min(i,r)};function T(t,a,o){var c=t[0].trace,f=c.marker||{},h=f.line||{},p=f.cornerradius?"M6,3a3,3,0,0,1-3,3H-3a3,3,0,0,1-3-3V-3a3,3,0,0,1,3-3H3a3,3,0,0,1,3,3Z":"M6,6H-6V-6H6Z",d=o?c.visible&&c.type===o:i.traceIs(c,"bar"),v=n.select(a).select("g.legendpoints").selectAll("path.legend"+o).data(d?[t]:[]);v.enter().append("path").classed("legend"+o,!0).attr("d",p).attr("transform",_),v.exit().remove(),v.each((function(t){var i=n.select(this),a=t[0],o=w(a.mlw,f.line,5,2);i.style("stroke-width",o+"px");var p=a.mcc;if(!r._inHover&&"mc"in a){var d=u(f),v=d.mid;void 0===v&&(v=(d.max+d.min)/2),p=s.tryColorscale(f,"")(v)}var y=p||a.mc||f.color,m=f.pattern,x=m&&s.getPatternAttr(m.shape,0,"");if(x){var b=s.getPatternAttr(m.bgcolor,0,null),_=s.getPatternAttr(m.fgcolor,0,null),T=m.fgopacity,k=g(m.size,8,10),A=g(m.solidity,.5,1),M="legend-"+c.uid;i.call(s.pattern,"legend",e,M,x,k,A,p,m.fillmode,b,_,T)}else i.call(l.fill,y);o&&l.stroke(i,a.mlc||h.color)}))}function k(t,r,o){var s=t[0],l=s.trace,u=o?l.visible&&l.type===o:i.traceIs(l,o),c=n.select(r).select("g.legendpoints").selectAll("path.legend"+o).data(u?[t]:[]);if(c.enter().append("path").classed("legend"+o,!0).attr("d","M6,6H-6V-6H6Z").attr("transform",_),c.exit().remove(),c.size()){var p=l.marker||{},d=w(h(p.line.width,s.pts),p.line,5,2),v="pieLike",g=a.minExtend(l,{marker:{line:{width:d}}},v),y=a.minExtend(s,{trace:g},v);f(c,y,g,e)}}t.each((function(t){var e=n.select(this),i=a.ensureSingle(e,"g","layers");i.style("opacity",t[0].trace.opacity);var s=r.indentation,l=r.valign,u=t[0].lineHeight,c=t[0].height;if("middle"===l&&0===s||!u||!c)i.attr("transform",null);else{var f={top:1,bottom:-1}[l]*(.5*(u-c+3))||0,h=r.indentation;i.attr("transform",o(h,f))}i.selectAll("g.legendfill").data([t]).enter().append("g").classed("legendfill",!0),i.selectAll("g.legendlines").data([t]).enter().append("g").classed("legendlines",!0);var p=i.selectAll("g.legendsymbols").data([t]);p.enter().append("g").classed("legendsymbols",!0),p.selectAll("g.legendpoints").data([t]).enter().append("g").classed("legendpoints",!0)})).each((function(t){var r,i=t[0].trace,o=[];if(i.visible)switch(i.type){case"histogram2d":case"heatmap":o=[["M-15,-2V4H15V-2Z"]],r=!0;break;case"choropleth":case"choroplethmapbox":o=[["M-6,-6V6H6V-6Z"]],r=!0;break;case"densitymapbox":o=[["M-6,0 a6,6 0 1,0 12,0 a 6,6 0 1,0 -12,0"]],r="radial";break;case"cone":o=[["M-6,2 A2,2 0 0,0 -6,6 V6L6,4Z"],["M-6,-6 A2,2 0 0,0 -6,-2 L6,-4Z"],["M-6,-2 A2,2 0 0,0 -6,2 L6,0Z"]],r=!1;break;case"streamtube":o=[["M-6,2 A2,2 0 0,0 -6,6 H6 A2,2 0 0,1 6,2 Z"],["M-6,-6 A2,2 0 0,0 -6,-2 H6 A2,2 0 0,1 6,-6 Z"],["M-6,-2 A2,2 0 0,0 -6,2 H6 A2,2 0 0,1 6,-2 Z"]],r=!1;break;case"surface":o=[["M-6,-6 A2,3 0 0,0 -6,0 H6 A2,3 0 0,1 6,-6 Z"],["M-6,1 A2,3 0 0,1 -6,6 H6 A2,3 0 0,0 6,0 Z"]],r=!0;break;case"mesh3d":o=[["M-6,6H0L-6,-6Z"],["M6,6H0L6,-6Z"],["M-6,-6H6L0,6Z"]],r=!1;break;case"volume":o=[["M-6,6H0L-6,-6Z"],["M6,6H0L6,-6Z"],["M-6,-6H6L0,6Z"]],r=!0;break;case"isosurface":o=[["M-6,6H0L-6,-6Z"],["M6,6H0L6,-6Z"],["M-6,-6 A12,24 0 0,0 6,-6 L0,6Z"]],r=!1}var c=n.select(this).select("g.legendpoints").selectAll("path.legend3dandfriends").data(o);c.enter().append("path").classed("legend3dandfriends",!0).attr("transform",_).style("stroke-miterlimit",1),c.exit().remove(),c.each((function(t,o){var c,f=n.select(this),h=u(i),p=h.colorscale,v=h.reversescale;if(p){if(!r){var g=p.length;c=0===o?p[v?g-1:0][1]:1===o?p[v?0:g-1][1]:p[Math.floor((g-1)/2)][1]}}else{var y=i.vertexcolor||i.facecolor||i.color;c=a.isArrayOrTypedArray(y)?y[o]||y[0]:y}f.attr("d",t[0]),c?f.call(l.fill,c):f.call((function(t){if(t.size()){var n="legendfill-"+i.uid;s.gradient(t,e,n,d(v,"radial"===r),p,"fill")}}))}))})).each((function(t){var e=t[0].trace,r="waterfall"===e.type;if(t[0]._distinct&&r){var i=t[0].trace[t[0].dir].marker;return t[0].mc=i.color,t[0].mlw=i.line.width,t[0].mlc=i.line.color,T(t,this,"waterfall")}var a=[];e.visible&&r&&(a=t[0].hasTotals?[["increasing","M-6,-6V6H0Z"],["totals","M6,6H0L-6,-6H-0Z"],["decreasing","M6,6V-6H0Z"]]:[["increasing","M-6,-6V6H6Z"],["decreasing","M6,6V-6H-6Z"]]);var o=n.select(this).select("g.legendpoints").selectAll("path.legendwaterfall").data(a);o.enter().append("path").classed("legendwaterfall",!0).attr("transform",_).style("stroke-miterlimit",1),o.exit().remove(),o.each((function(t){var r=n.select(this),i=e[t[0]].marker,a=w(void 0,i.line,5,2);r.attr("d",t[1]).style("stroke-width",a+"px").call(l.fill,i.color),a&&r.call(l.stroke,i.line.color)}))})).each((function(t){T(t,this,"funnel")})).each((function(t){T(t,this)})).each((function(t){var r=t[0].trace,o=n.select(this).select("g.legendpoints").selectAll("path.legendbox").data(r.visible&&i.traceIs(r,"box-violin")?[t]:[]);o.enter().append("path").classed("legendbox",!0).attr("d","M6,6H-6V-6H6Z").attr("transform",_),o.exit().remove(),o.each((function(){var t=n.select(this);if("all"!==r.boxpoints&&"all"!==r.points||0!==l.opacity(r.fillcolor)||0!==l.opacity((r.line||{}).color)){var i=w(void 0,r.line,5,2);t.style("stroke-width",i+"px").call(l.fill,r.fillcolor),i&&l.stroke(t,r.line.color)}else{var u=a.minExtend(r,{marker:{size:m?12:a.constrain(r.marker.size,2,16),sizeref:1,sizemin:1,sizemode:"diameter"}});o.call(s.pointStyle,u,e)}}))})).each((function(t){k(t,this,"funnelarea")})).each((function(t){k(t,this,"pie")})).each((function(t){var r,i,o=v(t),l=o.showFill,f=o.showLine,h=o.showGradientLine,p=o.showGradientFill,g=o.anyFill,y=o.anyLine,m=t[0],b=m.trace,_=u(b),T=_.colorscale,k=_.reversescale,A=c.hasMarkers(b)||!g?"M5,0":y?"M5,-2":"M5,-3",M=n.select(this),S=M.select(".legendfill").selectAll("path").data(l||p?[t]:[]);if(S.enter().append("path").classed("js-fill",!0),S.exit().remove(),S.attr("d",A+"h"+x+"v6h-"+x+"z").call((function(t){if(t.size())if(l)s.fillGroupStyle(t,e,!0);else{var r="legendfill-"+b.uid;s.gradient(t,e,r,d(k),T,"fill")}})),f||h){var E=w(void 0,b.line,10,5);i=a.minExtend(b,{line:{width:E}}),r=[a.minExtend(m,{trace:i})]}var L=M.select(".legendlines").selectAll("path").data(f||h?[r]:[]);L.enter().append("path").classed("js-line",!0),L.exit().remove(),L.attr("d",A+(h?"l"+x+",0.0001":"h"+x)).call(f?s.lineGroupStyle:function(t){if(t.size()){var r="legendline-"+b.uid;s.lineGroupStyle(t),s.gradient(t,e,r,d(k),T,"stroke")}})})).each((function(t){var r,i,o=v(t),l=o.anyFill,u=o.anyLine,f=o.showLine,h=o.showMarker,p=t[0],d=p.trace,g=!h&&!u&&!l&&c.hasText(d);function y(t,e,r,n){var i=a.nestedProperty(d,t).get(),o=a.isArrayOrTypedArray(i)&&e?e(i):i;if(m&&o&&void 0!==n&&(o=n),r){if(o<r[0])return r[0];if(o>r[1])return r[1]}return o}function x(t){return p._distinct&&p.index&&t[p.index]?t[p.index]:t[0]}if(h||g||f){var b={},w={};if(h){b.mc=y("marker.color",x),b.mx=y("marker.symbol",x),b.mo=y("marker.opacity",a.mean,[.2,1]),b.mlc=y("marker.line.color",x),b.mlw=y("marker.line.width",a.mean,[0,5],2),w.marker={sizeref:1,sizemin:1,sizemode:"diameter"};var T=y("marker.size",a.mean,[2,16],12);b.ms=T,w.marker.size=T}f&&(w.line={width:y("line.width",x,[0,10],5)}),g&&(b.tx="Aa",b.tp=y("textposition",x),b.ts=10,b.tc=y("textfont.color",x),b.tf=y("textfont.family",x),b.tw=y("textfont.weight",x),b.ty=y("textfont.style",x),b.tv=y("textfont.variant",x)),r=[a.minExtend(p,b)],(i=a.minExtend(d,w)).selectedpoints=null,i.texttemplate=null}var k=n.select(this).select("g.legendpoints"),A=k.selectAll("path.scatterpts").data(h?r:[]);A.enter().insert("path",":first-child").classed("scatterpts",!0).attr("transform",_),A.exit().remove(),A.call(s.pointStyle,i,e),h&&(r[0].mrc=3);var M=k.selectAll("g.pointtext").data(g?r:[]);M.enter().append("g").classed("pointtext",!0).append("text").attr("transform",_),M.exit().remove(),M.selectAll("text").call(s.textPointStyle,i,e)})).each((function(t){var e=t[0].trace,r=n.select(this).select("g.legendpoints").selectAll("path.legendcandle").data(e.visible&&"candlestick"===e.type?[t,t]:[]);r.enter().append("path").classed("legendcandle",!0).attr("d",(function(t,e){return e?"M-15,0H-8M-8,6V-6H8Z":"M15,0H8M8,-6V6H-8Z"})).attr("transform",_).style("stroke-miterlimit",1),r.exit().remove(),r.each((function(t,r){var i=n.select(this),a=e[r?"increasing":"decreasing"],o=w(void 0,a.line,5,2);i.style("stroke-width",o+"px").call(l.fill,a.fillcolor),o&&l.stroke(i,a.line.color)}))})).each((function(t){var e=t[0].trace,r=n.select(this).select("g.legendpoints").selectAll("path.legendohlc").data(e.visible&&"ohlc"===e.type?[t,t]:[]);r.enter().append("path").classed("legendohlc",!0).attr("d",(function(t,e){return e?"M-15,0H0M-8,-6V0":"M15,0H0M8,6V0"})).attr("transform",_).style("stroke-miterlimit",1),r.exit().remove(),r.each((function(t,r){var i=n.select(this),a=e[r?"increasing":"decreasing"],o=w(void 0,a.line,5,2);i.style("fill","none").call(s.dashLine,a.line.dash,o),o&&l.stroke(i,a.line.color)}))}))}},66540:function(t,e,r){"use strict";r(76052),t.exports={editType:"modebar",orientation:{valType:"enumerated",values:["v","h"],dflt:"h",editType:"modebar"},bgcolor:{valType:"color",editType:"modebar"},color:{valType:"color",editType:"modebar"},activecolor:{valType:"color",editType:"modebar"},uirevision:{valType:"any",editType:"none"},add:{valType:"string",arrayOk:!0,dflt:"",editType:"modebar"},remove:{valType:"string",arrayOk:!0,dflt:"",editType:"modebar"}}},44248:function(t,e,r){"use strict";var n=r(24040),i=r(7316),a=r(79811),o=r(9224),s=r(4016).eraseActiveShape,l=r(3400),u=l._,c=t.exports={};function f(t,e){var r,i,o=e.currentTarget,s=o.getAttribute("data-attr"),l=o.getAttribute("data-val")||!0,u=t._fullLayout,c={},f=a.list(t,null,!0),h=u._cartesianSpikesEnabled;if("zoom"===s){var p,d="in"===l?.5:2,v=(1+d)/2,g=(1-d)/2;for(i=0;i<f.length;i++)if(!(r=f[i]).fixedrange)if(p=r._name,"auto"===l)c[p+".autorange"]=!0;else if("reset"===l)void 0===r._rangeInitial0&&void 0===r._rangeInitial1?c[p+".autorange"]=!0:void 0===r._rangeInitial0?(c[p+".autorange"]=r._autorangeInitial,c[p+".range"]=[null,r._rangeInitial1]):void 0===r._rangeInitial1?(c[p+".range"]=[r._rangeInitial0,null],c[p+".autorange"]=r._autorangeInitial):c[p+".range"]=[r._rangeInitial0,r._rangeInitial1],void 0!==r._showSpikeInitial&&(c[p+".showspikes"]=r._showSpikeInitial,"on"!==h||r._showSpikeInitial||(h="off"));else{var y=[r.r2l(r.range[0]),r.r2l(r.range[1])],m=[v*y[0]+g*y[1],v*y[1]+g*y[0]];c[p+".range[0]"]=r.l2r(m[0]),c[p+".range[1]"]=r.l2r(m[1])}}else"hovermode"!==s||"x"!==l&&"y"!==l||(l=u._isHoriz?"y":"x",o.setAttribute("data-val",l)),c[s]=l;u._cartesianSpikesEnabled=h,n.call("_guiRelayout",t,c)}function h(t,e){for(var r=e.currentTarget,i=r.getAttribute("data-attr"),a=r.getAttribute("data-val")||!0,o=t._fullLayout._subplots.gl3d||[],s={},l=i.split("."),u=0;u<o.length;u++)s[o[u]+"."+l[1]]=a;var c="pan"===a?a:"zoom";s.dragmode=c,n.call("_guiRelayout",t,s)}function p(t,e){for(var r=e.currentTarget.getAttribute("data-attr"),i="resetLastSave"===r,a="resetDefault"===r,o=t._fullLayout,s=o._subplots.gl3d||[],l={},u=0;u<s.length;u++){var c,f=s[u],h=f+".camera",p=f+".aspectratio",d=f+".aspectmode",v=o[f]._scene;i?(l[h+".up"]=v.viewInitial.up,l[h+".eye"]=v.viewInitial.eye,l[h+".center"]=v.viewInitial.center,c=!0):a&&(l[h+".up"]=null,l[h+".eye"]=null,l[h+".center"]=null,c=!0),c&&(l[p+".x"]=v.viewInitial.aspectratio.x,l[p+".y"]=v.viewInitial.aspectratio.y,l[p+".z"]=v.viewInitial.aspectratio.z,l[d]=v.viewInitial.aspectmode)}n.call("_guiRelayout",t,l)}function d(t,e){var r=e.currentTarget,n=r._previousVal,i=t._fullLayout,a=i._subplots.gl3d||[],o=["xaxis","yaxis","zaxis"],s={},l={};if(n)l=n,r._previousVal=null;else{for(var u=0;u<a.length;u++){var c=a[u],f=i[c],h=c+".hovermode";s[h]=f.hovermode,l[h]=!1;for(var p=0;p<3;p++){var d=o[p],v=c+"."+d+".showspikes";l[v]=!1,s[v]=f[d].showspikes}}r._previousVal=s}return l}function v(t,e){for(var r=e.currentTarget,i=r.getAttribute("data-attr"),a=r.getAttribute("data-val")||!0,o=t._fullLayout,s=o._subplots.geo||[],l=0;l<s.length;l++){var u=s[l],c=o[u];if("zoom"===i){var f=c.projection.scale,h="in"===a?2*f:.5*f;n.call("_guiRelayout",t,u+".projection.scale",h)}}"reset"===i&&x(t,"geo")}function g(t){var e=t._fullLayout;return!e.hovermode&&(e._has("cartesian")?e._isHoriz?"y":"x":"closest")}function y(t){var e=g(t);n.call("_guiRelayout",t,"hovermode",e)}function m(t,e){for(var r=e.currentTarget.getAttribute("data-val"),i=t._fullLayout,a=i._subplots.mapbox||[],o={},s=0;s<a.length;s++){var l=a[s],u=i[l].zoom,c="in"===r?1.05*u:u/1.05;o[l+".zoom"]=c}n.call("_guiRelayout",t,o)}function x(t,e){for(var r=t._fullLayout,i=r._subplots[e]||[],a={},o=0;o<i.length;o++)for(var s=i[o],l=r[s]._subplot.viewInitial,u=Object.keys(l),c=0;c<u.length;c++){var f=u[c];a[s+"."+f]=l[f]}n.call("_guiRelayout",t,a)}c.toImage={name:"toImage",title:function(t){var e=(t._context.toImageButtonOptions||{}).format||"png";return u(t,"png"===e?"Download plot as a png":"Download plot")},icon:o.camera,click:function(t){var e=t._context.toImageButtonOptions,r={format:e.format||"png"};l.notifier(u(t,"Taking snapshot - this may take a few seconds"),"long"),"svg"!==r.format&&l.isIE()&&(l.notifier(u(t,"IE only supports svg.  Changing format to svg."),"long"),r.format="svg"),["filename","width","height","scale"].forEach((function(t){t in e&&(r[t]=e[t])})),n.call("downloadImage",t,r).then((function(e){l.notifier(u(t,"Snapshot succeeded")+" - "+e,"long")})).catch((function(){l.notifier(u(t,"Sorry, there was a problem downloading your snapshot!"),"long")}))}},c.sendDataToCloud={name:"sendDataToCloud",title:function(t){return u(t,"Edit in Chart Studio")},icon:o.disk,click:function(t){i.sendDataToCloud(t)}},c.editInChartStudio={name:"editInChartStudio",title:function(t){return u(t,"Edit in Chart Studio")},icon:o.pencil,click:function(t){i.sendDataToCloud(t)}},c.zoom2d={name:"zoom2d",_cat:"zoom",title:function(t){return u(t,"Zoom")},attr:"dragmode",val:"zoom",icon:o.zoombox,click:f},c.pan2d={name:"pan2d",_cat:"pan",title:function(t){return u(t,"Pan")},attr:"dragmode",val:"pan",icon:o.pan,click:f},c.select2d={name:"select2d",_cat:"select",title:function(t){return u(t,"Box Select")},attr:"dragmode",val:"select",icon:o.selectbox,click:f},c.lasso2d={name:"lasso2d",_cat:"lasso",title:function(t){return u(t,"Lasso Select")},attr:"dragmode",val:"lasso",icon:o.lasso,click:f},c.drawclosedpath={name:"drawclosedpath",title:function(t){return u(t,"Draw closed freeform")},attr:"dragmode",val:"drawclosedpath",icon:o.drawclosedpath,click:f},c.drawopenpath={name:"drawopenpath",title:function(t){return u(t,"Draw open freeform")},attr:"dragmode",val:"drawopenpath",icon:o.drawopenpath,click:f},c.drawline={name:"drawline",title:function(t){return u(t,"Draw line")},attr:"dragmode",val:"drawline",icon:o.drawline,click:f},c.drawrect={name:"drawrect",title:function(t){return u(t,"Draw rectangle")},attr:"dragmode",val:"drawrect",icon:o.drawrect,click:f},c.drawcircle={name:"drawcircle",title:function(t){return u(t,"Draw circle")},attr:"dragmode",val:"drawcircle",icon:o.drawcircle,click:f},c.eraseshape={name:"eraseshape",title:function(t){return u(t,"Erase active shape")},icon:o.eraseshape,click:s},c.zoomIn2d={name:"zoomIn2d",_cat:"zoomin",title:function(t){return u(t,"Zoom in")},attr:"zoom",val:"in",icon:o.zoom_plus,click:f},c.zoomOut2d={name:"zoomOut2d",_cat:"zoomout",title:function(t){return u(t,"Zoom out")},attr:"zoom",val:"out",icon:o.zoom_minus,click:f},c.autoScale2d={name:"autoScale2d",_cat:"autoscale",title:function(t){return u(t,"Autoscale")},attr:"zoom",val:"auto",icon:o.autoscale,click:f},c.resetScale2d={name:"resetScale2d",_cat:"resetscale",title:function(t){return u(t,"Reset axes")},attr:"zoom",val:"reset",icon:o.home,click:f},c.hoverClosestCartesian={name:"hoverClosestCartesian",_cat:"hoverclosest",title:function(t){return u(t,"Show closest data on hover")},attr:"hovermode",val:"closest",icon:o.tooltip_basic,gravity:"ne",click:f},c.hoverCompareCartesian={name:"hoverCompareCartesian",_cat:"hoverCompare",title:function(t){return u(t,"Compare data on hover")},attr:"hovermode",val:function(t){return t._fullLayout._isHoriz?"y":"x"},icon:o.tooltip_compare,gravity:"ne",click:f},c.zoom3d={name:"zoom3d",_cat:"zoom",title:function(t){return u(t,"Zoom")},attr:"scene.dragmode",val:"zoom",icon:o.zoombox,click:h},c.pan3d={name:"pan3d",_cat:"pan",title:function(t){return u(t,"Pan")},attr:"scene.dragmode",val:"pan",icon:o.pan,click:h},c.orbitRotation={name:"orbitRotation",title:function(t){return u(t,"Orbital rotation")},attr:"scene.dragmode",val:"orbit",icon:o["3d_rotate"],click:h},c.tableRotation={name:"tableRotation",title:function(t){return u(t,"Turntable rotation")},attr:"scene.dragmode",val:"turntable",icon:o["z-axis"],click:h},c.resetCameraDefault3d={name:"resetCameraDefault3d",_cat:"resetCameraDefault",title:function(t){return u(t,"Reset camera to default")},attr:"resetDefault",icon:o.home,click:p},c.resetCameraLastSave3d={name:"resetCameraLastSave3d",_cat:"resetCameraLastSave",title:function(t){return u(t,"Reset camera to last save")},attr:"resetLastSave",icon:o.movie,click:p},c.hoverClosest3d={name:"hoverClosest3d",_cat:"hoverclosest",title:function(t){return u(t,"Toggle show closest data on hover")},attr:"hovermode",val:null,toggle:!0,icon:o.tooltip_basic,gravity:"ne",click:function(t,e){var r=d(t,e);n.call("_guiRelayout",t,r)}},c.zoomInGeo={name:"zoomInGeo",_cat:"zoomin",title:function(t){return u(t,"Zoom in")},attr:"zoom",val:"in",icon:o.zoom_plus,click:v},c.zoomOutGeo={name:"zoomOutGeo",_cat:"zoomout",title:function(t){return u(t,"Zoom out")},attr:"zoom",val:"out",icon:o.zoom_minus,click:v},c.resetGeo={name:"resetGeo",_cat:"reset",title:function(t){return u(t,"Reset")},attr:"reset",val:null,icon:o.autoscale,click:v},c.hoverClosestGeo={name:"hoverClosestGeo",_cat:"hoverclosest",title:function(t){return u(t,"Toggle show closest data on hover")},attr:"hovermode",val:null,toggle:!0,icon:o.tooltip_basic,gravity:"ne",click:y},c.hoverClosestGl2d={name:"hoverClosestGl2d",_cat:"hoverclosest",title:function(t){return u(t,"Toggle show closest data on hover")},attr:"hovermode",val:null,toggle:!0,icon:o.tooltip_basic,gravity:"ne",click:y},c.hoverClosestPie={name:"hoverClosestPie",_cat:"hoverclosest",title:function(t){return u(t,"Toggle show closest data on hover")},attr:"hovermode",val:"closest",icon:o.tooltip_basic,gravity:"ne",click:y},c.resetViewSankey={name:"resetSankeyGroup",title:function(t){return u(t,"Reset view")},icon:o.home,click:function(t){for(var e={"node.groups":[],"node.x":[],"node.y":[]},r=0;r<t._fullData.length;r++){var i=t._fullData[r]._viewInitial;e["node.groups"].push(i.node.groups.slice()),e["node.x"].push(i.node.x.slice()),e["node.y"].push(i.node.y.slice())}n.call("restyle",t,e)}},c.toggleHover={name:"toggleHover",title:function(t){return u(t,"Toggle show closest data on hover")},attr:"hovermode",val:null,toggle:!0,icon:o.tooltip_basic,gravity:"ne",click:function(t,e){var r=d(t,e);r.hovermode=g(t),n.call("_guiRelayout",t,r)}},c.resetViews={name:"resetViews",title:function(t){return u(t,"Reset views")},icon:o.home,click:function(t,e){var r=e.currentTarget;r.setAttribute("data-attr","zoom"),r.setAttribute("data-val","reset"),f(t,e),r.setAttribute("data-attr","resetLastSave"),p(t,e),x(t,"geo"),x(t,"mapbox")}},c.toggleSpikelines={name:"toggleSpikelines",title:function(t){return u(t,"Toggle Spike Lines")},icon:o.spikeline,attr:"_cartesianSpikesEnabled",val:"on",click:function(t){var e=t._fullLayout,r=e._cartesianSpikesEnabled;e._cartesianSpikesEnabled="on"===r?"off":"on",n.call("_guiRelayout",t,function(t){for(var e="on"===t._fullLayout._cartesianSpikesEnabled,r=a.list(t,null,!0),n={},i=0;i<r.length;i++){var o=r[i];n[o._name+".showspikes"]=!!e||o._showSpikeInitial}return n}(t))}},c.resetViewMapbox={name:"resetViewMapbox",_cat:"resetView",title:function(t){return u(t,"Reset view")},attr:"reset",icon:o.home,click:function(t){x(t,"mapbox")}},c.zoomInMapbox={name:"zoomInMapbox",_cat:"zoomin",title:function(t){return u(t,"Zoom in")},attr:"zoom",val:"in",icon:o.zoom_plus,click:m},c.zoomOutMapbox={name:"zoomOutMapbox",_cat:"zoomout",title:function(t){return u(t,"Zoom out")},attr:"zoom",val:"out",icon:o.zoom_minus,click:m}},76052:function(t,e,r){"use strict";var n=r(44248),i=Object.keys(n),a=["drawline","drawopenpath","drawclosedpath","drawcircle","drawrect","eraseshape"],o=["v1hovermode","hoverclosest","hovercompare","togglehover","togglespikelines"].concat(a),s=[];i.forEach((function(t){!function(t){if(-1===o.indexOf(t._cat||t.name)){var e=t.name,r=(t._cat||t.name).toLowerCase();-1===s.indexOf(e)&&s.push(e),-1===s.indexOf(r)&&s.push(r)}}(n[t])})),s.sort(),t.exports={DRAW_MODES:a,backButtons:o,foreButtons:s}},90824:function(t,e,r){"use strict";var n=r(3400),i=r(76308),a=r(31780),o=r(66540);t.exports=function(t,e){var r=t.modebar||{},s=a.newContainer(e,"modebar");function l(t,e){return n.coerce(r,s,o,t,e)}l("orientation"),l("bgcolor",i.addOpacity(e.paper_bgcolor,.5));var u=i.contrast(i.rgb(e.modebar.bgcolor));l("color",i.addOpacity(u,.3)),l("activecolor",i.addOpacity(u,.7)),l("uirevision",e.uirevision),l("add"),l("remove")}},45460:function(t,e,r){"use strict";t.exports={moduleType:"component",name:"modebar",layoutAttributes:r(66540),supplyLayoutDefaults:r(90824),manage:r(18816)}},18816:function(t,e,r){"use strict";var n=r(79811),i=r(43028),a=r(24040),o=r(10624).isUnifiedHover,s=r(66400),l=r(44248),u=r(76052).DRAW_MODES,c=r(3400).extendDeep;t.exports=function(t){var e=t._fullLayout,r=t._context,f=e._modeBar;if(r.displayModeBar||r.watermark){if(!Array.isArray(r.modeBarButtonsToRemove))throw new Error(["*modeBarButtonsToRemove* configuration options","must be an array."].join(" "));if(!Array.isArray(r.modeBarButtonsToAdd))throw new Error(["*modeBarButtonsToAdd* configuration options","must be an array."].join(" "));var h,p=r.modeBarButtons;h=Array.isArray(p)&&p.length?function(t){for(var e=c([],t),r=0;r<e.length;r++)for(var n=e[r],i=0;i<n.length;i++){var a=n[i];if("string"==typeof a){if(void 0===l[a])throw new Error(["*modeBarButtons* configuration options","invalid button name"].join(" "));e[r][i]=l[a]}}return e}(p):!r.displayModeBar&&r.watermark?[]:function(t){var e=t._fullLayout,r=t._fullData,s=t._context;function c(t,e){if("string"==typeof e){if(e.toLowerCase()===t.toLowerCase())return!0}else{var r=e.name,n=e._cat||e.name;if(r===t||n===t.toLowerCase())return!0}return!1}var f=e.modebar.add;"string"==typeof f&&(f=[f]);var h=e.modebar.remove;"string"==typeof h&&(h=[h]);var p=s.modeBarButtonsToAdd.concat(f.filter((function(t){for(var e=0;e<s.modeBarButtonsToRemove.length;e++)if(c(t,s.modeBarButtonsToRemove[e]))return!1;return!0}))),d=s.modeBarButtonsToRemove.concat(h.filter((function(t){for(var e=0;e<s.modeBarButtonsToAdd.length;e++)if(c(t,s.modeBarButtonsToAdd[e]))return!1;return!0}))),v=e._has("cartesian"),g=e._has("gl3d"),y=e._has("geo"),m=e._has("pie"),x=e._has("funnelarea"),b=e._has("gl2d"),_=e._has("ternary"),w=e._has("mapbox"),T=e._has("polar"),k=e._has("smith"),A=e._has("sankey"),M=function(t){for(var e=n.list({_fullLayout:t},null,!0),r=0;r<e.length;r++)if(!e[r].fixedrange)return!1;return!0}(e),S=o(e.hovermode),E=[];function L(t){if(t.length){for(var e=[],r=0;r<t.length;r++){for(var n=t[r],i=l[n],a=i.name.toLowerCase(),o=(i._cat||i.name).toLowerCase(),s=!1,u=0;u<d.length;u++){var c=d[u].toLowerCase();if(c===a||c===o){s=!0;break}}s||e.push(l[n])}E.push(e)}}var C=["toImage"];s.showEditInChartStudio?C.push("editInChartStudio"):s.showSendToCloud&&C.push("sendDataToCloud"),L(C);var P=[],O=[],I=[],z=[];(v||b||m||x||_)+y+g+w+T+k>1?(O=["toggleHover"],I=["resetViews"]):y?(P=["zoomInGeo","zoomOutGeo"],O=["hoverClosestGeo"],I=["resetGeo"]):g?(O=["hoverClosest3d"],I=["resetCameraDefault3d","resetCameraLastSave3d"]):w?(P=["zoomInMapbox","zoomOutMapbox"],O=["toggleHover"],I=["resetViewMapbox"]):b?O=["hoverClosestGl2d"]:m?O=["hoverClosestPie"]:A?(O=["hoverClosestCartesian","hoverCompareCartesian"],I=["resetViewSankey"]):O=["toggleHover"],v&&(O=["toggleSpikelines","hoverClosestCartesian","hoverCompareCartesian"]),(function(t){for(var e=0;e<t.length;e++)if(!a.traceIs(t[e],"noHover"))return!1;return!0}(r)||S)&&(O=[]),!v&&!b||M||(P=["zoomIn2d","zoomOut2d","autoScale2d"],"resetViews"!==I[0]&&(I=["resetScale2d"])),g?z=["zoom3d","pan3d","orbitRotation","tableRotation"]:(v||b)&&!M||_?z=["zoom2d","pan2d"]:w||y?z=["pan2d"]:T&&(z=["zoom2d"]),function(t){for(var e=!1,r=0;r<t.length&&!e;r++){var n=t[r];n._module&&n._module.selectPoints&&(a.traceIs(n,"scatter-like")?(i.hasMarkers(n)||i.hasText(n))&&(e=!0):a.traceIs(n,"box-violin")&&"all"!==n.boxpoints&&"all"!==n.points||(e=!0))}return e}(r)&&z.push("select2d","lasso2d");var D=[],R=function(t){-1===D.indexOf(t)&&-1!==O.indexOf(t)&&D.push(t)};if(Array.isArray(p)){for(var F=[],B=0;B<p.length;B++){var N=p[B];"string"==typeof N?(N=N.toLowerCase(),-1!==u.indexOf(N)?(e._has("mapbox")||e._has("cartesian"))&&z.push(N):"togglespikelines"===N?R("toggleSpikelines"):"togglehover"===N?R("toggleHover"):"hovercompare"===N?R("hoverCompareCartesian"):"hoverclosest"===N?(R("hoverClosestCartesian"),R("hoverClosestGeo"),R("hoverClosest3d"),R("hoverClosestGl2d"),R("hoverClosestPie")):"v1hovermode"===N&&(R("toggleHover"),R("hoverClosestCartesian"),R("hoverCompareCartesian"),R("hoverClosestGeo"),R("hoverClosest3d"),R("hoverClosestGl2d"),R("hoverClosestPie"))):F.push(N)}p=F}return L(z),L(P.concat(I)),L(D),function(t,e){if(e.length)if(Array.isArray(e[0]))for(var r=0;r<e.length;r++)t.push(e[r]);else t.push(e);return t}(E,p)}(t),f?f.update(t,h):e._modeBar=s(t,h)}else f&&(f.destroy(),delete e._modeBar)}},66400:function(t,e,r){"use strict";var n=r(33428),i=r(38248),a=r(3400),o=r(9224),s=r(25788).version,l=new DOMParser;function u(t){this.container=t.container,this.element=document.createElement("div"),this.update(t.graphInfo,t.buttons),this.container.appendChild(this.element)}var c=u.prototype;c.update=function(t,e){this.graphInfo=t;var r=this.graphInfo._context,n=this.graphInfo._fullLayout,i="modebar-"+n._uid;this.element.setAttribute("id",i),this._uid=i,this.element.className="modebar","hover"===r.displayModeBar&&(this.element.className+=" modebar--hover ease-bg"),"v"===n.modebar.orientation&&(this.element.className+=" vertical",e=e.reverse());var o=n.modebar,s="hover"===r.displayModeBar?".js-plotly-plot .plotly:hover ":"";a.deleteRelatedStyleRule(i),a.addRelatedStyleRule(i,s+"#"+i+" .modebar-group","background-color: "+o.bgcolor),a.addRelatedStyleRule(i,"#"+i+" .modebar-btn .icon path","fill: "+o.color),a.addRelatedStyleRule(i,"#"+i+" .modebar-btn:hover .icon path","fill: "+o.activecolor),a.addRelatedStyleRule(i,"#"+i+" .modebar-btn.active .icon path","fill: "+o.activecolor);var l=!this.hasButtons(e),u=this.hasLogo!==r.displaylogo,c=this.locale!==r.locale;if(this.locale=r.locale,(l||u||c)&&(this.removeAllButtons(),this.updateButtons(e),r.watermark||r.displaylogo)){var f=this.getLogo();r.watermark&&(f.className=f.className+" watermark"),"v"===n.modebar.orientation?this.element.insertBefore(f,this.element.childNodes[0]):this.element.appendChild(f),this.hasLogo=!0}this.updateActiveButton()},c.updateButtons=function(t){var e=this;this.buttons=t,this.buttonElements=[],this.buttonsNames=[],this.buttons.forEach((function(t){var r=e.createGroup();t.forEach((function(t){var n=t.name;if(!n)throw new Error("must provide button 'name' in button config");if(-1!==e.buttonsNames.indexOf(n))throw new Error("button name '"+n+"' is taken");e.buttonsNames.push(n);var i=e.createButton(t);e.buttonElements.push(i),r.appendChild(i)})),e.element.appendChild(r)}))},c.createGroup=function(){var t=document.createElement("div");return t.className="modebar-group",t},c.createButton=function(t){var e=this,r=document.createElement("a");r.setAttribute("rel","tooltip"),r.className="modebar-btn";var i=t.title;void 0===i?i=t.name:"function"==typeof i&&(i=i(this.graphInfo)),(i||0===i)&&r.setAttribute("data-title",i),void 0!==t.attr&&r.setAttribute("data-attr",t.attr);var a=t.val;if(void 0!==a&&("function"==typeof a&&(a=a(this.graphInfo)),r.setAttribute("data-val",a)),"function"!=typeof t.click)throw new Error("must provide button 'click' function in button config");r.addEventListener("click",(function(r){t.click(e.graphInfo,r),e.updateActiveButton(r.currentTarget)})),r.setAttribute("data-toggle",t.toggle||!1),t.toggle&&n.select(r).classed("active",!0);var s=t.icon;return"function"==typeof s?r.appendChild(s()):r.appendChild(this.createIcon(s||o.question)),r.setAttribute("data-gravity",t.gravity||"n"),r},c.createIcon=function(t){var e,r=i(t.height)?Number(t.height):t.ascent-t.descent,n="http://www.w3.org/2000/svg";if(t.path){(e=document.createElementNS(n,"svg")).setAttribute("viewBox",[0,0,t.width,r].join(" ")),e.setAttribute("class","icon");var a=document.createElementNS(n,"path");a.setAttribute("d",t.path),t.transform?a.setAttribute("transform",t.transform):void 0!==t.ascent&&a.setAttribute("transform","matrix(1 0 0 -1 0 "+t.ascent+")"),e.appendChild(a)}return t.svg&&(e=l.parseFromString(t.svg,"application/xml").childNodes[0]),e.setAttribute("height","1em"),e.setAttribute("width","1em"),e},c.updateActiveButton=function(t){var e=this.graphInfo._fullLayout,r=void 0!==t?t.getAttribute("data-attr"):null;this.buttonElements.forEach((function(t){var i=t.getAttribute("data-val")||!0,o=t.getAttribute("data-attr"),s="true"===t.getAttribute("data-toggle"),l=n.select(t);if(s)o===r&&l.classed("active",!l.classed("active"));else{var u=null===o?o:a.nestedProperty(e,o).get();l.classed("active",u===i)}}))},c.hasButtons=function(t){var e=this.buttons;if(!e)return!1;if(t.length!==e.length)return!1;for(var r=0;r<t.length;++r){if(t[r].length!==e[r].length)return!1;for(var n=0;n<t[r].length;n++)if(t[r][n].name!==e[r][n].name)return!1}return!0},c.getLogo=function(){var t=this.createGroup(),e=document.createElement("a");return e.href="https://plotly.com/",e.target="_blank",e.setAttribute("data-title",a._(this.graphInfo,"Produced with Plotly.js")+" (v"+s+")"),e.className="modebar-btn plotlyjsicon modebar-btn--logo",e.appendChild(this.createIcon(o.newplotlylogo)),t.appendChild(e),t},c.removeAllButtons=function(){for(;this.element.firstChild;)this.element.removeChild(this.element.firstChild);this.hasLogo=!1},c.destroy=function(){a.removeElement(this.container.querySelector(".modebar")),a.deleteRelatedStyleRule(this._uid)},t.exports=function(t,e){var r=t._fullLayout,i=new u({graphInfo:t,container:r._modebardiv.node(),buttons:e});return r._privateplot&&n.select(i.element).append("span").classed("badge-private float--left",!0).text("PRIVATE"),i}},26680:function(t,e,r){"use strict";var n=r(25376),i=r(22548),a=(0,r(31780).templatedArray)("button",{visible:{valType:"boolean",dflt:!0,editType:"plot"},step:{valType:"enumerated",values:["month","year","day","hour","minute","second","all"],dflt:"month",editType:"plot"},stepmode:{valType:"enumerated",values:["backward","todate"],dflt:"backward",editType:"plot"},count:{valType:"number",min:0,dflt:1,editType:"plot"},label:{valType:"string",editType:"plot"},editType:"plot"});t.exports={visible:{valType:"boolean",editType:"plot"},buttons:a,x:{valType:"number",min:-2,max:3,editType:"plot"},xanchor:{valType:"enumerated",values:["auto","left","center","right"],dflt:"left",editType:"plot"},y:{valType:"number",min:-2,max:3,editType:"plot"},yanchor:{valType:"enumerated",values:["auto","top","middle","bottom"],dflt:"bottom",editType:"plot"},font:n({editType:"plot"}),bgcolor:{valType:"color",dflt:i.lightLine,editType:"plot"},activecolor:{valType:"color",editType:"plot"},bordercolor:{valType:"color",dflt:i.defaultLine,editType:"plot"},borderwidth:{valType:"number",min:0,dflt:0,editType:"plot"},editType:"plot"}},85984:function(t){"use strict";t.exports={yPad:.02,minButtonWidth:30,rx:3,ry:3,lightAmount:25,darkAmount:10}},22148:function(t,e,r){"use strict";var n=r(3400),i=r(76308),a=r(31780),o=r(51272),s=r(26680),l=r(85984);function u(t,e,r,i){var a=i.calendar;function o(r,i){return n.coerce(t,e,s.buttons,r,i)}if(o("visible")){var l=o("step");"all"!==l&&(!a||"gregorian"===a||"month"!==l&&"year"!==l?o("stepmode"):e.stepmode="backward",o("count")),o("label")}}t.exports=function(t,e,r,c,f){var h=t.rangeselector||{},p=a.newContainer(e,"rangeselector");function d(t,e){return n.coerce(h,p,s,t,e)}if(d("visible",o(h,p,{name:"buttons",handleItemDefaults:u,calendar:f}).length>0)){var v=function(t,e,r){for(var n=r.filter((function(r){return e[r].anchor===t._id})),i=0,a=0;a<n.length;a++){var o=e[n[a]].domain;o&&(i=Math.max(o[1],i))}return[t.domain[0],i+l.yPad]}(e,r,c);d("x",v[0]),d("y",v[1]),n.noneOrAll(t,e,["x","y"]),d("xanchor"),d("yanchor"),n.coerceFont(d,"font",r.font);var g=d("bgcolor");d("activecolor",i.contrast(g,l.lightAmount,l.darkAmount)),d("bordercolor"),d("borderwidth")}}},50216:function(t,e,r){"use strict";var n=r(33428),i=r(24040),a=r(7316),o=r(76308),s=r(43616),l=r(3400),u=l.strTranslate,c=r(72736),f=r(79811),h=r(84284),p=h.LINE_SPACING,d=h.FROM_TL,v=h.FROM_BR,g=r(85984),y=r(48040);function m(t){return t._id}function x(t,e,r){var n=l.ensureSingle(t,"rect","selector-rect",(function(t){t.attr("shape-rendering","crispEdges")}));n.attr({rx:g.rx,ry:g.ry}),n.call(o.stroke,e.bordercolor).call(o.fill,function(t,e){return e._isActive||e._isHovered?t.activecolor:t.bgcolor}(e,r)).style("stroke-width",e.borderwidth+"px")}function b(t,e,r,n){var i,a;l.ensureSingle(t,"text","selector-text",(function(t){t.attr("text-anchor","middle")})).call(s.font,e.font).text((i=r,a=n._fullLayout._meta,i.label?a?l.templateString(i.label,a):i.label:"all"===i.step?"all":i.count+i.step.charAt(0))).call((function(t){c.convertToTspans(t,n)}))}t.exports=function(t){var e=t._fullLayout._infolayer.selectAll(".rangeselector").data(function(t){for(var e=f.list(t,"x",!0),r=[],n=0;n<e.length;n++){var i=e[n];i.rangeselector&&i.rangeselector.visible&&r.push(i)}return r}(t),m);e.enter().append("g").classed("rangeselector",!0),e.exit().remove(),e.style({cursor:"pointer","pointer-events":"all"}),e.each((function(e){var r=n.select(this),o=e,f=o.rangeselector,h=r.selectAll("g.button").data(l.filterVisible(f.buttons));h.enter().append("g").classed("button",!0),h.exit().remove(),h.each((function(e){var r=n.select(this),a=y(o,e);e._isActive=function(t,e,r){if("all"===e.step)return!0===t.autorange;var n=Object.keys(r);return t.range[0]===r[n[0]]&&t.range[1]===r[n[1]]}(o,e,a),r.call(x,f,e),r.call(b,f,e,t),r.on("click",(function(){t._dragged||i.call("_guiRelayout",t,a)})),r.on("mouseover",(function(){e._isHovered=!0,r.call(x,f,e)})),r.on("mouseout",(function(){e._isHovered=!1,r.call(x,f,e)}))})),function(t,e,r,i,o){var f=0,h=0,y=r.borderwidth;e.each((function(){var t=n.select(this).select(".selector-text"),e=r.font.size*p,i=Math.max(e*c.lineCount(t),16)+3;h=Math.max(h,i)})),e.each((function(){var t=n.select(this),e=t.select(".selector-rect"),i=t.select(".selector-text"),a=i.node()&&s.bBox(i.node()).width,o=r.font.size*p,l=c.lineCount(i),d=Math.max(a+10,g.minButtonWidth);t.attr("transform",u(y+f,y)),e.attr({x:0,y:0,width:d,height:h}),c.positionText(i,d/2,h/2-(l-1)*o/2+3),f+=d+5}));var m=t._fullLayout._size,x=m.l+m.w*r.x,b=m.t+m.h*(1-r.y),_="left";l.isRightAnchor(r)&&(x-=f,_="right"),l.isCenterAnchor(r)&&(x-=f/2,_="center");var w="top";l.isBottomAnchor(r)&&(b-=h,w="bottom"),l.isMiddleAnchor(r)&&(b-=h/2,w="middle"),f=Math.ceil(f),h=Math.ceil(h),x=Math.round(x),b=Math.round(b),a.autoMargin(t,i+"-range-selector",{x:r.x,y:r.y,l:f*d[_],r:f*v[_],b:h*v[w],t:h*d[w]}),o.attr("transform",u(x,b))}(t,h,f,o._name,r)}))}},48040:function(t,e,r){"use strict";var n=r(73220),i=r(3400).titleCase;t.exports=function(t,e){var r=t._name,a={};if("all"===e.step)a[r+".autorange"]=!0;else{var o=function(t,e){var r,a=t.range,o=new Date(t.r2l(a[1])),s=e.step,l=n["utc"+i(s)],u=e.count;switch(e.stepmode){case"backward":r=t.l2r(+l.offset(o,-u));break;case"todate":var c=l.offset(o,-u);r=t.l2r(+l.ceil(c))}return[r,a[1]]}(t,e);a[r+".range[0]"]=o[0],a[r+".range[1]"]=o[1]}return a}},41152:function(t,e,r){"use strict";t.exports={moduleType:"component",name:"rangeselector",schema:{subplots:{xaxis:{rangeselector:r(26680)}}},layoutAttributes:r(26680),handleDefaults:r(22148),draw:r(50216)}},11200:function(t,e,r){"use strict";var n=r(22548);t.exports={bgcolor:{valType:"color",dflt:n.background,editType:"plot"},bordercolor:{valType:"color",dflt:n.defaultLine,editType:"plot"},borderwidth:{valType:"integer",dflt:0,min:0,editType:"plot"},autorange:{valType:"boolean",dflt:!0,editType:"calc",impliedEdits:{"range[0]":void 0,"range[1]":void 0}},range:{valType:"info_array",items:[{valType:"any",editType:"calc",impliedEdits:{"^autorange":!1}},{valType:"any",editType:"calc",impliedEdits:{"^autorange":!1}}],editType:"calc",impliedEdits:{autorange:!1}},thickness:{valType:"number",dflt:.15,min:0,max:1,editType:"plot"},visible:{valType:"boolean",dflt:!0,editType:"calc"},editType:"calc"}},26652:function(t,e,r){"use strict";var n=r(79811).list,i=r(19280).getAutoRange,a=r(74636);t.exports=function(t){for(var e=n(t,"x",!0),r=0;r<e.length;r++){var o=e[r],s=o[a.name];s&&s.visible&&s.autorange&&(s._input.autorange=!0,s._input.range=s.range=i(t,o))}}},74636:function(t){"use strict";t.exports={name:"rangeslider",containerClassName:"rangeslider-container",bgClassName:"rangeslider-bg",rangePlotClassName:"rangeslider-rangeplot",maskMinClassName:"rangeslider-mask-min",maskMaxClassName:"rangeslider-mask-max",slideBoxClassName:"rangeslider-slidebox",grabberMinClassName:"rangeslider-grabber-min",grabAreaMinClassName:"rangeslider-grabarea-min",handleMinClassName:"rangeslider-handle-min",grabberMaxClassName:"rangeslider-grabber-max",grabAreaMaxClassName:"rangeslider-grabarea-max",handleMaxClassName:"rangeslider-handle-max",maskMinOppAxisClassName:"rangeslider-mask-min-opp-axis",maskMaxOppAxisClassName:"rangeslider-mask-max-opp-axis",maskColor:"rgba(0,0,0,0.4)",maskOppAxisColor:"rgba(0,0,0,0.2)",slideBoxFill:"transparent",slideBoxCursor:"ew-resize",grabAreaFill:"transparent",grabAreaCursor:"col-resize",grabAreaWidth:10,handleWidth:4,handleRadius:1,handleStrokeWidth:1,extraPad:15}},94040:function(t,e,r){"use strict";var n=r(3400),i=r(31780),a=r(79811),o=r(11200),s=r(10936);t.exports=function(t,e,r){var l=t[r],u=e[r];if(l.rangeslider||e._requestRangeslider[u._id]){n.isPlainObject(l.rangeslider)||(l.rangeslider={});var c,f,h=l.rangeslider,p=i.newContainer(u,"rangeslider");if(_("visible")){_("bgcolor",e.plot_bgcolor),_("bordercolor"),_("borderwidth"),_("thickness"),_("autorange",!u.isValidRange(h.range)),_("range");var d=e._subplots;if(d)for(var v=d.cartesian.filter((function(t){return t.substr(0,t.indexOf("y"))===a.name2id(r)})).map((function(t){return t.substr(t.indexOf("y"),t.length)})),g=n.simpleMap(v,a.id2name),y=0;y<g.length;y++){var m=g[y];c=h[m]||{},f=i.newContainer(p,m,"yaxis");var x,b=e[m];c.range&&b.isValidRange(c.range)&&(x="fixed"),"match"!==w("rangemode",x)&&w("range",b.range.slice())}p._input=h}}function _(t,e){return n.coerce(h,p,o,t,e)}function w(t,e){return n.coerce(c,f,s,t,e)}}},20060:function(t,e,r){"use strict";var n=r(33428),i=r(24040),a=r(7316),o=r(3400),s=o.strTranslate,l=r(43616),u=r(76308),c=r(81668),f=r(57952),h=r(79811),p=r(86476),d=r(93972),v=r(74636);function g(t){return"number"==typeof t.clientX?t.clientX:t.touches&&t.touches.length>0?t.touches[0].clientX:0}function y(t,e,r,n){var i=o.ensureSingle(t,"rect",v.bgClassName,(function(t){t.attr({x:0,y:0,"shape-rendering":"crispEdges"})})),a=n.borderwidth%2==0?n.borderwidth:n.borderwidth-1,c=-n._offsetShift,f=l.crispRound(e,n.borderwidth);i.attr({width:n._width+a,height:n._height+a,transform:s(c,c),"stroke-width":f}).call(u.stroke,n.bordercolor).call(u.fill,n.bgcolor)}function m(t,e,r,n){var i=e._fullLayout;o.ensureSingleById(i._topdefs,"clipPath",n._clipId,(function(t){t.append("rect").attr({x:0,y:0})})).select("rect").attr({width:n._width,height:n._height})}function x(t,e,r,i){var s,u=e.calcdata,c=t.selectAll("g."+v.rangePlotClassName).data(r._subplotsWith,o.identity);c.enter().append("g").attr("class",(function(t){return v.rangePlotClassName+" "+t})).call(l.setClipUrl,i._clipId,e),c.order(),c.exit().remove(),c.each((function(t,o){var l=n.select(this),c=0===o,p=h.getFromId(e,t,"y"),d=p._name,v=i[d],g={data:[],layout:{xaxis:{type:r.type,domain:[0,1],range:i.range.slice(),calendar:r.calendar},width:i._width,height:i._height,margin:{t:0,b:0,l:0,r:0}},_context:e._context};r.rangebreaks&&(g.layout.xaxis.rangebreaks=r.rangebreaks),g.layout[d]={type:p.type,domain:[0,1],range:"match"!==v.rangemode?v.range.slice():p.range.slice(),calendar:p.calendar},p.rangebreaks&&(g.layout[d].rangebreaks=p.rangebreaks),a.supplyDefaults(g);var y=g._fullLayout.xaxis,m=g._fullLayout[d];y.clearCalc(),y.setScale(),m.clearCalc(),m.setScale();var x={id:t,plotgroup:l,xaxis:y,yaxis:m,isRangePlot:!0};c?s=x:(x.mainplot="xy",x.mainplotinfo=s),f.rangePlot(e,x,function(t,e){for(var r=[],n=0;n<t.length;n++){var i=t[n],a=i[0].trace;a.xaxis+a.yaxis===e&&r.push(i)}return r}(u,t))}))}function b(t,e,r,n,i){o.ensureSingle(t,"rect",v.maskMinClassName,(function(t){t.attr({x:0,y:0,"shape-rendering":"crispEdges"})})).attr("height",n._height).call(u.fill,v.maskColor),o.ensureSingle(t,"rect",v.maskMaxClassName,(function(t){t.attr({y:0,"shape-rendering":"crispEdges"})})).attr("height",n._height).call(u.fill,v.maskColor),"match"!==i.rangemode&&(o.ensureSingle(t,"rect",v.maskMinOppAxisClassName,(function(t){t.attr({y:0,"shape-rendering":"crispEdges"})})).attr("width",n._width).call(u.fill,v.maskOppAxisColor),o.ensureSingle(t,"rect",v.maskMaxOppAxisClassName,(function(t){t.attr({y:0,"shape-rendering":"crispEdges"})})).attr("width",n._width).style("border-top",v.maskOppBorder).call(u.fill,v.maskOppAxisColor))}function _(t,e,r,n){e._context.staticPlot||o.ensureSingle(t,"rect",v.slideBoxClassName,(function(t){t.attr({y:0,cursor:v.slideBoxCursor,"shape-rendering":"crispEdges"})})).attr({height:n._height,fill:v.slideBoxFill})}function w(t,e,r,n){var i=o.ensureSingle(t,"g",v.grabberMinClassName),a=o.ensureSingle(t,"g",v.grabberMaxClassName),s={x:0,width:v.handleWidth,rx:v.handleRadius,fill:u.background,stroke:u.defaultLine,"stroke-width":v.handleStrokeWidth,"shape-rendering":"crispEdges"},l={y:Math.round(n._height/4),height:Math.round(n._height/2)};o.ensureSingle(i,"rect",v.handleMinClassName,(function(t){t.attr(s)})).attr(l),o.ensureSingle(a,"rect",v.handleMaxClassName,(function(t){t.attr(s)})).attr(l);var c={width:v.grabAreaWidth,x:0,y:0,fill:v.grabAreaFill,cursor:e._context.staticPlot?void 0:v.grabAreaCursor};o.ensureSingle(i,"rect",v.grabAreaMinClassName,(function(t){t.attr(c)})).attr("height",n._height),o.ensureSingle(a,"rect",v.grabAreaMaxClassName,(function(t){t.attr(c)})).attr("height",n._height)}t.exports=function(t){for(var e=t._fullLayout,r=e._rangeSliderData,a=0;a<r.length;a++){var l=r[a][v.name];l._clipId=l._id+"-"+e._uid}var u=e._infolayer.selectAll("g."+v.containerClassName).data(r,(function(t){return t._name}));u.exit().each((function(t){var r=t[v.name];e._topdefs.select("#"+r._clipId).remove()})).remove(),0!==r.length&&(u.enter().append("g").classed(v.containerClassName,!0).attr("pointer-events","all"),u.each((function(r){var a=n.select(this),l=r[v.name],u=e[h.id2name(r.anchor)],f=l[h.id2name(r.anchor)];if(l.range){var T,k=o.simpleMap(l.range,r.r2l),A=o.simpleMap(r.range,r.r2l);T=A[0]<A[1]?[Math.min(k[0],A[0]),Math.max(k[1],A[1])]:[Math.max(k[0],A[0]),Math.min(k[1],A[1])],l.range=l._input.range=o.simpleMap(T,r.l2r)}r.cleanRange("rangeslider.range");var M=e._size,S=r.domain;l._width=M.w*(S[1]-S[0]);var E=Math.round(M.l+M.w*S[0]),L=Math.round(M.t+M.h*(1-r._counterDomainMin)+("bottom"===r.side?r._depth:0)+l._offsetShift+v.extraPad);a.attr("transform",s(E,L)),l._rl=o.simpleMap(l.range,r.r2l);var C=l._rl[0],P=l._rl[1],O=P-C;if(l.p2d=function(t){return t/l._width*O+C},l.d2p=function(t){return(t-C)/O*l._width},r.rangebreaks){var I=r.locateBreaks(C,P);if(I.length){var z,D,R=0;for(z=0;z<I.length;z++)R+=(D=I[z]).max-D.min;var F=l._width/(P-C-R),B=[-F*C];for(z=0;z<I.length;z++)D=I[z],B.push(B[B.length-1]-F*(D.max-D.min));for(l.d2p=function(t){for(var e=B[0],r=0;r<I.length;r++){var n=I[r];if(t>=n.max)e=B[r+1];else if(t<n.min)break}return e+F*t},z=0;z<I.length;z++)(D=I[z]).pmin=l.d2p(D.min),D.pmax=l.d2p(D.max);l.p2d=function(t){for(var e=B[0],r=0;r<I.length;r++){var n=I[r];if(t>=n.pmax)e=B[r+1];else if(t<n.pmin)break}return(t-e)/F}}}if("match"!==f.rangemode){var N=u.r2l(f.range[0]),j=u.r2l(f.range[1])-N;l.d2pOppAxis=function(t){return(t-N)/j*l._height}}a.call(y,t,r,l).call(m,t,r,l).call(x,t,r,l).call(b,t,r,l,f).call(_,t,r,l).call(w,t,r,l),function(t,e,r,a){if(!e._context.staticPlot){var s=t.select("rect."+v.slideBoxClassName).node(),l=t.select("rect."+v.grabAreaMinClassName).node(),u=t.select("rect."+v.grabAreaMaxClassName).node();t.on("mousedown",c),t.on("touchstart",c)}function c(){var c=n.event,f=c.target,h=g(c),v=h-t.node().getBoundingClientRect().left,y=a.d2p(r._rl[0]),m=a.d2p(r._rl[1]),x=p.coverSlip();function b(t){var c,p,b,_=+g(t)-h;switch(f){case s:if(b="ew-resize",y+_>r._length||m+_<0)return;c=y+_,p=m+_;break;case l:if(b="col-resize",y+_>r._length)return;c=y+_,p=m;break;case u:if(b="col-resize",m+_<0)return;c=y,p=m+_;break;default:b="ew-resize",c=v,p=v+_}if(p<c){var w=p;p=c,c=w}a._pixelMin=c,a._pixelMax=p,d(n.select(x),b),function(t,e,r,n){function a(t){return r.l2r(o.constrain(t,n._rl[0],n._rl[1]))}var s=a(n.p2d(n._pixelMin)),l=a(n.p2d(n._pixelMax));window.requestAnimationFrame((function(){i.call("_guiRelayout",e,r._name+".range",[s,l])}))}(0,e,r,a)}function _(){x.removeEventListener("mousemove",b),x.removeEventListener("mouseup",_),this.removeEventListener("touchmove",b),this.removeEventListener("touchend",_),o.removeElement(x)}this.addEventListener("touchmove",b),this.addEventListener("touchend",_),x.addEventListener("mousemove",b),x.addEventListener("mouseup",_)}}(a,t,r,l),function(t,e,r,n,i,a){var l=v.handleWidth/2;function u(t){return o.constrain(t,0,n._width)}function c(t){return o.constrain(t,0,n._height)}function f(t){return o.constrain(t,-l,n._width+l)}var h=u(n.d2p(r._rl[0])),p=u(n.d2p(r._rl[1]));if(t.select("rect."+v.slideBoxClassName).attr("x",h).attr("width",p-h),t.select("rect."+v.maskMinClassName).attr("width",h),t.select("rect."+v.maskMaxClassName).attr("x",p).attr("width",n._width-p),"match"!==a.rangemode){var d=n._height-c(n.d2pOppAxis(i._rl[1])),g=n._height-c(n.d2pOppAxis(i._rl[0]));t.select("rect."+v.maskMinOppAxisClassName).attr("x",h).attr("height",d).attr("width",p-h),t.select("rect."+v.maskMaxOppAxisClassName).attr("x",h).attr("y",g).attr("height",n._height-g).attr("width",p-h),t.select("rect."+v.slideBoxClassName).attr("y",d).attr("height",g-d)}var y=.5,m=Math.round(f(h-l))-y,x=Math.round(f(p-l))+y;t.select("g."+v.grabberMinClassName).attr("transform",s(m,y)),t.select("g."+v.grabberMaxClassName).attr("transform",s(x,y))}(a,0,r,l,u,f),"bottom"===r.side&&c.draw(t,r._id+"title",{propContainer:r,propName:r._name+".title",placeholder:e._dfltTitle.x,attributes:{x:r._offset+r._length/2,y:L+l._height+l._offsetShift+10+1.5*r.title.font.size,"text-anchor":"middle"}})})))}},97944:function(t,e,r){"use strict";var n=r(79811),i=r(72736),a=r(74636),o=r(84284).LINE_SPACING,s=a.name;function l(t){var e=t&&t[s];return e&&e.visible}e.isVisible=l,e.makeData=function(t){var e=n.list({_fullLayout:t},"x",!0),r=t.margin,i=[];if(!t._has("gl2d"))for(var a=0;a<e.length;a++){var o=e[a];if(l(o)){i.push(o);var u=o[s];u._id=s+o._id,u._height=(t.height-r.b-r.t)*u.thickness,u._offsetShift=Math.floor(u.borderwidth/2)}}t._rangeSliderData=i},e.autoMarginOpts=function(t,e){var r=t._fullLayout,n=e[s],l=e._id.charAt(0),u=0,c=0;return"bottom"===e.side&&(u=e._depth,e.title.text!==r._dfltTitle[l]&&(c=1.5*e.title.font.size+10+n._offsetShift,c+=(e.title.text.match(i.BR_TAG_ALL)||[]).length*e.title.font.size*o)),{x:0,y:e._counterDomainMin,l:0,r:0,t:0,b:n._height+u+Math.max(r.margin.b,c),pad:a.extraPad+2*n._offsetShift}}},49692:function(t,e,r){"use strict";var n=r(3400),i=r(11200),a=r(10936),o=r(97944);t.exports={moduleType:"component",name:"rangeslider",schema:{subplots:{xaxis:{rangeslider:n.extendFlat({},i,{yaxis:a})}}},layoutAttributes:r(11200),handleDefaults:r(94040),calcAutorange:r(26652),draw:r(20060),isVisible:o.isVisible,makeData:o.makeData,autoMarginOpts:o.autoMarginOpts}},10936:function(t){"use strict";t.exports={_isSubplotObj:!0,rangemode:{valType:"enumerated",values:["auto","fixed","match"],dflt:"match",editType:"calc"},range:{valType:"info_array",items:[{valType:"any",editType:"plot"},{valType:"any",editType:"plot"}],editType:"plot"},editType:"calc"}},93956:function(t,e,r){"use strict";var n=r(13916),i=r(52904).line,a=r(98192).u,o=r(92880).extendFlat,s=r(67824).overrideAll,l=r(31780).templatedArray;r(36208),t.exports=s(l("selection",{type:{valType:"enumerated",values:["rect","path"]},xref:o({},n.xref,{}),yref:o({},n.yref,{}),x0:{valType:"any"},x1:{valType:"any"},y0:{valType:"any"},y1:{valType:"any"},path:{valType:"string",editType:"arraydraw"},opacity:{valType:"number",min:0,max:1,dflt:.7,editType:"arraydraw"},line:{color:i.color,width:o({},i.width,{min:1,dflt:1}),dash:o({},a,{dflt:"dot"})}}),"arraydraw","from-root")},83280:function(t){"use strict";t.exports={BENDPX:1.5,MINSELECT:12,SELECTDELAY:100,SELECTID:"-select"}},74224:function(t,e,r){"use strict";var n=r(3400),i=r(54460),a=r(51272),o=r(93956),s=r(65152);function l(t,e,r){function a(r,i){return n.coerce(t,e,o,r,i)}var l=a("path"),u="path"!==a("type",l?"path":"rect");u&&delete e.path,a("opacity"),a("line.color"),a("line.width"),a("line.dash");for(var c=["x","y"],f=0;f<2;f++){var h,p,d,v=c[f],g={_fullLayout:r},y=i.coerceRef(t,e,g,v);if((h=i.getFromId(g,y))._selectionIndices.push(e._index),d=s.rangeToShapePosition(h),p=s.shapePositionToRange(h),u){var m=v+"0",x=v+"1",b=t[m],_=t[x];t[m]=p(t[m],!0),t[x]=p(t[x],!0),i.coercePosition(e,g,a,y,m),i.coercePosition(e,g,a,y,x);var w=e[m],T=e[x];void 0!==w&&void 0!==T&&(e[m]=d(w),e[x]=d(T),t[m]=b,t[x]=_)}}u&&n.noneOrAll(t,e,["x0","x1","y0","y1"])}t.exports=function(t,e){a(t,e,{name:"selections",handleItemDefaults:l});for(var r=e.selections,n=0;n<r.length;n++){var i=r[n];i&&void 0===i.path&&(void 0!==i.x0&&void 0!==i.x1&&void 0!==i.y0&&void 0!==i.y1||(e.selections[n]=null))}}},23640:function(t,e,r){"use strict";var n=r(9856).readPaths,i=r(55496),a=r(1936).clearOutlineControllers,o=r(76308),s=r(43616),l=r(31780).arrayEditor,u=r(65152),c=u.getPathString;function f(t){var e=t._fullLayout;for(var r in a(t),e._selectionLayer.selectAll("path").remove(),e._plots){var n=e._plots[r].selectionLayer;n&&n.selectAll("path").remove()}for(var i=0;i<e.selections.length;i++)p(t,i)}function h(t){return t._context.editSelection}function p(t,e){t._fullLayout._paperdiv.selectAll('.selectionlayer [data-index="'+e+'"]').remove();var r=u.makeSelectionsOptionsAndPlotinfo(t,e),a=r.options,p=r.plotinfo;a._input&&function(r){var u=c(t,a),g={"data-index":e,"fill-rule":"evenodd",d:u},y=a.opacity,m="rgba(0,0,0,0)",x=a.line.color||o.contrast(t._fullLayout.plot_bgcolor),b=a.line.width,_=a.line.dash;b||(b=5,_="solid");var w=h(t)&&t._fullLayout._activeSelectionIndex===e;w&&(m=t._fullLayout.activeselection.fillcolor,y=t._fullLayout.activeselection.opacity);for(var T=[],k=1;k>=0;k--){var A=r.append("path").attr(g).style("opacity",k?.1:y).call(o.stroke,x).call(o.fill,m).call(s.dashLine,k?"solid":_,k?4+b:b);if(d(A,t,a),w){var M=l(t.layout,"selections",a);A.style({cursor:"move"});var S={element:A.node(),plotinfo:p,gd:t,editHelpers:M,isActiveSelection:!0},E=n(u,t);i(E,A,S)}else A.style("pointer-events",k?"all":"none");T[k]=A}var L=T[0];T[1].node().addEventListener("click",(function(){return function(t,e){if(h(t)){var r=+e.node().getAttribute("data-index");if(r>=0){if(r===t._fullLayout._activeSelectionIndex)return void v(t);t._fullLayout._activeSelectionIndex=r,t._fullLayout._deactivateSelection=v,f(t)}}}(t,L)}))}(t._fullLayout._selectionLayer)}function d(t,e,r){var n=r.xref+r.yref;s.setClipUrl(t,"clip"+e._fullLayout._uid+n,e)}function v(t){h(t)&&t._fullLayout._activeSelectionIndex>=0&&(a(t),delete t._fullLayout._activeSelectionIndex,f(t))}t.exports={draw:f,drawOne:p,activateLastSelection:function(t){if(h(t)){var e=t._fullLayout.selections.length-1;t._fullLayout._activeSelectionIndex=e,t._fullLayout._deactivateSelection=v,f(t)}}}},34200:function(t,e,r){"use strict";var n=r(98192).u,i=r(92880).extendFlat;t.exports={newselection:{mode:{valType:"enumerated",values:["immediate","gradual"],dflt:"immediate",editType:"none"},line:{color:{valType:"color",editType:"none"},width:{valType:"number",min:1,dflt:1,editType:"none"},dash:i({},n,{dflt:"dot",editType:"none"}),editType:"none"},editType:"none"},activeselection:{fillcolor:{valType:"color",dflt:"rgba(0,0,0,0)",editType:"none"},opacity:{valType:"number",min:0,max:1,dflt:.5,editType:"none"},editType:"none"}}},81004:function(t){"use strict";t.exports=function(t,e,r){r("newselection.mode"),r("newselection.line.width")&&(r("newselection.line.color"),r("newselection.line.dash")),r("activeselection.fillcolor"),r("activeselection.opacity")}},5968:function(t,e,r){"use strict";var n=r(72760).selectMode,i=r(1936).clearOutline,a=r(9856),o=a.readPaths,s=a.writePaths,l=a.fixDatesForPaths;t.exports=function(t,e){if(t.length){var r=t[0][0];if(r){var a=r.getAttribute("d"),u=e.gd,c=u._fullLayout.newselection,f=e.plotinfo,h=f.xaxis,p=f.yaxis,d=e.isActiveSelection,v=e.dragmode,g=(u.layout||{}).selections||[];if(!n(v)&&void 0!==d){var y=u._fullLayout._activeSelectionIndex;if(y<g.length)switch(u._fullLayout.selections[y].type){case"rect":v="select";break;case"path":v="lasso"}}var m,x=o(a,u,f,d),b={xref:h._id,yref:p._id,opacity:c.opacity,line:{color:c.line.color,width:c.line.width,dash:c.line.dash}};1===x.length&&(m=x[0]),m&&5===m.length&&"select"===v?(b.type="rect",b.x0=m[0][1],b.y0=m[0][2],b.x1=m[2][1],b.y1=m[2][2]):(b.type="path",h&&p&&l(x,h,p),b.path=s(x),m=null),i(u);for(var _=e.editHelpers,w=(_||{}).modifyItem,T=[],k=0;k<g.length;k++){var A=u._fullLayout.selections[k];if(A){if(T[k]=A._input,void 0!==d&&k===u._fullLayout._activeSelectionIndex){var M=b;switch(A.type){case"rect":w("x0",M.x0),w("x1",M.x1),w("y0",M.y0),w("y1",M.y1);break;case"path":w("path",M.path)}}}else T[k]=A}return void 0===d?(T.push(b),T):_?_.getUpdateObj():{}}}}},5840:function(t,e,r){"use strict";var n=r(3400).strTranslate;function i(t,e){switch(t.type){case"log":return t.p2d(e);case"date":return t.p2r(e,0,t.calendar);default:return t.p2r(e)}}t.exports={p2r:i,r2p:function(t,e){switch(t.type){case"log":return t.d2p(e);case"date":return t.r2p(e,0,t.calendar);default:return t.r2p(e)}},axValue:function(t){var e="y"===t._id.charAt(0)?1:0;return function(r){return i(t,r[e])}},getTransform:function(t){return n(t.xaxis._offset,t.yaxis._offset)}}},22676:function(t,e,r){"use strict";var n=r(23640),i=r(43156);t.exports={moduleType:"component",name:"selections",layoutAttributes:r(93956),supplyLayoutDefaults:r(74224),supplyDrawNewSelectionDefaults:r(81004),includeBasePlot:r(36632)("selections"),draw:n.draw,drawOne:n.drawOne,reselect:i.reselect,prepSelect:i.prepSelect,clearOutline:i.clearOutline,clearSelectionsCache:i.clearSelectionsCache,selectOnClick:i.selectOnClick}},43156:function(t,e,r){"use strict";var n=r(14756),i=r(61456),a=r(24040),o=r(43616).dashStyle,s=r(76308),l=r(93024),u=r(10624).makeEventData,c=r(72760),f=c.freeMode,h=c.rectMode,p=c.drawMode,d=c.openMode,v=c.selectMode,g=r(65152),y=r(85448),m=r(55496),x=r(1936).clearOutline,b=r(9856),_=b.handleEllipse,w=b.readPaths,T=r(93940).newShapes,k=r(5968),A=r(23640).activateLastSelection,M=r(3400),S=M.sorterAsc,E=r(92065),L=r(91200),C=r(79811).getFromId,P=r(73696),O=r(39172).redrawReglTraces,I=r(83280),z=I.MINSELECT,D=E.filter,R=E.tester,F=r(5840),B=F.p2r,N=F.axValue,j=F.getTransform;function U(t){return void 0!==t.subplot}function V(t,e,r,n,i,a,o){var s,l,u,c,f,h,p,v,g,y=e._hoverdata,x=e._fullLayout.clickmode.indexOf("event")>-1,b=[];if(function(t){return t&&Array.isArray(t)&&!0!==t[0].hoverOnBox}(y)){W(t,e,a);var _=function(t,e){var r,n,i=t[0],a=-1,o=[];for(n=0;n<e.length;n++)if(r=e[n],i.fullData._expandedIndex===r.cd[0].trace._expandedIndex){if(!0===i.hoverOnBox)break;void 0!==i.pointNumber?a=i.pointNumber:void 0!==i.binNumber&&(a=i.binNumber,o=i.pointNumbers);break}return{pointNumber:a,pointNumbers:o,searchInfo:r}}(y,s=Z(e,r,n,i));if(_.pointNumbers.length>0?function(t,e){var r,n,i,a=[];for(i=0;i<t.length;i++)(r=t[i]).cd[0].trace.selectedpoints&&r.cd[0].trace.selectedpoints.length>0&&a.push(r);if(1===a.length&&a[0]===e.searchInfo&&(n=e.searchInfo.cd[0].trace).selectedpoints.length===e.pointNumbers.length){for(i=0;i<e.pointNumbers.length;i++)if(n.selectedpoints.indexOf(e.pointNumbers[i])<0)return!1;return!0}return!1}(s,_):function(t){var e,r,n=0;for(r=0;r<t.length;r++)if((e=t[r].cd[0].trace).selectedpoints){if(e.selectedpoints.length>1)return!1;if((n+=e.selectedpoints.length)>1)return!1}return 1===n}(s)&&(h=J(_))){for(o&&o.remove(),g=0;g<s.length;g++)(l=s[g])._module.selectPoints(l,!1);$(e,s),Y(a),x&&ht(e)}else{for(p=t.shiftKey&&(void 0!==h?h:J(_)),u=function(t,e,r){return{pointNumber:t,searchInfo:e,subtract:!!r}}(_.pointNumber,_.searchInfo,p),c=G(a.selectionDefs.concat([u])),g=0;g<s.length;g++)if(f=tt(s[g]._module.selectPoints(s[g],c),s[g]),b.length)for(var w=0;w<f.length;w++)b.push(f[w]);else b=f;if($(e,s,v={points:b}),u&&a&&a.selectionDefs.push(u),o){var T=a.mergedPolygons,k=d(a.dragmode);m(et(T,k),o,a)}x&&ft(e,v)}}}function q(t){return"pointNumber"in t&&"searchInfo"in t}function H(t){return{xmin:0,xmax:0,ymin:0,ymax:0,pts:[],contains:function(e,r,n,i){var a=t.searchInfo.cd[0].trace._expandedIndex;return i.cd[0].trace._expandedIndex===a&&n===t.pointNumber},isRect:!1,degenerate:!1,subtract:!!t.subtract}}function G(t){if(t.length){for(var e=[],r=q(t[0])?0:t[0][0][0],n=r,i=q(t[0])?0:t[0][0][1],a=i,o=0;o<t.length;o++)if(q(t[o]))e.push(H(t[o]));else{var s=R(t[o]);s.subtract=!!t[o].subtract,e.push(s),r=Math.min(r,s.xmin),n=Math.max(n,s.xmax),i=Math.min(i,s.ymin),a=Math.max(a,s.ymax)}return{xmin:r,xmax:n,ymin:i,ymax:a,pts:[],contains:function(t,r,n,i){for(var a=!1,o=0;o<e.length;o++)e[o].contains(t,r,n,i)&&(a=!e[o].subtract);return a},isRect:!1,degenerate:!1}}}function W(t,e,r){var n=e._fullLayout,i=r.plotinfo,a=r.dragmode,o=n._lastSelectedSubplot&&n._lastSelectedSubplot===i.id,s=(t.shiftKey||t.altKey)&&!(p(a)&&d(a));o&&s&&i.selection&&i.selection.selectionDefs&&!r.selectionDefs?(r.selectionDefs=i.selection.selectionDefs,r.mergedPolygons=i.selection.mergedPolygons):s&&i.selection||Y(r),o||(x(e),n._lastSelectedSubplot=i.id)}function Y(t,e){var r=t.dragmode,n=t.plotinfo,i=t.gd;(function(t){return t._fullLayout._activeShapeIndex>=0})(i)&&i._fullLayout._deactivateShape(i),function(t){return t._fullLayout._activeSelectionIndex>=0}(i)&&i._fullLayout._deactivateSelection(i);var o=i._fullLayout._zoomlayer,s=p(r),l=v(r);if(s||l){var u,c,f=o.selectAll(".select-outline-"+n.id);f&&i._fullLayout._outlining&&(s&&(u=T(f,t)),u&&a.call("_guiRelayout",i,{shapes:u}),l&&!U(t)&&(c=k(f,t)),c&&(i._fullLayout._noEmitSelectedAtStart=!0,a.call("_guiRelayout",i,{selections:c}).then((function(){e&&A(i)}))),i._fullLayout._outlining=!1)}n.selection={},n.selection.selectionDefs=t.selectionDefs=[],n.selection.mergedPolygons=t.mergedPolygons=[]}function X(t){return t._id}function Z(t,e,r,n){if(!t.calcdata)return[];var i,a,o,s=[],l=e.map(X),u=r.map(X);for(o=0;o<t.calcdata.length;o++)if(!0===(a=(i=t.calcdata[o])[0].trace).visible&&a._module&&a._module.selectPoints)if(!U({subplot:n})||a.subplot!==n&&a.geo!==n)if("splom"===a.type){if(a._xaxes[l[0]]&&a._yaxes[u[0]]){var c=K(a._module,i,e[0],r[0]);c.scene=t._fullLayout._splomScenes[a.uid],s.push(c)}}else if("sankey"===a.type){var f=K(a._module,i,e[0],r[0]);s.push(f)}else{if(!(-1!==l.indexOf(a.xaxis)||a._xA&&a._xA.overlaying))continue;if(!(-1!==u.indexOf(a.yaxis)||a._yA&&a._yA.overlaying))continue;s.push(K(a._module,i,C(t,a.xaxis),C(t,a.yaxis)))}else s.push(K(a._module,i,e[0],r[0]));return s}function K(t,e,r,n){return{_module:t,cd:e,xaxis:r,yaxis:n}}function J(t){var e=t.searchInfo.cd[0].trace,r=t.pointNumber,n=t.pointNumbers,i=n.length>0?n[0]:r;return!!e.selectedpoints&&e.selectedpoints.indexOf(i)>-1}function $(t,e,r){var n,i;for(n=0;n<e.length;n++){var o=e[n].cd[0].trace._fullInput,s=t._fullLayout._tracePreGUI[o.uid]||{};void 0===s.selectedpoints&&(s.selectedpoints=o._input.selectedpoints||null)}if(r){var l=r.points||[];for(n=0;n<e.length;n++)(i=e[n].cd[0].trace)._input.selectedpoints=i._fullInput.selectedpoints=[],i._fullInput!==i&&(i.selectedpoints=[]);for(var u=0;u<l.length;u++){var c=l[u],f=c.data,h=c.fullData,p=c.pointIndex,d=c.pointIndices;d?([].push.apply(f.selectedpoints,d),i._fullInput!==i&&[].push.apply(h.selectedpoints,d)):(f.selectedpoints.push(p),i._fullInput!==i&&h.selectedpoints.push(p))}}else for(n=0;n<e.length;n++)delete(i=e[n].cd[0].trace).selectedpoints,delete i._input.selectedpoints,i._fullInput!==i&&delete i._fullInput.selectedpoints;!function(t,e){for(var r=!1,n=0;n<e.length;n++){var i=e[n],o=i.cd;a.traceIs(o[0].trace,"regl")&&(r=!0);var s=i._module,l=s.styleOnSelect||s.style;l&&(l(t,o,o[0].node3),o[0].nodeRangePlot3&&l(t,o,o[0].nodeRangePlot3))}r&&(P(t),O(t))}(t,e)}function Q(t,e,r){for(var i=(r?n.difference:n.union)({regions:t},{regions:[e]}).regions.reverse(),a=0;a<i.length;a++){var o=i[a];o.subtract=st(o,i.slice(0,a))}return i}function tt(t,e){if(Array.isArray(t))for(var r=e.cd,n=e.cd[0].trace,i=0;i<t.length;i++)t[i]=u(t[i],n,r);return t}function et(t,e){for(var r=[],n=0;n<t.length;n++){r[n]=[];for(var i=0;i<t[n].length;i++){r[n][i]=[],r[n][i][0]=i?"L":"M";for(var a=0;a<t[n][i].length;a++)r[n][i].push(t[n][i][a])}e||r[n].push(["Z",r[n][0][1],r[n][0][2]])}return r}function rt(t,e){for(var r,n,i=[],a=[],o=0;o<e.length;o++){var s=e[o];n=s._module.selectPoints(s,t),a.push(n),r=tt(n,s),i=i.concat(r)}return i}function nt(t,e,r,n,i){var a,o,s,l=!!n;i&&(a=i.plotinfo,o=i.xaxes[0]._id,s=i.yaxes[0]._id);var u=[],c=[],f=ot(t),h=t._fullLayout;if(a){var d=h._zoomlayer,g=h.dragmode,y=p(g),m=v(g);if(y||m){var x=C(t,o,"x"),b=C(t,s,"y");if(x&&b){var _=d.selectAll(".select-outline-"+a.id);if(_&&t._fullLayout._outlining&&_.length){for(var T=_[0][0].getAttribute("d"),k=w(T,t,a),A=[],M=0;M<k.length;M++){for(var S=k[M],E=[],L=0;L<S.length;L++)E.push([lt(x,S[L][1]),lt(b,S[L][2])]);E.xref=o,E.yref=s,E.subtract=st(E,A),A.push(E)}f=f.concat(A)}}}}var P=o&&s?[o+s]:h._subplots.cartesian;!function(t){var e=t.calcdata;if(e)for(var r=0;r<e.length;r++){var n=e[r][0].trace,i=t._fullLayout._splomScenes;if(i){var a=i[n.uid];a&&(a.selectBatch=[])}}}(t);for(var O={},I=0;I<P.length;I++){var z=P[I],D=z.indexOf("y"),R=z.slice(0,D),F=z.slice(D),B=o&&s?r:void 0;if(B=at(f,R,F,B)){var N=n;if(!l){var j=C(t,R,"x"),U=C(t,F,"y");N=Z(t,[j],[U],z);for(var V=0;V<N.length;V++){var q=N[V],H=q.cd[0],G=H.trace;if("scattergl"===q._module.name&&!H.t.xpx){var W=G.x,Y=G.y,X=G._length;H.t.xpx=[],H.t.ypx=[];for(var K=0;K<X;K++)H.t.xpx[K]=j.c2p(W[K]),H.t.ypx[K]=U.c2p(Y[K])}"splom"===q._module.name&&(O[G.uid]||(O[G.uid]=!0))}}var J=rt(B,N);u=u.concat(J),c=c.concat(N)}}var Q={points:u};$(t,c,Q);var tt=h.clickmode.indexOf("event")>-1&&e;if(!a&&e){var et=ot(t,!0);if(et.length){var nt=et[0].xref,pt=et[0].yref;if(nt&&pt){var dt=ut(et);ct([C(t,nt,"x"),C(t,pt,"y")])(Q,dt)}}t._fullLayout._noEmitSelectedAtStart?t._fullLayout._noEmitSelectedAtStart=!1:tt&&ft(t,Q),h._reselect=!1}if(!a&&h._deselect){var vt=h._deselect;(function(t,e,r){for(var n=0;n<r.length;n++){var i=r[n];if(i.xaxis&&i.xaxis._id===t&&i.yaxis&&i.yaxis._id===e)return!0}return!1})(o=vt.xref,s=vt.yref,c)||it(t,o,s,n),tt&&(Q.points.length?ft(t,Q):ht(t)),h._deselect=!1}return{eventData:Q,selectionTesters:r}}function it(t,e,r,n){n=Z(t,[C(t,e,"x")],[C(t,r,"y")],e+r);for(var i=0;i<n.length;i++){var a=n[i];a._module.selectPoints(a,!1)}$(t,n)}function at(t,e,r,n){for(var i,a=0;a<t.length;a++){var o=t[a];e===o.xref&&r===o.yref&&(i?n=G(i=Q(i,o,!!o.subtract)):(i=[o],n=R(o)))}return n}function ot(t,e){for(var r=[],n=t._fullLayout,i=n.selections,a=i.length,o=0;o<a;o++)if(!e||o===n._activeSelectionIndex){var s=i[o];if(s){var l,u,c,f,h,p=s.xref,d=s.yref,v=C(t,p,"x"),m=C(t,d,"y");if("rect"===s.type){h=[];var x=lt(v,s.x0),b=lt(v,s.x1),_=lt(m,s.y0),w=lt(m,s.y1);h=[[x,_],[x,w],[b,w],[b,_]],l=Math.min(x,b),u=Math.max(x,b),c=Math.min(_,w),f=Math.max(_,w),h.xmin=l,h.xmax=u,h.ymin=c,h.ymax=f,h.xref=p,h.yref=d,h.subtract=!1,h.isRect=!0,r.push(h)}else if("path"===s.type)for(var T=s.path.split("Z"),k=[],A=0;A<T.length;A++){var M=T[A];if(M){M+="Z";var S=g.extractPathCoords(M,y.paramIsX,"raw"),E=g.extractPathCoords(M,y.paramIsY,"raw");l=1/0,u=-1/0,c=1/0,f=-1/0,h=[];for(var L=0;L<S.length;L++){var P=lt(v,S[L]),O=lt(m,E[L]);h.push([P,O]),l=Math.min(P,l),u=Math.max(P,u),c=Math.min(O,c),f=Math.max(O,f)}h.xmin=l,h.xmax=u,h.ymin=c,h.ymax=f,h.xref=p,h.yref=d,h.subtract=st(h,k),k.push(h),r.push(h)}}}}return r}function st(t,e){for(var r=!1,n=0;n<e.length;n++)for(var a=e[n],o=0;o<t.length;o++)if(i(t[o],a)){r=!r;break}return r}function lt(t,e){return"date"===t.type&&(e=e.replace("_"," ")),"log"===t.type?t.c2p(e):t.r2p(e,null,t.calendar)}function ut(t){for(var e=t.length,r=[],n=0;n<e;n++){var i=t[n];r=(r=r.concat(i)).concat([i[0]])}return(a=r).isRect=5===a.length&&a[0][0]===a[4][0]&&a[0][1]===a[4][1]&&a[0][0]===a[1][0]&&a[2][0]===a[3][0]&&a[0][1]===a[3][1]&&a[1][1]===a[2][1]||a[0][1]===a[1][1]&&a[2][1]===a[3][1]&&a[0][0]===a[3][0]&&a[1][0]===a[2][0],a.isRect&&(a.xmin=Math.min(a[0][0],a[2][0]),a.xmax=Math.max(a[0][0],a[2][0]),a.ymin=Math.min(a[0][1],a[2][1]),a.ymax=Math.max(a[0][1],a[2][1])),a;var a}function ct(t){return function(e,r){for(var n,i,a=0;a<t.length;a++){var o=t[a],s=o._id,l=s.charAt(0);if(r.isRect){n||(n={});var u=r[l+"min"],c=r[l+"max"];void 0!==u&&void 0!==c&&(n[s]=[B(o,u),B(o,c)].sort(S))}else i||(i={}),i[s]=r.map(N(o))}n&&(e.range=n),i&&(e.lassoPoints=i)}}function ft(t,e){e&&(e.selections=(t.layout||{}).selections||[]),t.emit("plotly_selected",e)}function ht(t){t.emit("plotly_deselect",null)}t.exports={reselect:nt,prepSelect:function(t,e,r,n,i){var u=!U(n),c=f(i),g=h(i),y=d(i),x=p(i),b=v(i),w="drawcircle"===i,T="drawline"===i||w,k=n.gd,A=k._fullLayout,S=b&&"immediate"===A.newselection.mode&&u,E=A._zoomlayer,C=n.element.getBoundingClientRect(),P=n.plotinfo,O=j(P),F=e-C.left,B=r-C.top;A._calcInverseTransform(k);var N=M.apply3DTransform(A._invTransform)(F,B);F=N[0],B=N[1];var q,H,X,K,J,tt,at,ot=A._invScaleX,st=A._invScaleY,lt=F,pt=B,dt="M"+F+","+B,vt=n.xaxes[0],gt=n.yaxes[0],yt=vt._length,mt=gt._length,xt=t.altKey&&!(p(i)&&y);W(t,k,n),c&&(q=D([[F,B]],I.BENDPX));var bt=E.selectAll("path.select-outline-"+P.id).data([1]),_t=x?A.newshape:A.newselection;x&&(n.hasText=_t.label.text||_t.label.texttemplate);var wt=x&&!y?_t.fillcolor:"rgba(0,0,0,0)",Tt=_t.line.color||(u?s.contrast(k._fullLayout.plot_bgcolor):"#7f7f7f");bt.enter().append("path").attr("class","select-outline select-outline-"+P.id).style({opacity:x?_t.opacity/2:1,"stroke-dasharray":o(_t.line.dash,_t.line.width),"stroke-width":_t.line.width+"px","shape-rendering":"crispEdges"}).call(s.stroke,Tt).call(s.fill,wt).attr("fill-rule","evenodd").classed("cursor-move",!!x).attr("transform",O).attr("d",dt+"Z");var kt=E.append("path").attr("class","zoombox-corners").style({fill:s.background,stroke:s.defaultLine,"stroke-width":1}).attr("transform",O).attr("d","M0,0Z");if(x&&n.hasText){var At=E.select(".label-temp");At.empty()&&(At=E.append("g").classed("label-temp",!0).classed("select-outline",!0).style({opacity:.8}))}var Mt=A._uid+I.SELECTID,St=[],Et=Z(k,n.xaxes,n.yaxes,n.subplot);S&&!t.shiftKey&&(n._clearSubplotSelections=function(){if(u){var t=vt._id,e=gt._id;it(k,t,e,Et);for(var r=(k.layout||{}).selections||[],n=[],i=!1,o=0;o<r.length;o++){var s=A.selections[o];s.xref!==t||s.yref!==e?n.push(r[o]):i=!0}i&&(k._fullLayout._noEmitSelectedAtStart=!0,a.call("_guiRelayout",k,{selections:n}))}});var Lt=function(t){return t.plotinfo.fillRangeItems||ct(t.xaxes.concat(t.yaxes))}(n);n.moveFn=function(t,e){n._clearSubplotSelections&&(n._clearSubplotSelections(),n._clearSubplotSelections=void 0),lt=Math.max(0,Math.min(yt,ot*t+F)),pt=Math.max(0,Math.min(mt,st*e+B));var r=Math.abs(lt-F),i=Math.abs(pt-B);if(g){var a,o,s;if(b){var l=A.selectdirection;switch(a="any"===l?i<Math.min(.6*r,z)?"h":r<Math.min(.6*i,z)?"v":"d":l){case"h":o=w?mt/2:0,s=mt;break;case"v":o=w?yt/2:0,s=yt}}if(x)switch(A.newshape.drawdirection){case"vertical":a="h",o=w?mt/2:0,s=mt;break;case"horizontal":a="v",o=w?yt/2:0,s=yt;break;case"ortho":r<i?(a="h",o=B,s=pt):(a="v",o=F,s=lt);break;default:a="d"}"h"===a?((K=T?_(w,[lt,o],[lt,s]):[[F,o],[F,s],[lt,s],[lt,o]]).xmin=T?lt:Math.min(F,lt),K.xmax=T?lt:Math.max(F,lt),K.ymin=Math.min(o,s),K.ymax=Math.max(o,s),kt.attr("d","M"+K.xmin+","+(B-z)+"h-4v"+2*z+"h4ZM"+(K.xmax-1)+","+(B-z)+"h4v"+2*z+"h-4Z")):"v"===a?((K=T?_(w,[o,pt],[s,pt]):[[o,B],[o,pt],[s,pt],[s,B]]).xmin=Math.min(o,s),K.xmax=Math.max(o,s),K.ymin=T?pt:Math.min(B,pt),K.ymax=T?pt:Math.max(B,pt),kt.attr("d","M"+(F-z)+","+K.ymin+"v-4h"+2*z+"v4ZM"+(F-z)+","+(K.ymax-1)+"v4h"+2*z+"v-4Z")):"d"===a&&((K=T?_(w,[F,B],[lt,pt]):[[F,B],[F,pt],[lt,pt],[lt,B]]).xmin=Math.min(F,lt),K.xmax=Math.max(F,lt),K.ymin=Math.min(B,pt),K.ymax=Math.max(B,pt),kt.attr("d","M0,0Z"))}else c&&(q.addPt([lt,pt]),K=q.filtered);if(n.selectionDefs&&n.selectionDefs.length?(X=Q(n.mergedPolygons,K,xt),K.subtract=xt,H=G(n.selectionDefs.concat([K]))):(X=[K],H=R(K)),m(et(X,y),bt,n),b){var u,f=nt(k,!1),h=f.eventData?f.eventData.points.slice():[];f=nt(k,!1,H,Et,n),H=f.selectionTesters,at=f.eventData,u=q?q.filtered:ut(X),L.throttle(Mt,I.SELECTDELAY,(function(){for(var t=(St=rt(H,Et)).slice(),e=0;e<h.length;e++){for(var r=h[e],n=!1,i=0;i<t.length;i++)if(t[i].curveNumber===r.curveNumber&&t[i].pointNumber===r.pointNumber){n=!0;break}n||t.push(r)}t.length&&(at||(at={}),at.points=t),Lt(at,u),function(t,e){t.emit("plotly_selecting",e)}(k,at)}))}},n.clickFn=function(t,e){if(kt.remove(),k._fullLayout._activeShapeIndex>=0)k._fullLayout._deactivateShape(k);else if(!x){var r=A.clickmode;L.done(Mt).then((function(){if(L.clear(Mt),2===t){for(bt.remove(),J=0;J<Et.length;J++)(tt=Et[J])._module.selectPoints(tt,!1);if($(k,Et),Y(n),ht(k),Et.length){var i=Et[0].xaxis,o=Et[0].yaxis;if(i&&o){for(var s=[],u=k._fullLayout.selections,c=0;c<u.length;c++){var f=u[c];f&&(f.xref===i._id&&f.yref===o._id||s.push(f))}s.length<u.length&&(k._fullLayout._noEmitSelectedAtStart=!0,a.call("_guiRelayout",k,{selections:s}))}}}else r.indexOf("select")>-1&&V(e,k,n.xaxes,n.yaxes,n.subplot,n,bt),"event"===r&&ft(k,void 0);l.click(k,e,P.id)})).catch(M.error)}},n.doneFn=function(){kt.remove(),L.done(Mt).then((function(){L.clear(Mt),!S&&K&&n.selectionDefs&&(K.subtract=xt,n.selectionDefs.push(K),n.mergedPolygons.length=0,[].push.apply(n.mergedPolygons,X)),(S||x)&&Y(n,S),n.doneFnCompleted&&n.doneFnCompleted(St),b&&ft(k,at)})).catch(M.error)}},clearOutline:x,clearSelectionsCache:Y,selectOnClick:V}},46056:function(t,e,r){"use strict";var n=r(13916),i=r(25376),a=r(52904).line,o=r(98192).u,s=r(92880).extendFlat,l=r(31780).templatedArray,u=(r(36208),r(45464)),c=r(21776).ye,f=r(97728);t.exports=l("shape",{visible:s({},u.visible,{editType:"calc+arraydraw"}),showlegend:{valType:"boolean",dflt:!1,editType:"calc+arraydraw"},legend:s({},u.legend,{editType:"calc+arraydraw"}),legendgroup:s({},u.legendgroup,{editType:"calc+arraydraw"}),legendgrouptitle:{text:s({},u.legendgrouptitle.text,{editType:"calc+arraydraw"}),font:i({editType:"calc+arraydraw"}),editType:"calc+arraydraw"},legendrank:s({},u.legendrank,{editType:"calc+arraydraw"}),legendwidth:s({},u.legendwidth,{editType:"calc+arraydraw"}),type:{valType:"enumerated",values:["circle","rect","path","line"],editType:"calc+arraydraw"},layer:{valType:"enumerated",values:["below","above","between"],dflt:"above",editType:"arraydraw"},xref:s({},n.xref,{}),xsizemode:{valType:"enumerated",values:["scaled","pixel"],dflt:"scaled",editType:"calc+arraydraw"},xanchor:{valType:"any",editType:"calc+arraydraw"},x0:{valType:"any",editType:"calc+arraydraw"},x1:{valType:"any",editType:"calc+arraydraw"},yref:s({},n.yref,{}),ysizemode:{valType:"enumerated",values:["scaled","pixel"],dflt:"scaled",editType:"calc+arraydraw"},yanchor:{valType:"any",editType:"calc+arraydraw"},y0:{valType:"any",editType:"calc+arraydraw"},y1:{valType:"any",editType:"calc+arraydraw"},path:{valType:"string",editType:"calc+arraydraw"},opacity:{valType:"number",min:0,max:1,dflt:1,editType:"arraydraw"},line:{color:s({},a.color,{editType:"arraydraw"}),width:s({},a.width,{editType:"calc+arraydraw"}),dash:s({},o,{editType:"arraydraw"}),editType:"calc+arraydraw"},fillcolor:{valType:"color",dflt:"rgba(0,0,0,0)",editType:"arraydraw"},fillrule:{valType:"enumerated",values:["evenodd","nonzero"],dflt:"evenodd",editType:"arraydraw"},editable:{valType:"boolean",dflt:!1,editType:"calc+arraydraw"},label:{text:{valType:"string",dflt:"",editType:"arraydraw"},texttemplate:c({},{keys:Object.keys(f)}),font:i({editType:"calc+arraydraw",colorEditType:"arraydraw"}),textposition:{valType:"enumerated",values:["top left","top center","top right","middle left","middle center","middle right","bottom left","bottom center","bottom right","start","middle","end"],editType:"arraydraw"},textangle:{valType:"angle",dflt:"auto",editType:"calc+arraydraw"},xanchor:{valType:"enumerated",values:["auto","left","center","right"],dflt:"auto",editType:"calc+arraydraw"},yanchor:{valType:"enumerated",values:["top","middle","bottom"],editType:"calc+arraydraw"},padding:{valType:"number",dflt:3,min:0,editType:"arraydraw"},editType:"arraydraw"},editType:"arraydraw"})},96084:function(t,e,r){"use strict";var n=r(3400),i=r(54460),a=r(85448),o=r(65152);function s(t){return u(t.line.width,t.xsizemode,t.x0,t.x1,t.path,!1)}function l(t){return u(t.line.width,t.ysizemode,t.y0,t.y1,t.path,!0)}function u(t,e,r,i,s,l){var u=t/2,c=l;if("pixel"===e){var f=s?o.extractPathCoords(s,l?a.paramIsY:a.paramIsX):[r,i],h=n.aggNums(Math.max,null,f),p=n.aggNums(Math.min,null,f),d=p<0?Math.abs(p)+u:u,v=h>0?h+u:u;return{ppad:u,ppadplus:c?d:v,ppadminus:c?v:d}}return{ppad:u}}function c(t,e,r,n,i){var s="category"===t.type||"multicategory"===t.type?t.r2c:t.d2c;if(void 0!==e)return[s(e),s(r)];if(n){var l,u,c,f,h=1/0,p=-1/0,d=n.match(a.segmentRE);for("date"===t.type&&(s=o.decodeDate(s)),l=0;l<d.length;l++)void 0!==(u=i[d[l].charAt(0)].drawn)&&(!(c=d[l].substr(1).match(a.paramRE))||c.length<u||((f=s(c[u]))<h&&(h=f),f>p&&(p=f)));return p>=h?[h,p]:void 0}}t.exports=function(t){var e=t._fullLayout,r=n.filterVisible(e.shapes);if(r.length&&t._fullData.length)for(var o=0;o<r.length;o++){var u,f,h=r[o];h._extremes={};var p=i.getRefType(h.xref),d=i.getRefType(h.yref);if("paper"!==h.xref&&"domain"!==p){var v="pixel"===h.xsizemode?h.xanchor:h.x0,g="pixel"===h.xsizemode?h.xanchor:h.x1;(f=c(u=i.getFromId(t,h.xref),v,g,h.path,a.paramIsX))&&(h._extremes[u._id]=i.findExtremes(u,f,s(h)))}if("paper"!==h.yref&&"domain"!==d){var y="pixel"===h.ysizemode?h.yanchor:h.y0,m="pixel"===h.ysizemode?h.yanchor:h.y1;(f=c(u=i.getFromId(t,h.yref),y,m,h.path,a.paramIsY))&&(h._extremes[u._id]=i.findExtremes(u,f,l(h)))}}}},85448:function(t){"use strict";t.exports={segmentRE:/[MLHVQCTSZ][^MLHVQCTSZ]*/g,paramRE:/[^\s,]+/g,paramIsX:{M:{0:!0,drawn:0},L:{0:!0,drawn:0},H:{0:!0,drawn:0},V:{},Q:{0:!0,2:!0,drawn:2},C:{0:!0,2:!0,4:!0,drawn:4},T:{0:!0,drawn:0},S:{0:!0,2:!0,drawn:2},Z:{}},paramIsY:{M:{1:!0,drawn:1},L:{1:!0,drawn:1},H:{},V:{0:!0,drawn:0},Q:{1:!0,3:!0,drawn:3},C:{1:!0,3:!0,5:!0,drawn:5},T:{1:!0,drawn:1},S:{1:!0,3:!0,drawn:5},Z:{}},numParams:{M:2,L:2,H:1,V:1,Q:4,C:6,T:2,S:4,Z:0}}},43712:function(t,e,r){"use strict";var n=r(3400),i=r(54460),a=r(51272),o=r(46056),s=r(65152);function l(t,e,r){function a(r,i){return n.coerce(t,e,o,r,i)}if(e._isShape=!0,a("visible")){a("showlegend")&&(a("legend"),a("legendwidth"),a("legendgroup"),a("legendgrouptitle.text"),n.coerceFont(a,"legendgrouptitle.font"),a("legendrank"));var l=a("path"),u=a("type",l?"path":"rect"),c="path"!==u;c&&delete e.path,a("editable"),a("layer"),a("opacity"),a("fillcolor"),a("fillrule"),a("line.width")&&(a("line.color"),a("line.dash"));for(var f=a("xsizemode"),h=a("ysizemode"),p=["x","y"],d=0;d<2;d++){var v,g,y,m=p[d],x=m+"anchor",b="x"===m?f:h,_={_fullLayout:r},w=i.coerceRef(t,e,_,m,void 0,"paper");if("range"===i.getRefType(w)?((v=i.getFromId(_,w))._shapeIndices.push(e._index),y=s.rangeToShapePosition(v),g=s.shapePositionToRange(v)):g=y=n.identity,c){var T=m+"0",k=m+"1",A=t[T],M=t[k];t[T]=g(t[T],!0),t[k]=g(t[k],!0),"pixel"===b?(a(T,0),a(k,10)):(i.coercePosition(e,_,a,w,T,.25),i.coercePosition(e,_,a,w,k,.75)),e[T]=y(e[T]),e[k]=y(e[k]),t[T]=A,t[k]=M}if("pixel"===b){var S=t[x];t[x]=g(t[x],!0),i.coercePosition(e,_,a,w,x,.25),e[x]=y(e[x]),t[x]=S}}c&&n.noneOrAll(t,e,["x0","x1","y0","y1"]);var E,L,C="line"===u;if(c&&(E=a("label.texttemplate")),E||(L=a("label.text")),L||E){a("label.textangle");var P=a("label.textposition",C?"middle":"middle center");a("label.xanchor"),a("label.yanchor",function(t,e){return t?"bottom":-1!==e.indexOf("top")?"top":-1!==e.indexOf("bottom")?"bottom":"middle"}(C,P)),a("label.padding"),n.coerceFont(a,"label.font",r.font)}}}t.exports=function(t,e){a(t,e,{name:"shapes",handleItemDefaults:l})}},60728:function(t,e,r){"use strict";var n=r(3400),i=r(54460),a=r(72736),o=r(43616),s=r(9856).readPaths,l=r(65152),u=l.getPathString,c=r(97728),f=r(84284).FROM_TL;t.exports=function(t,e,r,h){if(h.selectAll(".shape-label").remove(),r.label.text||r.label.texttemplate){var p;if(r.label.texttemplate){var d={};if("path"!==r.type){var v=i.getFromId(t,r.xref),g=i.getFromId(t,r.yref);for(var y in c){var m=c[y](r,v,g);void 0!==m&&(d[y]=m)}}p=n.texttemplateStringForShapes(r.label.texttemplate,{},t._fullLayout._d3locale,d)}else p=r.label.text;var x,b,_,w,T={"data-index":e},k=r.label.font,A=h.append("g").attr(T).classed("shape-label",!0).append("text").attr({"data-notex":1}).classed("shape-label-text",!0).text(p);if(r.path){var M=u(t,r),S=s(M,t);x=1/0,_=1/0,b=-1/0,w=-1/0;for(var E=0;E<S.length;E++)for(var L=0;L<S[E].length;L++)for(var C=S[E][L],P=1;P<C.length;P+=2){var O=C[P],I=C[P+1];x=Math.min(x,O),b=Math.max(b,O),_=Math.min(_,I),w=Math.max(w,I)}}else{var z=i.getFromId(t,r.xref),D=i.getRefType(r.xref),R=i.getFromId(t,r.yref),F=i.getRefType(r.yref),B=l.getDataToPixel(t,z,!1,D),N=l.getDataToPixel(t,R,!0,F);x=B(r.x0),b=B(r.x1),_=N(r.y0),w=N(r.y1)}var j=r.label.textangle;"auto"===j&&(j="line"===r.type?function(t,e,r,n){var i,a;return a=Math.abs(r-t),i=r>=t?e-n:n-e,-180/Math.PI*Math.atan2(i,a)}(x,_,b,w):0),A.call((function(e){return e.call(o.font,k).attr({}),a.convertToTspans(e,t),e}));var U=function(t,e,r,n,i,a,o){var s,l,u,c,h=i.label.textposition,p=i.label.textangle,d=i.label.padding,v=i.type,g=Math.PI/180*a,y=Math.sin(g),m=Math.cos(g),x=i.label.xanchor,b=i.label.yanchor;if("line"===v){"start"===h?(s=t,l=e):"end"===h?(s=r,l=n):(s=(t+r)/2,l=(e+n)/2),"auto"===x&&(x="start"===h?"auto"===p?r>t?"left":r<t?"right":"center":r>t?"right":r<t?"left":"center":"end"===h?"auto"===p?r>t?"right":r<t?"left":"center":r>t?"left":r<t?"right":"center":"center");var _={bottom:-1,middle:0,top:1};if("auto"===p){var w=_[b];u=-d*y*w,c=d*m*w}else u=d*{left:1,center:0,right:-1}[x],c=d*_[b];s+=u,l+=c}else u=d+3,-1!==h.indexOf("right")?(s=Math.max(t,r)-u,"auto"===x&&(x="right")):-1!==h.indexOf("left")?(s=Math.min(t,r)+u,"auto"===x&&(x="left")):(s=(t+r)/2,"auto"===x&&(x="center")),l=-1!==h.indexOf("top")?Math.min(e,n):-1!==h.indexOf("bottom")?Math.max(e,n):(e+n)/2,c=d,"bottom"===b?l-=c:"top"===b&&(l+=c);var T=f[b],k=i.label.font.size,A=o.height;return{textx:s+(A*T-k)*y,texty:l+-(A*T-k)*m,xanchor:x}}(x,_,b,w,r,j,o.bBox(A.node())),V=U.textx,q=U.texty,H=U.xanchor;A.attr({"text-anchor":{left:"start",center:"middle",right:"end"}[H],y:q,x:V,transform:"rotate("+j+","+V+","+q+")"}).call(a.positionText,V,q)}}},55496:function(t,e,r){"use strict";var n=r(3400).strTranslate,i=r(86476),a=r(72760),o=a.drawMode,s=a.selectMode,l=r(24040),u=r(76308),c=r(7e3),f=c.i000,h=c.i090,p=c.i180,d=c.i270,v=r(1936).clearOutlineControllers,g=r(9856),y=g.pointsOnRectangle,m=g.pointsOnEllipse,x=g.writePaths,b=r(93940).newShapes,_=r(93940).createShapeObj,w=r(5968),T=r(60728);function k(t,e){var r,n,i,a=t[e][1],o=t[e][2],s=t.length;return n=t[r=(e+1)%s][1],i=t[r][2],n===a&&i===o&&(n=t[r=(e+2)%s][1],i=t[r][2]),[r,n,i]}t.exports=function t(e,r,a,c){c||(c=0);var g=a.gd;function A(){t(e,r,a,c++),(m(e[0])||a.hasText)&&M({redrawing:!0})}function M(t){var e={};void 0!==a.isActiveShape&&(a.isActiveShape=!1,e=b(r,a)),void 0!==a.isActiveSelection&&(a.isActiveSelection=!1,e=w(r,a),g._fullLayout._reselect=!0),Object.keys(e).length&&l.call((t||{}).redrawing?"relayout":"_guiRelayout",g,e)}var S,E,L,C,P,O=g._fullLayout._zoomlayer,I=a.dragmode,z=o(I),D=s(I);if((z||D)&&(g._fullLayout._outlining=!0),v(g),r.attr("d",x(e)),c||!a.isActiveShape&&!a.isActiveSelection||(P=function(t,e){for(var r=0;r<e.length;r++){var n=e[r];t[r]=[];for(var i=0;i<n.length;i++){t[r][i]=[];for(var a=0;a<n[i].length;a++)t[r][i][a]=n[i][a]}}return t}([],e),function(t){S=[];for(var r=0;r<e.length;r++){var o=e[r],s=y(o),l=!s&&m(o);S[r]=[];for(var c=o.length,v=0;v<c;v++)if("Z"!==o[v][0]&&(!l||v===f||v===h||v===p||v===d)){var x,b=s&&a.isActiveSelection;b&&(x=k(o,v));var _=o[v][1],w=o[v][2],T=t.append(b?"rect":"circle").attr("data-i",r).attr("data-j",v).style({fill:u.background,stroke:u.defaultLine,"stroke-width":1,"shape-rendering":"crispEdges"});if(b){var A=x[1]-_,M=x[2]-w,E=M?5:Math.max(Math.min(25,Math.abs(A)-5),5),L=A?5:Math.max(Math.min(25,Math.abs(M)-5),5);T.classed(M?"cursor-ew-resize":"cursor-ns-resize",!0).attr("width",E).attr("height",L).attr("x",_-E/2).attr("y",w-L/2).attr("transform",n(A/2,M/2))}else T.classed("cursor-grab",!0).attr("r",5).attr("cx",_).attr("cy",w);S[r][v]={element:T.node(),gd:g,prepFn:B,doneFn:j,clickFn:U},i.init(S[r][v])}}}(O.append("g").attr("class","outline-controllers")),function(){if(E=[],e.length){E[0]={element:r[0][0],gd:g,prepFn:q,doneFn:H,clickFn:G},i.init(E[0])}}()),z&&a.hasText){var R=O.select(".label-temp"),F=_(r,a,a.dragmode);T(g,"label-temp",F,R)}function B(t){L=+t.srcElement.getAttribute("data-i"),C=+t.srcElement.getAttribute("data-j"),S[L][C].moveFn=N}function N(t,r){if(e.length){var n=P[L][C][1],i=P[L][C][2],o=e[L],s=o.length;if(y(o)){var l=t,u=r;a.isActiveSelection&&(k(o,C)[1]===o[C][1]?u=0:l=0);for(var c=0;c<s;c++)if(c!==C){var f=o[c];f[1]===o[C][1]&&(f[1]=n+l),f[2]===o[C][2]&&(f[2]=i+u)}if(o[C][1]=n+l,o[C][2]=i+u,!y(o))for(var h=0;h<s;h++)for(var p=0;p<o[h].length;p++)o[h][p]=P[L][h][p]}else o[C][1]=n+t,o[C][2]=i+r;A()}}function j(){M()}function U(t,r){if(2===t){L=+r.srcElement.getAttribute("data-i"),C=+r.srcElement.getAttribute("data-j");var n=e[L];y(n)||m(n)||function(){if(e.length&&e[L]&&e[L].length){for(var t=[],r=0;r<e[L].length;r++)r!==C&&t.push(e[L][r]);t.length>1&&(2!==t.length||"Z"!==t[1][0])&&(0===C&&(t[0][0]="M"),e[L]=t,A(),M())}}()}}function V(t,r){!function(t,r){if(e.length)for(var n=0;n<e.length;n++)for(var i=0;i<e[n].length;i++)for(var a=0;a+2<e[n][i].length;a+=2)e[n][i][a+1]=P[n][i][a+1]+t,e[n][i][a+2]=P[n][i][a+2]+r}(t,r),A()}function q(t){(L=+t.srcElement.getAttribute("data-i"))||(L=0),E[L].moveFn=V}function H(){M()}function G(t){2===t&&function(t){if(s(t._fullLayout.dragmode)){v(t);var e=t._fullLayout._activeSelectionIndex,r=(t.layout||{}).selections||[];if(e<r.length){for(var n=[],i=0;i<r.length;i++)i!==e&&n.push(r[i]);delete t._fullLayout._activeSelectionIndex;var a=t._fullLayout.selections[e];t._fullLayout._deselect={xref:a.xref,yref:a.yref},l.call("_guiRelayout",t,{selections:n})}}}(g)}}},4016:function(t,e,r){"use strict";var n=r(33428),i=r(24040),a=r(3400),o=r(54460),s=r(9856).readPaths,l=r(55496),u=r(60728),c=r(1936).clearOutlineControllers,f=r(76308),h=r(43616),p=r(31780).arrayEditor,d=r(86476),v=r(93972),g=r(85448),y=r(65152),m=y.getPathString;function x(t){var e=t._fullLayout;for(var r in e._shapeUpperLayer.selectAll("path").remove(),e._shapeLowerLayer.selectAll("path").remove(),e._shapeUpperLayer.selectAll("text").remove(),e._shapeLowerLayer.selectAll("text").remove(),e._plots){var n=e._plots[r].shapelayer;n&&(n.selectAll("path").remove(),n.selectAll("text").remove())}for(var i=0;i<e.shapes.length;i++)!0===e.shapes[i].visible&&w(t,i)}function b(t){return!!t._fullLayout._outlining}function _(t){return!t._context.edits.shapePosition}function w(t,e){t._fullLayout._paperdiv.selectAll('.shapelayer [data-index="'+e+'"]').remove();var r=y.makeShapesOptionsAndPlotinfo(t,e),c=r.options,w=r.plotinfo;function M(r){var M=m(t,c),S={"data-index":e,"fill-rule":c.fillrule,d:M},E=c.opacity,L=c.fillcolor,C=c.line.width?c.line.color:"rgba(0,0,0,0)",P=c.line.width,O=c.line.dash;P||!0!==c.editable||(P=5,O="solid");var I="Z"!==M[M.length-1],z=_(t)&&c.editable&&t._fullLayout._activeShapeIndex===e;z&&(L=I?"rgba(0,0,0,0)":t._fullLayout.activeshape.fillcolor,E=t._fullLayout.activeshape.opacity);var D,R=r.append("g").classed("shape-group",!0).attr({"data-index":e}),F=R.append("path").attr(S).style("opacity",E).call(f.stroke,C).call(f.fill,L).call(h.dashLine,O,P);if(T(R,t,c),u(t,e,c,R),(z||t._context.edits.shapePosition)&&(D=p(t.layout,"shapes",c)),z){F.style({cursor:"move"});var B={element:F.node(),plotinfo:w,gd:t,editHelpers:D,hasText:c.label.text||c.label.texttemplate,isActiveShape:!0},N=s(M,t);l(N,F,B)}else t._context.edits.shapePosition?function(t,e,r,s,l,c){var f,p,x,_,w,A,M,S,E,L,C,P,O,I,z,D,R=10,F=10,B="pixel"===r.xsizemode,N="pixel"===r.ysizemode,j="line"===r.type,U="path"===r.type,V=c.modifyItem,q=n.select(e.node().parentNode),H=o.getFromId(t,r.xref),G=o.getRefType(r.xref),W=o.getFromId(t,r.yref),Y=o.getRefType(r.yref),X=y.getDataToPixel(t,H,!1,G),Z=y.getDataToPixel(t,W,!0,Y),K=y.getPixelToData(t,H,!1,G),J=y.getPixelToData(t,W,!0,Y),$=j?function(){var t=10,n=Math.max(r.line.width,t),i=l.append("g").attr("data-index",s).attr("drag-helper",!0);i.append("path").attr("d",e.attr("d")).style({cursor:"move","stroke-width":n,"stroke-opacity":"0"});var a={"fill-opacity":"0"},o=Math.max(n/2,t);return i.append("circle").attr({"data-line-point":"start-point",cx:B?X(r.xanchor)+r.x0:X(r.x0),cy:N?Z(r.yanchor)-r.y0:Z(r.y0),r:o}).style(a).classed("cursor-grab",!0),i.append("circle").attr({"data-line-point":"end-point",cx:B?X(r.xanchor)+r.x1:X(r.x1),cy:N?Z(r.yanchor)-r.y1:Z(r.y1),r:o}).style(a).classed("cursor-grab",!0),i}():e,Q={element:$.node(),gd:t,prepFn:function(n){b(t)||(B&&(w=X(r.xanchor)),N&&(A=Z(r.yanchor)),"path"===r.type?z=r.path:(f=B?r.x0:X(r.x0),p=N?r.y0:Z(r.y0),x=B?r.x1:X(r.x1),_=N?r.y1:Z(r.y1)),f<x?(E=f,O="x0",L=x,I="x1"):(E=x,O="x1",L=f,I="x0"),!N&&p<_||N&&p>_?(M=p,C="y0",S=_,P="y1"):(M=_,C="y1",S=p,P="y0"),tt(n),nt(l,r),function(t,e,r){var n=e.xref,i=e.yref,a=o.getFromId(r,n),s=o.getFromId(r,i),l="";"paper"===n||a.autorange||(l+=n),"paper"===i||s.autorange||(l+=i),h.setClipUrl(t,l?"clip"+r._fullLayout._uid+l:null,r)}(e,r,t),Q.moveFn="move"===D?et:rt,Q.altKey=n.altKey)},doneFn:function(){b(t)||(v(e),it(l),T(e,t,r),i.call("_guiRelayout",t,c.getUpdateObj()))},clickFn:function(){b(t)||it(l)}};function tt(r){if(b(t))D=null;else if(j)D="path"===r.target.tagName?"move":"start-point"===r.target.attributes["data-line-point"].value?"resize-over-start-point":"resize-over-end-point";else{var n=Q.element.getBoundingClientRect(),i=n.right-n.left,a=n.bottom-n.top,o=r.clientX-n.left,s=r.clientY-n.top,l=!U&&i>R&&a>F&&!r.shiftKey?d.getCursor(o/i,1-s/a):"move";v(e,l),D=l.split("-")[0]}}function et(n,i){if("path"===r.type){var a=function(t){return t},o=a,c=a;B?V("xanchor",r.xanchor=K(w+n)):(o=function(t){return K(X(t)+n)},H&&"date"===H.type&&(o=y.encodeDate(o))),N?V("yanchor",r.yanchor=J(A+i)):(c=function(t){return J(Z(t)+i)},W&&"date"===W.type&&(c=y.encodeDate(c))),V("path",r.path=k(z,o,c))}else B?V("xanchor",r.xanchor=K(w+n)):(V("x0",r.x0=K(f+n)),V("x1",r.x1=K(x+n))),N?V("yanchor",r.yanchor=J(A+i)):(V("y0",r.y0=J(p+i)),V("y1",r.y1=J(_+i)));e.attr("d",m(t,r)),nt(l,r),u(t,s,r,q)}function rt(n,i){if(U){var a=function(t){return t},o=a,c=a;B?V("xanchor",r.xanchor=K(w+n)):(o=function(t){return K(X(t)+n)},H&&"date"===H.type&&(o=y.encodeDate(o))),N?V("yanchor",r.yanchor=J(A+i)):(c=function(t){return J(Z(t)+i)},W&&"date"===W.type&&(c=y.encodeDate(c))),V("path",r.path=k(z,o,c))}else if(j){if("resize-over-start-point"===D){var h=f+n,d=N?p-i:p+i;V("x0",r.x0=B?h:K(h)),V("y0",r.y0=N?d:J(d))}else if("resize-over-end-point"===D){var v=x+n,g=N?_-i:_+i;V("x1",r.x1=B?v:K(v)),V("y1",r.y1=N?g:J(g))}}else{var b=function(t){return-1!==D.indexOf(t)},T=b("n"),G=b("s"),Y=b("w"),$=b("e"),Q=T?M+i:M,tt=G?S+i:S,et=Y?E+n:E,rt=$?L+n:L;N&&(T&&(Q=M-i),G&&(tt=S-i)),(!N&&tt-Q>F||N&&Q-tt>F)&&(V(C,r[C]=N?Q:J(Q)),V(P,r[P]=N?tt:J(tt))),rt-et>R&&(V(O,r[O]=B?et:K(et)),V(I,r[I]=B?rt:K(rt)))}e.attr("d",m(t,r)),nt(l,r),u(t,s,r,q)}function nt(t,e){(B||N)&&function(){var r="path"!==e.type,n=t.selectAll(".visual-cue").data([0]);n.enter().append("path").attr({fill:"#fff","fill-rule":"evenodd",stroke:"#000","stroke-width":1}).classed("visual-cue",!0);var i=X(B?e.xanchor:a.midRange(r?[e.x0,e.x1]:y.extractPathCoords(e.path,g.paramIsX))),o=Z(N?e.yanchor:a.midRange(r?[e.y0,e.y1]:y.extractPathCoords(e.path,g.paramIsY)));if(i=y.roundPositionForSharpStrokeRendering(i,1),o=y.roundPositionForSharpStrokeRendering(o,1),B&&N){var s="M"+(i-1-1)+","+(o-1-1)+"h-8v2h8 v8h2v-8 h8v-2h-8 v-8h-2 Z";n.attr("d",s)}else if(B){var l="M"+(i-1-1)+","+(o-9-1)+"v18 h2 v-18 Z";n.attr("d",l)}else{var u="M"+(i-9-1)+","+(o-1-1)+"h18 v2 h-18 Z";n.attr("d",u)}}()}function it(t){t.selectAll(".visual-cue").remove()}d.init(Q),$.node().onmousemove=tt}(t,F,c,e,r,D):!0===c.editable&&F.style("pointer-events",I||f.opacity(L)*E<=.5?"stroke":"all");F.node().addEventListener("click",(function(){return function(t,e){if(_(t)){var r=+e.node().getAttribute("data-index");if(r>=0){if(r===t._fullLayout._activeShapeIndex)return void A(t);t._fullLayout._activeShapeIndex=r,t._fullLayout._deactivateShape=A,x(t)}}}(t,F)}))}c._input&&!0===c.visible&&("above"===c.layer?M(t._fullLayout._shapeUpperLayer):"paper"===c.xref||"paper"===c.yref?M(t._fullLayout._shapeLowerLayer):"between"===c.layer?M(w.shapelayerBetween):w._hadPlotinfo?M((w.mainplotinfo||w).shapelayer):M(t._fullLayout._shapeLowerLayer))}function T(t,e,r){var n=(r.xref+r.yref).replace(/paper/g,"").replace(/[xyz][1-9]* *domain/g,"");h.setClipUrl(t,n?"clip"+e._fullLayout._uid+n:null,e)}function k(t,e,r){return t.replace(g.segmentRE,(function(t){var n=0,i=t.charAt(0),a=g.paramIsX[i],o=g.paramIsY[i],s=g.numParams[i];return i+t.substr(1).replace(g.paramRE,(function(t){return n>=s||(a[n]?t=e(t):o[n]&&(t=r(t)),n++),t}))}))}function A(t){_(t)&&t._fullLayout._activeShapeIndex>=0&&(c(t),delete t._fullLayout._activeShapeIndex,x(t))}t.exports={draw:x,drawOne:w,eraseActiveShape:function(t){if(_(t)){c(t);var e=t._fullLayout._activeShapeIndex,r=(t.layout||{}).shapes||[];if(e<r.length){for(var n=[],a=0;a<r.length;a++)a!==e&&n.push(r[a]);return delete t._fullLayout._activeShapeIndex,i.call("_guiRelayout",t,{shapes:n})}}},drawLabel:u}},92872:function(t,e,r){"use strict";var n=r(67824).overrideAll,i=r(45464),a=r(25376),o=r(98192).u,s=r(92880).extendFlat,l=r(21776).ye,u=r(97728);t.exports=n({newshape:{visible:s({},i.visible,{}),showlegend:{valType:"boolean",dflt:!1},legend:s({},i.legend,{}),legendgroup:s({},i.legendgroup,{}),legendgrouptitle:{text:s({},i.legendgrouptitle.text,{}),font:a({})},legendrank:s({},i.legendrank,{}),legendwidth:s({},i.legendwidth,{}),line:{color:{valType:"color"},width:{valType:"number",min:0,dflt:4},dash:s({},o,{dflt:"solid"})},fillcolor:{valType:"color",dflt:"rgba(0,0,0,0)"},fillrule:{valType:"enumerated",values:["evenodd","nonzero"],dflt:"evenodd"},opacity:{valType:"number",min:0,max:1,dflt:1},layer:{valType:"enumerated",values:["below","above","between"],dflt:"above"},drawdirection:{valType:"enumerated",values:["ortho","horizontal","vertical","diagonal"],dflt:"diagonal"},name:s({},i.name,{}),label:{text:{valType:"string",dflt:""},texttemplate:l({newshape:!0},{keys:Object.keys(u)}),font:a({}),textposition:{valType:"enumerated",values:["top left","top center","top right","middle left","middle center","middle right","bottom left","bottom center","bottom right","start","middle","end"]},textangle:{valType:"angle",dflt:"auto"},xanchor:{valType:"enumerated",values:["auto","left","center","right"],dflt:"auto"},yanchor:{valType:"enumerated",values:["top","middle","bottom"]},padding:{valType:"number",dflt:3,min:0}}},activeshape:{fillcolor:{valType:"color",dflt:"rgb(255,0,255)"},opacity:{valType:"number",min:0,max:1,dflt:.5}}},"none","from-root")},7e3:function(t){"use strict";t.exports={CIRCLE_SIDES:32,i000:0,i090:8,i180:16,i270:24,cos45:Math.cos(Math.PI/4),sin45:Math.sin(Math.PI/4),SQRT2:Math.sqrt(2)}},65144:function(t,e,r){"use strict";var n=r(76308),i=r(3400);t.exports=function(t,e,r){if(r("newshape.visible"),r("newshape.name"),r("newshape.showlegend"),r("newshape.legend"),r("newshape.legendwidth"),r("newshape.legendgroup"),r("newshape.legendgrouptitle.text"),i.coerceFont(r,"newshape.legendgrouptitle.font"),r("newshape.legendrank"),r("newshape.drawdirection"),r("newshape.layer"),r("newshape.fillcolor"),r("newshape.fillrule"),r("newshape.opacity"),r("newshape.line.width")){var a=(t||{}).plot_bgcolor||"#FFF";r("newshape.line.color",n.contrast(a)),r("newshape.line.dash")}var o="drawline"===t.dragmode,s=r("newshape.label.text"),l=r("newshape.label.texttemplate");if(s||l){r("newshape.label.textangle");var u=r("newshape.label.textposition",o?"middle":"middle center");r("newshape.label.xanchor"),r("newshape.label.yanchor",function(t,e){return t?"bottom":-1!==e.indexOf("top")?"top":-1!==e.indexOf("bottom")?"bottom":"middle"}(o,u)),r("newshape.label.padding"),i.coerceFont(r,"newshape.label.font",e.font)}r("activeshape.fillcolor"),r("activeshape.opacity")}},9856:function(t,e,r){"use strict";var n=r(21984),i=r(7e3),a=i.CIRCLE_SIDES,o=i.SQRT2,s=r(5840),l=s.p2r,u=s.r2p,c=[0,3,4,5,6,1,2],f=[0,3,4,1,2];function h(t,e){return Math.abs(t-e)<=1e-6}function p(t,e){var r=e[1]-t[1],n=e[2]-t[2];return Math.sqrt(r*r+n*n)}e.writePaths=function(t){var e=t.length;if(!e)return"M0,0Z";for(var r="",n=0;n<e;n++)for(var i=t[n].length,a=0;a<i;a++){var o=t[n][a][0];if("Z"===o)r+="Z";else for(var s=t[n][a].length,l=0;l<s;l++){var u=l;"Q"===o||"S"===o?u=f[l]:"C"===o&&(u=c[l]),r+=t[n][a][u],l>0&&l<s-1&&(r+=",")}}return r},e.readPaths=function(t,e,r,i){var o,s,c,f=n(t),h=[],p=-1,d=0,v=0,g=function(){s=d,c=v};g();for(var y=0;y<f.length;y++){var m,x,b,_,w=[],T=f[y][0],k=T;switch(T){case"M":h[++p]=[],d=+f[y][1],v=+f[y][2],w.push([k,d,v]),g();break;case"Q":case"S":m=+f[y][1],b=+f[y][2],d=+f[y][3],v=+f[y][4],w.push([k,d,v,m,b]);break;case"C":m=+f[y][1],b=+f[y][2],x=+f[y][3],_=+f[y][4],d=+f[y][5],v=+f[y][6],w.push([k,d,v,m,b,x,_]);break;case"T":case"L":d=+f[y][1],v=+f[y][2],w.push([k,d,v]);break;case"H":k="L",d=+f[y][1],w.push([k,d,v]);break;case"V":k="L",v=+f[y][1],w.push([k,d,v]);break;case"A":k="L";var A=+f[y][1],M=+f[y][2];+f[y][4]||(A=-A,M=-M);var S=d-A,E=v;for(o=1;o<=a/2;o++){var L=2*Math.PI*o/a;w.push([k,S+A*Math.cos(L),E+M*Math.sin(L)])}break;case"Z":d===s&&v===c||(d=s,v=c,w.push([k,d,v]))}for(var C=(r||{}).domain,P=e._fullLayout._size,O=r&&"pixel"===r.xsizemode,I=r&&"pixel"===r.ysizemode,z=!1===i,D=0;D<w.length;D++){for(o=0;o+2<7;o+=2){var R=w[D][o+1],F=w[D][o+2];void 0!==R&&void 0!==F&&(d=R,v=F,r&&(r.xaxis&&r.xaxis.p2r?(z&&(R-=r.xaxis._offset),R=O?u(r.xaxis,r.xanchor)+R:l(r.xaxis,R)):(z&&(R-=P.l),C?R=C.x[0]+R/P.w:R/=P.w),r.yaxis&&r.yaxis.p2r?(z&&(F-=r.yaxis._offset),F=I?u(r.yaxis,r.yanchor)-F:l(r.yaxis,F)):(z&&(F-=P.t),F=C?C.y[1]-F/P.h:1-F/P.h)),w[D][o+1]=R,w[D][o+2]=F)}h[p].push(w[D].slice())}}return h},e.pointsOnRectangle=function(t){if(5!==t.length)return!1;for(var e=1;e<3;e++){if(!h(t[0][e]-t[1][e],t[3][e]-t[2][e]))return!1;if(!h(t[0][e]-t[3][e],t[1][e]-t[2][e]))return!1}return!(!h(t[0][1],t[1][1])&&!h(t[0][1],t[3][1])||!(p(t[0],t[1])*p(t[0],t[3])))},e.pointsOnEllipse=function(t){var e=t.length;if(e!==a+1)return!1;e=a;for(var r=0;r<e;r++){var n=(2*e-r)%e,i=(e/2+n)%e,o=(e/2+r)%e;if(!h(p(t[r],t[o]),p(t[n],t[i])))return!1}return!0},e.handleEllipse=function(t,r,n){if(!t)return[r,n];var i=e.ellipseOver({x0:r[0],y0:r[1],x1:n[0],y1:n[1]}),s=(i.x1+i.x0)/2,l=(i.y1+i.y0)/2,u=(i.x1-i.x0)/2,c=(i.y1-i.y0)/2;u||(u=c/=o),c||(c=u/=o);for(var f=[],h=0;h<a;h++){var p=2*h*Math.PI/a;f.push([s+u*Math.cos(p),l+c*Math.sin(p)])}return f},e.ellipseOver=function(t){var e=t.x0,r=t.y0,n=t.x1,i=t.y1,a=n-e,s=i-r,l=((e-=a)+n)/2,u=((r-=s)+i)/2;return{x0:l-(a*=o),y0:u-(s*=o),x1:l+a,y1:u+s}},e.fixDatesForPaths=function(t,e,r){var n="date"===e.type,i="date"===r.type;if(!n&&!i)return t;for(var a=0;a<t.length;a++)for(var o=0;o<t[a].length;o++)for(var s=0;s+2<t[a][o].length;s+=2)n&&(t[a][o][s+1]=t[a][o][s+1].replace(" ","_")),i&&(t[a][o][s+2]=t[a][o][s+2].replace(" ","_"));return t}},93940:function(t,e,r){"use strict";var n=r(72760),i=n.drawMode,a=n.openMode,o=r(7e3),s=o.i000,l=o.i090,u=o.i180,c=o.i270,f=o.cos45,h=o.sin45,p=r(5840),d=p.p2r,v=p.r2p,g=r(1936).clearOutline,y=r(9856),m=y.readPaths,x=y.writePaths,b=y.ellipseOver,_=y.fixDatesForPaths;function w(t,e,r){var n,i=t[0][0],o=e.gd,p=i.getAttribute("d"),g=o._fullLayout.newshape,y=e.plotinfo,w=e.isActiveShape,T=y.xaxis,k=y.yaxis,A=!!y.domain||!y.xaxis,M=!!y.domain||!y.yaxis,S=a(r),E=m(p,o,y,w),L={editable:!0,visible:g.visible,name:g.name,showlegend:g.showlegend,legend:g.legend,legendwidth:g.legendwidth,legendgroup:g.legendgroup,legendgrouptitle:{text:g.legendgrouptitle.text,font:g.legendgrouptitle.font},legendrank:g.legendrank,label:g.label,xref:A?"paper":T._id,yref:M?"paper":k._id,layer:g.layer,opacity:g.opacity,line:{color:g.line.color,width:g.line.width,dash:g.line.dash}};if(S||(L.fillcolor=g.fillcolor,L.fillrule=g.fillrule),1===E.length&&(n=E[0]),n&&5===n.length&&"drawrect"===r)L.type="rect",L.x0=n[0][1],L.y0=n[0][2],L.x1=n[2][1],L.y1=n[2][2];else if(n&&"drawline"===r)L.type="line",L.x0=n[0][1],L.y0=n[0][2],L.x1=n[1][1],L.y1=n[1][2];else if(n&&"drawcircle"===r){L.type="circle";var C=n[s][1],P=n[l][1],O=n[u][1],I=n[c][1],z=n[s][2],D=n[l][2],R=n[u][2],F=n[c][2],B=y.xaxis&&("date"===y.xaxis.type||"log"===y.xaxis.type),N=y.yaxis&&("date"===y.yaxis.type||"log"===y.yaxis.type);B&&(C=v(y.xaxis,C),P=v(y.xaxis,P),O=v(y.xaxis,O),I=v(y.xaxis,I)),N&&(z=v(y.yaxis,z),D=v(y.yaxis,D),R=v(y.yaxis,R),F=v(y.yaxis,F));var j=(P+I)/2,U=(z+R)/2,V=b({x0:j,y0:U,x1:j+(I-P+O-C)/2*f,y1:U+(F-D+R-z)/2*h});B&&(V.x0=d(y.xaxis,V.x0),V.x1=d(y.xaxis,V.x1)),N&&(V.y0=d(y.yaxis,V.y0),V.y1=d(y.yaxis,V.y1)),L.x0=V.x0,L.y0=V.y0,L.x1=V.x1,L.y1=V.y1}else L.type="path",T&&k&&_(E,T,k),L.path=x(E),n=null;return L}t.exports={newShapes:function(t,e){if(t.length&&t[0][0]){var r=e.gd,n=e.isActiveShape,a=e.dragmode,o=(r.layout||{}).shapes||[];if(!i(a)&&void 0!==n){var s=r._fullLayout._activeShapeIndex;if(s<o.length)switch(r._fullLayout.shapes[s].type){case"rect":a="drawrect";break;case"circle":a="drawcircle";break;case"line":a="drawline";break;case"path":var l=o[s].path||"";a="Z"===l[l.length-1]?"drawclosedpath":"drawopenpath"}}var u=w(t,e,a);g(r);for(var c=e.editHelpers,f=(c||{}).modifyItem,h=[],p=0;p<o.length;p++){var d=r._fullLayout.shapes[p];if(h[p]=d._input,void 0!==n&&p===r._fullLayout._activeShapeIndex){var v=u;switch(d.type){case"line":case"rect":case"circle":f("x0",v.x0),f("x1",v.x1),f("y0",v.y0),f("y1",v.y1);break;case"path":f("path",v.path)}}}return void 0===n?(h.push(u),h):c?c.getUpdateObj():{}}},createShapeObj:w}},1936:function(t){"use strict";t.exports={clearOutlineControllers:function(t){var e=t._fullLayout._zoomlayer;e&&e.selectAll(".outline-controllers").remove()},clearOutline:function(t){var e=t._fullLayout._zoomlayer;e&&e.selectAll(".select-outline").remove(),t._fullLayout._outlining=!1}}},65152:function(t,e,r){"use strict";var n=r(85448),i=r(3400),a=r(54460);e.rangeToShapePosition=function(t){return"log"===t.type?t.r2d:function(t){return t}},e.shapePositionToRange=function(t){return"log"===t.type?t.d2r:function(t){return t}},e.decodeDate=function(t){return function(e){return e.replace&&(e=e.replace("_"," ")),t(e)}},e.encodeDate=function(t){return function(e){return t(e).replace(" ","_")}},e.extractPathCoords=function(t,e,r){var a=[];return t.match(n.segmentRE).forEach((function(t){var o=e[t.charAt(0)].drawn;if(void 0!==o){var s=t.substr(1).match(n.paramRE);if(s&&!(s.length<o)){var l=s[o],u=r?l:i.cleanNumber(l);a.push(u)}}})),a},e.getDataToPixel=function(t,r,n,i){var a,o=t._fullLayout._size;if(r)if("domain"===i)a=function(t){return r._length*(n?1-t:t)+r._offset};else{var s=e.shapePositionToRange(r);a=function(t){return r._offset+r.r2p(s(t,!0))},"date"===r.type&&(a=e.decodeDate(a))}else a=n?function(t){return o.t+o.h*(1-t)}:function(t){return o.l+o.w*t};return a},e.getPixelToData=function(t,r,n,i){var a,o=t._fullLayout._size;if(r)if("domain"===i)a=function(t){var e=(t-r._offset)/r._length;return n?1-e:e};else{var s=e.rangeToShapePosition(r);a=function(t){return s(r.p2r(t-r._offset))}}else a=n?function(t){return 1-(t-o.t)/o.h}:function(t){return(t-o.l)/o.w};return a},e.roundPositionForSharpStrokeRendering=function(t,e){var r=1===Math.round(e%2),n=Math.round(t);return r?n+.5:n},e.makeShapesOptionsAndPlotinfo=function(t,e){var r=t._fullLayout.shapes[e]||{},n=t._fullLayout._plots[r.xref+r.yref];return n?n._hadPlotinfo=!0:(n={},r.xref&&"paper"!==r.xref&&(n.xaxis=t._fullLayout[r.xref+"axis"]),r.yref&&"paper"!==r.yref&&(n.yaxis=t._fullLayout[r.yref+"axis"])),n.xsizemode=r.xsizemode,n.ysizemode=r.ysizemode,n.xanchor=r.xanchor,n.yanchor=r.yanchor,{options:r,plotinfo:n}},e.makeSelectionsOptionsAndPlotinfo=function(t,e){var r=t._fullLayout.selections[e]||{},n=t._fullLayout._plots[r.xref+r.yref];return n?n._hadPlotinfo=!0:(n={},r.xref&&(n.xaxis=t._fullLayout[r.xref+"axis"]),r.yref&&(n.yaxis=t._fullLayout[r.yref+"axis"])),{options:r,plotinfo:n}},e.getPathString=function(t,r){var o,s,l,u,c,f,h,p,d=r.type,v=a.getRefType(r.xref),g=a.getRefType(r.yref),y=a.getFromId(t,r.xref),m=a.getFromId(t,r.yref),x=t._fullLayout._size;if(y?"domain"===v?s=function(t){return y._offset+y._length*t}:(o=e.shapePositionToRange(y),s=function(t){return y._offset+y.r2p(o(t,!0))}):s=function(t){return x.l+x.w*t},m?"domain"===g?u=function(t){return m._offset+m._length*(1-t)}:(l=e.shapePositionToRange(m),u=function(t){return m._offset+m.r2p(l(t,!0))}):u=function(t){return x.t+x.h*(1-t)},"path"===d)return y&&"date"===y.type&&(s=e.decodeDate(s)),m&&"date"===m.type&&(u=e.decodeDate(u)),function(t,e,r){var a=t.path,o=t.xsizemode,s=t.ysizemode,l=t.xanchor,u=t.yanchor;return a.replace(n.segmentRE,(function(t){var a=0,c=t.charAt(0),f=n.paramIsX[c],h=n.paramIsY[c],p=n.numParams[c],d=t.substr(1).replace(n.paramRE,(function(t){return f[a]?t="pixel"===o?e(l)+Number(t):e(t):h[a]&&(t="pixel"===s?r(u)-Number(t):r(t)),++a>p&&(t="X"),t}));return a>p&&(d=d.replace(/[\s,]*X.*/,""),i.log("Ignoring extra params in segment "+t)),c+d}))}(r,s,u);if("pixel"===r.xsizemode){var b=s(r.xanchor);c=b+r.x0,f=b+r.x1}else c=s(r.x0),f=s(r.x1);if("pixel"===r.ysizemode){var _=u(r.yanchor);h=_-r.y0,p=_-r.y1}else h=u(r.y0),p=u(r.y1);if("line"===d)return"M"+c+","+h+"L"+f+","+p;if("rect"===d)return"M"+c+","+h+"H"+f+"V"+p+"H"+c+"Z";var w=(c+f)/2,T=(h+p)/2,k=Math.abs(w-c),A=Math.abs(T-h),M="A"+k+","+A,S=w+k+","+T;return"M"+S+M+" 0 1,1 "+w+","+(T-A)+M+" 0 0,1 "+S+"Z"}},41592:function(t,e,r){"use strict";var n=r(4016);t.exports={moduleType:"component",name:"shapes",layoutAttributes:r(46056),supplyLayoutDefaults:r(43712),supplyDrawNewShapeDefaults:r(65144),includeBasePlot:r(36632)("shapes"),calcAutorange:r(96084),draw:n.draw,drawOne:n.drawOne}},97728:function(t){"use strict";function e(t,e){return e?e.d2l(t):t}function r(t,e){return e?e.l2d(t):t}function n(t,r){return e(t.x1,r)-e(t.x0,r)}function i(t,r,n){return e(t.y1,n)-e(t.y0,n)}t.exports={x0:function(t){return t.x0},x1:function(t){return t.x1},y0:function(t){return t.y0},y1:function(t){return t.y1},slope:function(t,e,r){return"line"!==t.type?void 0:i(t,0,r)/n(t,e)},dx:n,dy:i,width:function(t,e){return Math.abs(n(t,e))},height:function(t,e,r){return Math.abs(i(t,0,r))},length:function(t,e,r){return"line"!==t.type?void 0:Math.sqrt(Math.pow(n(t,e),2)+Math.pow(i(t,0,r),2))},xcenter:function(t,n){return r((e(t.x1,n)+e(t.x0,n))/2,n)},ycenter:function(t,n,i){return r((e(t.y1,i)+e(t.y0,i))/2,i)}}},89861:function(t,e,r){"use strict";var n=r(25376),i=r(66741),a=r(92880).extendDeepAll,o=r(67824).overrideAll,s=r(85656),l=r(31780).templatedArray,u=r(60876),c=l("step",{visible:{valType:"boolean",dflt:!0},method:{valType:"enumerated",values:["restyle","relayout","animate","update","skip"],dflt:"restyle"},args:{valType:"info_array",freeLength:!0,items:[{valType:"any"},{valType:"any"},{valType:"any"}]},label:{valType:"string"},value:{valType:"string"},execute:{valType:"boolean",dflt:!0}});t.exports=o(l("slider",{visible:{valType:"boolean",dflt:!0},active:{valType:"number",min:0,dflt:0},steps:c,lenmode:{valType:"enumerated",values:["fraction","pixels"],dflt:"fraction"},len:{valType:"number",min:0,dflt:1},x:{valType:"number",min:-2,max:3,dflt:0},pad:a(i({editType:"arraydraw"}),{},{t:{dflt:20}}),xanchor:{valType:"enumerated",values:["auto","left","center","right"],dflt:"left"},y:{valType:"number",min:-2,max:3,dflt:0},yanchor:{valType:"enumerated",values:["auto","top","middle","bottom"],dflt:"top"},transition:{duration:{valType:"number",min:0,dflt:150},easing:{valType:"enumerated",values:s.transition.easing.values,dflt:"cubic-in-out"}},currentvalue:{visible:{valType:"boolean",dflt:!0},xanchor:{valType:"enumerated",values:["left","center","right"],dflt:"left"},offset:{valType:"number",dflt:10},prefix:{valType:"string"},suffix:{valType:"string"},font:n({})},font:n({}),activebgcolor:{valType:"color",dflt:u.gripBgActiveColor},bgcolor:{valType:"color",dflt:u.railBgColor},bordercolor:{valType:"color",dflt:u.railBorderColor},borderwidth:{valType:"number",min:0,dflt:u.railBorderWidth},ticklen:{valType:"number",min:0,dflt:u.tickLength},tickcolor:{valType:"color",dflt:u.tickColor},tickwidth:{valType:"number",min:0,dflt:1},minorticklen:{valType:"number",min:0,dflt:u.minorTickLength}}),"arraydraw","from-root")},60876:function(t){"use strict";t.exports={name:"sliders",containerClassName:"slider-container",groupClassName:"slider-group",inputAreaClass:"slider-input-area",railRectClass:"slider-rail-rect",railTouchRectClass:"slider-rail-touch-rect",gripRectClass:"slider-grip-rect",tickRectClass:"slider-tick-rect",inputProxyClass:"slider-input-proxy",labelsClass:"slider-labels",labelGroupClass:"slider-label-group",labelClass:"slider-label",currentValueClass:"slider-current-value",railHeight:5,menuIndexAttrName:"slider-active-index",autoMarginIdRoot:"slider-",minWidth:30,minHeight:30,textPadX:40,arrowOffsetX:4,railRadius:2,railWidth:5,railBorder:4,railBorderWidth:1,railBorderColor:"#bec8d9",railBgColor:"#f8fafc",railInset:8,stepInset:10,gripRadius:10,gripWidth:20,gripHeight:20,gripBorder:20,gripBorderWidth:1,gripBorderColor:"#bec8d9",gripBgColor:"#f6f8fa",gripBgActiveColor:"#dbdde0",labelPadding:8,labelOffset:0,tickWidth:1,tickColor:"#333",tickOffset:25,tickLength:7,minorTickOffset:25,minorTickColor:"#333",minorTickLength:4,currentValuePadding:8,currentValueInset:0}},8132:function(t,e,r){"use strict";var n=r(3400),i=r(51272),a=r(89861),o=r(60876).name,s=a.steps;function l(t,e,r){function o(r,i){return n.coerce(t,e,a,r,i)}for(var s=i(t,e,{name:"steps",handleItemDefaults:u}),l=0,c=0;c<s.length;c++)s[c].visible&&l++;if(l<2?e.visible=!1:o("visible")){e._stepCount=l;var f=e._visibleSteps=n.filterVisible(s);(s[o("active")]||{}).visible||(e.active=f[0]._index),o("x"),o("y"),n.noneOrAll(t,e,["x","y"]),o("xanchor"),o("yanchor"),o("len"),o("lenmode"),o("pad.t"),o("pad.r"),o("pad.b"),o("pad.l"),n.coerceFont(o,"font",r.font),o("currentvalue.visible")&&(o("currentvalue.xanchor"),o("currentvalue.prefix"),o("currentvalue.suffix"),o("currentvalue.offset"),n.coerceFont(o,"currentvalue.font",e.font)),o("transition.duration"),o("transition.easing"),o("bgcolor"),o("activebgcolor"),o("bordercolor"),o("borderwidth"),o("ticklen"),o("tickwidth"),o("tickcolor"),o("minorticklen")}}function u(t,e){function r(r,i){return n.coerce(t,e,s,r,i)}if("skip"===t.method||Array.isArray(t.args)?r("visible"):e.visible=!1){r("method"),r("args");var i=r("label","step-"+e._index);r("value",i),r("execute")}}t.exports=function(t,e){i(t,e,{name:o,handleItemDefaults:l})}},79664:function(t,e,r){"use strict";var n=r(33428),i=r(7316),a=r(76308),o=r(43616),s=r(3400),l=s.strTranslate,u=r(72736),c=r(31780).arrayEditor,f=r(60876),h=r(84284),p=h.LINE_SPACING,d=h.FROM_TL,v=h.FROM_BR;function g(t){return f.autoMarginIdRoot+t._index}function y(t){return t._index}function m(t,e){var r=o.tester.selectAll("g."+f.labelGroupClass).data(e._visibleSteps);r.enter().append("g").classed(f.labelGroupClass,!0);var a=0,l=0;r.each((function(t){var r=_(n.select(this),{step:t},e).node();if(r){var i=o.bBox(r);l=Math.max(l,i.height),a=Math.max(a,i.width)}})),r.remove();var c=e._dims={};c.inputAreaWidth=Math.max(f.railWidth,f.gripHeight);var h=t._fullLayout._size;c.lx=h.l+h.w*e.x,c.ly=h.t+h.h*(1-e.y),"fraction"===e.lenmode?c.outerLength=Math.round(h.w*e.len):c.outerLength=e.len,c.inputAreaStart=0,c.inputAreaLength=Math.round(c.outerLength-e.pad.l-e.pad.r);var p=(c.inputAreaLength-2*f.stepInset)/(e._stepCount-1),y=a+f.labelPadding;if(c.labelStride=Math.max(1,Math.ceil(y/p)),c.labelHeight=l,c.currentValueMaxWidth=0,c.currentValueHeight=0,c.currentValueTotalHeight=0,c.currentValueMaxLines=1,e.currentvalue.visible){var m=o.tester.append("g");r.each((function(t){var r=x(m,e,t.label),n=r.node()&&o.bBox(r.node())||{width:0,height:0},i=u.lineCount(r);c.currentValueMaxWidth=Math.max(c.currentValueMaxWidth,Math.ceil(n.width)),c.currentValueHeight=Math.max(c.currentValueHeight,Math.ceil(n.height)),c.currentValueMaxLines=Math.max(c.currentValueMaxLines,i)})),c.currentValueTotalHeight=c.currentValueHeight+e.currentvalue.offset,m.remove()}c.height=c.currentValueTotalHeight+f.tickOffset+e.ticklen+f.labelOffset+c.labelHeight+e.pad.t+e.pad.b;var b="left";s.isRightAnchor(e)&&(c.lx-=c.outerLength,b="right"),s.isCenterAnchor(e)&&(c.lx-=c.outerLength/2,b="center");var w="top";s.isBottomAnchor(e)&&(c.ly-=c.height,w="bottom"),s.isMiddleAnchor(e)&&(c.ly-=c.height/2,w="middle"),c.outerLength=Math.ceil(c.outerLength),c.height=Math.ceil(c.height),c.lx=Math.round(c.lx),c.ly=Math.round(c.ly);var T={y:e.y,b:c.height*v[w],t:c.height*d[w]};"fraction"===e.lenmode?(T.l=0,T.xl=e.x-e.len*d[b],T.r=0,T.xr=e.x+e.len*v[b]):(T.x=e.x,T.l=c.outerLength*d[b],T.r=c.outerLength*v[b]),i.autoMargin(t,g(e),T)}function x(t,e,r){if(e.currentvalue.visible){var n,i,a=e._dims;switch(e.currentvalue.xanchor){case"right":n=a.inputAreaLength-f.currentValueInset-a.currentValueMaxWidth,i="left";break;case"center":n=.5*a.inputAreaLength,i="middle";break;default:n=f.currentValueInset,i="left"}var l=s.ensureSingle(t,"text",f.labelClass,(function(t){t.attr({"text-anchor":i,"data-notex":1})})),c=e.currentvalue.prefix?e.currentvalue.prefix:"";if("string"==typeof r)c+=r;else{var h=e.steps[e.active].label,d=e._gd._fullLayout._meta;d&&(h=s.templateString(h,d)),c+=h}e.currentvalue.suffix&&(c+=e.currentvalue.suffix),l.call(o.font,e.currentvalue.font).text(c).call(u.convertToTspans,e._gd);var v=u.lineCount(l),g=(a.currentValueMaxLines+1-v)*e.currentvalue.font.size*p;return u.positionText(l,n,g),l}}function b(t,e,r){s.ensureSingle(t,"rect",f.gripRectClass,(function(n){n.call(A,e,t,r).style("pointer-events","all")})).attr({width:f.gripWidth,height:f.gripHeight,rx:f.gripRadius,ry:f.gripRadius}).call(a.stroke,r.bordercolor).call(a.fill,r.bgcolor).style("stroke-width",r.borderwidth+"px")}function _(t,e,r){var n=s.ensureSingle(t,"text",f.labelClass,(function(t){t.attr({"text-anchor":"middle","data-notex":1})})),i=e.step.label,a=r._gd._fullLayout._meta;return a&&(i=s.templateString(i,a)),n.call(o.font,r.font).text(i).call(u.convertToTspans,r._gd),n}function w(t,e){var r=s.ensureSingle(t,"g",f.labelsClass),i=e._dims,a=r.selectAll("g."+f.labelGroupClass).data(i.labelSteps);a.enter().append("g").classed(f.labelGroupClass,!0),a.exit().remove(),a.each((function(t){var r=n.select(this);r.call(_,t,e),o.setTranslate(r,E(e,t.fraction),f.tickOffset+e.ticklen+e.font.size*p+f.labelOffset+i.currentValueTotalHeight)}))}function T(t,e,r,n,i){var a=Math.round(n*(r._stepCount-1)),o=r._visibleSteps[a]._index;o!==r.active&&k(t,e,r,o,!0,i)}function k(t,e,r,n,a,o){var s=r.active;r.active=n,c(t.layout,f.name,r).applyUpdate("active",n);var l=r.steps[r.active];e.call(S,r,o),e.call(x,r),t.emit("plotly_sliderchange",{slider:r,step:r.steps[r.active],interaction:a,previousActive:s}),l&&l.method&&a&&(e._nextMethod?(e._nextMethod.step=l,e._nextMethod.doCallback=a,e._nextMethod.doTransition=o):(e._nextMethod={step:l,doCallback:a,doTransition:o},e._nextMethodRaf=window.requestAnimationFrame((function(){var r=e._nextMethod.step;r.method&&(r.execute&&i.executeAPICommand(t,r.method,r.args),e._nextMethod=null,e._nextMethodRaf=null)}))))}function A(t,e,r){if(!e._context.staticPlot){var i=r.node(),o=n.select(e);t.on("mousedown",l),t.on("touchstart",l)}function s(){return r.data()[0]}function l(){var t=s();e.emit("plotly_sliderstart",{slider:t});var l=r.select("."+f.gripRectClass);n.event.stopPropagation(),n.event.preventDefault(),l.call(a.fill,t.activebgcolor);var u=L(t,n.mouse(i)[0]);function c(){var t=s(),a=L(t,n.mouse(i)[0]);T(e,r,t,a,!1)}function h(){var t=s();t._dragging=!1,l.call(a.fill,t.bgcolor),o.on("mouseup",null),o.on("mousemove",null),o.on("touchend",null),o.on("touchmove",null),e.emit("plotly_sliderend",{slider:t,step:t.steps[t.active]})}T(e,r,t,u,!0),t._dragging=!0,o.on("mousemove",c),o.on("touchmove",c),o.on("mouseup",h),o.on("touchend",h)}}function M(t,e){var r=t.selectAll("rect."+f.tickRectClass).data(e._visibleSteps),i=e._dims;r.enter().append("rect").classed(f.tickRectClass,!0),r.exit().remove(),r.attr({width:e.tickwidth+"px","shape-rendering":"crispEdges"}),r.each((function(t,r){var s=r%i.labelStride==0,l=n.select(this);l.attr({height:s?e.ticklen:e.minorticklen}).call(a.fill,e.tickcolor),o.setTranslate(l,E(e,r/(e._stepCount-1))-.5*e.tickwidth,(s?f.tickOffset:f.minorTickOffset)+i.currentValueTotalHeight)}))}function S(t,e,r){for(var n=t.select("rect."+f.gripRectClass),i=0,a=0;a<e._stepCount;a++)if(e._visibleSteps[a]._index===e.active){i=a;break}var o=E(e,i/(e._stepCount-1));if(!e._invokingCommand){var s=n;r&&e.transition.duration>0&&(s=s.transition().duration(e.transition.duration).ease(e.transition.easing)),s.attr("transform",l(o-.5*f.gripWidth,e._dims.currentValueTotalHeight))}}function E(t,e){var r=t._dims;return r.inputAreaStart+f.stepInset+(r.inputAreaLength-2*f.stepInset)*Math.min(1,Math.max(0,e))}function L(t,e){var r=t._dims;return Math.min(1,Math.max(0,(e-f.stepInset-r.inputAreaStart)/(r.inputAreaLength-2*f.stepInset-2*r.inputAreaStart)))}function C(t,e,r){var n=r._dims,i=s.ensureSingle(t,"rect",f.railTouchRectClass,(function(n){n.call(A,e,t,r).style("pointer-events","all")}));i.attr({width:n.inputAreaLength,height:Math.max(n.inputAreaWidth,f.tickOffset+r.ticklen+n.labelHeight)}).call(a.fill,r.bgcolor).attr("opacity",0),o.setTranslate(i,0,n.currentValueTotalHeight)}function P(t,e){var r=e._dims,n=r.inputAreaLength-2*f.railInset,i=s.ensureSingle(t,"rect",f.railRectClass);i.attr({width:n,height:f.railWidth,rx:f.railRadius,ry:f.railRadius,"shape-rendering":"crispEdges"}).call(a.stroke,e.bordercolor).call(a.fill,e.bgcolor).style("stroke-width",e.borderwidth+"px"),o.setTranslate(i,f.railInset,.5*(r.inputAreaWidth-f.railWidth)+r.currentValueTotalHeight)}t.exports=function(t){var e=t._context.staticPlot,r=t._fullLayout,a=function(t,e){for(var r=t[f.name],n=[],i=0;i<r.length;i++){var a=r[i];a.visible&&(a._gd=e,n.push(a))}return n}(r,t),s=r._infolayer.selectAll("g."+f.containerClassName).data(a.length>0?[0]:[]);function l(e){e._commandObserver&&(e._commandObserver.remove(),delete e._commandObserver),i.autoMargin(t,g(e))}if(s.enter().append("g").classed(f.containerClassName,!0).style("cursor",e?null:"ew-resize"),s.exit().each((function(){n.select(this).selectAll("g."+f.groupClassName).each(l)})).remove(),0!==a.length){var u=s.selectAll("g."+f.groupClassName).data(a,y);u.enter().append("g").classed(f.groupClassName,!0),u.exit().each(l).remove();for(var c=0;c<a.length;c++){var h=a[c];m(t,h)}u.each((function(e){var r=n.select(this);!function(t){var e=t._dims;e.labelSteps=[];for(var r=t._stepCount,n=0;n<r;n+=e.labelStride)e.labelSteps.push({fraction:n/(r-1),step:t._visibleSteps[n]})}(e),i.manageCommandObserver(t,e,e._visibleSteps,(function(e){var n=r.data()[0];n.active!==e.index&&(n._dragging||k(t,r,n,e.index,!1,!0))})),function(t,e,r){(r.steps[r.active]||{}).visible||(r.active=r._visibleSteps[0]._index),e.call(x,r).call(P,r).call(w,r).call(M,r).call(C,t,r).call(b,t,r);var n=r._dims;o.setTranslate(e,n.lx+r.pad.l,n.ly+r.pad.t),e.call(S,r,!1),e.call(x,r)}(t,n.select(this),e)}))}}},97544:function(t,e,r){"use strict";var n=r(60876);t.exports={moduleType:"component",name:n.name,layoutAttributes:r(89861),supplyLayoutDefaults:r(8132),draw:r(79664)}},81668:function(t,e,r){"use strict";var n=r(33428),i=r(38248),a=r(7316),o=r(24040),s=r(3400),l=s.strTranslate,u=r(43616),c=r(76308),f=r(72736),h=r(13448),p=r(84284).OPPOSITE_SIDE,d=/ [XY][0-9]* /;t.exports={draw:function(t,e,r){var v,g=r.propContainer,y=r.propName,m=r.placeholder,x=r.traceIndex,b=r.avoid||{},_=r.attributes,w=r.transform,T=r.containerGroup,k=t._fullLayout,A=1,M=!1,S=g.title,E=(S&&S.text?S.text:"").trim(),L=S&&S.font?S.font:{},C=L.family,P=L.size,O=L.color,I=L.weight,z=L.style,D=L.variant;"title.text"===y?v="titleText":-1!==y.indexOf("axis")?v="axisTitleText":y.indexOf(!0)&&(v="colorbarTitleText");var R=t._context.edits[v];""===E?A=0:E.replace(d," % ")===m.replace(d," % ")&&(A=.2,M=!0,R||(E="")),r._meta?E=s.templateString(E,r._meta):k._meta&&(E=s.templateString(E,k._meta));var F,B=E||R;T||(T=s.ensureSingle(k._infolayer,"g","g-"+e),F=k._hColorbarMoveTitle);var N=T.selectAll("text").data(B?[0]:[]);if(N.enter().append("text"),N.text(E).attr("class",e),N.exit().remove(),!B)return T;function j(t){s.syncOrAsync([U,V],t)}function U(e){var r;return!w&&F&&(w={}),w?(r="",w.rotate&&(r+="rotate("+[w.rotate,_.x,_.y]+")"),(w.offset||F)&&(r+=l(0,(w.offset||0)-(F||0)))):r=null,e.attr("transform",r),e.style({"font-family":C,"font-size":n.round(P,2)+"px",fill:c.rgb(O),opacity:A*c.opacity(O),"font-weight":I,"font-style":z,"font-variant":D}).attr(_).call(f.convertToTspans,t),a.previousPromises(t)}function V(e){var r=n.select(e.node().parentNode);if(b&&b.selection&&b.side&&E){r.attr("transform",null);var a=p[b.side],o="left"===b.side||"top"===b.side?-1:1,c=i(b.pad)?b.pad:2,f=u.bBox(r.node()),h={t:0,b:0,l:0,r:0},d=t._fullLayout._reservedMargin;for(var v in d)for(var y in d[v]){var m=d[v][y];h[y]=Math.max(h[y],m)}var x={left:h.l,top:h.t,right:k.width-h.r,bottom:k.height-h.b},_=b.maxShift||o*(x[b.side]-f[b.side]),w=0;if(_<0)w=_;else{var T=b.offsetLeft||0,A=b.offsetTop||0;f.left-=T,f.right-=T,f.top-=A,f.bottom-=A,b.selection.each((function(){var t=u.bBox(this);s.bBoxIntersect(f,t,c)&&(w=Math.max(w,o*(t[b.side]-f[a])+c))})),w=Math.min(_,w),g._titleScoot=Math.abs(w)}if(w>0||_<0){var M={left:[-w,0],right:[w,0],top:[0,-w],bottom:[0,w]}[b.side];r.attr("transform",l(M[0],M[1]))}}}return N.call(j),R&&(E?N.on(".opacity",null):(A=0,M=!0,N.text(m).on("mouseover.opacity",(function(){n.select(this).transition().duration(h.SHOW_PLACEHOLDER).style("opacity",1)})).on("mouseout.opacity",(function(){n.select(this).transition().duration(h.HIDE_PLACEHOLDER).style("opacity",0)}))),N.call(f.makeEditable,{gd:t}).on("edit",(function(e){void 0!==x?o.call("_guiRestyle",t,y,e,x):o.call("_guiRelayout",t,y,e)})).on("cancel",(function(){this.text(this.attr("data-unformatted")).call(j)})).on("input",(function(t){this.text(t||" ").call(f.positionText,_.x,_.y)}))),N.classed("js-placeholder",M),T}}},88444:function(t,e,r){"use strict";var n=r(25376),i=r(22548),a=r(92880).extendFlat,o=r(67824).overrideAll,s=r(66741),l=r(31780).templatedArray,u=l("button",{visible:{valType:"boolean"},method:{valType:"enumerated",values:["restyle","relayout","animate","update","skip"],dflt:"restyle"},args:{valType:"info_array",freeLength:!0,items:[{valType:"any"},{valType:"any"},{valType:"any"}]},args2:{valType:"info_array",freeLength:!0,items:[{valType:"any"},{valType:"any"},{valType:"any"}]},label:{valType:"string",dflt:""},execute:{valType:"boolean",dflt:!0}});t.exports=o(l("updatemenu",{_arrayAttrRegexps:[/^updatemenus\[(0|[1-9][0-9]+)\]\.buttons/],visible:{valType:"boolean"},type:{valType:"enumerated",values:["dropdown","buttons"],dflt:"dropdown"},direction:{valType:"enumerated",values:["left","right","up","down"],dflt:"down"},active:{valType:"integer",min:-1,dflt:0},showactive:{valType:"boolean",dflt:!0},buttons:u,x:{valType:"number",min:-2,max:3,dflt:-.05},xanchor:{valType:"enumerated",values:["auto","left","center","right"],dflt:"right"},y:{valType:"number",min:-2,max:3,dflt:1},yanchor:{valType:"enumerated",values:["auto","top","middle","bottom"],dflt:"top"},pad:a(s({editType:"arraydraw"}),{}),font:n({}),bgcolor:{valType:"color"},bordercolor:{valType:"color",dflt:i.borderLine},borderwidth:{valType:"number",min:0,dflt:1,editType:"arraydraw"}}),"arraydraw","from-root")},73712:function(t){"use strict";t.exports={name:"updatemenus",containerClassName:"updatemenu-container",headerGroupClassName:"updatemenu-header-group",headerClassName:"updatemenu-header",headerArrowClassName:"updatemenu-header-arrow",dropdownButtonGroupClassName:"updatemenu-dropdown-button-group",dropdownButtonClassName:"updatemenu-dropdown-button",buttonClassName:"updatemenu-button",itemRectClassName:"updatemenu-item-rect",itemTextClassName:"updatemenu-item-text",menuIndexAttrName:"updatemenu-active-index",autoMarginIdRoot:"updatemenu-",blankHeaderOpts:{label:"  "},minWidth:30,minHeight:30,textPadX:24,arrowPadX:16,rx:2,ry:2,textOffsetX:12,textOffsetY:3,arrowOffsetX:4,gapButtonHeader:5,gapButton:2,activeColor:"#F4FAFF",hoverColor:"#F4FAFF",arrowSymbol:{left:"◄",right:"►",up:"▲",down:"▼"}}},91384:function(t,e,r){"use strict";var n=r(3400),i=r(51272),a=r(88444),o=r(73712).name,s=a.buttons;function l(t,e,r){function o(r,i){return n.coerce(t,e,a,r,i)}o("visible",i(t,e,{name:"buttons",handleItemDefaults:u}).length>0)&&(o("active"),o("direction"),o("type"),o("showactive"),o("x"),o("y"),n.noneOrAll(t,e,["x","y"]),o("xanchor"),o("yanchor"),o("pad.t"),o("pad.r"),o("pad.b"),o("pad.l"),n.coerceFont(o,"font",r.font),o("bgcolor",r.paper_bgcolor),o("bordercolor"),o("borderwidth"))}function u(t,e){function r(r,i){return n.coerce(t,e,s,r,i)}r("visible","skip"===t.method||Array.isArray(t.args))&&(r("method"),r("args"),r("args2"),r("label"),r("execute"))}t.exports=function(t,e){i(t,e,{name:o,handleItemDefaults:l})}},14420:function(t,e,r){"use strict";var n=r(33428),i=r(7316),a=r(76308),o=r(43616),s=r(3400),l=r(72736),u=r(31780).arrayEditor,c=r(84284).LINE_SPACING,f=r(73712),h=r(37400);function p(t){return t._index}function d(t,e){return+t.attr(f.menuIndexAttrName)===e._index}function v(t,e,r,n,i,a,o,s){e.active=o,u(t.layout,f.name,e).applyUpdate("active",o),"buttons"===e.type?y(t,n,null,null,e):"dropdown"===e.type&&(i.attr(f.menuIndexAttrName,"-1"),g(t,n,i,a,e),s||y(t,n,i,a,e))}function g(t,e,r,n,i){var a=s.ensureSingle(e,"g",f.headerClassName,(function(t){t.style("pointer-events","all")})),l=i._dims,u=i.active,c=i.buttons[u]||f.blankHeaderOpts,h={y:i.pad.t,yPad:0,x:i.pad.l,xPad:0,index:0},p={width:l.headerWidth,height:l.headerHeight};a.call(m,i,c,t).call(M,i,h,p),s.ensureSingle(e,"text",f.headerArrowClassName,(function(t){t.attr("text-anchor","end").call(o.font,i.font).text(f.arrowSymbol[i.direction])})).attr({x:l.headerWidth-f.arrowOffsetX+i.pad.l,y:l.headerHeight/2+f.textOffsetY+i.pad.t}),a.on("click",(function(){r.call(S,String(d(r,i)?-1:i._index)),y(t,e,r,n,i)})),a.on("mouseover",(function(){a.call(w)})),a.on("mouseout",(function(){a.call(T,i)})),o.setTranslate(e,l.lx,l.ly)}function y(t,e,r,a,o){r||(r=e).attr("pointer-events","all");var l=function(t){return-1==+t.attr(f.menuIndexAttrName)}(r)&&"buttons"!==o.type?[]:o.buttons,u="dropdown"===o.type?f.dropdownButtonClassName:f.buttonClassName,c=r.selectAll("g."+u).data(s.filterVisible(l)),h=c.enter().append("g").classed(u,!0),p=c.exit();"dropdown"===o.type?(h.attr("opacity","0").transition().attr("opacity","1"),p.transition().attr("opacity","0").remove()):p.remove();var d=0,g=0,y=o._dims,x=-1!==["up","down"].indexOf(o.direction);"dropdown"===o.type&&(x?g=y.headerHeight+f.gapButtonHeader:d=y.headerWidth+f.gapButtonHeader),"dropdown"===o.type&&"up"===o.direction&&(g=-f.gapButtonHeader+f.gapButton-y.openHeight),"dropdown"===o.type&&"left"===o.direction&&(d=-f.gapButtonHeader+f.gapButton-y.openWidth);var b={x:y.lx+d+o.pad.l,y:y.ly+g+o.pad.t,yPad:f.gapButton,xPad:f.gapButton,index:0},k={l:b.x+o.borderwidth,t:b.y+o.borderwidth};c.each((function(s,l){var u=n.select(this);u.call(m,o,s,t).call(M,o,b),u.on("click",(function(){n.event.defaultPrevented||(s.execute&&(s.args2&&o.active===l?(v(t,o,0,e,r,a,-1),i.executeAPICommand(t,s.method,s.args2)):(v(t,o,0,e,r,a,l),i.executeAPICommand(t,s.method,s.args))),t.emit("plotly_buttonclicked",{menu:o,button:s,active:o.active}))})),u.on("mouseover",(function(){u.call(w)})),u.on("mouseout",(function(){u.call(T,o),c.call(_,o)}))})),c.call(_,o),x?(k.w=Math.max(y.openWidth,y.headerWidth),k.h=b.y-k.t):(k.w=b.x-k.l,k.h=Math.max(y.openHeight,y.headerHeight)),k.direction=o.direction,a&&(c.size()?function(t,e,r,n,i,a){var o,s,l,u=i.direction,c="up"===u||"down"===u,h=i._dims,p=i.active;if(c)for(s=0,l=0;l<p;l++)s+=h.heights[l]+f.gapButton;else for(o=0,l=0;l<p;l++)o+=h.widths[l]+f.gapButton;n.enable(a,o,s),n.hbar&&n.hbar.attr("opacity","0").transition().attr("opacity","1"),n.vbar&&n.vbar.attr("opacity","0").transition().attr("opacity","1")}(0,0,0,a,o,k):function(t){var e=!!t.hbar,r=!!t.vbar;e&&t.hbar.transition().attr("opacity","0").each("end",(function(){e=!1,r||t.disable()})),r&&t.vbar.transition().attr("opacity","0").each("end",(function(){r=!1,e||t.disable()}))}(a))}function m(t,e,r,n){t.call(x,e).call(b,e,r,n)}function x(t,e){s.ensureSingle(t,"rect",f.itemRectClassName,(function(t){t.attr({rx:f.rx,ry:f.ry,"shape-rendering":"crispEdges"})})).call(a.stroke,e.bordercolor).call(a.fill,e.bgcolor).style("stroke-width",e.borderwidth+"px")}function b(t,e,r,n){var i=s.ensureSingle(t,"text",f.itemTextClassName,(function(t){t.attr({"text-anchor":"start","data-notex":1})})),a=r.label,u=n._fullLayout._meta;u&&(a=s.templateString(a,u)),i.call(o.font,e.font).text(a).call(l.convertToTspans,n)}function _(t,e){var r=e.active;t.each((function(t,i){var o=n.select(this);i===r&&e.showactive&&o.select("rect."+f.itemRectClassName).call(a.fill,f.activeColor)}))}function w(t){t.select("rect."+f.itemRectClassName).call(a.fill,f.hoverColor)}function T(t,e){t.select("rect."+f.itemRectClassName).call(a.fill,e.bgcolor)}function k(t,e){var r=e._dims={width1:0,height1:0,heights:[],widths:[],totalWidth:0,totalHeight:0,openWidth:0,openHeight:0,lx:0,ly:0},a=o.tester.selectAll("g."+f.dropdownButtonClassName).data(s.filterVisible(e.buttons));a.enter().append("g").classed(f.dropdownButtonClassName,!0);var u=-1!==["up","down"].indexOf(e.direction);a.each((function(i,a){var s=n.select(this);s.call(m,e,i,t);var h=s.select("."+f.itemTextClassName),p=h.node()&&o.bBox(h.node()).width,d=Math.max(p+f.textPadX,f.minWidth),v=e.font.size*c,g=l.lineCount(h),y=Math.max(v*g,f.minHeight)+f.textOffsetY;y=Math.ceil(y),d=Math.ceil(d),r.widths[a]=d,r.heights[a]=y,r.height1=Math.max(r.height1,y),r.width1=Math.max(r.width1,d),u?(r.totalWidth=Math.max(r.totalWidth,d),r.openWidth=r.totalWidth,r.totalHeight+=y+f.gapButton,r.openHeight+=y+f.gapButton):(r.totalWidth+=d+f.gapButton,r.openWidth+=d+f.gapButton,r.totalHeight=Math.max(r.totalHeight,y),r.openHeight=r.totalHeight)})),u?r.totalHeight-=f.gapButton:r.totalWidth-=f.gapButton,r.headerWidth=r.width1+f.arrowPadX,r.headerHeight=r.height1,"dropdown"===e.type&&(u?(r.width1+=f.arrowPadX,r.totalHeight=r.height1):r.totalWidth=r.width1,r.totalWidth+=f.arrowPadX),a.remove();var h=r.totalWidth+e.pad.l+e.pad.r,p=r.totalHeight+e.pad.t+e.pad.b,d=t._fullLayout._size;r.lx=d.l+d.w*e.x,r.ly=d.t+d.h*(1-e.y);var v="left";s.isRightAnchor(e)&&(r.lx-=h,v="right"),s.isCenterAnchor(e)&&(r.lx-=h/2,v="center");var g="top";s.isBottomAnchor(e)&&(r.ly-=p,g="bottom"),s.isMiddleAnchor(e)&&(r.ly-=p/2,g="middle"),r.totalWidth=Math.ceil(r.totalWidth),r.totalHeight=Math.ceil(r.totalHeight),r.lx=Math.round(r.lx),r.ly=Math.round(r.ly),i.autoMargin(t,A(e),{x:e.x,y:e.y,l:h*({right:1,center:.5}[v]||0),r:h*({left:1,center:.5}[v]||0),b:p*({top:1,middle:.5}[g]||0),t:p*({bottom:1,middle:.5}[g]||0)})}function A(t){return f.autoMarginIdRoot+t._index}function M(t,e,r,n){n=n||{};var i=t.select("."+f.itemRectClassName),a=t.select("."+f.itemTextClassName),s=e.borderwidth,u=r.index,h=e._dims;o.setTranslate(t,s+r.x,s+r.y);var p=-1!==["up","down"].indexOf(e.direction),d=n.height||(p?h.heights[u]:h.height1);i.attr({x:0,y:0,width:n.width||(p?h.width1:h.widths[u]),height:d});var v=e.font.size*c,g=(l.lineCount(a)-1)*v/2;l.positionText(a,f.textOffsetX,d/2-g+f.textOffsetY),p?r.y+=h.heights[u]+r.yPad:r.x+=h.widths[u]+r.xPad,r.index++}function S(t,e){t.attr(f.menuIndexAttrName,e||"-1").selectAll("g."+f.dropdownButtonClassName).remove()}t.exports=function(t){var e=t._fullLayout,r=s.filterVisible(e[f.name]);function a(e){i.autoMargin(t,A(e))}var o=e._menulayer.selectAll("g."+f.containerClassName).data(r.length>0?[0]:[]);if(o.enter().append("g").classed(f.containerClassName,!0).style("cursor","pointer"),o.exit().each((function(){n.select(this).selectAll("g."+f.headerGroupClassName).each(a)})).remove(),0!==r.length){var l=o.selectAll("g."+f.headerGroupClassName).data(r,p);l.enter().append("g").classed(f.headerGroupClassName,!0);for(var u=s.ensureSingle(o,"g",f.dropdownButtonGroupClassName,(function(t){t.style("pointer-events","all")})),c=0;c<r.length;c++){var m=r[c];k(t,m)}var x="updatemenus"+e._uid,b=new h(t,u,x);l.enter().size()&&(u.node().parentNode.appendChild(u.node()),u.call(S)),l.exit().each((function(t){u.call(S),a(t)})).remove(),l.each((function(e){var r=n.select(this),a="dropdown"===e.type?u:null;i.manageCommandObserver(t,e,e.buttons,(function(n){v(t,e,e.buttons[n.index],r,a,b,n.index,!0)})),"dropdown"===e.type?(g(t,r,u,b,e),d(u,e)&&y(t,r,u,b,e)):y(t,r,null,null,e)}))}}},76908:function(t,e,r){"use strict";var n=r(73712);t.exports={moduleType:"component",name:n.name,layoutAttributes:r(88444),supplyLayoutDefaults:r(91384),draw:r(14420)}},37400:function(t,e,r){"use strict";t.exports=s;var n=r(33428),i=r(76308),a=r(43616),o=r(3400);function s(t,e,r){this.gd=t,this.container=e,this.id=r,this.position=null,this.translateX=null,this.translateY=null,this.hbar=null,this.vbar=null,this.bg=this.container.selectAll("rect.scrollbox-bg").data([0]),this.bg.exit().on(".drag",null).on("wheel",null).remove(),this.bg.enter().append("rect").classed("scrollbox-bg",!0).style("pointer-events","all").attr({opacity:0,x:0,y:0,width:0,height:0})}s.barWidth=2,s.barLength=20,s.barRadius=2,s.barPad=1,s.barColor="#808BA4",s.prototype.enable=function(t,e,r){var o=this.gd._fullLayout,l=o.width,u=o.height;this.position=t;var c,f,h,p,d=this.position.l,v=this.position.w,g=this.position.t,y=this.position.h,m=this.position.direction,x="down"===m,b="left"===m,_="up"===m,w=v,T=y;x||b||"right"===m||_||(this.position.direction="down",x=!0),x||_?(f=(c=d)+w,x?(h=g,T=(p=Math.min(h+T,u))-h):T=(p=g+T)-(h=Math.max(p-T,0))):(p=(h=g)+T,b?w=(f=d+w)-(c=Math.max(f-w,0)):(c=d,w=(f=Math.min(c+w,l))-c)),this._box={l:c,t:h,w:w,h:T};var k=v>w,A=s.barLength+2*s.barPad,M=s.barWidth+2*s.barPad,S=d,E=g+y;E+M>u&&(E=u-M);var L=this.container.selectAll("rect.scrollbar-horizontal").data(k?[0]:[]);L.exit().on(".drag",null).remove(),L.enter().append("rect").classed("scrollbar-horizontal",!0).call(i.fill,s.barColor),k?(this.hbar=L.attr({rx:s.barRadius,ry:s.barRadius,x:S,y:E,width:A,height:M}),this._hbarXMin=S+A/2,this._hbarTranslateMax=w-A):(delete this.hbar,delete this._hbarXMin,delete this._hbarTranslateMax);var C=y>T,P=s.barWidth+2*s.barPad,O=s.barLength+2*s.barPad,I=d+v,z=g;I+P>l&&(I=l-P);var D=this.container.selectAll("rect.scrollbar-vertical").data(C?[0]:[]);D.exit().on(".drag",null).remove(),D.enter().append("rect").classed("scrollbar-vertical",!0).call(i.fill,s.barColor),C?(this.vbar=D.attr({rx:s.barRadius,ry:s.barRadius,x:I,y:z,width:P,height:O}),this._vbarYMin=z+O/2,this._vbarTranslateMax=T-O):(delete this.vbar,delete this._vbarYMin,delete this._vbarTranslateMax);var R=this.id,F=c-.5,B=C?f+P+.5:f+.5,N=h-.5,j=k?p+M+.5:p+.5,U=o._topdefs.selectAll("#"+R).data(k||C?[0]:[]);if(U.exit().remove(),U.enter().append("clipPath").attr("id",R).append("rect"),k||C?(this._clipRect=U.select("rect").attr({x:Math.floor(F),y:Math.floor(N),width:Math.ceil(B)-Math.floor(F),height:Math.ceil(j)-Math.floor(N)}),this.container.call(a.setClipUrl,R,this.gd),this.bg.attr({x:d,y:g,width:v,height:y})):(this.bg.attr({width:0,height:0}),this.container.on("wheel",null).on(".drag",null).call(a.setClipUrl,null),delete this._clipRect),k||C){var V=n.behavior.drag().on("dragstart",(function(){n.event.sourceEvent.preventDefault()})).on("drag",this._onBoxDrag.bind(this));this.container.on("wheel",null).on("wheel",this._onBoxWheel.bind(this)).on(".drag",null).call(V);var q=n.behavior.drag().on("dragstart",(function(){n.event.sourceEvent.preventDefault(),n.event.sourceEvent.stopPropagation()})).on("drag",this._onBarDrag.bind(this));k&&this.hbar.on(".drag",null).call(q),C&&this.vbar.on(".drag",null).call(q)}this.setTranslate(e,r)},s.prototype.disable=function(){(this.hbar||this.vbar)&&(this.bg.attr({width:0,height:0}),this.container.on("wheel",null).on(".drag",null).call(a.setClipUrl,null),delete this._clipRect),this.hbar&&(this.hbar.on(".drag",null),this.hbar.remove(),delete this.hbar,delete this._hbarXMin,delete this._hbarTranslateMax),this.vbar&&(this.vbar.on(".drag",null),this.vbar.remove(),delete this.vbar,delete this._vbarYMin,delete this._vbarTranslateMax)},s.prototype._onBoxDrag=function(){var t=this.translateX,e=this.translateY;this.hbar&&(t-=n.event.dx),this.vbar&&(e-=n.event.dy),this.setTranslate(t,e)},s.prototype._onBoxWheel=function(){var t=this.translateX,e=this.translateY;this.hbar&&(t+=n.event.deltaY),this.vbar&&(e+=n.event.deltaY),this.setTranslate(t,e)},s.prototype._onBarDrag=function(){var t=this.translateX,e=this.translateY;if(this.hbar){var r=t+this._hbarXMin,i=r+this._hbarTranslateMax;t=(o.constrain(n.event.x,r,i)-r)/(i-r)*(this.position.w-this._box.w)}if(this.vbar){var a=e+this._vbarYMin,s=a+this._vbarTranslateMax;e=(o.constrain(n.event.y,a,s)-a)/(s-a)*(this.position.h-this._box.h)}this.setTranslate(t,e)},s.prototype.setTranslate=function(t,e){var r=this.position.w-this._box.w,n=this.position.h-this._box.h;if(t=o.constrain(t||0,0,r),e=o.constrain(e||0,0,n),this.translateX=t,this.translateY=e,this.container.call(a.setTranslate,this._box.l-this.position.l-t,this._box.t-this.position.t-e),this._clipRect&&this._clipRect.attr({x:Math.floor(this.position.l+t-.5),y:Math.floor(this.position.t+e-.5)}),this.hbar){var i=t/r;this.hbar.call(a.setTranslate,t+i*this._hbarTranslateMax,e)}if(this.vbar){var s=e/n;this.vbar.call(a.setTranslate,t,e+s*this._vbarTranslateMax)}}},84284:function(t){"use strict";t.exports={FROM_BL:{left:0,center:.5,right:1,bottom:0,middle:.5,top:1},FROM_TL:{left:0,center:.5,right:1,bottom:1,middle:.5,top:0},FROM_BR:{left:1,center:.5,right:0,bottom:0,middle:.5,top:1},LINE_SPACING:1.3,CAP_SHIFT:.7,MID_SHIFT:.35,OPPOSITE_SIDE:{left:"right",right:"left",top:"bottom",bottom:"top"}}},36208:function(t){"use strict";t.exports={axisRefDescription:function(t,e,r){return["If set to a",t,"axis id (e.g. *"+t+"* or","*"+t+"2*), the `"+t+"` position refers to a",t,"coordinate. If set to *paper*, the `"+t+"`","position refers to the distance from the",e,"of the plotting","area in normalized coordinates where *0* (*1*) corresponds to the",e,"("+r+"). If set to a",t,"axis ID followed by","*domain* (separated by a space), the position behaves like for","*paper*, but refers to the distance in fractions of the domain","length from the",e,"of the domain of that axis: e.g.,","*"+t+"2 domain* refers to the domain of the second",t," axis and a",t,"position of 0.5 refers to the","point between the",e,"and the",r,"of the domain of the","second",t,"axis."].join(" ")}}},48164:function(t){"use strict";t.exports={INCREASING:{COLOR:"#3D9970",SYMBOL:"▲"},DECREASING:{COLOR:"#FF4136",SYMBOL:"▼"}}},26880:function(t){"use strict";t.exports={FORMAT_LINK:"https://github.com/d3/d3-format/tree/v1.4.5#d3-format",DATE_FORMAT_LINK:"https://github.com/d3/d3-time-format/tree/v2.2.3#locale_format"}},69104:function(t){"use strict";t.exports={COMPARISON_OPS:["=","!=","<",">=",">","<="],COMPARISON_OPS2:["=","<",">=",">","<="],INTERVAL_OPS:["[]","()","[)","(]","][",")(","](",")["],SET_OPS:["{}","}{"],CONSTRAINT_REDUCTION:{"=":"=","<":"<","<=":"<",">":">",">=":">","[]":"[]","()":"[]","[)":"[]","(]":"[]","][":"][",")(":"][","](":"][",")[":"]["}}},99168:function(t){"use strict";t.exports={solid:[[],0],dot:[[.5,1],200],dash:[[.5,1],50],longdash:[[.5,1],10],dashdot:[[.5,.625,.875,1],50],longdashdot:[[.5,.7,.8,1],10]}},87792:function(t){"use strict";t.exports={circle:"●","circle-open":"○",square:"■","square-open":"□",diamond:"◆","diamond-open":"◇",cross:"+",x:"❌"}},13448:function(t){"use strict";t.exports={SHOW_PLACEHOLDER:100,HIDE_PLACEHOLDER:1e3,DESELECTDIM:.2}},39032:function(t){"use strict";t.exports={BADNUM:void 0,FP_SAFE:1e-4*Number.MAX_VALUE,ONEMAXYEAR:316224e5,ONEAVGYEAR:315576e5,ONEMINYEAR:31536e6,ONEMAXQUARTER:79488e5,ONEAVGQUARTER:78894e5,ONEMINQUARTER:76896e5,ONEMAXMONTH:26784e5,ONEAVGMONTH:26298e5,ONEMINMONTH:24192e5,ONEWEEK:6048e5,ONEDAY:864e5,ONEHOUR:36e5,ONEMIN:6e4,ONESEC:1e3,EPOCHJD:2440587.5,ALMOST_EQUAL:.999999,LOG_CLIP:10,MINUS_SIGN:"−"}},2264:function(t,e){"use strict";e.CSS_DECLARATIONS=[["image-rendering","optimizeSpeed"],["image-rendering","-moz-crisp-edges"],["image-rendering","-o-crisp-edges"],["image-rendering","-webkit-optimize-contrast"],["image-rendering","optimize-contrast"],["image-rendering","crisp-edges"],["image-rendering","pixelated"]],e.STYLE=e.CSS_DECLARATIONS.map((function(t){return t.join(": ")+"; "})).join("")},9616:function(t,e){"use strict";e.xmlns="http://www.w3.org/2000/xmlns/",e.svg="http://www.w3.org/2000/svg",e.xlink="http://www.w3.org/1999/xlink",e.svgAttrs={xmlns:e.svg,"xmlns:xlink":e.xlink}},64884:function(t,e,r){"use strict";e.version=r(25788).version,r(88324),r(79288);for(var n=r(24040),i=e.register=n.register,a=r(22448),o=Object.keys(a),s=0;s<o.length;s++){var l=o[s];"_"!==l.charAt(0)&&(e[l]=a[l]),i({moduleType:"apiMethod",name:l,fn:a[l]})}i(r(65875)),i([r(79180),r(56864),r(22676),r(41592),r(7402),r(76908),r(97544),r(49692),r(41152),r(12704),r(64968),r(8932),r(55080),r(2780),r(93024),r(45460)]),i([r(6580),r(11680)]),window.PlotlyLocales&&Array.isArray(window.PlotlyLocales)&&(i(window.PlotlyLocales),delete window.PlotlyLocales),e.Icons=r(9224);var u=r(93024),c=r(7316);e.Plots={resize:c.resize,graphJson:c.graphJson,sendDataToCloud:c.sendDataToCloud},e.Fx={hover:u.hover,unhover:u.unhover,loneHover:u.loneHover,loneUnhover:u.loneUnhover},e.Snapshot=r(78904),e.PlotSchema=r(73060)},9224:function(t){"use strict";t.exports={undo:{width:857.1,height:1e3,path:"m857 350q0-87-34-166t-91-137-137-92-166-34q-96 0-183 41t-147 114q-4 6-4 13t5 11l76 77q6 5 14 5 9-1 13-7 41-53 100-82t126-29q58 0 110 23t92 61 61 91 22 111-22 111-61 91-92 61-110 23q-55 0-105-20t-90-57l77-77q17-16 8-38-10-23-33-23h-250q-15 0-25 11t-11 25v250q0 24 22 33 22 10 39-8l72-72q60 57 137 88t159 31q87 0 166-34t137-92 91-137 34-166z",transform:"matrix(1 0 0 -1 0 850)"},home:{width:928.6,height:1e3,path:"m786 296v-267q0-15-11-26t-25-10h-214v214h-143v-214h-214q-15 0-25 10t-11 26v267q0 1 0 2t0 2l321 264 321-264q1-1 1-4z m124 39l-34-41q-5-5-12-6h-2q-7 0-12 3l-386 322-386-322q-7-4-13-4-7 2-12 7l-35 41q-4 5-3 13t6 12l401 334q18 15 42 15t43-15l136-114v109q0 8 5 13t13 5h107q8 0 13-5t5-13v-227l122-102q5-5 6-12t-4-13z",transform:"matrix(1 0 0 -1 0 850)"},"camera-retro":{width:1e3,height:1e3,path:"m518 386q0 8-5 13t-13 5q-37 0-63-27t-26-63q0-8 5-13t13-5 12 5 5 13q0 23 16 38t38 16q8 0 13 5t5 13z m125-73q0-59-42-101t-101-42-101 42-42 101 42 101 101 42 101-42 42-101z m-572-320h858v71h-858v-71z m643 320q0 89-62 152t-152 62-151-62-63-152 63-151 151-63 152 63 62 151z m-571 358h214v72h-214v-72z m-72-107h858v143h-462l-36-71h-360v-72z m929 143v-714q0-30-21-51t-50-21h-858q-29 0-50 21t-21 51v714q0 30 21 51t50 21h858q29 0 50-21t21-51z",transform:"matrix(1 0 0 -1 0 850)"},zoombox:{width:1e3,height:1e3,path:"m1000-25l-250 251c40 63 63 138 63 218 0 224-182 406-407 406-224 0-406-182-406-406s183-406 407-406c80 0 155 22 218 62l250-250 125 125z m-812 250l0 438 437 0 0-438-437 0z m62 375l313 0 0-312-313 0 0 312z",transform:"matrix(1 0 0 -1 0 850)"},pan:{width:1e3,height:1e3,path:"m1000 350l-187 188 0-125-250 0 0 250 125 0-188 187-187-187 125 0 0-250-250 0 0 125-188-188 186-187 0 125 252 0 0-250-125 0 187-188 188 188-125 0 0 250 250 0 0-126 187 188z",transform:"matrix(1 0 0 -1 0 850)"},zoom_plus:{width:875,height:1e3,path:"m1 787l0-875 875 0 0 875-875 0z m687-500l-187 0 0-187-125 0 0 187-188 0 0 125 188 0 0 187 125 0 0-187 187 0 0-125z",transform:"matrix(1 0 0 -1 0 850)"},zoom_minus:{width:875,height:1e3,path:"m0 788l0-876 875 0 0 876-875 0z m688-500l-500 0 0 125 500 0 0-125z",transform:"matrix(1 0 0 -1 0 850)"},autoscale:{width:1e3,height:1e3,path:"m250 850l-187 0-63 0 0-62 0-188 63 0 0 188 187 0 0 62z m688 0l-188 0 0-62 188 0 0-188 62 0 0 188 0 62-62 0z m-875-938l0 188-63 0 0-188 0-62 63 0 187 0 0 62-187 0z m875 188l0-188-188 0 0-62 188 0 62 0 0 62 0 188-62 0z m-125 188l-1 0-93-94-156 156 156 156 92-93 2 0 0 250-250 0 0-2 93-92-156-156-156 156 94 92 0 2-250 0 0-250 0 0 93 93 157-156-157-156-93 94 0 0 0-250 250 0 0 0-94 93 156 157 156-157-93-93 0 0 250 0 0 250z",transform:"matrix(1 0 0 -1 0 850)"},tooltip_basic:{width:1500,height:1e3,path:"m375 725l0 0-375-375 375-374 0-1 1125 0 0 750-1125 0z",transform:"matrix(1 0 0 -1 0 850)"},tooltip_compare:{width:1125,height:1e3,path:"m187 786l0 2-187-188 188-187 0 0 937 0 0 373-938 0z m0-499l0 1-187-188 188-188 0 0 937 0 0 376-938-1z",transform:"matrix(1 0 0 -1 0 850)"},plotlylogo:{width:1542,height:1e3,path:"m0-10h182v-140h-182v140z m228 146h183v-286h-183v286z m225 714h182v-1000h-182v1000z m225-285h182v-715h-182v715z m225 142h183v-857h-183v857z m231-428h182v-429h-182v429z m225-291h183v-138h-183v138z",transform:"matrix(1 0 0 -1 0 850)"},"z-axis":{width:1e3,height:1e3,path:"m833 5l-17 108v41l-130-65 130-66c0 0 0 38 0 39 0-1 36-14 39-25 4-15-6-22-16-30-15-12-39-16-56-20-90-22-187-23-279-23-261 0-341 34-353 59 3 60 228 110 228 110-140-8-351-35-351-116 0-120 293-142 474-142 155 0 477 22 477 142 0 50-74 79-163 96z m-374 94c-58-5-99-21-99-40 0-24 65-43 144-43 79 0 143 19 143 43 0 19-42 34-98 40v216h87l-132 135-133-135h88v-216z m167 515h-136v1c16 16 31 34 46 52l84 109v54h-230v-71h124v-1c-16-17-28-32-44-51l-89-114v-51h245v72z",transform:"matrix(1 0 0 -1 0 850)"},"3d_rotate":{width:1e3,height:1e3,path:"m922 660c-5 4-9 7-14 11-359 263-580-31-580-31l-102 28 58-400c0 1 1 1 2 2 118 108 351 249 351 249s-62 27-100 42c88 83 222 183 347 122 16-8 30-17 44-27-2 1-4 2-6 4z m36-329c0 0 64 229-88 296-62 27-124 14-175-11 157-78 225-208 249-266 8-19 11-31 11-31 2 5 6 15 11 32-5-13-8-20-8-20z m-775-239c70-31 117-50 198-32-121 80-199 346-199 346l-96-15-58-12c0 0 55-226 155-287z m603 133l-317-139c0 0 4-4 19-14 7-5 24-15 24-15s-177-147-389 4c235-287 536-112 536-112l31-22 100 299-4-1z m-298-153c6-4 14-9 24-15 0 0-17 10-24 15z",transform:"matrix(1 0 0 -1 0 850)"},camera:{width:1e3,height:1e3,path:"m500 450c-83 0-150-67-150-150 0-83 67-150 150-150 83 0 150 67 150 150 0 83-67 150-150 150z m400 150h-120c-16 0-34 13-39 29l-31 93c-6 15-23 28-40 28h-340c-16 0-34-13-39-28l-31-94c-6-15-23-28-40-28h-120c-55 0-100-45-100-100v-450c0-55 45-100 100-100h800c55 0 100 45 100 100v450c0 55-45 100-100 100z m-400-550c-138 0-250 112-250 250 0 138 112 250 250 250 138 0 250-112 250-250 0-138-112-250-250-250z m365 380c-19 0-35 16-35 35 0 19 16 35 35 35 19 0 35-16 35-35 0-19-16-35-35-35z",transform:"matrix(1 0 0 -1 0 850)"},movie:{width:1e3,height:1e3,path:"m938 413l-188-125c0 37-17 71-44 94 64 38 107 107 107 187 0 121-98 219-219 219-121 0-219-98-219-219 0-61 25-117 66-156h-115c30 33 49 76 49 125 0 103-84 187-187 187s-188-84-188-187c0-57 26-107 65-141-38-22-65-62-65-109v-250c0-70 56-126 125-126h500c69 0 125 56 125 126l188-126c34 0 62 28 62 63v375c0 35-28 63-62 63z m-750 0c-69 0-125 56-125 125s56 125 125 125 125-56 125-125-56-125-125-125z m406-1c-87 0-157 70-157 157 0 86 70 156 157 156s156-70 156-156-70-157-156-157z",transform:"matrix(1 0 0 -1 0 850)"},question:{width:857.1,height:1e3,path:"m500 82v107q0 8-5 13t-13 5h-107q-8 0-13-5t-5-13v-107q0-8 5-13t13-5h107q8 0 13 5t5 13z m143 375q0 49-31 91t-77 65-95 23q-136 0-207-119-9-14 4-24l74-55q4-4 10-4 9 0 14 7 30 38 48 51 19 14 48 14 27 0 48-15t21-33q0-21-11-34t-38-25q-35-16-65-48t-29-70v-20q0-8 5-13t13-5h107q8 0 13 5t5 13q0 10 12 27t30 28q18 10 28 16t25 19 25 27 16 34 7 45z m214-107q0-117-57-215t-156-156-215-58-216 58-155 156-58 215 58 215 155 156 216 58 215-58 156-156 57-215z",transform:"matrix(1 0 0 -1 0 850)"},disk:{width:857.1,height:1e3,path:"m214-7h429v214h-429v-214z m500 0h72v500q0 8-6 21t-11 20l-157 156q-5 6-19 12t-22 5v-232q0-22-15-38t-38-16h-322q-22 0-37 16t-16 38v232h-72v-714h72v232q0 22 16 38t37 16h465q22 0 38-16t15-38v-232z m-214 518v178q0 8-5 13t-13 5h-107q-7 0-13-5t-5-13v-178q0-8 5-13t13-5h107q7 0 13 5t5 13z m357-18v-518q0-22-15-38t-38-16h-750q-23 0-38 16t-16 38v750q0 22 16 38t38 16h517q23 0 50-12t42-26l156-157q16-15 27-42t11-49z",transform:"matrix(1 0 0 -1 0 850)"},drawopenpath:{width:70,height:70,path:"M33.21,85.65a7.31,7.31,0,0,1-2.59-.48c-8.16-3.11-9.27-19.8-9.88-41.3-.1-3.58-.19-6.68-.35-9-.15-2.1-.67-3.48-1.43-3.79-2.13-.88-7.91,2.32-12,5.86L3,32.38c1.87-1.64,11.55-9.66,18.27-6.9,2.13.87,4.75,3.14,5.17,9,.17,2.43.26,5.59.36,9.25a224.17,224.17,0,0,0,1.5,23.4c1.54,10.76,4,12.22,4.48,12.4.84.32,2.79-.46,5.76-3.59L43,80.07C41.53,81.57,37.68,85.64,33.21,85.65ZM74.81,69a11.34,11.34,0,0,0,6.09-6.72L87.26,44.5,74.72,32,56.9,38.35c-2.37.86-5.57,3.42-6.61,6L38.65,72.14l8.42,8.43ZM55,46.27a7.91,7.91,0,0,1,3.64-3.17l14.8-5.3,8,8L76.11,60.6l-.06.19a6.37,6.37,0,0,1-3,3.43L48.25,74.59,44.62,71Zm16.57,7.82A6.9,6.9,0,1,0,64.64,61,6.91,6.91,0,0,0,71.54,54.09Zm-4.05,0a2.85,2.85,0,1,1-2.85-2.85A2.86,2.86,0,0,1,67.49,54.09Zm-4.13,5.22L60.5,56.45,44.26,72.7l2.86,2.86ZM97.83,35.67,84.14,22l-8.57,8.57L89.26,44.24Zm-13.69-8,8,8-2.85,2.85-8-8Z",transform:"matrix(1 0 0 1 -15 -15)"},drawclosedpath:{width:90,height:90,path:"M88.41,21.12a26.56,26.56,0,0,0-36.18,0l-2.07,2-2.07-2a26.57,26.57,0,0,0-36.18,0,23.74,23.74,0,0,0,0,34.8L48,90.12a3.22,3.22,0,0,0,4.42,0l36-34.21a23.73,23.73,0,0,0,0-34.79ZM84,51.24,50.16,83.35,16.35,51.25a17.28,17.28,0,0,1,0-25.47,20,20,0,0,1,27.3,0l4.29,4.07a3.23,3.23,0,0,0,4.44,0l4.29-4.07a20,20,0,0,1,27.3,0,17.27,17.27,0,0,1,0,25.46ZM66.76,47.68h-33v6.91h33ZM53.35,35H46.44V68h6.91Z",transform:"matrix(1 0 0 1 -5 -5)"},lasso:{width:1031,height:1e3,path:"m1018 538c-36 207-290 336-568 286-277-48-473-256-436-463 10-57 36-108 76-151-13-66 11-137 68-183 34-28 75-41 114-42l-55-70 0 0c-2-1-3-2-4-3-10-14-8-34 5-45 14-11 34-8 45 4 1 1 2 3 2 5l0 0 113 140c16 11 31 24 45 40 4 3 6 7 8 11 48-3 100 0 151 9 278 48 473 255 436 462z m-624-379c-80 14-149 48-197 96 42 42 109 47 156 9 33-26 47-66 41-105z m-187-74c-19 16-33 37-39 60 50-32 109-55 174-68-42-25-95-24-135 8z m360 75c-34-7-69-9-102-8 8 62-16 128-68 170-73 59-175 54-244-5-9 20-16 40-20 61-28 159 121 317 333 354s407-60 434-217c28-159-121-318-333-355z",transform:"matrix(1 0 0 -1 0 850)"},selectbox:{width:1e3,height:1e3,path:"m0 850l0-143 143 0 0 143-143 0z m286 0l0-143 143 0 0 143-143 0z m285 0l0-143 143 0 0 143-143 0z m286 0l0-143 143 0 0 143-143 0z m-857-286l0-143 143 0 0 143-143 0z m857 0l0-143 143 0 0 143-143 0z m-857-285l0-143 143 0 0 143-143 0z m857 0l0-143 143 0 0 143-143 0z m-857-286l0-143 143 0 0 143-143 0z m286 0l0-143 143 0 0 143-143 0z m285 0l0-143 143 0 0 143-143 0z m286 0l0-143 143 0 0 143-143 0z",transform:"matrix(1 0 0 -1 0 850)"},drawline:{width:70,height:70,path:"M60.64,62.3a11.29,11.29,0,0,0,6.09-6.72l6.35-17.72L60.54,25.31l-17.82,6.4c-2.36.86-5.57,3.41-6.6,6L24.48,65.5l8.42,8.42ZM40.79,39.63a7.89,7.89,0,0,1,3.65-3.17l14.79-5.31,8,8L61.94,54l-.06.19a6.44,6.44,0,0,1-3,3.43L34.07,68l-3.62-3.63Zm16.57,7.81a6.9,6.9,0,1,0-6.89,6.9A6.9,6.9,0,0,0,57.36,47.44Zm-4,0a2.86,2.86,0,1,1-2.85-2.85A2.86,2.86,0,0,1,53.32,47.44Zm-4.13,5.22L46.33,49.8,30.08,66.05l2.86,2.86ZM83.65,29,70,15.34,61.4,23.9,75.09,37.59ZM70,21.06l8,8-2.84,2.85-8-8ZM87,80.49H10.67V87H87Z",transform:"matrix(1 0 0 1 -15 -15)"},drawrect:{width:80,height:80,path:"M78,22V79H21V22H78m9-9H12V88H87V13ZM68,46.22H31V54H68ZM53,32H45.22V69H53Z",transform:"matrix(1 0 0 1 -10 -10)"},drawcircle:{width:80,height:80,path:"M50,84.72C26.84,84.72,8,69.28,8,50.3S26.84,15.87,50,15.87,92,31.31,92,50.3,73.16,84.72,50,84.72Zm0-60.59c-18.6,0-33.74,11.74-33.74,26.17S31.4,76.46,50,76.46,83.74,64.72,83.74,50.3,68.6,24.13,50,24.13Zm17.15,22h-34v7.11h34Zm-13.8-13H46.24v34h7.11Z",transform:"matrix(1 0 0 1 -10 -10)"},eraseshape:{width:80,height:80,path:"M82.77,78H31.85L6,49.57,31.85,21.14H82.77a8.72,8.72,0,0,1,8.65,8.77V69.24A8.72,8.72,0,0,1,82.77,78ZM35.46,69.84H82.77a.57.57,0,0,0,.49-.6V29.91a.57.57,0,0,0-.49-.61H35.46L17,49.57Zm32.68-34.7-24,24,5,5,24-24Zm-19,.53-5,5,24,24,5-5Z",transform:"matrix(1 0 0 1 -10 -10)"},spikeline:{width:1e3,height:1e3,path:"M512 409c0-57-46-104-103-104-57 0-104 47-104 104 0 57 47 103 104 103 57 0 103-46 103-103z m-327-39l92 0 0 92-92 0z m-185 0l92 0 0 92-92 0z m370-186l92 0 0 93-92 0z m0-184l92 0 0 92-92 0z",transform:"matrix(1.5 0 0 -1.5 0 850)"},pencil:{width:1792,height:1792,path:"M491 1536l91-91-235-235-91 91v107h128v128h107zm523-928q0-22-22-22-10 0-17 7l-542 542q-7 7-7 17 0 22 22 22 10 0 17-7l542-542q7-7 7-17zm-54-192l416 416-832 832h-416v-416zm683 96q0 53-37 90l-166 166-416-416 166-165q36-38 90-38 53 0 91 38l235 234q37 39 37 91z",transform:"matrix(1 0 0 1 0 1)"},newplotlylogo:{name:"newplotlylogo",svg:["<svg xmlns='http://www.w3.org/2000/svg' viewBox='0 0 132 132'>","<defs>"," <style>","  .cls-0{fill:#000;}","  .cls-1{fill:#FFF;}","  .cls-2{fill:#F26;}","  .cls-3{fill:#D69;}","  .cls-4{fill:#BAC;}","  .cls-5{fill:#9EF;}"," </style>","</defs>"," <title>plotly-logomark</title>"," <g id='symbol'>","  <rect class='cls-0' x='0' y='0' width='132' height='132' rx='18' ry='18'/>","  <circle class='cls-5' cx='102' cy='30' r='6'/>","  <circle class='cls-4' cx='78' cy='30' r='6'/>","  <circle class='cls-4' cx='78' cy='54' r='6'/>","  <circle class='cls-3' cx='54' cy='30' r='6'/>","  <circle class='cls-2' cx='30' cy='30' r='6'/>","  <circle class='cls-2' cx='30' cy='54' r='6'/>","  <path class='cls-1' d='M30,72a6,6,0,0,0-6,6v24a6,6,0,0,0,12,0V78A6,6,0,0,0,30,72Z'/>","  <path class='cls-1' d='M78,72a6,6,0,0,0-6,6v24a6,6,0,0,0,12,0V78A6,6,0,0,0,78,72Z'/>","  <path class='cls-1' d='M54,48a6,6,0,0,0-6,6v48a6,6,0,0,0,12,0V54A6,6,0,0,0,54,48Z'/>","  <path class='cls-1' d='M102,48a6,6,0,0,0-6,6v48a6,6,0,0,0,12,0V54A6,6,0,0,0,102,48Z'/>"," </g>","</svg>"].join("")}}},98308:function(t,e){"use strict";e.isLeftAnchor=function(t){return"left"===t.xanchor||"auto"===t.xanchor&&t.x<=1/3},e.isCenterAnchor=function(t){return"center"===t.xanchor||"auto"===t.xanchor&&t.x>1/3&&t.x<2/3},e.isRightAnchor=function(t){return"right"===t.xanchor||"auto"===t.xanchor&&t.x>=2/3},e.isTopAnchor=function(t){return"top"===t.yanchor||"auto"===t.yanchor&&t.y>=2/3},e.isMiddleAnchor=function(t){return"middle"===t.yanchor||"auto"===t.yanchor&&t.y>1/3&&t.y<2/3},e.isBottomAnchor=function(t){return"bottom"===t.yanchor||"auto"===t.yanchor&&t.y<=1/3}},11864:function(t,e,r){"use strict";var n=r(20435),i=n.mod,a=n.modHalf,o=Math.PI,s=2*o;function l(t){return Math.abs(t[1]-t[0])>s-1e-14}function u(t,e){return a(e-t,s)}function c(t,e){if(l(e))return!0;var r,n;e[0]<e[1]?(r=e[0],n=e[1]):(r=e[1],n=e[0]),(r=i(r,s))>(n=i(n,s))&&(n+=s);var a=i(t,s),o=a+s;return a>=r&&a<=n||o>=r&&o<=n}function f(t,e,r,n,i,a,u){i=i||0,a=a||0;var c,f,h,p,d,v=l([r,n]);function g(t,e){return[t*Math.cos(e)+i,a-t*Math.sin(e)]}v?(c=0,f=o,h=s):r<n?(c=r,h=n):(c=n,h=r),t<e?(p=t,d=e):(p=e,d=t);var y,m=Math.abs(h-c)<=o?0:1;function x(t,e,r){return"A"+[t,t]+" "+[0,m,r]+" "+g(t,e)}return v?y=null===p?"M"+g(d,c)+x(d,f,0)+x(d,h,0)+"Z":"M"+g(p,c)+x(p,f,0)+x(p,h,0)+"ZM"+g(d,c)+x(d,f,1)+x(d,h,1)+"Z":null===p?(y="M"+g(d,c)+x(d,h,0),u&&(y+="L0,0Z")):y="M"+g(p,c)+"L"+g(d,c)+x(d,h,0)+"L"+g(p,h)+x(p,c,1)+"Z",y}t.exports={deg2rad:function(t){return t/180*o},rad2deg:function(t){return t/o*180},angleDelta:u,angleDist:function(t,e){return Math.abs(u(t,e))},isFullCircle:l,isAngleInsideSector:c,isPtInsideSector:function(t,e,r,n){return!!c(e,n)&&(r[0]<r[1]?(i=r[0],a=r[1]):(i=r[1],a=r[0]),t>=i&&t<=a);var i,a},pathArc:function(t,e,r,n,i){return f(null,t,e,r,n,i,0)},pathSector:function(t,e,r,n,i){return f(null,t,e,r,n,i,1)},pathAnnulus:function(t,e,r,n,i,a){return f(t,e,r,n,i,a,1)}}},38116:function(t,e,r){"use strict";var n=r(83160).decode,i=r(63620),a=Array.isArray,o=ArrayBuffer,s=DataView;function l(t){return o.isView(t)&&!(t instanceof s)}function u(t){return a(t)||l(t)}e.isTypedArray=l,e.isArrayOrTypedArray=u,e.isArray1D=function(t){return!u(t[0])},e.ensureArray=function(t,e){return a(t)||(t=[]),t.length=e,t};var c={u1c:"undefined"==typeof Uint8ClampedArray?void 0:Uint8ClampedArray,i1:"undefined"==typeof Int8Array?void 0:Int8Array,u1:"undefined"==typeof Uint8Array?void 0:Uint8Array,i2:"undefined"==typeof Int16Array?void 0:Int16Array,u2:"undefined"==typeof Uint16Array?void 0:Uint16Array,i4:"undefined"==typeof Int32Array?void 0:Int32Array,u4:"undefined"==typeof Uint32Array?void 0:Uint32Array,f4:"undefined"==typeof Float32Array?void 0:Float32Array,f8:"undefined"==typeof Float64Array?void 0:Float64Array};function f(t){return t.constructor===ArrayBuffer}function h(t,e,r){if(u(t)){if(u(t[0])){for(var n=r,i=0;i<t.length;i++)n=e(n,t[i].length);return n}return t.length}return 0}c.uint8c=c.u1c,c.uint8=c.u1,c.int8=c.i1,c.uint16=c.u2,c.int16=c.i2,c.uint32=c.u4,c.int32=c.i4,c.float32=c.f4,c.float64=c.f8,e.isArrayBuffer=f,e.decodeTypedArraySpec=function(t){var e=[],r=function(t){return{bdata:t.bdata,dtype:t.dtype,shape:t.shape}}(t),i=r.dtype,a=c[i];if(!a)throw new Error('Error in dtype: "'+i+'"');var o=a.BYTES_PER_ELEMENT,s=r.bdata;f(s)||(s=n(s));var l=void 0===r.shape?[s.byteLength/o]:(""+r.shape).split(",");l.reverse();var u,h,p=l.length,d=+l[0],v=o*d,g=0;if(1===p)e=new a(s);else if(2===p)for(u=+l[1],h=0;h<u;h++)e[h]=new a(s,g,d),g+=v;else{if(3!==p)throw new Error("ndim: "+p+'is not supported with the shape:"'+r.shape+'"');u=+l[1];for(var y=+l[2],m=0;m<y;m++)for(e[m]=[],h=0;h<u;h++)e[m][h]=new a(s,g,d),g+=v}return e.bdata=r.bdata,e.dtype=r.dtype,e.shape=l.reverse().join(","),t._inputArray=e,e},e.isTypedArraySpec=function(t){return i(t)&&t.hasOwnProperty("dtype")&&"string"==typeof t.dtype&&t.hasOwnProperty("bdata")&&("string"==typeof t.bdata||f(t.bdata))&&(void 0===t.shape||t.hasOwnProperty("shape")&&("string"==typeof t.shape||"number"==typeof t.shape))},e.concat=function(){var t,e,r,n,i,o,s,l,u=[],c=!0,f=0;for(r=0;r<arguments.length;r++)(o=(n=arguments[r]).length)&&(e?u.push(n):(e=n,i=o),a(n)?t=!1:(c=!1,f?t!==n.constructor&&(t=!1):t=n.constructor),f+=o);if(!f)return[];if(!u.length)return e;if(c)return e.concat.apply(e,u);if(t){for((s=new t(f)).set(e),r=0;r<u.length;r++)n=u[r],s.set(n,i),i+=n.length;return s}for(s=new Array(f),l=0;l<e.length;l++)s[l]=e[l];for(r=0;r<u.length;r++){for(n=u[r],l=0;l<n.length;l++)s[i+l]=n[l];i+=l}return s},e.maxRowLength=function(t){return h(t,Math.max,0)},e.minRowLength=function(t){return h(t,Math.min,1/0)}},54037:function(t,e,r){"use strict";var n=r(38248),i=r(39032).BADNUM,a=/^['"%,$#\s']+|[, ]|['"%,$#\s']+$/g;t.exports=function(t){return"string"==typeof t&&(t=t.replace(a,"")),n(t)?Number(t):i}},73696:function(t){"use strict";t.exports=function(t){var e=t._fullLayout;e._glcanvas&&e._glcanvas.size()&&e._glcanvas.each((function(t){t.regl&&t.regl.clear({color:!0,depth:!0})}))}},75352:function(t){"use strict";t.exports=function(t){t._responsiveChartHandler&&(window.removeEventListener("resize",t._responsiveChartHandler),delete t._responsiveChartHandler)}},63064:function(t,e,r){"use strict";var n=r(38248),i=r(49760),a=r(45464),o=r(88304),s=r(76308),l=r(13448).DESELECTDIM,u=r(22296),c=r(53756).counter,f=r(20435).modHalf,h=r(38116).isArrayOrTypedArray,p=r(38116).isTypedArraySpec,d=r(38116).decodeTypedArraySpec;function v(t,r){var n=e.valObjectMeta[r.valType];if(r.arrayOk&&h(t))return!0;if(n.validateFunction)return n.validateFunction(t,r);var i={},a=i,o={set:function(t){a=t}};return n.coerceFunction(t,o,i,r),a!==i}e.valObjectMeta={data_array:{coerceFunction:function(t,e,r){e.set(h(t)?t:p(t)?d(t):r)}},enumerated:{coerceFunction:function(t,e,r,n){n.coerceNumber&&(t=+t),-1===n.values.indexOf(t)?e.set(r):e.set(t)},validateFunction:function(t,e){e.coerceNumber&&(t=+t);for(var r=e.values,n=0;n<r.length;n++){var i=String(r[n]);if("/"===i.charAt(0)&&"/"===i.charAt(i.length-1)){if(new RegExp(i.substr(1,i.length-2)).test(t))return!0}else if(t===r[n])return!0}return!1}},boolean:{coerceFunction:function(t,e,r){!0===t||!1===t?e.set(t):e.set(r)}},number:{coerceFunction:function(t,e,r,i){!n(t)||void 0!==i.min&&t<i.min||void 0!==i.max&&t>i.max?e.set(r):e.set(+t)}},integer:{coerceFunction:function(t,e,r,i){t%1||!n(t)||void 0!==i.min&&t<i.min||void 0!==i.max&&t>i.max?e.set(r):e.set(+t)}},string:{coerceFunction:function(t,e,r,n){if("string"!=typeof t){var i="number"==typeof t;!0!==n.strict&&i?e.set(String(t)):e.set(r)}else n.noBlank&&!t?e.set(r):e.set(t)}},color:{coerceFunction:function(t,e,r){i(t).isValid()?e.set(t):e.set(r)}},colorlist:{coerceFunction:function(t,e,r){Array.isArray(t)&&t.length&&t.every((function(t){return i(t).isValid()}))?e.set(t):e.set(r)}},colorscale:{coerceFunction:function(t,e,r){e.set(o.get(t,r))}},angle:{coerceFunction:function(t,e,r){"auto"===t?e.set("auto"):n(t)?e.set(f(+t,360)):e.set(r)}},subplotid:{coerceFunction:function(t,e,r,n){var i=n.regex||c(r);"string"==typeof t&&i.test(t)?e.set(t):e.set(r)},validateFunction:function(t,e){var r=e.dflt;return t===r||"string"==typeof t&&!!c(r).test(t)}},flaglist:{coerceFunction:function(t,e,r,n){if(-1===(n.extras||[]).indexOf(t))if("string"==typeof t){for(var i=t.split("+"),a=0;a<i.length;){var o=i[a];-1===n.flags.indexOf(o)||i.indexOf(o)<a?i.splice(a,1):a++}i.length?e.set(i.join("+")):e.set(r)}else e.set(r);else e.set(t)}},any:{coerceFunction:function(t,e,r){void 0===t?e.set(r):e.set(p(t)?d(t):t)}},info_array:{coerceFunction:function(t,r,n,i){function a(t,r,n){var i,a={set:function(t){i=t}};return void 0===n&&(n=r.dflt),e.valObjectMeta[r.valType].coerceFunction(t,a,n,r),i}if(p(t)&&(t=d(t)),h(t)){var o,s,l,u,c,f,v=2===i.dimensions||"1-2"===i.dimensions&&Array.isArray(t)&&h(t[0]),g=i.items,y=[],m=Array.isArray(g),x=m&&v&&h(g[0]),b=v&&m&&!x,_=m&&!b?g.length:t.length;if(n=Array.isArray(n)?n:[],v)for(o=0;o<_;o++)for(y[o]=[],l=h(t[o])?t[o]:[],c=b?g.length:m?g[o].length:l.length,s=0;s<c;s++)u=b?g[s]:m?g[o][s]:g,void 0!==(f=a(l[s],u,(n[o]||[])[s]))&&(y[o][s]=f);else for(o=0;o<_;o++)void 0!==(f=a(t[o],m?g[o]:g,n[o]))&&(y[o]=f);r.set(y)}else r.set(n)},validateFunction:function(t,e){if(!h(t))return!1;var r=e.items,n=Array.isArray(r),i=2===e.dimensions;if(!e.freeLength&&t.length!==r.length)return!1;for(var a=0;a<t.length;a++)if(i){if(!h(t[a])||!e.freeLength&&t[a].length!==r[a].length)return!1;for(var o=0;o<t[a].length;o++)if(!v(t[a][o],n?r[a][o]:r))return!1}else if(!v(t[a],n?r[a]:r))return!1;return!0}}},e.coerce=function(t,r,n,i,a){var o=u(n,i).get(),s=u(t,i),l=u(r,i),c=s.get(),f=r._template;if(void 0===c&&f&&(c=u(f,i).get(),f=0),void 0===a&&(a=o.dflt),o.arrayOk){if(h(c))return l.set(c),c;if(p(c))return c=d(c),l.set(c),c}var g=e.valObjectMeta[o.valType].coerceFunction;g(c,l,a,o);var y=l.get();return f&&y===a&&!v(c,o)&&(g(c=u(f,i).get(),l,a,o),y=l.get()),y},e.coerce2=function(t,r,n,i,a){var o=u(t,i),s=e.coerce(t,r,n,i,a);return null!=o.get()&&s},e.coerceFont=function(t,e,r,n){n||(n={});var i={};return r=r||{},i.family=t(e+".family",r.family),i.size=t(e+".size",r.size),i.color=t(e+".color",r.color),i.weight=t(e+".weight",r.weight),i.style=t(e+".style",r.style),n.noFontVariant||(i.variant=t(e+".variant",r.variant)),i},e.coercePattern=function(t,e,r,n){if(t(e+".shape")){t(e+".solidity"),t(e+".size");var i="overlay"===t(e+".fillmode");if(!n){var a=t(e+".bgcolor",i?r:void 0);t(e+".fgcolor",i?s.contrast(a):r)}t(e+".fgopacity",i?.5:1)}},e.coerceHoverinfo=function(t,r,n){var i,o=r._module.attributes,s=o.hoverinfo?o:a,l=s.hoverinfo;if(1===n._dataLength){var u="all"===l.dflt?l.flags.slice():l.dflt.split("+");u.splice(u.indexOf("name"),1),i=u.join("+")}return e.coerce(t,r,s,"hoverinfo",i)},e.coerceSelectionMarkerOpacity=function(t,e){if(t.marker){var r,n,i=t.marker.opacity;void 0!==i&&(h(i)||t.selected||t.unselected||(r=i,n=l*i),e("selected.marker.opacity",r),e("unselected.marker.opacity",n))}},e.validate=v},67555:function(t,e,r){"use strict";var n,i,a=r(94336).Yn,o=r(38248),s=r(24248),l=r(20435).mod,u=r(39032),c=u.BADNUM,f=u.ONEDAY,h=u.ONEHOUR,p=u.ONEMIN,d=u.ONESEC,v=u.EPOCHJD,g=r(24040),y=r(94336).E9,m=/^\s*(-?\d\d\d\d|\d\d)(-(\d?\d)(-(\d?\d)([ Tt]([01]?\d|2[0-3])(:([0-5]\d)(:([0-5]\d(\.\d+)?))?(Z|z|[+\-]\d\d(:?\d\d)?)?)?)?)?)?\s*$/m,x=/^\s*(-?\d\d\d\d|\d\d)(-(\d?\di?)(-(\d?\d)([ Tt]([01]?\d|2[0-3])(:([0-5]\d)(:([0-5]\d(\.\d+)?))?(Z|z|[+\-]\d\d(:?\d\d)?)?)?)?)?)?\s*$/m,b=(new Date).getFullYear()-70;function _(t){return t&&g.componentsRegistry.calendars&&"string"==typeof t&&"gregorian"!==t}function w(t,e){return String(t+Math.pow(10,e)).substr(1)}e.dateTick0=function(t,r){var n=function(t,e){return _(t)?e?g.getComponentMethod("calendars","CANONICAL_SUNDAY")[t]:g.getComponentMethod("calendars","CANONICAL_TICK")[t]:e?"2000-01-02":"2000-01-01"}(t,!!r);if(r<2)return n;var i=e.dateTime2ms(n,t);return i+=f*(r-1),e.ms2DateTime(i,0,t)},e.dfltRange=function(t){return _(t)?g.getComponentMethod("calendars","DFLTRANGE")[t]:["2000-01-01","2001-01-01"]},e.isJSDate=function(t){return"object"==typeof t&&null!==t&&"function"==typeof t.getTime},e.dateTime2ms=function(t,r){if(e.isJSDate(t)){var a=t.getTimezoneOffset()*p,o=(t.getUTCMinutes()-t.getMinutes())*p+(t.getUTCSeconds()-t.getSeconds())*d+(t.getUTCMilliseconds()-t.getMilliseconds());if(o){var s=3*p;a=a-s/2+l(o-a+s/2,s)}return(t=Number(t)-a)>=n&&t<=i?t:c}if("string"!=typeof t&&"number"!=typeof t)return c;t=String(t);var u=_(r),y=t.charAt(0);!u||"G"!==y&&"g"!==y||(t=t.substr(1),r="");var w=u&&"chinese"===r.substr(0,7),T=t.match(w?x:m);if(!T)return c;var k=T[1],A=T[3]||"1",M=Number(T[5]||1),S=Number(T[7]||0),E=Number(T[9]||0),L=Number(T[11]||0);if(u){if(2===k.length)return c;var C;k=Number(k);try{var P=g.getComponentMethod("calendars","getCal")(r);if(w){var O="i"===A.charAt(A.length-1);A=parseInt(A,10),C=P.newDate(k,P.toMonthIndex(k,A,O),M)}else C=P.newDate(k,Number(A),M)}catch(t){return c}return C?(C.toJD()-v)*f+S*h+E*p+L*d:c}k=2===k.length?(Number(k)+2e3-b)%100+b:Number(k),A-=1;var I=new Date(Date.UTC(2e3,A,M,S,E));return I.setUTCFullYear(k),I.getUTCMonth()!==A||I.getUTCDate()!==M?c:I.getTime()+L*d},n=e.MIN_MS=e.dateTime2ms("-9999"),i=e.MAX_MS=e.dateTime2ms("9999-12-31 23:59:59.9999"),e.isDateTime=function(t,r){return e.dateTime2ms(t,r)!==c};var T=90*f,k=3*h,A=5*p;function M(t,e,r,n,i){if((e||r||n||i)&&(t+=" "+w(e,2)+":"+w(r,2),(n||i)&&(t+=":"+w(n,2),i))){for(var a=4;i%10==0;)a-=1,i/=10;t+="."+w(i,a)}return t}e.ms2DateTime=function(t,e,r){if("number"!=typeof t||!(t>=n&&t<=i))return c;e||(e=0);var a,o,s,u,m,x,b=Math.floor(10*l(t+.05,1)),w=Math.round(t-b/10);if(_(r)){var S=Math.floor(w/f)+v,E=Math.floor(l(t,f));try{a=g.getComponentMethod("calendars","getCal")(r).fromJD(S).formatDate("yyyy-mm-dd")}catch(t){a=y("G%Y-%m-%d")(new Date(w))}if("-"===a.charAt(0))for(;a.length<11;)a="-0"+a.substr(1);else for(;a.length<10;)a="0"+a;o=e<T?Math.floor(E/h):0,s=e<T?Math.floor(E%h/p):0,u=e<k?Math.floor(E%p/d):0,m=e<A?E%d*10+b:0}else x=new Date(w),a=y("%Y-%m-%d")(x),o=e<T?x.getUTCHours():0,s=e<T?x.getUTCMinutes():0,u=e<k?x.getUTCSeconds():0,m=e<A?10*x.getUTCMilliseconds()+b:0;return M(a,o,s,u,m)},e.ms2DateTimeLocal=function(t){if(!(t>=n+f&&t<=i-f))return c;var e=Math.floor(10*l(t+.05,1)),r=new Date(Math.round(t-e/10));return M(a("%Y-%m-%d")(r),r.getHours(),r.getMinutes(),r.getSeconds(),10*r.getUTCMilliseconds()+e)},e.cleanDate=function(t,r,n){if(t===c)return r;if(e.isJSDate(t)||"number"==typeof t&&isFinite(t)){if(_(n))return s.error("JS Dates and milliseconds are incompatible with world calendars",t),r;if(!(t=e.ms2DateTimeLocal(+t))&&void 0!==r)return r}else if(!e.isDateTime(t,n))return s.error("unrecognized date",t),r;return t};var S=/%\d?f/g,E=/%h/g,L={1:"1",2:"1",3:"2",4:"2"};function C(t,e,r,n){t=t.replace(S,(function(t){var r=Math.min(+t.charAt(1)||6,6);return(e/1e3%1+2).toFixed(r).substr(2).replace(/0+$/,"")||"0"}));var i=new Date(Math.floor(e+.05));if(t=t.replace(E,(function(){return L[r("%q")(i)]})),_(n))try{t=g.getComponentMethod("calendars","worldCalFmt")(t,e,n)}catch(t){return"Invalid"}return r(t)(i)}var P=[59,59.9,59.99,59.999,59.9999];e.formatDate=function(t,e,r,n,i,a){if(i=_(i)&&i,!e)if("y"===r)e=a.year;else if("m"===r)e=a.month;else{if("d"!==r)return function(t,e){var r=l(t+.05,f),n=w(Math.floor(r/h),2)+":"+w(l(Math.floor(r/p),60),2);if("M"!==e){o(e)||(e=0);var i=(100+Math.min(l(t/d,60),P[e])).toFixed(e).substr(1);e>0&&(i=i.replace(/0+$/,"").replace(/[\.]$/,"")),n+=":"+i}return n}(t,r)+"\n"+C(a.dayMonthYear,t,n,i);e=a.dayMonth+"\n"+a.year}return C(e,t,n,i)};var O=3*f;e.incrementMonth=function(t,e,r){r=_(r)&&r;var n=l(t,f);if(t=Math.round(t-n),r)try{var i=Math.round(t/f)+v,a=g.getComponentMethod("calendars","getCal")(r),o=a.fromJD(i);return e%12?a.add(o,e,"m"):a.add(o,e/12,"y"),(o.toJD()-v)*f+n}catch(e){s.error("invalid ms "+t+" in calendar "+r)}var u=new Date(t+O);return u.setUTCMonth(u.getUTCMonth()+e)+n-O},e.findExactDates=function(t,e){for(var r,n,i=0,a=0,s=0,l=0,u=_(e)&&g.getComponentMethod("calendars","getCal")(e),c=0;c<t.length;c++)if(n=t[c],o(n)){if(!(n%f))if(u)try{1===(r=u.fromJD(n/f+v)).day()?1===r.month()?i++:a++:s++}catch(t){}else 1===(r=new Date(n)).getUTCDate()?0===r.getUTCMonth()?i++:a++:s++}else l++;s+=a+=i;var h=t.length-l;return{exactYears:i/h,exactMonths:a/h,exactDays:s/h}}},52200:function(t,e,r){"use strict";var n=r(33428),i=r(24248),a=r(52248),o=r(36524);function s(t){var e=t&&t.parentNode;e&&e.removeChild(t)}function l(t,e,r){var n="plotly.js-style-"+t,a=document.getElementById(n);a||((a=document.createElement("style")).setAttribute("id",n),a.appendChild(document.createTextNode("")),document.head.appendChild(a));var o=a.sheet;o.insertRule?o.insertRule(e+"{"+r+"}",0):o.addRule?o.addRule(e,r,0):i.warn("addStyleRule failed")}function u(t){var e=window.getComputedStyle(t,null),r=e.getPropertyValue("-webkit-transform")||e.getPropertyValue("-moz-transform")||e.getPropertyValue("-ms-transform")||e.getPropertyValue("-o-transform")||e.getPropertyValue("transform");return"none"===r?null:r.replace("matrix","").replace("3d","").slice(1,-1).split(",").map((function(t){return+t}))}function c(t){for(var e=[];f(t);)e.push(t),t=t.parentNode;return e}function f(t){return t&&(t instanceof Element||t instanceof HTMLElement)}t.exports={getGraphDiv:function(t){var e;if("string"==typeof t){if(null===(e=document.getElementById(t)))throw new Error("No DOM element with id '"+t+"' exists on the page.");return e}if(null==t)throw new Error("DOM element provided is null or undefined");return t},isPlotDiv:function(t){var e=n.select(t);return e.node()instanceof HTMLElement&&e.size()&&e.classed("js-plotly-plot")},removeElement:s,addStyleRule:function(t,e){l("global",t,e)},addRelatedStyleRule:l,deleteRelatedStyleRule:function(t){var e="plotly.js-style-"+t,r=document.getElementById(e);r&&s(r)},getFullTransformMatrix:function(t){var e=c(t),r=[1,0,0,0,0,1,0,0,0,0,1,0,0,0,0,1];return e.forEach((function(t){var e=u(t);if(e){var n=a.convertCssMatrix(e);r=o.multiply(r,r,n)}})),r},getElementTransformMatrix:u,getElementAndAncestors:c,equalDomRects:function(t,e){return t&&e&&t.top===e.top&&t.left===e.left&&t.right===e.right&&t.bottom===e.bottom}}},95924:function(t,e,r){"use strict";var n=r(61252).EventEmitter,i={init:function(t){if(t._ev instanceof n)return t;var e=new n,r=new n;return t._ev=e,t._internalEv=r,t.on=e.on.bind(e),t.once=e.once.bind(e),t.removeListener=e.removeListener.bind(e),t.removeAllListeners=e.removeAllListeners.bind(e),t._internalOn=r.on.bind(r),t._internalOnce=r.once.bind(r),t._removeInternalListener=r.removeListener.bind(r),t._removeAllInternalListeners=r.removeAllListeners.bind(r),t.emit=function(n,i){"undefined"!=typeof jQuery&&jQuery(t).trigger(n,i),e.emit(n,i),r.emit(n,i)},t},triggerHandler:function(t,e,r){var n,i;"undefined"!=typeof jQuery&&(n=jQuery(t).triggerHandler(e,r));var a=t._ev;if(!a)return n;var o,s=a._events[e];if(!s)return n;function l(t){return t.listener?(a.removeListener(e,t.listener),t.fired?void 0:(t.fired=!0,t.listener.apply(a,[r]))):t.apply(a,[r])}for(s=Array.isArray(s)?s:[s],o=0;o<s.length-1;o++)l(s[o]);return i=l(s[o]),void 0!==n?n:i},purge:function(t){return delete t._ev,delete t.on,delete t.once,delete t.removeListener,delete t.removeAllListeners,delete t.emit,delete t._ev,delete t._internalEv,delete t._internalOn,delete t._internalOnce,delete t._removeInternalListener,delete t._removeAllInternalListeners,t}};t.exports=i},92880:function(t,e,r){"use strict";var n=r(63620),i=Array.isArray;function a(t,e,r,o){var s,l,u,c,f,h,p,d=t[0],v=t.length;if(2===v&&i(d)&&i(t[1])&&0===d.length){if(p=function(t,e){var r,n;for(r=0;r<t.length;r++){if(null!==(n=t[r])&&"object"==typeof n)return!1;void 0!==n&&(e[r]=n)}return!0}(t[1],d),p)return d;d.splice(0,d.length)}for(var g=1;g<v;g++)for(l in s=t[g])u=d[l],c=s[l],o&&i(c)?d[l]=c:e&&c&&(n(c)||(f=i(c)))?(f?(f=!1,h=u&&i(u)?u:[]):h=u&&n(u)?u:{},d[l]=a([h,c],e,r,o)):(void 0!==c||r)&&(d[l]=c);return d}e.extendFlat=function(){return a(arguments,!1,!1,!1)},e.extendDeep=function(){return a(arguments,!0,!1,!1)},e.extendDeepAll=function(){return a(arguments,!0,!0,!1)},e.extendDeepNoArrays=function(){return a(arguments,!0,!1,!0)}},68944:function(t){"use strict";t.exports=function(t){for(var e={},r=[],n=0,i=0;i<t.length;i++){var a=t[i];1!==e[a]&&(e[a]=1,r[n++]=a)}return r}},43880:function(t){"use strict";function e(t){return!0===t.visible}function r(t){var e=t[0].trace;return!0===e.visible&&0!==e._length}t.exports=function(t){for(var n,i=(n=t,Array.isArray(n)&&Array.isArray(n[0])&&n[0][0]&&n[0][0].trace?r:e),a=[],o=0;o<t.length;o++){var s=t[o];i(s)&&a.push(s)}return a}},27144:function(t,e,r){"use strict";var n=r(33428),i=r(36116),a=r(40440),o=r(77844),s=r(42428),l=r(35536),u=r(24248),c=r(63620),f=r(22296),h=r(92065),p=Object.keys(i),d={"ISO-3":l,"USA-states":l,"country names":function(t){for(var e=0;e<p.length;e++){var r=p[e];if(new RegExp(i[r]).test(t.trim().toLowerCase()))return r}return u.log("Unrecognized country name: "+t+"."),!1}};function v(t){var e=t.geojson,r=window.PlotlyGeoAssets||{},n="string"==typeof e?r[e]:e;return c(n)?n:(u.error("Oops ... something went wrong when fetching "+e),!1)}t.exports={locationToFeature:function(t,e,r){if(!e||"string"!=typeof e)return!1;var n,i,a,o=d[t](e);if(o){if("USA-states"===t)for(n=[],a=0;a<r.length;a++)(i=r[a]).properties&&i.properties.gu&&"USA"===i.properties.gu&&n.push(i);else n=r;for(a=0;a<n.length;a++)if((i=n[a]).id===o)return i;u.log(["Location with id",o,"does not have a matching topojson feature at this resolution."].join(" "))}return!1},feature2polygons:function(t){var e,r,n,i,a=t.geometry,o=a.coordinates,s=t.id,l=[];function u(t){for(var e=0;e<t.length-1;e++)if(t[e][0]>0&&t[e+1][0]<0)return e;return null}switch(e="RUS"===s||"FJI"===s?function(t){var e;if(null===u(t))e=t;else for(e=new Array(t.length),i=0;i<t.length;i++)e[i]=[t[i][0]<0?t[i][0]+360:t[i][0],t[i][1]];l.push(h.tester(e))}:"ATA"===s?function(t){var e=u(t);if(null===e)return l.push(h.tester(t));var r=new Array(t.length+1),n=0;for(i=0;i<t.length;i++)i>e?r[n++]=[t[i][0]+360,t[i][1]]:i===e?(r[n++]=t[i],r[n++]=[t[i][0],-90]):r[n++]=t[i];var a=h.tester(r);a.pts.pop(),l.push(a)}:function(t){l.push(h.tester(t))},a.type){case"MultiPolygon":for(r=0;r<o.length;r++)for(n=0;n<o[r].length;n++)e(o[r][n]);break;case"Polygon":for(r=0;r<o.length;r++)e(o[r])}return l},getTraceGeojson:v,extractTraceFeature:function(t){var e=t[0].trace,r=v(e);if(!r)return!1;var n,i={},s=[];for(n=0;n<e._length;n++){var l=t[n];(l.loc||0===l.loc)&&(i[l.loc]=l)}function c(t){var r=f(t,e.featureidkey||"id").get(),n=i[r];if(n){var l=t.geometry;if("Polygon"===l.type||"MultiPolygon"===l.type){var c={type:"Feature",id:r,geometry:l,properties:{}};c.properties.ct=function(t){var e,r=t.geometry;if("MultiPolygon"===r.type)for(var n=r.coordinates,i=0,s=0;s<n.length;s++){var l={type:"Polygon",coordinates:n[s]},u=a.default(l);u>i&&(i=u,e=l)}else e=r;return o.default(e).geometry.coordinates}(c),n.fIn=t,n.fOut=c,s.push(c)}else u.log(["Location",n.loc,"does not have a valid GeoJSON geometry.","Traces with locationmode *geojson-id* only support","*Polygon* and *MultiPolygon* geometries."].join(" "))}delete i[r]}switch(r.type){case"FeatureCollection":var h=r.features;for(n=0;n<h.length;n++)c(h[n]);break;case"Feature":c(r);break;default:return u.warn(["Invalid GeoJSON type",(r.type||"none")+".","Traces with locationmode *geojson-id* only support","*FeatureCollection* and *Feature* types."].join(" ")),!1}for(var p in i)u.log(["Location *"+p+"*","does not have a matching feature with id-key","*"+e.featureidkey+"*."].join(" "));return s},fetchTraceGeoData:function(t){var e=window.PlotlyGeoAssets||{},r=[];function i(t){return new Promise((function(r,i){n.json(t,(function(n,a){if(n){delete e[t];var o=404===n.status?'GeoJSON at URL "'+t+'" does not exist.':"Unexpected error while fetching from "+t;return i(new Error(o))}return e[t]=a,r(a)}))}))}function a(t){return new Promise((function(r,n){var i=0,a=setInterval((function(){return e[t]&&"pending"!==e[t]?(clearInterval(a),r(e[t])):i>100?(clearInterval(a),n("Unexpected error while fetching from "+t)):void i++}),50)}))}for(var o=0;o<t.length;o++){var s=t[o][0].trace.geojson;"string"==typeof s&&(e[s]?"pending"===e[s]&&r.push(a(s)):(e[s]="pending",r.push(i(s))))}return r},computeBbox:function(t){return s.default(t)}}},44808:function(t,e,r){"use strict";var n=r(39032).BADNUM;e.calcTraceToLineCoords=function(t){for(var e=t[0].trace.connectgaps,r=[],i=[],a=0;a<t.length;a++){var o=t[a].lonlat;o[0]!==n?i.push(o):!e&&i.length>0&&(r.push(i),i=[])}return i.length>0&&r.push(i),r},e.makeLine=function(t){return 1===t.length?{type:"LineString",coordinates:t[0]}:{type:"MultiLineString",coordinates:t}},e.makePolygon=function(t){if(1===t.length)return{type:"Polygon",coordinates:t};for(var e=new Array(t.length),r=0;r<t.length;r++)e[r]=[t[r]];return{type:"MultiPolygon",coordinates:e}},e.makeBlank=function(){return{type:"Point",coordinates:[]}}},92348:function(t,e,r){"use strict";var n,i,a,o=r(20435).mod;function s(t,e,r,n,i,a,o,s){var l=r-t,u=i-t,c=o-i,f=n-e,h=a-e,p=s-a,d=l*p-c*f;if(0===d)return null;var v=(u*p-c*h)/d,g=(u*f-l*h)/d;return g<0||g>1||v<0||v>1?null:{x:t+l*v,y:e+f*v}}function l(t,e,r,n,i){var a=n*t+i*e;if(a<0)return n*n+i*i;if(a>r){var o=n-t,s=i-e;return o*o+s*s}var l=n*e-i*t;return l*l/r}e.segmentsIntersect=s,e.segmentDistance=function(t,e,r,n,i,a,o,u){if(s(t,e,r,n,i,a,o,u))return 0;var c=r-t,f=n-e,h=o-i,p=u-a,d=c*c+f*f,v=h*h+p*p,g=Math.min(l(c,f,d,i-t,a-e),l(c,f,d,o-t,u-e),l(h,p,v,t-i,e-a),l(h,p,v,r-i,n-a));return Math.sqrt(g)},e.getTextLocation=function(t,e,r,s){if(t===i&&s===a||(n={},i=t,a=s),n[r])return n[r];var l=t.getPointAtLength(o(r-s/2,e)),u=t.getPointAtLength(o(r+s/2,e)),c=Math.atan((u.y-l.y)/(u.x-l.x)),f=t.getPointAtLength(o(r,e)),h={x:(4*f.x+l.x+u.x)/6,y:(4*f.y+l.y+u.y)/6,theta:c};return n[r]=h,h},e.clearLocationCache=function(){i=null},e.getVisibleSegment=function(t,e,r){var n,i,a=e.left,o=e.right,s=e.top,l=e.bottom,u=0,c=t.getTotalLength(),f=c;function h(e){var r=t.getPointAtLength(e);0===e?n=r:e===c&&(i=r);var u=r.x<a?a-r.x:r.x>o?r.x-o:0,f=r.y<s?s-r.y:r.y>l?r.y-l:0;return Math.sqrt(u*u+f*f)}for(var p=h(u);p;){if((u+=p+r)>f)return;p=h(u)}for(p=h(f);p;){if(u>(f-=p+r))return;p=h(f)}return{min:u,max:f,len:f-u,total:c,isClosed:0===u&&f===c&&Math.abs(n.x-i.x)<.1&&Math.abs(n.y-i.y)<.1}},e.findPointOnPath=function(t,e,r,n){for(var i,a,o,s=(n=n||{}).pathLength||t.getTotalLength(),l=n.tolerance||.001,u=n.iterationLimit||30,c=t.getPointAtLength(0)[r]>t.getPointAtLength(s)[r]?-1:1,f=0,h=0,p=s;f<u;){if(i=(h+p)/2,o=(a=t.getPointAtLength(i))[r]-e,Math.abs(o)<l)return a;c*o>0?p=i:h=i,f++}return a}},33040:function(t,e,r){"use strict";var n=r(38248),i=r(49760),a=r(72160),o=r(8932),s=r(22548).defaultLine,l=r(38116).isArrayOrTypedArray,u=a(s);function c(t,e){var r=t;return r[3]*=e,r}function f(t){if(n(t))return u;var e=a(t);return e.length?e:u}function h(t){return n(t)?t:1}t.exports={formatColor:function(t,e,r){var n=t.color;n&&n._inputArray&&(n=n._inputArray);var i,s,p,d,v,g=l(n),y=l(e),m=o.extractOpts(t),x=[];if(i=void 0!==m.colorscale?o.makeColorScaleFuncFromTrace(t):f,s=g?function(t,e){return void 0===t[e]?u:a(i(t[e]))}:f,p=y?function(t,e){return void 0===t[e]?1:h(t[e])}:h,g||y)for(var b=0;b<r;b++)d=s(n,b),v=p(e,b),x[b]=c(d,v);else x=c(a(n),e);return x},parseColorScale:function(t){var e=o.extractOpts(t),r=e.colorscale;return e.reversescale&&(r=o.flipScale(e.colorscale)),r.map((function(t){var e=t[0],r=i(t[1]).toRgb();return{index:e,rgb:[r.r,r.g,r.b,r.a]}}))}}},71688:function(t,e,r){"use strict";var n=r(35536);function i(t){return[t]}t.exports={keyFun:function(t){return t.key},repeat:i,descend:n,wrap:i,unwrap:function(t){return t[0]}}},35536:function(t){"use strict";t.exports=function(t){return t}},1396:function(t){"use strict";t.exports=function(t,e){if(!e)return t;var r=1/Math.abs(e),n=r>1?(r*t+r*e)/r:t+e,i=String(n).length;if(i>16){var a=String(e).length;if(i>=String(t).length+a){var o=parseFloat(n).toPrecision(12);-1===o.indexOf("e+")&&(n=+o)}}return n}},3400:function(t,e,r){"use strict";var n=r(33428),i=r(94336).E9,a=r(57624).E9,o=r(38248),s=r(39032),l=s.FP_SAFE,u=-l,c=s.BADNUM,f=t.exports={};f.adjustFormat=function(t){return!t||/^\d[.]\df/.test(t)||/[.]\d%/.test(t)?t:"0.f"===t?"~f":/^\d%/.test(t)?"~%":/^\ds/.test(t)?"~s":!/^[~,.0$]/.test(t)&&/[&fps]/.test(t)?"~"+t:t};var h={};f.warnBadFormat=function(t){var e=String(t);h[e]||(h[e]=1,f.warn('encountered bad format: "'+e+'"'))},f.noFormat=function(t){return String(t)},f.numberFormat=function(t){var e;try{e=a(f.adjustFormat(t))}catch(e){return f.warnBadFormat(t),f.noFormat}return e},f.nestedProperty=r(22296),f.keyedContainer=r(37804),f.relativeAttr=r(23193),f.isPlainObject=r(63620),f.toLogRange=r(36896),f.relinkPrivateKeys=r(51528);var p=r(38116);f.isArrayBuffer=p.isArrayBuffer,f.isTypedArray=p.isTypedArray,f.isArrayOrTypedArray=p.isArrayOrTypedArray,f.isArray1D=p.isArray1D,f.ensureArray=p.ensureArray,f.concat=p.concat,f.maxRowLength=p.maxRowLength,f.minRowLength=p.minRowLength;var d=r(20435);f.mod=d.mod,f.modHalf=d.modHalf;var v=r(63064);f.valObjectMeta=v.valObjectMeta,f.coerce=v.coerce,f.coerce2=v.coerce2,f.coerceFont=v.coerceFont,f.coercePattern=v.coercePattern,f.coerceHoverinfo=v.coerceHoverinfo,f.coerceSelectionMarkerOpacity=v.coerceSelectionMarkerOpacity,f.validate=v.validate;var g=r(67555);f.dateTime2ms=g.dateTime2ms,f.isDateTime=g.isDateTime,f.ms2DateTime=g.ms2DateTime,f.ms2DateTimeLocal=g.ms2DateTimeLocal,f.cleanDate=g.cleanDate,f.isJSDate=g.isJSDate,f.formatDate=g.formatDate,f.incrementMonth=g.incrementMonth,f.dateTick0=g.dateTick0,f.dfltRange=g.dfltRange,f.findExactDates=g.findExactDates,f.MIN_MS=g.MIN_MS,f.MAX_MS=g.MAX_MS;var y=r(14952);f.findBin=y.findBin,f.sorterAsc=y.sorterAsc,f.sorterDes=y.sorterDes,f.distinctVals=y.distinctVals,f.roundUp=y.roundUp,f.sort=y.sort,f.findIndexOfMin=y.findIndexOfMin,f.sortObjectKeys=r(95376);var m=r(63084);f.aggNums=m.aggNums,f.len=m.len,f.mean=m.mean,f.median=m.median,f.midRange=m.midRange,f.variance=m.variance,f.stdev=m.stdev,f.interp=m.interp;var x=r(52248);f.init2dArray=x.init2dArray,f.transposeRagged=x.transposeRagged,f.dot=x.dot,f.translationMatrix=x.translationMatrix,f.rotationMatrix=x.rotationMatrix,f.rotationXYMatrix=x.rotationXYMatrix,f.apply3DTransform=x.apply3DTransform,f.apply2DTransform=x.apply2DTransform,f.apply2DTransform2=x.apply2DTransform2,f.convertCssMatrix=x.convertCssMatrix,f.inverseTransformMatrix=x.inverseTransformMatrix;var b=r(11864);f.deg2rad=b.deg2rad,f.rad2deg=b.rad2deg,f.angleDelta=b.angleDelta,f.angleDist=b.angleDist,f.isFullCircle=b.isFullCircle,f.isAngleInsideSector=b.isAngleInsideSector,f.isPtInsideSector=b.isPtInsideSector,f.pathArc=b.pathArc,f.pathSector=b.pathSector,f.pathAnnulus=b.pathAnnulus;var _=r(98308);f.isLeftAnchor=_.isLeftAnchor,f.isCenterAnchor=_.isCenterAnchor,f.isRightAnchor=_.isRightAnchor,f.isTopAnchor=_.isTopAnchor,f.isMiddleAnchor=_.isMiddleAnchor,f.isBottomAnchor=_.isBottomAnchor;var w=r(92348);f.segmentsIntersect=w.segmentsIntersect,f.segmentDistance=w.segmentDistance,f.getTextLocation=w.getTextLocation,f.clearLocationCache=w.clearLocationCache,f.getVisibleSegment=w.getVisibleSegment,f.findPointOnPath=w.findPointOnPath;var T=r(92880);f.extendFlat=T.extendFlat,f.extendDeep=T.extendDeep,f.extendDeepAll=T.extendDeepAll,f.extendDeepNoArrays=T.extendDeepNoArrays;var k=r(24248);f.log=k.log,f.warn=k.warn,f.error=k.error;var A=r(53756);f.counterRegex=A.counter;var M=r(91200);f.throttle=M.throttle,f.throttleDone=M.done,f.clearThrottle=M.clear;var S=r(52200);function E(t){var e={};for(var r in t)for(var n=t[r],i=0;i<n.length;i++)e[n[i]]=+r;return e}f.getGraphDiv=S.getGraphDiv,f.isPlotDiv=S.isPlotDiv,f.removeElement=S.removeElement,f.addStyleRule=S.addStyleRule,f.addRelatedStyleRule=S.addRelatedStyleRule,f.deleteRelatedStyleRule=S.deleteRelatedStyleRule,f.getFullTransformMatrix=S.getFullTransformMatrix,f.getElementTransformMatrix=S.getElementTransformMatrix,f.getElementAndAncestors=S.getElementAndAncestors,f.equalDomRects=S.equalDomRects,f.clearResponsive=r(75352),f.preserveDrawingBuffer=r(34296),f.makeTraceGroups=r(30988),f._=r(98356),f.notifier=r(41792),f.filterUnique=r(68944),f.filterVisible=r(43880),f.pushUnique=r(52416),f.increment=r(1396),f.cleanNumber=r(54037),f.ensureNumber=function(t){return o(t)?(t=Number(t))>l||t<u?c:t:c},f.isIndex=function(t,e){return!(void 0!==e&&t>=e)&&o(t)&&t>=0&&t%1==0},f.noop=r(16628),f.identity=r(35536),f.repeat=function(t,e){for(var r=new Array(e),n=0;n<e;n++)r[n]=t;return r},f.swapAttrs=function(t,e,r,n){r||(r="x"),n||(n="y");for(var i=0;i<e.length;i++){var a=e[i],o=f.nestedProperty(t,a.replace("?",r)),s=f.nestedProperty(t,a.replace("?",n)),l=o.get();o.set(s.get()),s.set(l)}},f.raiseToTop=function(t){t.parentNode.appendChild(t)},f.cancelTransition=function(t){return t.transition().duration(0)},f.constrain=function(t,e,r){return e>r?Math.max(r,Math.min(e,t)):Math.max(e,Math.min(r,t))},f.bBoxIntersect=function(t,e,r){return r=r||0,t.left<=e.right+r&&e.left<=t.right+r&&t.top<=e.bottom+r&&e.top<=t.bottom+r},f.simpleMap=function(t,e,r,n,i){for(var a=t.length,o=new Array(a),s=0;s<a;s++)o[s]=e(t[s],r,n,i);return o},f.randstr=function t(e,r,n,i){if(n||(n=16),void 0===r&&(r=24),r<=0)return"0";var a,o,s=Math.log(Math.pow(2,r))/Math.log(n),l="";for(a=2;s===1/0;a*=2)s=Math.log(Math.pow(2,r/a))/Math.log(n)*a;var u=s-Math.floor(s);for(a=0;a<Math.floor(s);a++)l=Math.floor(Math.random()*n).toString(n)+l;u&&(o=Math.pow(n,u),l=Math.floor(Math.random()*o).toString(n)+l);var c=parseInt(l,n);return e&&e[l]||c!==1/0&&c>=Math.pow(2,r)?i>10?(f.warn("randstr failed uniqueness"),l):t(e,r,n,(i||0)+1):l},f.OptionControl=function(t,e){t||(t={}),e||(e="opt");var r={optionList:[],_newoption:function(n){n[e]=t,r[n.name]=n,r.optionList.push(n)}};return r["_"+e]=t,r},f.smooth=function(t,e){if((e=Math.round(e)||0)<2)return t;var r,n,i,a,o=t.length,s=2*o,l=2*e-1,u=new Array(l),c=new Array(o);for(r=0;r<l;r++)u[r]=(1-Math.cos(Math.PI*(r+1)/e))/(2*e);for(r=0;r<o;r++){for(a=0,n=0;n<l;n++)(i=r+n+1-e)<-o?i-=s*Math.round(i/s):i>=s&&(i-=s*Math.floor(i/s)),i<0?i=-1-i:i>=o&&(i=s-1-i),a+=t[i]*u[n];c[r]=a}return c},f.syncOrAsync=function(t,e,r){var n;function i(){return f.syncOrAsync(t,e,r)}for(;t.length;)if((n=(0,t.splice(0,1)[0])(e))&&n.then)return n.then(i);return r&&r(e)},f.stripTrailingSlash=function(t){return"/"===t.substr(-1)?t.substr(0,t.length-1):t},f.noneOrAll=function(t,e,r){if(t){var n,i=!1,a=!0;for(n=0;n<r.length;n++)null!=t[r[n]]?i=!0:a=!1;if(i&&!a)for(n=0;n<r.length;n++)t[r[n]]=e[r[n]]}},f.mergeArray=function(t,e,r,n){var i="function"==typeof n;if(f.isArrayOrTypedArray(t))for(var a=Math.min(t.length,e.length),o=0;o<a;o++){var s=t[o];e[o][r]=i?n(s):s}},f.mergeArrayCastPositive=function(t,e,r){return f.mergeArray(t,e,r,(function(t){var e=+t;return isFinite(e)&&e>0?e:0}))},f.fillArray=function(t,e,r,n){if(n=n||f.identity,f.isArrayOrTypedArray(t))for(var i=0;i<e.length;i++)e[i][r]=n(t[i])},f.castOption=function(t,e,r,n){n=n||f.identity;var i=f.nestedProperty(t,r).get();return f.isArrayOrTypedArray(i)?Array.isArray(e)&&f.isArrayOrTypedArray(i[e[0]])?n(i[e[0]][e[1]]):n(i[e]):i},f.extractOption=function(t,e,r,n){if(r in t)return t[r];var i=f.nestedProperty(e,n).get();return Array.isArray(i)?void 0:i},f.tagSelected=function(t,e,r){var n,i,a=e.selectedpoints,o=e._indexToPoints;o&&(n=E(o));for(var s=0;s<a.length;s++){var l=a[s];if(f.isIndex(l)||f.isArrayOrTypedArray(l)&&f.isIndex(l[0])&&f.isIndex(l[1])){var u=n?n[l]:l,c=r?r[u]:u;void 0!==(i=c)&&i<t.length&&(t[c].selected=1)}}},f.selIndices2selPoints=function(t){var e=t.selectedpoints,r=t._indexToPoints;if(r){for(var n=E(r),i=[],a=0;a<e.length;a++){var o=e[a];if(f.isIndex(o)){var s=n[o];f.isIndex(s)&&i.push(s)}}return i}return e},f.getTargetArray=function(t,e){var r=e.target;if("string"==typeof r&&r){var n=f.nestedProperty(t,r).get();return!!f.isArrayOrTypedArray(n)&&n}return!!f.isArrayOrTypedArray(r)&&r},f.minExtend=function t(e,r,n){var i={};"object"!=typeof r&&(r={});var a,o,s,l="pieLike"===n?-1:3,u=Object.keys(e);for(a=0;a<u.length;a++)s=e[o=u[a]],"_"!==o.charAt(0)&&"function"!=typeof s&&("module"===o?i[o]=s:Array.isArray(s)?i[o]="colorscale"===o||-1===l?s.slice():s.slice(0,l):f.isTypedArray(s)?i[o]=-1===l?s.subarray():s.subarray(0,l):i[o]=s&&"object"==typeof s?t(e[o],r[o],n):s);for(u=Object.keys(r),a=0;a<u.length;a++)"object"==typeof(s=r[o=u[a]])&&o in i&&"object"==typeof i[o]||(i[o]=s);return i},f.titleCase=function(t){return t.charAt(0).toUpperCase()+t.substr(1)},f.containsAny=function(t,e){for(var r=0;r<e.length;r++)if(-1!==t.indexOf(e[r]))return!0;return!1},f.isIE=function(){return void 0!==window.navigator.msSaveBlob};var L=/Version\/[\d\.]+.*Safari/;f.isSafari=function(){return L.test(window.navigator.userAgent)};var C=/iPad|iPhone|iPod/;f.isIOS=function(){return C.test(window.navigator.userAgent)};var P=/Firefox\/(\d+)\.\d+/;f.getFirefoxVersion=function(){var t=P.exec(window.navigator.userAgent);if(t&&2===t.length){var e=parseInt(t[1]);if(!isNaN(e))return e}return null},f.isD3Selection=function(t){return t instanceof n.selection},f.ensureSingle=function(t,e,r,n){var i=t.select(e+(r?"."+r:""));if(i.size())return i;var a=t.append(e);return r&&a.classed(r,!0),n&&a.call(n),a},f.ensureSingleById=function(t,e,r,n){var i=t.select(e+"#"+r);if(i.size())return i;var a=t.append(e).attr("id",r);return n&&a.call(n),a},f.objectFromPath=function(t,e){for(var r,n=t.split("."),i=r={},a=0;a<n.length;a++){var o=n[a],s=null,l=n[a].match(/(.*)\[([0-9]+)\]/);l?(o=l[1],s=l[2],r=r[o]=[],a===n.length-1?r[s]=e:r[s]={},r=r[s]):(a===n.length-1?r[o]=e:r[o]={},r=r[o])}return i};var O=/^([^\[\.]+)\.(.+)?/,I=/^([^\.]+)\[([0-9]+)\](\.)?(.+)?/;function z(t){return"__"===t.slice(0,2)}f.expandObjectPaths=function(t){var e,r,n,i,a,o,s;if("object"==typeof t&&!Array.isArray(t))for(r in t)if(t.hasOwnProperty(r))if(e=r.match(O)){if(i=t[r],z(n=e[1]))continue;delete t[r],t[n]=f.extendDeepNoArrays(t[n]||{},f.objectFromPath(r,f.expandObjectPaths(i))[n])}else if(e=r.match(I)){if(i=t[r],z(n=e[1]))continue;if(a=parseInt(e[2]),delete t[r],t[n]=t[n]||[],"."===e[3])s=e[4],o=t[n][a]=t[n][a]||{},f.extendDeepNoArrays(o,f.objectFromPath(s,f.expandObjectPaths(i)));else{if(z(n))continue;t[n][a]=f.expandObjectPaths(i)}}else{if(z(r))continue;t[r]=f.expandObjectPaths(t[r])}return t},f.numSeparate=function(t,e,r){if(r||(r=!1),"string"!=typeof e||0===e.length)throw new Error("Separator string required for formatting!");"number"==typeof t&&(t=String(t));var n=/(\d+)(\d{3})/,i=e.charAt(0),a=e.charAt(1),o=t.split("."),s=o[0],l=o.length>1?i+o[1]:"";if(a&&(o.length>1||s.length>4||r))for(;n.test(s);)s=s.replace(n,"$1"+a+"$2");return s+l},f.TEMPLATE_STRING_REGEX=/%{([^\s%{}:]*)([:|\|][^}]*)?}/g;var D=/^\w*$/;f.templateString=function(t,e){var r={};return t.replace(f.TEMPLATE_STRING_REGEX,(function(t,n){var i;return D.test(n)?i=e[n]:(r[n]=r[n]||f.nestedProperty(e,n).get,i=r[n]()),f.isValidTextValue(i)?i:""}))};var R={max:10,count:0,name:"hovertemplate"};f.hovertemplateString=function(){return U.apply(R,arguments)};var F={max:10,count:0,name:"texttemplate"};f.texttemplateString=function(){return U.apply(F,arguments)};var B=/^(\S+)([\*\/])(-?\d+(\.\d+)?)$/,N={max:10,count:0,name:"texttemplate",parseMultDiv:!0};f.texttemplateStringForShapes=function(){return U.apply(N,arguments)};var j=/^[:|\|]/;function U(t,e,r){var n=this,a=arguments;e||(e={});var o={};return t.replace(f.TEMPLATE_STRING_REGEX,(function(t,s,l){var u="_xother"===s||"_yother"===s,c="_xother_"===s||"_yother_"===s,h="xother_"===s||"yother_"===s,p="xother"===s||"yother"===s||u||h||c,d=s;(u||c)&&(d=d.substring(1)),(h||c)&&(d=d.substring(0,d.length-1));var v,g,y,m=null,x=null;if(n.parseMultDiv){var b=function(t){var e=t.match(B);return e?{key:e[1],op:e[2],number:Number(e[3])}:{key:t,op:null,number:null}}(d);d=b.key,m=b.op,x=b.number}if(p){if(void 0===(v=e[d]))return""}else for(y=3;y<a.length;y++)if(g=a[y]){if(g.hasOwnProperty(d)){v=g[d];break}if(D.test(d)||(v=f.nestedProperty(g,d).get(),(v=o[d]||f.nestedProperty(g,d).get())&&(o[d]=v)),void 0!==v)break}if(void 0!==v&&("*"===m&&(v*=x),"/"===m&&(v/=x)),void 0===v&&n)return n.count<n.max&&(f.warn("Variable '"+d+"' in "+n.name+" could not be found!"),v=t),n.count===n.max&&f.warn("Too many "+n.name+" warnings - additional warnings will be suppressed"),n.count++,t;if(l){var _;if(":"===l[0]&&(_=r?r.numberFormat:f.numberFormat,""!==v&&(v=_(l.replace(j,""))(v))),"|"===l[0]){_=r?r.timeFormat:i;var w=f.dateTime2ms(v);v=f.formatDate(w,l.replace(j,""),!1,_)}}else{var T=d+"Label";e.hasOwnProperty(T)&&(v=e[T])}return p&&(v="("+v+")",(u||c)&&(v=" "+v),(h||c)&&(v+=" ")),v}))}f.subplotSort=function(t,e){for(var r=Math.min(t.length,e.length)+1,n=0,i=0,a=0;a<r;a++){var o=t.charCodeAt(a)||0,s=e.charCodeAt(a)||0,l=o>=48&&o<=57,u=s>=48&&s<=57;if(l&&(n=10*n+o-48),u&&(i=10*i+s-48),!l||!u){if(n!==i)return n-i;if(o!==s)return o-s}}return i-n};var V=2e9;f.seedPseudoRandom=function(){V=2e9},f.pseudoRandom=function(){var t=V;return V=(69069*V+1)%4294967296,Math.abs(V-t)<429496729?f.pseudoRandom():V/4294967296},f.fillText=function(t,e,r){var n=Array.isArray(r)?function(t){r.push(t)}:function(t){r.text=t},i=f.extractOption(t,e,"htx","hovertext");if(f.isValidTextValue(i))return n(i);var a=f.extractOption(t,e,"tx","text");return f.isValidTextValue(a)?n(a):void 0},f.isValidTextValue=function(t){return t||0===t},f.formatPercent=function(t,e){e=e||0;for(var r=(Math.round(100*t*Math.pow(10,e))*Math.pow(.1,e)).toFixed(e)+"%",n=0;n<e;n++)-1!==r.indexOf(".")&&(r=(r=r.replace("0%","%")).replace(".%","%"));return r},f.isHidden=function(t){var e=window.getComputedStyle(t).display;return!e||"none"===e},f.strTranslate=function(t,e){return t||e?"translate("+t+","+e+")":""},f.strRotate=function(t){return t?"rotate("+t+")":""},f.strScale=function(t){return 1!==t?"scale("+t+")":""},f.getTextTransform=function(t){var e=t.noCenter,r=t.textX,n=t.textY,i=t.targetX,a=t.targetY,o=t.anchorX||0,s=t.anchorY||0,l=t.rotate,u=t.scale;return u?u>1&&(u=1):u=0,f.strTranslate(i-u*(r+o),a-u*(n+s))+f.strScale(u)+(l?"rotate("+l+(e?"":" "+r+" "+n)+")":"")},f.setTransormAndDisplay=function(t,e){t.attr("transform",f.getTextTransform(e)),t.style("display",e.scale?null:"none")},f.ensureUniformFontSize=function(t,e){var r=f.extendFlat({},e);return r.size=Math.max(e.size,t._fullLayout.uniformtext.minsize||0),r},f.join2=function(t,e,r){var n=t.length;return n>1?t.slice(0,-1).join(e)+r+t[n-1]:t.join(e)},f.bigFont=function(t){return Math.round(1.2*t)};var q=f.getFirefoxVersion(),H=null!==q&&q<86;f.getPositionFromD3Event=function(){return H?[n.event.layerX,n.event.layerY]:[n.event.offsetX,n.event.offsetY]}},63620:function(t){"use strict";t.exports=function(t){return window&&window.process&&window.process.versions?"[object Object]"===Object.prototype.toString.call(t):"[object Object]"===Object.prototype.toString.call(t)&&Object.getPrototypeOf(t).hasOwnProperty("hasOwnProperty")}},37804:function(t,e,r){"use strict";var n=r(22296),i=/^\w*$/;t.exports=function(t,e,r,a){var o,s,l;r=r||"name",a=a||"value";var u={};e&&e.length?(l=n(t,e),s=l.get()):s=t,e=e||"";var c={};if(s)for(o=0;o<s.length;o++)c[s[o][r]]=o;var f=i.test(a),h={set:function(t,e){var i=null===e?4:0;if(!s){if(!l||4===i)return;s=[],l.set(s)}var o=c[t];if(void 0===o){if(4===i)return;i|=3,o=s.length,c[t]=o}else e!==(f?s[o][a]:n(s[o],a).get())&&(i|=2);var p=s[o]=s[o]||{};return p[r]=t,f?p[a]=e:n(p,a).set(e),null!==e&&(i&=-5),u[o]=u[o]|i,h},get:function(t){if(s){var e=c[t];return void 0===e?void 0:f?s[e][a]:n(s[e],a).get()}},rename:function(t,e){var n=c[t];return void 0===n||(u[n]=1|u[n],c[e]=n,delete c[t],s[n][r]=e),h},remove:function(t){var e=c[t];if(void 0===e)return h;var i=s[e];if(Object.keys(i).length>2)return u[e]=2|u[e],h.set(t,null);if(f){for(o=e;o<s.length;o++)u[o]=3|u[o];for(o=e;o<s.length;o++)c[s[o][r]]--;s.splice(e,1),delete c[t]}else n(i,a).set(null),u[e]=6|u[e];return h},constructUpdate:function(){for(var t,i,o={},l=Object.keys(u),c=0;c<l.length;c++)i=l[c],t=e+"["+i+"]",s[i]?(1&u[i]&&(o[t+"."+r]=s[i][r]),2&u[i]&&(o[t+"."+a]=f?4&u[i]?null:s[i][a]:4&u[i]?null:n(s[i],a).get())):o[t]=null;return o}};return h}},98356:function(t,e,r){"use strict";var n=r(24040);t.exports=function(t,e){for(var r=t._context.locale,i=0;i<2;i++){for(var a=t._context.locales,o=0;o<2;o++){var s=(a[r]||{}).dictionary;if(s){var l=s[e];if(l)return l}a=n.localeRegistry}var u=r.split("-")[0];if(u===r)break;r=u}return e}},24248:function(t,e,r){"use strict";var n=r(20556).dfltConfig,i=r(41792),a=t.exports={};a.log=function(){var t;if(n.logging>1){var e=["LOG:"];for(t=0;t<arguments.length;t++)e.push(arguments[t]);console.trace.apply(console,e)}if(n.notifyOnLogging>1){var r=[];for(t=0;t<arguments.length;t++)r.push(arguments[t]);i(r.join("<br>"),"long")}},a.warn=function(){var t;if(n.logging>0){var e=["WARN:"];for(t=0;t<arguments.length;t++)e.push(arguments[t]);console.trace.apply(console,e)}if(n.notifyOnLogging>0){var r=[];for(t=0;t<arguments.length;t++)r.push(arguments[t]);i(r.join("<br>"),"stick")}},a.error=function(){var t;if(n.logging>0){var e=["ERROR:"];for(t=0;t<arguments.length;t++)e.push(arguments[t]);console.error.apply(console,e)}if(n.notifyOnLogging>0){var r=[];for(t=0;t<arguments.length;t++)r.push(arguments[t]);i(r.join("<br>"),"stick")}}},30988:function(t,e,r){"use strict";var n=r(33428);t.exports=function(t,e,r){var i=t.selectAll("g."+r.replace(/\s/g,".")).data(e,(function(t){return t[0].trace.uid}));i.exit().remove(),i.enter().append("g").attr("class",r),i.order();var a=t.classed("rangeplot")?"nodeRangePlot3":"node3";return i.each((function(t){t[0][a]=n.select(this)})),i}},52248:function(t,e,r){"use strict";var n=r(36524);e.init2dArray=function(t,e){for(var r=new Array(t),n=0;n<t;n++)r[n]=new Array(e);return r},e.transposeRagged=function(t){var e,r,n=0,i=t.length;for(e=0;e<i;e++)n=Math.max(n,t[e].length);var a=new Array(n);for(e=0;e<n;e++)for(a[e]=new Array(i),r=0;r<i;r++)a[e][r]=t[r][e];return a},e.dot=function(t,r){if(!t.length||!r.length||t.length!==r.length)return null;var n,i,a=t.length;if(t[0].length)for(n=new Array(a),i=0;i<a;i++)n[i]=e.dot(t[i],r);else if(r[0].length){var o=e.transposeRagged(r);for(n=new Array(o.length),i=0;i<o.length;i++)n[i]=e.dot(t,o[i])}else for(n=0,i=0;i<a;i++)n+=t[i]*r[i];return n},e.translationMatrix=function(t,e){return[[1,0,t],[0,1,e],[0,0,1]]},e.rotationMatrix=function(t){var e=t*Math.PI/180;return[[Math.cos(e),-Math.sin(e),0],[Math.sin(e),Math.cos(e),0],[0,0,1]]},e.rotationXYMatrix=function(t,r,n){return e.dot(e.dot(e.translationMatrix(r,n),e.rotationMatrix(t)),e.translationMatrix(-r,-n))},e.apply3DTransform=function(t){return function(){var r=arguments,n=1===arguments.length?r[0]:[r[0],r[1],r[2]||0];return e.dot(t,[n[0],n[1],n[2],1]).slice(0,3)}},e.apply2DTransform=function(t){return function(){var r=arguments;3===r.length&&(r=r[0]);var n=1===arguments.length?r[0]:[r[0],r[1]];return e.dot(t,[n[0],n[1],1]).slice(0,2)}},e.apply2DTransform2=function(t){var r=e.apply2DTransform(t);return function(t){return r(t.slice(0,2)).concat(r(t.slice(2,4)))}},e.convertCssMatrix=function(t){if(t){var e=t.length;if(16===e)return t;if(6===e)return[t[0],t[1],0,0,t[2],t[3],0,0,0,0,1,0,t[4],t[5],0,1]}return[1,0,0,0,0,1,0,0,0,0,1,0,0,0,0,1]},e.inverseTransformMatrix=function(t){var e=[];return n.invert(e,t),[[e[0],e[1],e[2],e[3]],[e[4],e[5],e[6],e[7]],[e[8],e[9],e[10],e[11]],[e[12],e[13],e[14],e[15]]]}},20435:function(t){"use strict";t.exports={mod:function(t,e){var r=t%e;return r<0?r+e:r},modHalf:function(t,e){return Math.abs(t)>e/2?t-Math.round(t/e)*e:t}}},22296:function(t,e,r){"use strict";var n=r(38248),i=r(38116).isArrayOrTypedArray;function a(t,e){return function(){var r,n,o,s,l,u=t;for(s=0;s<e.length-1;s++){if(-1===(r=e[s])){for(n=!0,o=[],l=0;l<u.length;l++)o[l]=a(u[l],e.slice(s+1))(),o[l]!==o[0]&&(n=!1);return n?o[0]:o}if("number"==typeof r&&!i(u))return;if("object"!=typeof(u=u[r])||null===u)return}if("object"==typeof u&&null!==u&&null!==(o=u[e[s]]))return o}}t.exports=function(t,e){if(n(e))e=String(e);else if("string"!=typeof e||"[-1]"===e.substr(e.length-4))throw"bad property string";var r,i,o,s,u=e.split(".");for(s=0;s<u.length;s++)if("__"===String(u[s]).slice(0,2))throw"bad property string";for(s=0;s<u.length;){if(r=String(u[s]).match(/^([^\[\]]*)((\[\-?[0-9]*\])+)$/)){if(r[1])u[s]=r[1];else{if(0!==s)throw"bad property string";u.splice(0,1)}for(i=r[2].substr(1,r[2].length-2).split("]["),o=0;o<i.length;o++)s++,u.splice(s,0,Number(i[o]))}s++}return"object"!=typeof t?function(t,e,r){return{set:function(){throw"bad container"},get:function(){},astr:e,parts:r,obj:t}}(t,e,u):{set:l(t,u,e),get:a(t,u),astr:e,parts:u,obj:t}};var o=/(^|\.)args\[/;function s(t,e){return void 0===t||null===t&&!e.match(o)}function l(t,e,r){return function(n){var a,o,l=t,h="",p=[[t,h]],d=s(n,r);for(o=0;o<e.length-1;o++){if("number"==typeof(a=e[o])&&!i(l))throw"array index but container is not an array";if(-1===a){if(d=!c(l,e.slice(o+1),n,r))break;return}if(!f(l,a,e[o+1],d))break;if("object"!=typeof(l=l[a])||null===l)throw"container is not an object";h=u(h,a),p.push([l,h])}if(d){if(o===e.length-1&&(delete l[e[o]],Array.isArray(l)&&+e[o]==l.length-1))for(;l.length&&void 0===l[l.length-1];)l.pop()}else l[e[o]]=n}}function u(t,e){var r=e;return n(e)?r="["+e+"]":t&&(r="."+e),t+r}function c(t,e,r,n){var a,o=i(r),u=!0,c=r,h=n.replace("-1",0),p=!o&&s(r,h),d=e[0];for(a=0;a<t.length;a++)h=n.replace("-1",a),o&&(p=s(c=r[a%r.length],h)),p&&(u=!1),f(t,a,d,p)&&l(t[a],e,n.replace("-1",a))(c);return u}function f(t,e,r,n){if(void 0===t[e]){if(n)return!1;t[e]="number"==typeof r?[]:{}}return!0}},16628:function(t){"use strict";t.exports=function(){}},41792:function(t,e,r){"use strict";var n=r(33428),i=r(38248),a=[];t.exports=function(t,e){if(-1===a.indexOf(t)){a.push(t);var r=1e3;i(e)?r=e:"long"===e&&(r=3e3);var o=n.select("body").selectAll(".plotly-notifier").data([0]);o.enter().append("div").classed("plotly-notifier",!0),o.selectAll(".notifier-note").data(a).enter().append("div").classed("notifier-note",!0).style("opacity",0).each((function(t){var i=n.select(this);i.append("button").classed("notifier-close",!0).html("&times;").on("click",(function(){i.transition().call(s)}));for(var a=i.append("p"),o=t.split(/<br\s*\/?>/g),l=0;l<o.length;l++)l&&a.append("br"),a.append("span").text(o[l]);"stick"===e?i.transition().duration(350).style("opacity",1):i.transition().duration(700).style("opacity",1).transition().delay(r).call(s)}))}function s(t){t.duration(700).style("opacity",0).each("end",(function(t){var e=a.indexOf(t);-1!==e&&a.splice(e,1),n.select(this).remove()}))}}},72213:function(t,e,r){"use strict";var n=r(93972),i="data-savedcursor";t.exports=function(t,e){var r=t.attr(i);if(e){if(!r){for(var a=(t.attr("class")||"").split(" "),o=0;o<a.length;o++){var s=a[o];0===s.indexOf("cursor-")&&t.attr(i,s.substr(7)).classed(s,!1)}t.attr(i)||t.attr(i,"!!")}n(t,e)}else r&&(t.attr(i,null),"!!"===r?n(t):n(t,r))}},92065:function(t,e,r){"use strict";var n=r(52248).dot,i=r(39032).BADNUM,a=t.exports={};a.tester=function(t){var e,r=t.slice(),n=r[0][0],a=n,o=r[0][1],s=o;for(r[r.length-1][0]===r[0][0]&&r[r.length-1][1]===r[0][1]||r.push(r[0]),e=1;e<r.length;e++)n=Math.min(n,r[e][0]),a=Math.max(a,r[e][0]),o=Math.min(o,r[e][1]),s=Math.max(s,r[e][1]);var l,u=!1;5===r.length&&(r[0][0]===r[1][0]?r[2][0]===r[3][0]&&r[0][1]===r[3][1]&&r[1][1]===r[2][1]&&(u=!0,l=function(t){return t[0]===r[0][0]}):r[0][1]===r[1][1]&&r[2][1]===r[3][1]&&r[0][0]===r[3][0]&&r[1][0]===r[2][0]&&(u=!0,l=function(t){return t[1]===r[0][1]}));var c=!0,f=r[0];for(e=1;e<r.length;e++)if(f[0]!==r[e][0]||f[1]!==r[e][1]){c=!1;break}return{xmin:n,xmax:a,ymin:o,ymax:s,pts:r,contains:u?function(t,e){var r=t[0],u=t[1];return!(r===i||r<n||r>a||u===i||u<o||u>s||e&&l(t))}:function(t,e){var l=t[0],u=t[1];if(l===i||l<n||l>a||u===i||u<o||u>s)return!1;var c,f,h,p,d,v=r.length,g=r[0][0],y=r[0][1],m=0;for(c=1;c<v;c++)if(f=g,h=y,g=r[c][0],y=r[c][1],!(l<(p=Math.min(f,g))||l>Math.max(f,g)||u>Math.max(h,y)))if(u<Math.min(h,y))l!==p&&m++;else{if(u===(d=g===f?u:h+(l-f)*(y-h)/(g-f)))return 1!==c||!e;u<=d&&l!==p&&m++}return m%2==1},isRect:u,degenerate:c}},a.isSegmentBent=function(t,e,r,i){var a,o,s,l=t[e],u=[t[r][0]-l[0],t[r][1]-l[1]],c=n(u,u),f=Math.sqrt(c),h=[-u[1]/f,u[0]/f];for(a=e+1;a<r;a++)if(o=[t[a][0]-l[0],t[a][1]-l[1]],(s=n(o,u))<0||s>c||Math.abs(n(o,h))>i)return!0;return!1},a.filter=function(t,e){var r=[t[0]],n=0,i=0;function o(o){t.push(o);var s=r.length,l=n;r.splice(i+1);for(var u=l+1;u<t.length;u++)(u===t.length-1||a.isSegmentBent(t,l,u+1,e))&&(r.push(t[u]),r.length<s-2&&(n=u,i=r.length-1),l=u)}return t.length>1&&o(t.pop()),{addPt:o,raw:t,filtered:r}}},5048:function(t,e,r){"use strict";var n=r(16576),i=r(28624);t.exports=function(t,e,a){var o=t._fullLayout,s=!0;return o._glcanvas.each((function(n){if(n.regl)n.regl.preloadCachedCode(a);else if(!n.pick||o._has("parcoords")){try{n.regl=i({canvas:this,attributes:{antialias:!n.pick,preserveDrawingBuffer:!0},pixelRatio:t._context.plotGlPixelRatio||r.g.devicePixelRatio,extensions:e||[],cachedCode:a||{}})}catch(t){s=!1}n.regl||(s=!1),s&&this.addEventListener("webglcontextlost",(function(e){t&&t.emit&&t.emit("plotly_webglcontextlost",{event:e,layer:n.key})}),!1)}})),s||n({container:o._glcontainer.node()}),s}},34296:function(t,e,r){"use strict";var n=r(38248),i=r(25928);t.exports=function(t){var e;if("string"!=typeof(e=t&&t.hasOwnProperty("userAgent")?t.userAgent:function(){var t;return"undefined"!=typeof navigator&&(t=navigator.userAgent),t&&t.headers&&"string"==typeof t.headers["user-agent"]&&(t=t.headers["user-agent"]),t}()))return!0;var r=i({ua:{headers:{"user-agent":e}},tablet:!0,featureDetect:!1});if(!r)for(var a=e.split(" "),o=1;o<a.length;o++)if(-1!==a[o].indexOf("Safari"))for(var s=o-1;s>-1;s--){var l=a[s];if("Version/"===l.substr(0,8)){var u=l.substr(8).split(".")[0];if(n(u)&&(u=+u),u>=13)return!0}}return r}},52416:function(t){"use strict";t.exports=function(t,e){if(e instanceof RegExp){for(var r=e.toString(),n=0;n<t.length;n++)if(t[n]instanceof RegExp&&t[n].toString()===r)return t;t.push(e)}else!e&&0!==e||-1!==t.indexOf(e)||t.push(e);return t}},94552:function(t,e,r){"use strict";var n=r(3400),i=r(20556).dfltConfig,a={add:function(t,e,r,n,a){var o,s;t.undoQueue=t.undoQueue||{index:0,queue:[],sequence:!1},s=t.undoQueue.index,t.autoplay?t.undoQueue.inSequence||(t.autoplay=!1):(!t.undoQueue.sequence||t.undoQueue.beginSequence?(o={undo:{calls:[],args:[]},redo:{calls:[],args:[]}},t.undoQueue.queue.splice(s,t.undoQueue.queue.length-s,o),t.undoQueue.index+=1):o=t.undoQueue.queue[s-1],t.undoQueue.beginSequence=!1,o&&(o.undo.calls.unshift(e),o.undo.args.unshift(r),o.redo.calls.push(n),o.redo.args.push(a)),t.undoQueue.queue.length>i.queueLength&&(t.undoQueue.queue.shift(),t.undoQueue.index--))},startSequence:function(t){t.undoQueue=t.undoQueue||{index:0,queue:[],sequence:!1},t.undoQueue.sequence=!0,t.undoQueue.beginSequence=!0},stopSequence:function(t){t.undoQueue=t.undoQueue||{index:0,queue:[],sequence:!1},t.undoQueue.sequence=!1,t.undoQueue.beginSequence=!1},undo:function(t){var e,r;if(!(void 0===t.undoQueue||isNaN(t.undoQueue.index)||t.undoQueue.index<=0)){for(t.undoQueue.index--,e=t.undoQueue.queue[t.undoQueue.index],t.undoQueue.inSequence=!0,r=0;r<e.undo.calls.length;r++)a.plotDo(t,e.undo.calls[r],e.undo.args[r]);t.undoQueue.inSequence=!1,t.autoplay=!1}},redo:function(t){var e,r;if(!(void 0===t.undoQueue||isNaN(t.undoQueue.index)||t.undoQueue.index>=t.undoQueue.queue.length)){for(e=t.undoQueue.queue[t.undoQueue.index],t.undoQueue.inSequence=!0,r=0;r<e.redo.calls.length;r++)a.plotDo(t,e.redo.calls[r],e.redo.args[r]);t.undoQueue.inSequence=!1,t.autoplay=!1,t.undoQueue.index++}},plotDo:function(t,e,r){t.autoplay=!0,r=function(t,e){for(var r,i=[],a=0;a<e.length;a++)r=e[a],i[a]=r===t?r:"object"==typeof r?Array.isArray(r)?n.extendDeep([],r):n.extendDeepAll({},r):r;return i}(t,r),e.apply(null,r)}};t.exports=a},53756:function(t,e){"use strict";e.counter=function(t,e,r,n){var i=(e||"")+(r?"":"$"),a=!1===n?"":"^";return"xy"===t?new RegExp(a+"x([2-9]|[1-9][0-9]+)?y([2-9]|[1-9][0-9]+)?"+i):new RegExp(a+t+"([2-9]|[1-9][0-9]+)?"+i)}},23193:function(t){"use strict";var e=/^(.*)(\.[^\.\[\]]+|\[\d\])$/,r=/^[^\.\[\]]+$/;t.exports=function(t,n){for(;n;){var i=t.match(e);if(i)t=i[1];else{if(!t.match(r))throw new Error("bad relativeAttr call:"+[t,n]);t=""}if("^"!==n.charAt(0))break;n=n.slice(1)}return t&&"["!==n.charAt(0)?t+"."+n:t+n}},51528:function(t,e,r){"use strict";var n=r(38116).isArrayOrTypedArray,i=r(63620);t.exports=function t(e,r){for(var a in r){var o=r[a],s=e[a];if(s!==o)if("_"===a.charAt(0)||"function"==typeof o){if(a in e)continue;e[a]=o}else if(n(o)&&n(s)&&i(o[0])){if("customdata"===a||"ids"===a)continue;for(var l=Math.min(o.length,s.length),u=0;u<l;u++)s[u]!==o[u]&&i(o[u])&&i(s[u])&&t(s[u],o[u])}else i(o)&&i(s)&&(t(s,o),Object.keys(s).length||delete e[a])}}},14952:function(t,e,r){"use strict";var n=r(38248),i=r(24248),a=r(35536),o=r(39032).BADNUM,s=1e-9;function l(t,e){return t<e}function u(t,e){return t<=e}function c(t,e){return t>e}function f(t,e){return t>=e}e.findBin=function(t,e,r){if(n(e.start))return r?Math.ceil((t-e.start)/e.size-s)-1:Math.floor((t-e.start)/e.size+s);var a,o,h=0,p=e.length,d=0,v=p>1?(e[p-1]-e[0])/(p-1):1;for(o=v>=0?r?l:u:r?f:c,t+=v*s*(r?-1:1)*(v>=0?1:-1);h<p&&d++<100;)o(e[a=Math.floor((h+p)/2)],t)?h=a+1:p=a;return d>90&&i.log("Long binary search..."),h-1},e.sorterAsc=function(t,e){return t-e},e.sorterDes=function(t,e){return e-t},e.distinctVals=function(t){var r,n=t.slice();for(n.sort(e.sorterAsc),r=n.length-1;r>-1&&n[r]===o;r--);for(var i,a=n[r]-n[0]||1,s=a/(r||1)/1e4,l=[],u=0;u<=r;u++){var c=n[u],f=c-i;void 0===i?(l.push(c),i=c):f>s&&(a=Math.min(a,f),l.push(c),i=c)}return{vals:l,minDiff:a}},e.roundUp=function(t,e,r){for(var n,i=0,a=e.length-1,o=0,s=r?0:1,l=r?1:0,u=r?Math.ceil:Math.floor;i<a&&o++<100;)e[n=u((i+a)/2)]<=t?i=n+s:a=n-l;return e[i]},e.sort=function(t,e){for(var r=0,n=0,i=1;i<t.length;i++){var a=e(t[i],t[i-1]);if(a<0?r=1:a>0&&(n=1),r&&n)return t.sort(e)}return n?t:t.reverse()},e.findIndexOfMin=function(t,e){e=e||a;for(var r,n=1/0,i=0;i<t.length;i++){var o=e(t[i]);o<n&&(n=o,r=i)}return r}},93972:function(t){"use strict";t.exports=function(t,e){(t.attr("class")||"").split(" ").forEach((function(e){0===e.indexOf("cursor-")&&t.classed(e,!1)})),e&&t.classed("cursor-"+e,!0)}},16576:function(t,e,r){"use strict";var n=r(76308),i=function(){};t.exports=function(t){for(var e in t)"function"==typeof t[e]&&(t[e]=i);t.destroy=function(){t.container.parentNode.removeChild(t.container)};var r=document.createElement("div");r.className="no-webgl",r.style.cursor="pointer",r.style.fontSize="24px",r.style.color=n.defaults[0],r.style.position="absolute",r.style.left=r.style.top="0px",r.style.width=r.style.height="100%",r.style["background-color"]=n.lightLine,r.style["z-index"]=30;var a=document.createElement("p");return a.textContent="WebGL is not supported by your browser - visit https://get.webgl.org for more info",a.style.position="relative",a.style.top="50%",a.style.left="50%",a.style.height="30%",a.style.width="50%",a.style.margin="-15% 0 0 -25%",r.appendChild(a),t.container.appendChild(r),t.container.style.background="#FFFFFF",t.container.onclick=function(){window.open("https://get.webgl.org")},!1}},95376:function(t){"use strict";t.exports=function(t){return Object.keys(t).sort()}},63084:function(t,e,r){"use strict";var n=r(38248),i=r(38116).isArrayOrTypedArray;e.aggNums=function(t,r,a,o){var s,l;if((!o||o>a.length)&&(o=a.length),n(r)||(r=!1),i(a[0])){for(l=new Array(o),s=0;s<o;s++)l[s]=e.aggNums(t,r,a[s]);a=l}for(s=0;s<o;s++)n(r)?n(a[s])&&(r=t(+r,+a[s])):r=a[s];return r},e.len=function(t){return e.aggNums((function(t){return t+1}),0,t)},e.mean=function(t,r){return r||(r=e.len(t)),e.aggNums((function(t,e){return t+e}),0,t)/r},e.midRange=function(t){if(void 0!==t&&0!==t.length)return(e.aggNums(Math.max,null,t)+e.aggNums(Math.min,null,t))/2},e.variance=function(t,r,i){return r||(r=e.len(t)),n(i)||(i=e.mean(t,r)),e.aggNums((function(t,e){return t+Math.pow(e-i,2)}),0,t)/r},e.stdev=function(t,r,n){return Math.sqrt(e.variance(t,r,n))},e.median=function(t){var r=t.slice().sort();return e.interp(r,.5)},e.interp=function(t,e){if(!n(e))throw"n should be a finite number";if((e=e*t.length-.5)<0)return t[0];if(e>t.length-1)return t[t.length-1];var r=e%1;return r*t[Math.ceil(e)]+(1-r)*t[Math.floor(e)]}},43080:function(t,e,r){"use strict";var n=r(72160);t.exports=function(t){return t?n(t):[0,0,0,1]}},9188:function(t,e,r){"use strict";var n=r(2264),i=r(43616),a=r(3400),o=null;t.exports=function(){if(null!==o)return o;o=!1;var t=a.isIE()||a.isSafari()||a.isIOS();if(window.navigator.userAgent&&!t){var e=Array.from(n.CSS_DECLARATIONS).reverse(),r=window.CSS&&window.CSS.supports||window.supportsCSS;if("function"==typeof r)o=e.some((function(t){return r.apply(null,t)}));else{var s=i.tester.append("image").attr("style",n.STYLE),l=window.getComputedStyle(s.node()).imageRendering;o=e.some((function(t){var e=t[1];return l===e||l===e.toLowerCase()})),s.remove()}}return o}},72736:function(t,e,r){"use strict";var n=r(33428),i=r(3400),a=i.strTranslate,o=r(9616),s=r(84284).LINE_SPACING,l=/([^$]*)([$]+[^$]*[$]+)([^$]*)/;e.convertToTspans=function(t,r,g){var S=t.text(),E=!t.attr("data-notex")&&r&&r._context.typesetMath&&"undefined"!=typeof MathJax&&S.match(l),P=n.select(t.node().parentNode);if(!P.empty()){var O=t.attr("class")?t.attr("class").split(" ")[0]:"text";return O+="-math",P.selectAll("svg."+O).remove(),P.selectAll("g."+O+"-group").remove(),t.style("display",null).attr({"data-unformatted":S,"data-math":"N"}),E?(r&&r._promises||[]).push(new Promise((function(e){t.style("display","none");var r=parseInt(t.node().style.fontSize,10),o={fontSize:r};!function(t,e,r){var a,o,s,l,h=parseInt((MathJax.version||"").split(".")[0]);if(2===h||3===h){var p=function(){var r="math-output-"+i.randstr({},64),a=(l=n.select("body").append("div").attr({id:r}).style({visibility:"hidden",position:"absolute","font-size":e.fontSize+"px"}).text(t.replace(u,"\\lt ").replace(c,"\\gt "))).node();return 2===h?MathJax.Hub.Typeset(a):MathJax.typeset([a])},d=function(){var e=l.select(2===h?".MathJax_SVG":".MathJax"),a=!e.empty()&&l.select("svg").node();if(a){var o,s=a.getBoundingClientRect();o=2===h?n.select("body").select("#MathJax_SVG_glyphs"):e.select("defs"),r(e,o,s)}else i.log("There was an error in the tex syntax.",t),r();l.remove()};2===h?MathJax.Hub.Queue((function(){return o=i.extendDeepAll({},MathJax.Hub.config),s=MathJax.Hub.processSectionDelay,void 0!==MathJax.Hub.processSectionDelay&&(MathJax.Hub.processSectionDelay=0),MathJax.Hub.Config({messageStyle:"none",tex2jax:{inlineMath:f},displayAlign:"left"})}),(function(){if("SVG"!==(a=MathJax.Hub.config.menuSettings.renderer))return MathJax.Hub.setRenderer("SVG")}),p,d,(function(){if("SVG"!==a)return MathJax.Hub.setRenderer(a)}),(function(){return void 0!==s&&(MathJax.Hub.processSectionDelay=s),MathJax.Hub.Config(o)})):3===h&&(o=i.extendDeepAll({},MathJax.config),MathJax.config.tex||(MathJax.config.tex={}),MathJax.config.tex.inlineMath=f,"svg"!==(a=MathJax.config.startup.output)&&(MathJax.config.startup.output="svg"),MathJax.startup.defaultReady(),MathJax.startup.promise.then((function(){p(),d(),"svg"!==a&&(MathJax.config.startup.output=a),MathJax.config=o})))}else i.warn("No MathJax version:",MathJax.version)}(E[2],o,(function(n,i,o){P.selectAll("svg."+O).remove(),P.selectAll("g."+O+"-group").remove();var s=n&&n.select("svg");if(!s||!s.node())return I(),void e();var l=P.append("g").classed(O+"-group",!0).attr({"pointer-events":"none","data-unformatted":S,"data-math":"Y"});l.node().appendChild(s.node()),i&&i.node()&&s.node().insertBefore(i.node().cloneNode(!0),s.node().firstChild);var u=o.width,c=o.height;s.attr({class:O,height:c,preserveAspectRatio:"xMinYMin meet"}).style({overflow:"visible","pointer-events":"none"});var f=t.node().style.fill||"black",h=s.select("g");h.attr({fill:f,stroke:f});var p=h.node().getBoundingClientRect(),d=p.width,v=p.height;(d>u||v>c)&&(s.style("overflow","hidden"),d=(p=s.node().getBoundingClientRect()).width,v=p.height);var y=+t.attr("x"),m=+t.attr("y"),x=-(r||t.node().getBoundingClientRect().height)/4;if("y"===O[0])l.attr({transform:"rotate("+[-90,y,m]+")"+a(-d/2,x-v/2)});else if("l"===O[0])m=x-v/2;else if("a"===O[0]&&0!==O.indexOf("atitle"))y=0,m=x;else{var b=t.attr("text-anchor");y-=d*("middle"===b?.5:"end"===b?1:0),m=m+x-v/2}s.attr({x:y,y:m}),g&&g.call(t,l),e(l)}))}))):I(),t}function I(){P.empty()||(O=t.attr("class")+"-math",P.select("svg."+O).remove()),t.text("").style("white-space","pre");var r=function(t,e){e=e.replace(y," ");var r,a=!1,l=[],u=-1;function c(){u++;var e=document.createElementNS(o.svg,"tspan");n.select(e).attr({class:"line",dy:u*s+"em"}),t.appendChild(e),r=e;var i=l;if(l=[{node:e}],i.length>1)for(var a=1;a<i.length;a++)f(i[a])}function f(t){var e,i=t.type,a={};if("a"===i){e="a";var s=t.target,u=t.href,c=t.popup;u&&(a={"xlink:xlink:show":"_blank"===s||"_"!==s.charAt(0)?"new":"replace",target:s,"xlink:xlink:href":u},c&&(a.onclick='window.open(this.href.baseVal,this.target.baseVal,"'+c+'");return false;'))}else e="tspan";t.style&&(a.style=t.style);var f=document.createElementNS(o.svg,e);if("sup"===i||"sub"===i){g(r,v),r.appendChild(f);var h=document.createElementNS(o.svg,"tspan");g(h,v),n.select(h).attr("dy",d[i]),a.dy=p[i],r.appendChild(f),r.appendChild(h)}else r.appendChild(f);n.select(f).attr(a),r=t.node=f,l.push(t)}function g(t,e){t.appendChild(document.createTextNode(e))}function S(t){if(1!==l.length){var n=l.pop();t!==n.type&&i.log("Start tag <"+n.type+"> doesnt match end tag <"+t+">. Pretending it did match.",e),r=l[l.length-1].node}else i.log("Ignoring unexpected end tag </"+t+">.",e)}b.test(e)?c():(r=t,l=[{node:t}]);for(var E=e.split(m),P=0;P<E.length;P++){var O=E[P],I=O.match(x),z=I&&I[2].toLowerCase(),D=h[z];if("br"===z)c();else if(void 0===D)g(r,L(O));else if(I[1])S(z);else{var R=I[4],F={type:z},B=A(R,_);if(B?(B=B.replace(M,"$1 fill:"),D&&(B+=";"+D)):D&&(B=D),B&&(F.style=B),"a"===z){a=!0;var N=A(R,w);if(N){var j=C(N);j&&(F.href=j,F.target=A(R,T)||"_blank",F.popup=A(R,k))}}f(F)}}return a}(t.node(),S);r&&t.style("pointer-events","all"),e.positionText(t),g&&g.call(t)}};var u=/(<|&lt;|&#60;)/g,c=/(>|&gt;|&#62;)/g,f=[["$","$"],["\\(","\\)"]],h={sup:"font-size:70%",sub:"font-size:70%",b:"font-weight:bold",i:"font-style:italic",a:"cursor:pointer",span:"",em:"font-style:italic;font-weight:bold"},p={sub:"0.3em",sup:"-0.6em"},d={sub:"-0.21em",sup:"0.42em"},v="​",g=["http:","https:","mailto:","",void 0,":"],y=e.NEWLINES=/(\r\n?|\n)/g,m=/(<[^<>]*>)/,x=/<(\/?)([^ >]*)(\s+(.*))?>/i,b=/<br(\s+.*)?>/i;e.BR_TAG_ALL=/<br(\s+.*)?>/gi;var _=/(^|[\s"'])style\s*=\s*("([^"]*);?"|'([^']*);?')/i,w=/(^|[\s"'])href\s*=\s*("([^"]*)"|'([^']*)')/i,T=/(^|[\s"'])target\s*=\s*("([^"\s]*)"|'([^'\s]*)')/i,k=/(^|[\s"'])popup\s*=\s*("([\w=,]*)"|'([\w=,]*)')/i;function A(t,e){if(!t)return null;var r=t.match(e),n=r&&(r[3]||r[4]);return n&&L(n)}var M=/(^|;)\s*color:/;e.plainText=function(t,e){for(var r=void 0!==(e=e||{}).len&&-1!==e.len?e.len:1/0,n=void 0!==e.allowedTags?e.allowedTags:["br"],i=t.split(m),a=[],o="",s=0,l=0;l<i.length;l++){var u=i[l],c=u.match(x),f=c&&c[2].toLowerCase();if(f)-1!==n.indexOf(f)&&(a.push(u),o=f);else{var h=u.length;if(s+h<r)a.push(u),s+=h;else if(s<r){var p=r-s;o&&("br"!==o||p<=3||h<=3)&&a.pop(),r>3?a.push(u.substr(0,p-3)+"..."):a.push(u.substr(0,p));break}o=""}}return a.join("")};var S={mu:"μ",amp:"&",lt:"<",gt:">",nbsp:" ",times:"×",plusmn:"±",deg:"°"},E=/&(#\d+|#x[\da-fA-F]+|[a-z]+);/g;function L(t){return t.replace(E,(function(t,e){return("#"===e.charAt(0)?function(t){if(!(t>1114111)){var e=String.fromCodePoint;if(e)return e(t);var r=String.fromCharCode;return t<=65535?r(t):r(55232+(t>>10),t%1024+56320)}}("x"===e.charAt(1)?parseInt(e.substr(2),16):parseInt(e.substr(1),10)):S[e])||t}))}function C(t){var e=encodeURI(decodeURI(t)),r=document.createElement("a"),n=document.createElement("a");r.href=t,n.href=e;var i=r.protocol,a=n.protocol;return-1!==g.indexOf(i)&&-1!==g.indexOf(a)?e:""}function P(t,e,r){var n,a,o,s=r.horizontalAlign,l=r.verticalAlign||"top",u=t.node().getBoundingClientRect(),c=e.node().getBoundingClientRect();return a="bottom"===l?function(){return u.bottom-n.height}:"middle"===l?function(){return u.top+(u.height-n.height)/2}:function(){return u.top},o="right"===s?function(){return u.right-n.width}:"center"===s?function(){return u.left+(u.width-n.width)/2}:function(){return u.left},function(){n=this.node().getBoundingClientRect();var t=o()-c.left,e=a()-c.top,s=r.gd||{};if(r.gd){s._fullLayout._calcInverseTransform(s);var l=i.apply3DTransform(s._fullLayout._invTransform)(t,e);t=l[0],e=l[1]}return this.style({top:e+"px",left:t+"px","z-index":1e3}),this}}e.convertEntities=L,e.sanitizeHTML=function(t){t=t.replace(y," ");for(var e=document.createElement("p"),r=e,i=[],a=t.split(m),o=0;o<a.length;o++){var s=a[o],l=s.match(x),u=l&&l[2].toLowerCase();if(u in h)if(l[1])i.length&&(r=i.pop());else{var c=l[4],f=A(c,_),p=f?{style:f}:{};if("a"===u){var d=A(c,w);if(d){var v=C(d);if(v){p.href=v;var g=A(c,T);g&&(p.target=g)}}}var b=document.createElement(u);r.appendChild(b),n.select(b).attr(p),r=b,i.push(b)}else r.appendChild(document.createTextNode(L(s)))}return e.innerHTML},e.lineCount=function(t){return t.selectAll("tspan.line").size()||1},e.positionText=function(t,e,r){return t.each((function(){var t=n.select(this);function i(e,r){return void 0===r?null===(r=t.attr(e))&&(t.attr(e,0),r=0):t.attr(e,r),r}var a=i("x",e),o=i("y",r);"text"===this.nodeName&&t.selectAll("tspan.line").attr({x:a,y:o})}))};var O="1px ";e.makeTextShadow=function(t){return O+O+O+t+", -"+O+"-"+O+O+t+", "+O+"-"+O+O+t+", -"+O+O+O+t},e.makeEditable=function(t,e){var r=e.gd,i=e.delegate,a=n.dispatch("edit","input","cancel"),o=i||t;if(t.style({"pointer-events":i?"none":"all"}),1!==t.size())throw new Error("boo");function s(){var i,s,u,c,f;i=n.select(r).select(".svg-container"),s=i.append("div"),u=t.node().style,c=parseFloat(u.fontSize||12),void 0===(f=e.text)&&(f=t.attr("data-unformatted")),s.classed("plugin-editable editable",!0).style({position:"absolute","font-family":u.fontFamily||"Arial","font-size":c,color:e.fill||u.fill||"black",opacity:1,"background-color":e.background||"transparent",outline:"#ffffff33 1px solid",margin:[-c/8+1,0,0,-1].join("px ")+"px",padding:"0","box-sizing":"border-box"}).attr({contenteditable:!0}).text(f).call(P(t,i,e)).on("blur",(function(){r._editing=!1,t.text(this.textContent).style({opacity:1});var e,i=n.select(this).attr("class");(e=i?"."+i.split(" ")[0]+"-math-group":"[class*=-math-group]")&&n.select(t.node().parentNode).select(e).style({opacity:0});var o=this.textContent;n.select(this).transition().duration(0).remove(),n.select(document).on("mouseup",null),a.edit.call(t,o)})).on("focus",(function(){var t=this;r._editing=!0,n.select(document).on("mouseup",(function(){if(n.event.target===t)return!1;document.activeElement===s.node()&&s.node().blur()}))})).on("keyup",(function(){27===n.event.which?(r._editing=!1,t.style({opacity:1}),n.select(this).style({opacity:0}).on("blur",(function(){return!1})).transition().remove(),a.cancel.call(t,this.textContent)):(a.input.call(t,this.textContent),n.select(this).call(P(t,i,e)))})).on("keydown",(function(){13===n.event.which&&this.blur()})).call(l),t.style({opacity:0});var h,p=o.attr("class");(h=p?"."+p.split(" ")[0]+"-math-group":"[class*=-math-group]")&&n.select(t.node().parentNode).select(h).style({opacity:0})}function l(t){var e=t.node(),r=document.createRange();r.selectNodeContents(e);var n=window.getSelection();n.removeAllRanges(),n.addRange(r),e.focus()}return e.immediate?s():o.on("click",s),n.rebind(t,a,"on")}},91200:function(t,e){"use strict";var r={};function n(t){t&&null!==t.timer&&(clearTimeout(t.timer),t.timer=null)}e.throttle=function(t,e,i){var a=r[t],o=Date.now();if(!a){for(var s in r)r[s].ts<o-6e4&&delete r[s];a=r[t]={ts:0,timer:null}}function l(){i(),a.ts=Date.now(),a.onDone&&(a.onDone(),a.onDone=null)}n(a),o>a.ts+e?l():a.timer=setTimeout((function(){l(),a.timer=null}),e)},e.done=function(t){var e=r[t];return e&&e.timer?new Promise((function(t){var r=e.onDone;e.onDone=function(){r&&r(),t(),e.onDone=null}})):Promise.resolve()},e.clear=function(t){if(t)n(r[t]),delete r[t];else for(var i in r)e.clear(i)}},36896:function(t,e,r){"use strict";var n=r(38248);t.exports=function(t,e){if(t>0)return Math.log(t)/Math.LN10;var r=Math.log(Math.min(e[0],e[1]))/Math.LN10;return n(r)||(r=Math.log(Math.max(e[0],e[1]))/Math.LN10-6),r}},59972:function(t,e,r){"use strict";var n=t.exports={},i=r(79552).locationmodeToLayer,a=r(55712).NO;n.getTopojsonName=function(t){return[t.scope.replace(/ /g,"-"),"_",t.resolution.toString(),"m"].join("")},n.getTopojsonPath=function(t,e){return t+e+".json"},n.getTopojsonFeatures=function(t,e){var r=i[t.locationmode],n=e.objects[r];return a(e,n).features}},11680:function(t){"use strict";t.exports={moduleType:"locale",name:"en-US",dictionary:{"Click to enter Colorscale title":"Click to enter Colorscale title"},format:{date:"%m/%d/%Y"}}},6580:function(t){"use strict";t.exports={moduleType:"locale",name:"en",dictionary:{"Click to enter Colorscale title":"Click to enter Colourscale title"},format:{days:["Sunday","Monday","Tuesday","Wednesday","Thursday","Friday","Saturday"],shortDays:["Sun","Mon","Tue","Wed","Thu","Fri","Sat"],months:["January","February","March","April","May","June","July","August","September","October","November","December"],shortMonths:["Jan","Feb","Mar","Apr","May","Jun","Jul","Aug","Sep","Oct","Nov","Dec"],periods:["AM","PM"],dateTime:"%a %b %e %X %Y",date:"%d/%m/%Y",time:"%H:%M:%S",decimal:".",thousands:",",grouping:[3],currency:["$",""],year:"%Y",month:"%b %Y",dayMonth:"%b %-d",dayMonthYear:"%b %-d, %Y"}}},69820:function(t,e,r){"use strict";var n=r(24040);t.exports=function(t){for(var e,r,i=n.layoutArrayContainers,a=n.layoutArrayRegexes,o=t.split("[")[0],s=0;s<a.length;s++)if((r=t.match(a[s]))&&0===r.index){e=r[0];break}if(e||(e=i[i.indexOf(o)]),!e)return!1;var l=t.substr(e.length);return l?!!(r=l.match(/^\[(0|[1-9][0-9]*)\](\.(.+))?$/))&&{array:e,index:Number(r[1]),property:r[3]||""}:{array:e,index:"",property:""}}},67824:function(t,e,r){"use strict";var n=r(92880).extendFlat,i=r(63620),a={valType:"flaglist",extras:["none"],flags:["calc","clearAxisTypes","plot","style","markerSize","colorbars"]},o={valType:"flaglist",extras:["none"],flags:["calc","plot","legend","ticks","axrange","layoutstyle","modebar","camera","arraydraw","colorbars"]},s=a.flags.slice().concat(["fullReplot"]),l=o.flags.slice().concat("layoutReplot");function u(t){for(var e={},r=0;r<t.length;r++)e[t[r]]=!1;return e}function c(t,e,r){var a=n({},t);for(var o in a){var s=a[o];i(s)&&(a[o]=f(s,e,0,o))}return"from-root"===r&&(a.editType=e),a}function f(t,e,r,i){if(t.valType){var a=n({},t);if(a.editType=e,Array.isArray(t.items)){a.items=new Array(t.items.length);for(var o=0;o<t.items.length;o++)a.items[o]=f(t.items[o],e)}return a}return c(t,e,"_"===i.charAt(0)?"nested":"from-root")}t.exports={traces:a,layout:o,traceFlags:function(){return u(s)},layoutFlags:function(){return u(l)},update:function(t,e){var r=e.editType;if(r&&"none"!==r)for(var n=r.split("+"),i=0;i<n.length;i++)t[n[i]]=!0},overrideAll:c}},93404:function(t,e,r){"use strict";var n=r(38248),i=r(61784),a=r(24040),o=r(3400),s=r(7316),l=r(79811),u=r(76308),c=l.cleanId,f=l.getFromTrace,h=a.traceIs;function p(t,e){var r=t[e],n=e.charAt(0);r&&"paper"!==r&&(t[e]=c(r,n,!0))}function d(t){function e(e,r){var n=t[e],i=t.title&&t.title[r];n&&!i&&(t.title||(t.title={}),t.title[r]=t[e],delete t[e])}t&&("string"!=typeof t.title&&"number"!=typeof t.title||(t.title={text:t.title}),e("titlefont","font"),e("titleposition","position"),e("titleside","side"),e("titleoffset","offset"))}function v(t){if(!o.isPlainObject(t))return!1;var e=t.name;return delete t.name,delete t.showlegend,("string"==typeof e||"number"==typeof e)&&String(e)}function g(t,e,r,n){if(r&&!n)return t;if(n&&!r)return e;if(!t.trim())return e;if(!e.trim())return t;var i,a=Math.min(t.length,e.length);for(i=0;i<a&&t.charAt(i)===e.charAt(i);i++);return t.substr(0,i).trim()}function y(t){var e="middle",r="center";return"string"==typeof t&&(-1!==t.indexOf("top")?e="top":-1!==t.indexOf("bottom")&&(e="bottom"),-1!==t.indexOf("left")?r="left":-1!==t.indexOf("right")&&(r="right")),e+" "+r}function m(t,e){return e in t&&"object"==typeof t[e]&&0===Object.keys(t[e]).length}e.clearPromiseQueue=function(t){Array.isArray(t._promises)&&t._promises.length>0&&o.log("Clearing previous rejected promises from queue."),t._promises=[]},e.cleanLayout=function(t){var r,n;t||(t={}),t.xaxis1&&(t.xaxis||(t.xaxis=t.xaxis1),delete t.xaxis1),t.yaxis1&&(t.yaxis||(t.yaxis=t.yaxis1),delete t.yaxis1),t.scene1&&(t.scene||(t.scene=t.scene1),delete t.scene1);var a=(s.subplotsRegistry.cartesian||{}).attrRegex,l=(s.subplotsRegistry.polar||{}).attrRegex,f=(s.subplotsRegistry.ternary||{}).attrRegex,h=(s.subplotsRegistry.gl3d||{}).attrRegex,v=Object.keys(t);for(r=0;r<v.length;r++){var g=v[r];if(a&&a.test(g)){var y=t[g];y.anchor&&"free"!==y.anchor&&(y.anchor=c(y.anchor)),y.overlaying&&(y.overlaying=c(y.overlaying)),y.type||(y.isdate?y.type="date":y.islog?y.type="log":!1===y.isdate&&!1===y.islog&&(y.type="linear")),"withzero"!==y.autorange&&"tozero"!==y.autorange||(y.autorange=!0,y.rangemode="tozero"),y.insiderange&&delete y.range,delete y.islog,delete y.isdate,delete y.categories,m(y,"domain")&&delete y.domain,void 0!==y.autotick&&(void 0===y.tickmode&&(y.tickmode=y.autotick?"auto":"linear"),delete y.autotick),d(y)}else if(l&&l.test(g))d(t[g].radialaxis);else if(f&&f.test(g)){var x=t[g];d(x.aaxis),d(x.baxis),d(x.caxis)}else if(h&&h.test(g)){var b=t[g],_=b.cameraposition;if(Array.isArray(_)&&4===_[0].length){var w=_[0],T=_[1],k=_[2],A=i([],w),M=[];for(n=0;n<3;++n)M[n]=T[n]+k*A[2+4*n];b.camera={eye:{x:M[0],y:M[1],z:M[2]},center:{x:T[0],y:T[1],z:T[2]},up:{x:0,y:0,z:1}},delete b.cameraposition}d(b.xaxis),d(b.yaxis),d(b.zaxis)}}var S=Array.isArray(t.annotations)?t.annotations.length:0;for(r=0;r<S;r++){var E=t.annotations[r];o.isPlainObject(E)&&(E.ref&&("paper"===E.ref?(E.xref="paper",E.yref="paper"):"data"===E.ref&&(E.xref="x",E.yref="y"),delete E.ref),p(E,"xref"),p(E,"yref"))}var L=Array.isArray(t.shapes)?t.shapes.length:0;for(r=0;r<L;r++){var C=t.shapes[r];o.isPlainObject(C)&&(p(C,"xref"),p(C,"yref"))}var P=Array.isArray(t.
```
