## Supplementary File 1 for "A shared MHC immunogenetic signal connects aging, rheumatoid arthritis, and herpes zoster through chronic high inflammatory burden–compensatory immune tolerance dysregulation": select2.min.js

```
/*! Select2 4.1.0-rc.0 | https://github.com/select2/select2/blob/master/LICENSE.md */
!function(n){"function"==typeof define&&define.amd?define(["jquery"],n):"object"==typeof module&&module.exports?module.exports=function(e,t){return void 0===t&&(t="undefined"!=typeof window?require("jquery"):require("jquery")(e)),n(t),t}:n(jQuery)}(function(t){var e,n,s,p,r,o,h,f,g,m,y,v,i,a,_,s=((u=t&&t.fn&&t.fn.select2&&t.fn.select2.amd?t.fn.select2.amd:u)&&u.requirejs||(u?n=u:u={},g={},m={},y={},v={},i=Object.prototype.hasOwnProperty,a=[].slice,_=/\.js$/,h=function(e,t){var n,s,i=c(e),r=i[0],t=t[1];return e=i[1],r&&(n=x(r=l(r,t))),r?e=n&&n.normalize?n.normalize(e,(s=t,function(e){return l(e,s)})):l(e,t):(r=(i=c(e=l(e,t)))[0],e=i[1],r&&(n=x(r))),{f:r?r+"!"+e:e,n:e,pr:r,p:n}},f={require:function(e){return w(e)},exports:function(e){var t=g[e];return void 0!==t?t:g[e]={}},module:function(e){return{id:e,uri:"",exports:g[e],config:(t=e,function(){return y&&y.config&&y.config[t]||{}})};var t}},r=function(e,t,n,s){var i,r,o,a,l,c=[],u=typeof n,d=A(s=s||e);if("undefined"==u||"function"==u){for(t=!t.length&&n.length?["require","exports","module"]:t,a=0;a<t.length;a+=1)if("require"===(r=(o=h(t[a],d)).f))c[a]=f.require(e);else if("exports"===r)c[a]=f.exports(e),l=!0;else if("module"===r)i=c[a]=f.module(e);else if(b(g,r)||b(m,r)||b(v,r))c[a]=x(r);else{if(!o.p)throw new Error(e+" missing "+r);o.p.load(o.n,w(s,!0),function(t){return function(e){g[t]=e}}(r),{}),c[a]=g[r]}u=n?n.apply(g[e],c):void 0,e&&(i&&i.exports!==p&&i.exports!==g[e]?g[e]=i.exports:u===p&&l||(g[e]=u))}else e&&(g[e]=n)},e=n=o=function(e,t,n,s,i){if("string"==typeof e)return f[e]?f[e](t):x(h(e,A(t)).f);if(!e.splice){if((y=e).deps&&o(y.deps,y.callback),!t)return;t.splice?(e=t,t=n,n=null):e=p}return t=t||function(){},"function"==typeof n&&(n=s,s=i),s?r(p,e,t,n):setTimeout(function(){r(p,e,t,n)},4),o},o.config=function(e){return o(e)},e._defined=g,(s=function(e,t,n){if("string"!=typeof e)throw new Error("See almond README: incorrect module build, no module name");t.splice||(n=t,t=[]),b(g,e)||b(m,e)||(m[e]=[e,t,n])}).amd={jQuery:!0},u.requirejs=e,u.require=n,u.define=s),u.define("almond",function(){}),u.define("jquery",[],function(){var e=t||$;return null==e&&console&&console.error&&console.error("Select2: An instance of jQuery or a jQuery-compatible library was not found. Make sure that you are including jQuery before Select2 on your web page."),e}),u.define("select2/utils",["jquery"],function(r){var s={};function c(e){var t,n=e.prototype,s=[];for(t in n)"function"==typeof n[t]&&"constructor"!==t&&s.push(t);return s}s.Extend=function(e,t){var n,s={}.hasOwnProperty;function i(){this.constructor=e}for(n in t)s.call(t,n)&&(e[n]=t[n]);return i.prototype=t.prototype,e.prototype=new i,e.__super__=t.prototype,e},s.Decorate=function(s,i){var e=c(i),t=c(s);function r(){var e=Array.prototype.unshift,t=i.prototype.constructor.length,n=s.prototype.constructor;0<t&&(e.call(arguments,s.prototype.constructor),n=i.prototype.constructor),n.apply(this,arguments)}i.displayName=s.displayName,r.prototype=new function(){this.constructor=r};for(var n=0;n<t.length;n++){var o=t[n];r.prototype[o]=s.prototype[o]}for(var a=0;a<e.length;a++){var l=e[a];r.prototype[l]=function(e){var t=function(){};e in r.prototype&&(t=r.prototype[e]);var n=i.prototype[e];return function(){return Array.prototype.unshift.call(arguments,t),n.apply(this,arguments)}}(l)}return r};function e(){this.listeners={}}e.prototype.on=function(e,t){this.listeners=this.listeners||{},e in this.listeners?this.listeners[e].push(t):this.listeners[e]=[t]},e.prototype.trigger=function(e){var t=Array.prototype.slice,n=t.call(arguments,1);this.listeners=this.listeners||{},0===(n=null==n?[]:n).length&&n.push({}),(n[0]._type=e)in this.listeners&&this.invoke(this.listeners[e],t.call(arguments,1)),"*"in this.listeners&&this.invoke(this.listeners["*"],arguments)},e.prototype.invoke=function(e,t){for(var n=0,s=e.length;n<s;n++)e[n].apply(this,t)},s.Observable=e,s.generateChars=function(e){for(var t="",n=0;n<e;n++)t+=Math.floor(36*Math.random()).toString(36);return t},s.bind=function(e,t){return function(){e.apply(t,arguments)}},s._convertData=function(e){for(var t in e){var n=t.split("-"),s=e;if(1!==n.length){for(var i=0;i<n.length;i++){var r=n[i];(r=r.substring(0,1).toLowerCase()+r.substring(1))in s||(s[r]={}),i==n.length-1&&(s[r]=e[t]),s=s[r]}delete e[t]}}return e},s.hasScroll=function(e,t){var n=r(t),s=t.style.overflowX,i=t.style.overflowY;return(s!==i||"hidden"!==i&&"visible"!==i)&&("scroll"===s||"scroll"===i||(n.innerHeight()<t.scrollHeight||n.innerWidth()<t.scrollWidth))},s.escapeMarkup=function(e){var t={"\\":"&#92;","&":"&amp;","<":"&lt;",">":"&gt;",'"':"&quot;","'":"&#39;","/":"&#47;"};return"string"!=typeof e?e:String(e).replace(/[&<>"'\/\\]/g,function(e){return t[e]})},s.__cache={};var n=0;return s.GetUniqueElementId=function(e){var t=e.getAttribute("data-select2-id");return null!=t||(t=e.id?"select2-data-"+e.id:"select2-data-"+(++n).toString()+"-"+s.generateChars(4),e.setAttribute("data-select2-id",t)),t},s.StoreData=function(e,t,n){e=s.GetUniqueElementId(e);s.__cache[e]||(s.__cache[e]={}),s.__cache[e][t]=n},s.GetData=function(e,t){var n=s.GetUniqueElementId(e);return t?s.__cache[n]&&null!=s.__cache[n][t]?s.__cache[n][t]:r(e).data(t):s.__cache[n]},s.RemoveData=function(e){var t=s.GetUniqueElementId(e);null!=s.__cache[t]&&delete s.__cache[t],e.removeAttribute("data-select2-id")},s.copyNonInternalCssClasses=function(e,t){var n=(n=e.getAttribute("class").trim().split(/\s+/)).filter(function(e){return 0===e.indexOf("select2-")}),t=(t=t.getAttribute("class").trim().split(/\s+/)).filter(function(e){return 0!==e.indexOf("select2-")}),t=n.concat(t);e.setAttribute("class",t.join(" "))},s}),u.define("select2/results",["jquery","./utils"],function(d,p){function s(e,t,n){this.$element=e,this.data=n,this.options=t,s.__super__.constructor.call(this)}return p.Extend(s,p.Observable),s.prototype.render=function(){var e=d('<ul class="select2-results__options" role="listbox"></ul>');return this.options.get("multiple")&&e.attr("aria-multiselectable","true"),this.$results=e},s.prototype.clear=function(){this.$results.empty()},s.prototype.displayMessage=function(e){var t=this.options.get("escapeMarkup");this.clear(),this.hideLoading();var n=d('<li role="alert" aria-live="assertive" class="select2-results__option"></li>'),s=this.options.get("translations").get(e.message);n.append(t(s(e.args))),n[0].className+=" select2-results__message",this.$results.append(n)},s.prototype.hideMessages=function(){this.$results.find(".select2-results__message").remove()},s.prototype.append=function(e){this.hideLoading();var t=[];if(null!=e.results&&0!==e.results.length){e.results=this.sort(e.results);for(var n=0;n<e.results.length;n++){var s=e.results[n],s=this.option(s);t.push(s)}this.$results.append(t)}else 0===this.$results.children().length&&this.trigger("results:message",{message:"noResults"})},s.prototype.position=function(e,t){t.find(".select2-results").append(e)},s.prototype.sort=function(e){return this.options.get("sorter")(e)},s.prototype.highlightFirstItem=function(){var e=this.$results.find(".select2-results__option--selectable"),t=e.filter(".select2-results__option--selected");(0<t.length?t:e).first().trigger("mouseenter"),this.ensureHighlightVisible()},s.prototype.setClasses=function(){var t=this;this.data.current(function(e){var s=e.map(function(e){return e.id.toString()});t.$results.find(".select2-results__option--selectable").each(function(){var e=d(this),t=p.GetData(this,"data"),n=""+t.id;null!=t.element&&t.element.selected||null==t.element&&-1<s.indexOf(n)?(this.classList.add("select2-results__option--selected"),e.attr("aria-selected","true")):(this.classList.remove("select2-results__option--selected"),e.attr("aria-selected","false"))})})},s.prototype.showLoading=function(e){this.hideLoading();e={disabled:!0,loading:!0,text:this.options.get("translations").get("searching")(e)},e=this.option(e);e.className+=" loading-results",this.$results.prepend(e)},s.prototype.hideLoading=function(){this.$results.find(".loading-results").remove()},s.prototype.option=function(e){var t=document.createElement("li");t.classList.add("select2-results__option"),t.classList.add("select2-results__option--selectable");var n,s={role:"option"},i=window.Element.prototype.matches||window.Element.prototype.msMatchesSelector||window.Element.prototype.webkitMatchesSelector;for(n in(null!=e.element&&i.call(e.element,":disabled")||null==e.element&&e.disabled)&&(s["aria-disabled"]="true",t.classList.remove("select2-results__option--selectable"),t.classList.add("select2-results__option--disabled")),null==e.id&&t.classList.remove("select2-results__option--selectable"),null!=e._resultId&&(t.id=e._resultId),e.title&&(t.title=e.title),e.children&&(s.role="group",s["aria-label"]=e.text,t.classList.remove("select2-results__option--selectable"),t.classList.add("select2-results__option--group")),s){var r=s[n];t.setAttribute(n,r)}if(e.children){var o=d(t),a=document.createElement("strong");a.className="select2-results__group",this.template(e,a);for(var l=[],c=0;c<e.children.length;c++){var u=e.children[c],u=this.option(u);l.push(u)}i=d("<ul></ul>",{class:"select2-results__options select2-results__options--nested",role:"none"});i.append(l),o.append(a),o.append(i)}else this.template(e,t);return p.StoreData(t,"data",e),t},s.prototype.bind=function(t,e){var i=this,n=t.id+"-results";this.$results.attr("id",n),t.on("results:all",function(e){i.clear(),i.append(e.data),t.isOpen()&&(i.setClasses(),i.highlightFirstItem())}),t.on("results:append",function(e){i.append(e.data),t.isOpen()&&i.setClasses()}),t.on("query",function(e){i.hideMessages(),i.showLoading(e)}),t.on("select",function(){t.isOpen()&&(i.setClasses(),i.options.get("scrollAfterSelect")&&i.highlightFirstItem())}),t.on("unselect",function(){t.isOpen()&&(i.setClasses(),i.options.get("scrollAfterSelect")&&i.highlightFirstItem())}),t.on("open",function(){i.$results.attr("aria-expanded","true"),i.$results.attr("aria-hidden","false"),i.setClasses(),i.ensureHighlightVisible()}),t.on("close",function(){i.$results.attr("aria-expanded","false"),i.$results.attr("aria-hidden","true"),i.$results.removeAttr("aria-activedescendant")}),t.on("results:toggle",function(){var e=i.getHighlightedResults();0!==e.length&&e.trigger("mouseup")}),t.on("results:select",function(){var e,t=i.getHighlightedResults();0!==t.length&&(e=p.GetData(t[0],"data"),t.hasClass("select2-results__option--selected")?i.trigger("close",{}):i.trigger("select",{data:e}))}),t.on("results:previous",function(){var e,t=i.getHighlightedResults(),n=i.$results.find(".select2-results__option--selectable"),s=n.index(t);s<=0||(e=s-1,0===t.length&&(e=0),(s=n.eq(e)).trigger("mouseenter"),t=i.$results.offset().top,n=s.offset().top,s=i.$results.scrollTop()+(n-t),0===e?i.$results.scrollTop(0):n-t<0&&i.$results.scrollTop(s))}),t.on("results:next",function(){var e,t=i.getHighlightedResults(),n=i.$results.find(".select2-results__option--selectable"),s=n.index(t)+1;s>=n.length||((e=n.eq(s)).trigger("mouseenter"),t=i.$results.offset().top+i.$results.outerHeight(!1),n=e.offset().top+e.outerHeight(!1),e=i.$results.scrollTop()+n-t,0===s?i.$results.scrollTop(0):t<n&&i.$results.scrollTop(e))}),t.on("results:focus",function(e){e.element[0].classList.add("select2-results__option--highlighted"),e.element[0].setAttribute("aria-selected","true")}),t.on("results:message",function(e){i.displayMessage(e)}),d.fn.mousewheel&&this.$results.on("mousewheel",function(e){var t=i.$results.scrollTop(),n=i.$results.get(0).scrollHeight-t+e.deltaY,t=0<e.deltaY&&t-e.deltaY<=0,n=e.deltaY<0&&n<=i.$results.height();t?(i.$results.scrollTop(0),e.preventDefault(),e.stopPropagation()):n&&(i.$results.scrollTop(i.$results.get(0).scrollHeight-i.$results.height()),e.preventDefault(),e.stopPropagation())}),this.$results.on("mouseup",".select2-results__option--selectable",function(e){var t=d(this),n=p.GetData(this,"data");t.hasClass("select2-results__option--selected")?i.options.get("multiple")?i.trigger("unselect",{originalEvent:e,data:n}):i.trigger("close",{}):i.trigger("select",{originalEvent:e,data:n})}),this.$results.on("mouseenter",".select2-results__option--selectable",function(e){var t=p.GetData(this,"data");i.getHighlightedResults().removeClass("select2-results__option--highlighted").attr("aria-selected","false"),i.trigger("results:focus",{data:t,element:d(this)})})},s.prototype.getHighlightedResults=function(){return this.$results.find(".select2-results__option--highlighted")},s.prototype.destroy=function(){this.$results.remove()},s.prototype.ensureHighlightVisible=function(){var e,t,n,s,i=this.getHighlightedResults();0!==i.length&&(e=this.$results.find(".select2-results__option--selectable").index(i),s=this.$results.offset().top,t=i.offset().top,n=this.$results.scrollTop()+(t-s),s=t-s,n-=2*i.outerHeight(!1),e<=2?this.$results.scrollTop(0):(s>this.$results.outerHeight()||s<0)&&this.$results.scrollTop(n))},s.prototype.template=function(e,t){var n=this.options.get("templateResult"),s=this.options.get("escapeMarkup"),e=n(e,t);null==e?t.style.display="none":"string"==typeof e?t.innerHTML=s(e):d(t).append(e)},s}),u.define("select2/keys",[],function(){return{BACKSPACE:8,TAB:9,ENTER:13,SHIFT:16,CTRL:17,ALT:18,ESC:27,SPACE:32,PAGE_UP:33,PAGE_DOWN:34,END:35,HOME:36,LEFT:37,UP:38,RIGHT:39,DOWN:40,DELETE:46}}),u.define("select2/selection/base",["jquery","../utils","../keys"],function(n,s,i){function r(e,t){this.$element=e,this.options=t,r.__super__.constructor.call(this)}return s.Extend(r,s.Observable),r.prototype.render=function(){var e=n('<span class="select2-selection" role="combobox"  aria-haspopup="true" aria-expanded="false"></span>');return this._tabindex=0,null!=s.GetData(this.$element[0],"old-tabindex")?this._tabindex=s.GetData(this.$element[0],"old-tabindex"):null!=this.$element.attr("tabindex")&&(this._tabindex=this.$element.attr("tabindex")),e.attr("title",this.$element.attr("title")),e.attr("tabindex",this._tabindex),e.attr("aria-disabled","false"),this.$selection=e},r.prototype.bind=function(e,t){var n=this,s=e.id+"-results";this.container=e,this.$selection.on("focus",function(e){n.trigger("focus",e)}),this.$selection.on("blur",function(e){n._handleBlur(e)}),this.$selection.on("keydown",function(e){n.trigger("keypress",e),e.which===i.SPACE&&e.preventDefault()}),e.on("results:focus",function(e){n.$selection.attr("aria-activedescendant",e.data._resultId)}),e.on("selection:update",function(e){n.update(e.data)}),e.on("open",function(){n.$selection.attr("aria-expanded","true"),n.$selection.attr("aria-owns",s),n._attachCloseHandler(e)}),e.on("close",function(){n.$selection.attr("aria-expanded","false"),n.$selection.removeAttr("aria-activedescendant"),n.$selection.removeAttr("aria-owns"),n.$selection.trigger("focus"),n._detachCloseHandler(e)}),e.on("enable",function(){n.$selection.attr("tabindex",n._tabindex),n.$selection.attr("aria-disabled","false")}),e.on("disable",function(){n.$selection.attr("tabindex","-1"),n.$selection.attr("aria-disabled","true")})},r.prototype._handleBlur=function(e){var t=this;window.setTimeout(function(){document.activeElement==t.$selection[0]||n.contains(t.$selection[0],document.activeElement)||t.trigger("blur",e)},1)},r.prototype._attachCloseHandler=function(e){n(document.body).on("mousedown.select2."+e.id,function(e){var t=n(e.target).closest(".select2");n(".select2.select2-container--open").each(function(){this!=t[0]&&s.GetData(this,"element").select2("close")})})},r.prototype._detachCloseHandler=function(e){n(document.body).off("mousedown.select2."+e.id)},r.prototype.position=function(e,t){t.find(".selection").append(e)},r.prototype.destroy=function(){this._detachCloseHandler(this.container)},r.prototype.update=function(e){throw new Error("The `update` method must be defined in child classes.")},r.prototype.isEnabled=function(){return!this.isDisabled()},r.prototype.isDisabled=function(){return this.options.get("disabled")},r}),u.define("select2/selection/single",["jquery","./base","../utils","../keys"],function(e,t,n,s){function i(){i.__super__.constructor.apply(this,arguments)}return n.Extend(i,t),i.prototype.render=function(){var e=i.__super__.render.call(this);return e[0].classList.add("select2-selection--single"),e.html('<span class="select2-selection__rendered"></span><span class="select2-selection__arrow" role="presentation"><b role="presentation"></b></span>'),e},i.prototype.bind=function(t,e){var n=this;i.__super__.bind.apply(this,arguments);var s=t.id+"-container";this.$selection.find(".select2-selection__rendered").attr("id",s).attr("role","textbox").attr("aria-readonly","true"),this.$selection.attr("aria-labelledby",s),this.$selection.attr("aria-controls",s),this.$selection.on("mousedown",function(e){1===e.which&&n.trigger("toggle",{originalEvent:e})}),this.$selection.on("focus",function(e){}),this.$selection.on("blur",function(e){}),t.on("focus",function(e){t.isOpen()||n.$selection.trigger("focus")})},i.prototype.clear=function(){var e=this.$selection.find(".select2-selection__rendered");e.empty(),e.removeAttr("title")},i.prototype.display=function(e,t){var n=this.options.get("templateSelection");return this.options.get("escapeMarkup")(n(e,t))},i.prototype.selectionContainer=function(){return e("<span></span>")},i.prototype.update=function(e){var t,n;0!==e.length?(n=e[0],t=this.$selection.find(".select2-selection__rendered"),e=this.display(n,t),t.empty().append(e),(n=n.title||n.text)?t.attr("title",n):t.removeAttr("title")):this.clear()},i}),u.define("select2/selection/multiple",["jquery","./base","../utils"],function(i,e,c){function r(e,t){r.__super__.constructor.apply(this,arguments)}return c.Extend(r,e),r.prototype.render=function(){var e=r.__super__.render.call(this);return e[0].classList.add("select2-selection--multiple"),e.html('<ul class="select2-selection__rendered"></ul>'),e},r.prototype.bind=function(e,t){var n=this;r.__super__.bind.apply(this,arguments);var s=e.id+"-container";this.$selection.find(".select2-selection__rendered").attr("id",s),this.$selection.on("click",function(e){n.trigger("toggle",{originalEvent:e})}),this.$selection.on("click",".select2-selection__choice__remove",function(e){var t;n.isDisabled()||(t=i(this).parent(),t=c.GetData(t[0],"data"),n.trigger("unselect",{originalEvent:e,data:t}))}),this.$selection.on("keydown",".select2-selection__choice__remove",function(e){n.isDisabled()||e.stopPropagation()})},r.prototype.clear=function(){var e=this.$selection.find(".select2-selection__rendered");e.empty(),e.removeAttr("title")},r.prototype.display=function(e,t){var n=this.options.get("templateSelection");return this.options.get("escapeMarkup")(n(e,t))},r.prototype.selectionContainer=function(){return i('<li class="select2-selection__choice"><button type="button" class="select2-selection__choice__remove" tabindex="-1"><span aria-hidden="true">&times;</span></button><span class="select2-selection__choice__display"></span></li>')},r.prototype.update=function(e){if(this.clear(),0!==e.length){for(var t=[],n=this.$selection.find(".select2-selection__rendered").attr("id")+"-choice-",s=0;s<e.length;s++){var i=e[s],r=this.selectionContainer(),o=this.display(i,r),a=n+c.generateChars(4)+"-";i.id?a+=i.id:a+=c.generateChars(4),r.find(".select2-selection__choice__display").append(o).attr("id",a);var l=i.title||i.text;l&&r.attr("title",l);o=this.options.get("translations").get("removeItem"),l=r.find(".select2-selection__choice__remove");l.attr("title",o()),l.attr("aria-label",o()),l.attr("aria-describedby",a),c.StoreData(r[0],"data",i),t.push(r)}this.$selection.find(".select2-selection__rendered").append(t)}},r}),u.define("select2/selection/placeholder",[],function(){function e(e,t,n){this.placeholder=this.normalizePlaceholder(n.get("placeholder")),e.call(this,t,n)}return e.prototype.normalizePlaceholder=function(e,t){return t="string"==typeof t?{id:"",text:t}:t},e.prototype.createPlaceholder=function(e,t){var n=this.selectionContainer();n.html(this.display(t)),n[0].classList.add("select2-selection__placeholder"),n[0].classList.remove("select2-selection__choice");t=t.title||t.text||n.text();return this.$selection.find(".select2-selection__rendered").attr("title",t),n},e.prototype.update=function(e,t){var n=1==t.length&&t[0].id!=this.placeholder.id;if(1<t.length||n)return e.call(this,t);this.clear();t=this.createPlaceholder(this.placeholder);this.$selection.find(".select2-selection__rendered").append(t)},e}),u.define("select2/selection/allowClear",["jquery","../keys","../utils"],function(i,s,a){function e(){}return e.prototype.bind=function(e,t,n){var s=this;e.call(this,t,n),null==this.placeholder&&this.options.get("debug")&&window.console&&console.error&&console.error("Select2: The `allowClear` option should be used in combination with the `placeholder` option."),this.$selection.on("mousedown",".select2-selection__clear",function(e){s._handleClear(e)}),t.on("keypress",function(e){s._handleKeyboardClear(e,t)})},e.prototype._handleClear=function(e,t){if(!this.isDisabled()){var n=this.$selection.find(".select2-selection__clear");if(0!==n.length){t.stopPropagation();var s=a.GetData(n[0],"data"),i=this.$element.val();this.$element.val(this.placeholder.id);var r={data:s};if(this.trigger("clear",r),r.prevented)this.$element.val(i);else{for(var o=0;o<s.length;o++)if(r={data:s[o]},this.trigger("unselect",r),r.prevented)return void this.$element.val(i);this.$element.trigger("input").trigger("change"),this.trigger("toggle",{})}}}},e.prototype._handleKeyboardClear=function(e,t,n){n.isOpen()||t.which!=s.DELETE&&t.which!=s.BACKSPACE||this._handleClear(t)},e.prototype.update=function(e,t){var n,s;e.call(this,t),this.$selection.find(".select2-selection__clear").remove(),this.$selection[0].classList.remove("select2-selection--clearable"),0<this.$selection.find(".select2-selection__placeholder").length||0===t.length||(n=this.$selection.find(".select2-selection__rendered").attr("id"),s=this.options.get("translations").get("removeAllItems"),(e=i('<button type="button" class="select2-selection__clear" tabindex="-1"><span aria-hidden="true">&times;</span></button>')).attr("title",s()),e.attr("aria-label",s()),e.attr("aria-describedby",n),a.StoreData(e[0],"data",t),this.$selection.prepend(e),this.$selection[0].classList.add("select2-selection--clearable"))},e}),u.define("select2/selection/search",["jquery","../utils","../keys"],function(s,a,l){function e(e,t,n){e.call(this,t,n)}return e.prototype.render=function(e){var t=this.options.get("translations").get("search"),n=s('<span class="select2-search select2-search--inline"><textarea class="select2-search__field" type="search" tabindex="-1" autocorrect="off" autocapitalize="none" spellcheck="false" role="searchbox" aria-autocomplete="list" ></textarea></span>');this.$searchContainer=n,this.$search=n.find("textarea"),this.$search.prop("autocomplete",this.options.get("autocomplete")),this.$search.attr("aria-label",t());e=e.call(this);return this._transferTabIndex(),e.append(this.$searchContainer),e},e.prototype.bind=function(e,t,n){var s=this,i=t.id+"-results",r=t.id+"-container";e.call(this,t,n),s.$search.attr("aria-describedby",r),t.on("open",function(){s.$search.attr("aria-controls",i),s.$search.trigger("focus")}),t.on("close",function(){s.$search.val(""),s.resizeSearch(),s.$search.removeAttr("aria-controls"),s.$search.removeAttr("aria-activedescendant"),s.$search.trigger("focus")}),t.on("enable",function(){s.$search.prop("disabled",!1),s._transferTabIndex()}),t.on("disable",function(){s.$search.prop("disabled",!0)}),t.on("focus",function(e){s.$search.trigger("focus")}),t.on("results:focus",function(e){e.data._resultId?s.$search.attr("aria-activedescendant",e.data._resultId):s.$search.removeAttr("aria-activedescendant")}),this.$selection.on("focusin",".select2-search--inline",function(e){s.trigger("focus",e)}),this.$selection.on("focusout",".select2-search--inline",function(e){s._handleBlur(e)}),this.$selection.on("keydown",".select2-search--inline",function(e){var t;e.stopPropagation(),s.trigger("keypress",e),s._keyUpPrevented=e.isDefaultPrevented(),e.which!==l.BACKSPACE||""!==s.$search.val()||0<(t=s.$selection.find(".select2-selection__choice").last()).length&&(t=a.GetData(t[0],"data"),s.searchRemoveChoice(t),e.preventDefault())}),this.$selection.on("click",".select2-search--inline",function(e){s.$search.val()&&e.stopPropagation()});var t=document.documentMode,o=t&&t<=11;this.$selection.on("input.searchcheck",".select2-search--inline",function(e){o?s.$selection.off("input.search input.searchcheck"):s.$selection.off("keyup.search")}),this.$selection.on("keyup.search input.search",".select2-search--inline",function(e){var t;o&&"input"===e.type?s.$selection.off("input.search input.searchcheck"):(t=e.which)!=l.SHIFT&&t!=l.CTRL&&t!=l.ALT&&t!=l.TAB&&s.handleSearch(e)})},e.prototype._transferTabIndex=function(e){this.$search.attr("tabindex",this.$selection.attr("tabindex")),this.$selection.attr("tabindex","-1")},e.prototype.createPlaceholder=function(e,t){this.$search.attr("placeholder",t.text)},e.prototype.update=function(e,t){var n=this.$search[0]==document.activeElement;this.$search.attr("placeholder",""),e.call(this,t),this.resizeSearch(),n&&this.$search.trigger("focus")},e.prototype.handleSearch=function(){var e;this.resizeSearch(),this._keyUpPrevented||(e=this.$search.val(),this.trigger("query",{term:e})),this._keyUpPrevented=!1},e.prototype.searchRemoveChoice=function(e,t){this.trigger("unselect",{data:t}),this.$search.val(t.text),this.handleSearch()},e.prototype.resizeSearch=function(){this.$search.css("width","25px");var e="100%";""===this.$search.attr("placeholder")&&(e=.75*(this.$search.val().length+1)+"em"),this.$search.css("width",e)},e}),u.define("select2/selection/selectionCss",["../utils"],function(n){function e(){}return e.prototype.render=function(e){var t=e.call(this),e=this.options.get("selectionCssClass")||"";return-1!==e.indexOf(":all:")&&(e=e.replace(":all:",""),n.copyNonInternalCssClasses(t[0],this.$element[0])),t.addClass(e),t},e}),u.define("select2/selection/eventRelay",["jquery"],function(o){function e(){}return e.prototype.bind=function(e,t,n){var s=this,i=["open","opening","close","closing","select","selecting","unselect","unselecting","clear","clearing"],r=["opening","closing","selecting","unselecting","clearing"];e.call(this,t,n),t.on("*",function(e,t){var n;-1!==i.indexOf(e)&&(t=t||{},n=o.Event("select2:"+e,{params:t}),s.$element.trigger(n),-1!==r.indexOf(e)&&(t.prevented=n.isDefaultPrevented()))})},e}),u.define("select2/translation",["jquery","require"],function(t,n){function s(e){this.dict=e||{}}return s.prototype.all=function(){return this.dict},s.prototype.get=function(e){return this.dict[e]},s.prototype.extend=function(e){this.dict=t.extend({},e.all(),this.dict)},s._cache={},s.loadPath=function(e){var t;return e in s._cache||(t=n(e),s._cache[e]=t),new s(s._cache[e])},s}),u.define("select2/diacritics",[],function(){return{"Ⓐ":"A","Ａ":"A","À":"A","Á":"A","Â":"A","Ầ":"A","Ấ":"A","Ẫ":"A","Ẩ":"A","Ã":"A","Ā":"A","Ă":"A","Ằ":"A","Ắ":"A","Ẵ":"A","Ẳ":"A","Ȧ":"A","Ǡ":"A","Ä":"A","Ǟ":"A","Ả":"A","Å":"A","Ǻ":"A","Ǎ":"A","Ȁ":"A","Ȃ":"A","Ạ":"A","Ậ":"A","Ặ":"A","Ḁ":"A","Ą":"A","Ⱥ":"A","Ɐ":"A","Ꜳ":"AA","Æ":"AE","Ǽ":"AE","Ǣ":"AE","Ꜵ":"AO","Ꜷ":"AU","Ꜹ":"AV","Ꜻ":"AV","Ꜽ":"AY","Ⓑ":"B","Ｂ":"B","Ḃ":"B","Ḅ":"B","Ḇ":"B","Ƀ":"B","Ƃ":"B","Ɓ":"B","Ⓒ":"C","Ｃ":"C","Ć":"C","Ĉ":"C","Ċ":"C","Č":"C","Ç":"C","Ḉ":"C","Ƈ":"C","Ȼ":"C","Ꜿ":"C","Ⓓ":"D","Ｄ":"D","Ḋ":"D","Ď":"D","Ḍ":"D","Ḑ":"D","Ḓ":"D","Ḏ":"D","Đ":"D","Ƌ":"D","Ɗ":"D","Ɖ":"D","Ꝺ":"D","Ǳ":"DZ","Ǆ":"DZ","ǲ":"Dz","ǅ":"Dz","Ⓔ":"E","Ｅ":"E","È":"E","É":"E","Ê":"E","Ề":"E","Ế":"E","Ễ":"E","Ể":"E","Ẽ":"E","Ē":"E","Ḕ":"E","Ḗ":"E","Ĕ":"E","Ė":"E","Ë":"E","Ẻ":"E","Ě":"E","Ȅ":"E","Ȇ":"E","Ẹ":"E","Ệ":"E","Ȩ":"E","Ḝ":"E","Ę":"E","Ḙ":"E","Ḛ":"E","Ɛ":"E","Ǝ":"E","Ⓕ":"F","Ｆ":"F","Ḟ":"F","Ƒ":"F","Ꝼ":"F","Ⓖ":"G","Ｇ":"G","Ǵ":"G","Ĝ":"G","Ḡ":"G","Ğ":"G","Ġ":"G","Ǧ":"G","Ģ":"G","Ǥ":"G","Ɠ":"G","Ꞡ":"G","Ᵹ":"G","Ꝿ":"G","Ⓗ":"H","Ｈ":"H","Ĥ":"H","Ḣ":"H","Ḧ":"H","Ȟ":"H","Ḥ":"H","Ḩ":"H","Ḫ":"H","Ħ":"H","Ⱨ":"H","Ⱶ":"H","Ɥ":"H","Ⓘ":"I","Ｉ":"I","Ì":"I","Í":"I","Î":"I","Ĩ":"I","Ī":"I","Ĭ":"I","İ":"I","Ï":"I","Ḯ":"I","Ỉ":"I","Ǐ":"I","Ȉ":"I","Ȋ":"I","Ị":"I","Į":"I","Ḭ":"I","Ɨ":"I","Ⓙ":"J","Ｊ":"J","Ĵ":"J","Ɉ":"J","Ⓚ":"K","Ｋ":"K","Ḱ":"K","Ǩ":"K","Ḳ":"K","Ķ":"K","Ḵ":"K","Ƙ":"K","Ⱪ":"K","Ꝁ":"K","Ꝃ":"K","Ꝅ":"K","Ꞣ":"K","Ⓛ":"L","Ｌ":"L","Ŀ":"L","Ĺ":"L","Ľ":"L","Ḷ":"L","Ḹ":"L","Ļ":"L","Ḽ":"L","Ḻ":"L","Ł":"L","Ƚ":"L","Ɫ":"L","Ⱡ":"L","Ꝉ":"L","Ꝇ":"L","Ꞁ":"L","Ǉ":"LJ","ǈ":"Lj","Ⓜ":"M","Ｍ":"M","Ḿ":"M","Ṁ":"M","Ṃ":"M","Ɱ":"M","Ɯ":"M","Ⓝ":"N","Ｎ":"N","Ǹ":"N","Ń":"N","Ñ":"N","Ṅ":"N","Ň":"N","Ṇ":"N","Ņ":"N","Ṋ":"N","Ṉ":"N","Ƞ":"N","Ɲ":"N","Ꞑ":"N","Ꞥ":"N","Ǌ":"NJ","ǋ":"Nj","Ⓞ":"O","Ｏ":"O","Ò":"O","Ó":"O","Ô":"O","Ồ":"O","Ố":"O","Ỗ":"O","Ổ":"O","Õ":"O","Ṍ":"O","Ȭ":"O","Ṏ":"O","Ō":"O","Ṑ":"O","Ṓ":"O","Ŏ":"O","Ȯ":"O","Ȱ":"O","Ö":"O","Ȫ":"O","Ỏ":"O","Ő":"O","Ǒ":"O","Ȍ":"O","Ȏ":"O","Ơ":"O","Ờ":"O","Ớ":"O","Ỡ":"O","Ở":"O","Ợ":"O","Ọ":"O","Ộ":"O","Ǫ":"O","Ǭ":"O","Ø":"O","Ǿ":"O","Ɔ":"O","Ɵ":"O","Ꝋ":"O","Ꝍ":"O","Œ":"OE","Ƣ":"OI","Ꝏ":"OO","Ȣ":"OU","Ⓟ":"P","Ｐ":"P","Ṕ":"P","Ṗ":"P","Ƥ":"P","Ᵽ":"P","Ꝑ":"P","Ꝓ":"P","Ꝕ":"P","Ⓠ":"Q","Ｑ":"Q","Ꝗ":"Q","Ꝙ":"Q","Ɋ":"Q","Ⓡ":"R","Ｒ":"R","Ŕ":"R","Ṙ":"R","Ř":"R","Ȑ":"R","Ȓ":"R","Ṛ":"R","Ṝ":"R","Ŗ":"R","Ṟ":"R","Ɍ":"R","Ɽ":"R","Ꝛ":"R","Ꞧ":"R","Ꞃ":"R","Ⓢ":"S","Ｓ":"S","ẞ":"S","Ś":"S","Ṥ":"S","Ŝ":"S","Ṡ":"S","Š":"S","Ṧ":"S","Ṣ":"S","Ṩ":"S","Ș":"S","Ş":"S","Ȿ":"S","Ꞩ":"S","Ꞅ":"S","Ⓣ":"T","Ｔ":"T","Ṫ":"T","Ť":"T","Ṭ":"T","Ț":"T","Ţ":"T","Ṱ":"T","Ṯ":"T","Ŧ":"T","Ƭ":"T","Ʈ":"T","Ⱦ":"T","Ꞇ":"T","Ꜩ":"TZ","Ⓤ":"U","Ｕ":"U","Ù":"U","Ú":"U","Û":"U","Ũ":"U","Ṹ":"U","Ū":"U","Ṻ":"U","Ŭ":"U","Ü":"U","Ǜ":"U","Ǘ":"U","Ǖ":"U","Ǚ":"U","Ủ":"U","Ů":"U","Ű":"U","Ǔ":"U","Ȕ":"U","Ȗ":"U","Ư":"U","Ừ":"U","Ứ":"U","Ữ":"U","Ử":"U","Ự":"U","Ụ":"U","Ṳ":"U","Ų":"U","Ṷ":"U","Ṵ":"U","Ʉ":"U","Ⓥ":"V","Ｖ":"V","Ṽ":"V","Ṿ":"V","Ʋ":"V","Ꝟ":"V","Ʌ":"V","Ꝡ":"VY","Ⓦ":"W","Ｗ":"W","Ẁ":"W","Ẃ":"W","Ŵ":"W","Ẇ":"W","Ẅ":"W","Ẉ":"W","Ⱳ":"W","Ⓧ":"X","Ｘ":"X","Ẋ":"X","Ẍ":"X","Ⓨ":"Y","Ｙ":"Y","Ỳ":"Y","Ý":"Y","Ŷ":"Y","Ỹ":"Y","Ȳ":"Y","Ẏ":"Y","Ÿ":"Y","Ỷ":"Y","Ỵ":"Y","Ƴ":"Y","Ɏ":"Y","Ỿ":"Y","Ⓩ":"Z","Ｚ":"Z","Ź":"Z","Ẑ":"Z","Ż":"Z","Ž":"Z","Ẓ":"Z","Ẕ":"Z","Ƶ":"Z","Ȥ":"Z","Ɀ":"Z","Ⱬ":"Z","Ꝣ":"Z","ⓐ":"a","ａ":"a","ẚ":"a","à":"a","á":"a","â":"a","ầ":"a","ấ":"a","ẫ":"a","ẩ":"a","ã":"a","ā":"a","ă":"a","ằ":"a","ắ":"a","ẵ":"a","ẳ":"a","ȧ":"a","ǡ":"a","ä":"a","ǟ":"a","ả":"a","å":"a","ǻ":"a","ǎ":"a","ȁ":"a","ȃ":"a","ạ":"a","ậ":"a","ặ":"a","ḁ":"a","ą":"a","ⱥ":"a","ɐ":"a","ꜳ":"aa","æ":"ae","ǽ":"ae","ǣ":"ae","ꜵ":"ao","ꜷ":"au","ꜹ":"av","ꜻ":"av","ꜽ":"ay","ⓑ":"b","ｂ":"b","ḃ":"b","ḅ":"b","ḇ":"b","ƀ":"b","ƃ":"b","ɓ":"b","ⓒ":"c","ｃ":"c","ć":"c","ĉ":"c","ċ":"c","č":"c","ç":"c","ḉ":"c","ƈ":"c","ȼ":"c","ꜿ":"c","ↄ":"c","ⓓ":"d","ｄ":"d","ḋ":"d","ď":"d","ḍ":"d","ḑ":"d","ḓ":"d","ḏ":"d","đ":"d","ƌ":"d","ɖ":"d","ɗ":"d","ꝺ":"d","ǳ":"dz","ǆ":"dz","ⓔ":"e","ｅ":"e","è":"e","é":"e","ê":"e","ề":"e","ế":"e","ễ":"e","ể":"e","ẽ":"e","ē":"e","ḕ":"e","ḗ":"e","ĕ":"e","ė":"e","ë":"e","ẻ":"e","ě":"e","ȅ":"e","ȇ":"e","ẹ":"e","ệ":"e","ȩ":"e","ḝ":"e","ę":"e","ḙ":"e","ḛ":"e","ɇ":"e","ɛ":"e","ǝ":"e","ⓕ":"f","ｆ":"f","ḟ":"f","ƒ":"f","ꝼ":"f","ⓖ":"g","ｇ":"g","ǵ":"g","ĝ":"g","ḡ":"g","ğ":"g","ġ":"g","ǧ":"g","ģ":"g","ǥ":"g","ɠ":"g","ꞡ":"g","ᵹ":"g","ꝿ":"g","ⓗ":"h","ｈ":"h","ĥ":"h","ḣ":"h","ḧ":"h","ȟ":"h","ḥ":"h","ḩ":"h","ḫ":"h","ẖ":"h","ħ":"h","ⱨ":"h","ⱶ":"h","ɥ":"h","ƕ":"hv","ⓘ":"i","ｉ":"i","ì":"i","í":"i","î":"i","ĩ":"i","ī":"i","ĭ":"i","ï":"i","ḯ":"i","ỉ":"i","ǐ":"i","ȉ":"i","ȋ":"i","ị":"i","į":"i","ḭ":"i","ɨ":"i","ı":"i","ⓙ":"j","ｊ":"j","ĵ":"j","ǰ":"j","ɉ":"j","ⓚ":"k","ｋ":"k","ḱ":"k","ǩ":"k","ḳ":"k","ķ":"k","ḵ":"k","ƙ":"k","ⱪ":"k","ꝁ":"k","ꝃ":"k","ꝅ":"k","ꞣ":"k","ⓛ":"l","ｌ":"l","ŀ":"l","ĺ":"l","ľ":"l","ḷ":"l","ḹ":"l","ļ":"l","ḽ":"l","ḻ":"l","ſ":"l","ł":"l","ƚ":"l","ɫ":"l","ⱡ":"l","ꝉ":"l","ꞁ":"l","ꝇ":"l","ǉ":"lj","ⓜ":"m","ｍ":"m","ḿ":"m","ṁ":"m","ṃ":"m","ɱ":"m","ɯ":"m","ⓝ":"n","ｎ":"n","ǹ":"n","ń":"n","ñ":"n","ṅ":"n","ň":"n","ṇ":"n","ņ":"n","ṋ":"n","ṉ":"n","ƞ":"n","ɲ":"n","ŉ":"n","ꞑ":"n","ꞥ":"n","ǌ":"nj","ⓞ":"o","ｏ":"o","ò":"o","ó":"o","ô":"o","ồ":"o","ố":"o","ỗ":"o","ổ":"o","õ":"o","ṍ":"o","ȭ":"o","ṏ":"o","ō":"o","ṑ":"o","ṓ":"o","ŏ":"o","ȯ":"o","ȱ":"o","ö":"o","ȫ":"o","ỏ":"o","ő":"o","ǒ":"o","ȍ":"o","ȏ":"o","ơ":"o","ờ":"o","ớ":"o","ỡ":"o","ở":"o","ợ":"o","ọ":"o","ộ":"o","ǫ":"o","ǭ":"o","ø":"o","ǿ":"o","ɔ":"o","ꝋ":"o","ꝍ":"o","ɵ":"o","œ":"oe","ƣ":"oi","ȣ":"ou","ꝏ":"oo","ⓟ":"p","ｐ":"p","ṕ":"p","ṗ":"p","ƥ":"p","ᵽ":"p","ꝑ":"p","ꝓ":"p","ꝕ":"p","ⓠ":"q","ｑ":"q","ɋ":"q","ꝗ":"q","ꝙ":"q","ⓡ":"r","ｒ":"r","ŕ":"r","ṙ":"r","ř":"r","ȑ":"r","ȓ":"r","ṛ":"r","ṝ":"r","ŗ":"r","ṟ":"r","ɍ":"r","ɽ":"r","ꝛ":"r","ꞧ":"r","ꞃ":"r","ⓢ":"s","ｓ":"s","ß":"s","ś":"s","ṥ":"s","ŝ":"s","ṡ":"s","š":"s","ṧ":"s","ṣ":"s","ṩ":"s","ș":"s","ş":"s","ȿ":"s","ꞩ":"s","ꞅ":"s","ẛ":"s","ⓣ":"t","ｔ":"t","ṫ":"t","ẗ":"t","ť":"t","ṭ":"t","ț":"t","ţ":"t","ṱ":"t","ṯ":"t","ŧ":"t","ƭ":"t","ʈ":"t","ⱦ":"t","ꞇ":"t","ꜩ":"tz","ⓤ":"u","ｕ":"u","ù":"u","ú":"u","û":"u","ũ":"u","ṹ":"u","ū":"u","ṻ":"u","ŭ":"u","ü":"u","ǜ":"u","ǘ":"u","ǖ":"u","ǚ":"u","ủ":"u","ů":"u","ű":"u","ǔ":"u","ȕ":"u","ȗ":"u","ư":"u","ừ":"u","ứ":"u","ữ":"u","ử":"u","ự":"u","ụ":"u","ṳ":"u","ų":"u","ṷ":"u","ṵ":"u","ʉ":"u","ⓥ":"v","ｖ":"v","ṽ":"v","ṿ":"v","ʋ":"v","ꝟ":"v","ʌ":"v","ꝡ":"vy","ⓦ":"w","ｗ":"w","ẁ":"w","ẃ":"w","ŵ":"w","ẇ":"w","ẅ":"w","ẘ":"w","ẉ":"w","ⱳ":"w","ⓧ":"x","ｘ":"x","ẋ":"x","ẍ":"x","ⓨ":"y","ｙ":"y","ỳ":"y","ý":"y","ŷ":"y","ỹ":"y","ȳ":"y","ẏ":"y","ÿ":"y","ỷ":"y","ẙ":"y","ỵ":"y","ƴ":"y","ɏ":"y","ỿ":"y","ⓩ":"z","ｚ":"z","ź":"z","ẑ":"z","ż":"z","ž":"z","ẓ":"z","ẕ":"z","ƶ":"z","ȥ":"z","ɀ":"z","ⱬ":"z","ꝣ":"z","Ά":"Α","Έ":"Ε","Ή":"Η","Ί":"Ι","Ϊ":"Ι","Ό":"Ο","Ύ":"Υ","Ϋ":"Υ","Ώ":"Ω","ά":"α","έ":"ε","ή":"η","ί":"ι","ϊ":"ι","ΐ":"ι","ό":"ο","ύ":"υ","ϋ":"υ","ΰ":"υ","ώ":"ω","ς":"σ","’":"'"}}),u.define("select2/data/base",["../utils"],function(n){function s(e,t){s.__super__.constructor.call(this)}return n.Extend(s,n.Observable),s.prototype.current=function(e){throw new Error("The `current` method must be defined in child classes.")},s.prototype.query=function(e,t){throw new Error("The `query` method must be defined in child classes.")},s.prototype.bind=function(e,t){},s.prototype.destroy=function(){},s.prototype.generateResultId=function(e,t){e=e.id+"-result-";return e+=n.generateChars(4),null!=t.id?e+="-"+t.id.toString():e+="-"+n.generateChars(4),e},s}),u.define("select2/data/select",["./base","../utils","jquery"],function(e,a,l){function n(e,t){this.$element=e,this.options=t,n.__super__.constructor.call(this)}return a.Extend(n,e),n.prototype.current=function(e){var t=this;e(Array.prototype.map.call(this.$element[0].querySelectorAll(":checked"),function(e){return t.item(l(e))}))},n.prototype.select=function(i){var e,r=this;if(i.selected=!0,null!=i.element&&"option"===i.element.tagName.toLowerCase())return i.element.selected=!0,void this.$element.trigger("input").trigger("change");this.$element.prop("multiple")?this.current(function(e){var t=[];(i=[i]).push.apply(i,e);for(var n=0;n<i.length;n++){var s=i[n].id;-1===t.indexOf(s)&&t.push(s)}r.$element.val(t),r.$element.trigger("input").trigger("change")}):(e=i.id,this.$element.val(e),this.$element.trigger("input").trigger("change"))},n.prototype.unselect=function(i){var r=this;if(this.$element.prop("multiple")){if(i.selected=!1,null!=i.element&&"option"===i.element.tagName.toLowerCase())return i.element.selected=!1,void this.$element.trigger("input").trigger("change");this.current(function(e){for(var t=[],n=0;n<e.length;n++){var s=e[n].id;s!==i.id&&-1===t.indexOf(s)&&t.push(s)}r.$element.val(t),r.$element.trigger("input").trigger("change")})}},n.prototype.bind=function(e,t){var n=this;(this.container=e).on("select",function(e){n.select(e.data)}),e.on("unselect",function(e){n.unselect(e.data)})},n.prototype.destroy=function(){this.$element.find("*").each(function(){a.RemoveData(this)})},n.prototype.query=function(t,e){var n=[],s=this;this.$element.children().each(function(){var e;"option"!==this.tagName.toLowerCase()&&"optgroup"!==this.tagName.toLowerCase()||(e=l(this),e=s.item(e),null!==(e=s.matches(t,e))&&n.push(e))}),e({results:n})},n.prototype.addOptions=function(e){this.$element.append(e)},n.prototype.option=function(e){var t;e.children?(t=document.createElement("optgroup")).label=e.text:void 0!==(t=document.createElement("option")).textContent?t.textContent=e.text:t.innerText=e.text,void 0!==e.id&&(t.value=e.id),e.disabled&&(t.disabled=!0),e.selected&&(t.selected=!0),e.title&&(t.title=e.title);e=this._normalizeItem(e);return e.element=t,a.StoreData(t,"data",e),l(t)},n.prototype.item=function(e){var t={};if(null!=(t=a.GetData(e[0],"data")))return t;var n=e[0];if("option"===n.tagName.toLowerCase())t={id:e.val(),text:e.text(),disabled:e.prop("disabled"),selected:e.prop("selected"),title:e.prop("title")};else if("optgroup"===n.tagName.toLowerCase()){t={text:e.prop("label"),children:[],title:e.prop("title")};for(var s=e.children("option"),i=[],r=0;r<s.length;r++){var o=l(s[r]),o=this.item(o);i.push(o)}t.children=i}return(t=this._normalizeItem(t)).element=e[0],a.StoreData(e[0],"data",t),t},n.prototype._normalizeItem=function(e){e!==Object(e)&&(e={id:e,text:e});return null!=(e=l.extend({},{text:""},e)).id&&(e.id=e.id.toString()),null!=e.text&&(e.text=e.text.toString()),null==e._resultId&&e.id&&null!=this.container&&(e._resultId=this.generateResultId(this.container,e)),l.extend({},{selected:!1,disabled:!1},e)},n.prototype.matches=function(e,t){return this.options.get("matcher")(e,t)},n}),u.define("select2/data/array",["./select","../utils","jquery"],function(e,t,c){function s(e,t){this._dataToConvert=t.get("data")||[],s.__super__.constructor.call(this,e,t)}return t.Extend(s,e),s.prototype.bind=function(e,t){s.__super__.bind.call(this,e,t),this.addOptions(this.convertToOptions(this._dataToConvert))},s.prototype.select=function(n){var e=this.$element.find("option").filter(function(e,t){return t.value==n.id.toString()});0===e.length&&(e=this.option(n),this.addOptions(e)),s.__super__.select.call(this,n)},s.prototype.convertToOptions=function(e){var t=this,n=this.$element.find("option"),s=n.map(function(){return t.item(c(this)).id}).get(),i=[];for(var r=0;r<e.length;r++){var o,a,l=this._normalizeItem(e[r]);0<=s.indexOf(l.id)?(o=n.filter(function(e){return function(){return c(this).val()==e.id}}(l)),a=this.item(o),a=c.extend(!0,{},l,a),a=this.option(a),o.replaceWith(a)):(a=this.option(l),l.children&&(l=this.convertToOptions(l.children),a.append(l)),i.push(a))}return i},s}),u.define("select2/data/ajax",["./array","../utils","jquery"],function(e,t,r){function n(e,t){this.ajaxOptions=this._applyDefaults(t.get("ajax")),null!=this.ajaxOptions.processResults&&(this.processResults=this.ajaxOptions.processResults),n.__super__.constructor.call(this,e,t)}return t.Extend(n,e),n.prototype._applyDefaults=function(e){var t={data:function(e){return r.extend({},e,{q:e.term})},transport:function(e,t,n){e=r.ajax(e);return e.then(t),e.fail(n),e}};return r.extend({},t,e,!0)},n.prototype.processResults=function(e){return e},n.prototype.query=function(t,n){var s=this;null!=this._request&&("function"==typeof this._request.abort&&this._request.abort(),this._request=null);var i=r.extend({type:"GET"},this.ajaxOptions);function e(){var e=i.transport(i,function(e){e=s.processResults(e,t);s.options.get("debug")&&window.console&&console.error&&(e&&e.results&&Array.isArray(e.results)||console.error("Select2: The AJAX results did not return an array in the `results` key of the response.")),n(e)},function(){"status"in e&&(0===e.status||"0"===e.status)||s.trigger("results:message",{message:"errorLoading"})});s._request=e}"function"==typeof i.url&&(i.url=i.url.call(this.$element,t)),"function"==typeof i.data&&(i.data=i.data.call(this.$element,t)),this.ajaxOptions.delay&&null!=t.term?(this._queryTimeout&&window.clearTimeout(this._queryTimeout),this._queryTimeout=window.setTimeout(e,this.ajaxOptions.delay)):e()},n}),u.define("select2/data/tags",["jquery"],function(t){function e(e,t,n){var s=n.get("tags"),i=n.get("createTag");void 0!==i&&(this.createTag=i);i=n.get("insertTag");if(void 0!==i&&(this.insertTag=i),e.call(this,t,n),Array.isArray(s))for(var r=0;r<s.length;r++){var o=s[r],o=this._normalizeItem(o),o=this.option(o);this.$element.append(o)}}return e.prototype.query=function(e,c,u){var d=this;this._removeOldTags(),null!=c.term&&null==c.page?e.call(this,c,function e(t,n){for(var s=t.results,i=0;i<s.length;i++){var r=s[i],o=null!=r.children&&!e({results:r.children},!0);if((r.text||"").toUpperCase()===(c.term||"").toUpperCase()||o)return!n&&(t.data=s,void u(t))}if(n)return!0;var a,l=d.createTag(c);null!=l&&((a=d.option(l)).attr("data-select2-tag","true"),d.addOptions([a]),d.insertTag(s,l)),t.results=s,u(t)}):e.call(this,c,u)},e.prototype.createTag=function(e,t){if(null==t.term)return null;t=t.term.trim();return""===t?null:{id:t,text:t}},e.prototype.insertTag=function(e,t,n){t.unshift(n)},e.prototype._removeOldTags=function(e){this.$element.find("option[data-select2-tag]").each(function(){this.selected||t(this).remove()})},e}),u.define("select2/data/tokenizer",["jquery"],function(c){function e(e,t,n){var s=n.get("tokenizer");void 0!==s&&(this.tokenizer=s),e.call(this,t,n)}return e.prototype.bind=function(e,t,n){e.call(this,t,n),this.$search=t.dropdown.$search||t.selection.$search||n.find(".select2-search__field")},e.prototype.query=function(e,t,n){var s=this;t.term=t.term||"";var i=this.tokenizer(t,this.options,function(e){var t,n=s._normalizeItem(e);s.$element.find("option").filter(function(){return c(this).val()===n.id}).length||((t=s.option(n)).attr("data-select2-tag",!0),s._removeOldTags(),s.addOptions([t])),t=n,s.trigger("select",{data:t})});i.term!==t.term&&(this.$search.length&&(this.$search.val(i.term),this.$search.trigger("focus")),t.term=i.term),e.call(this,t,n)},e.prototype.tokenizer=function(e,t,n,s){for(var i=n.get("tokenSeparators")||[],r=t.term,o=0,a=this.createTag||function(e){return{id:e.term,text:e.term}};o<r.length;){var l=r[o];-1!==i.indexOf(l)?(l=r.substr(0,o),null!=(l=a(c.extend({},t,{term:l})))?(s(l),r=r.substr(o+1)||"",o=0):o++):o++}return{term:r}},e}),u.define("select2/data/minimumInputLength",[],function(){function e(e,t,n){this.minimumInputLength=n.get("minimumInputLength"),e.call(this,t,n)}return e.prototype.query=function(e,t,n){t.term=t.term||"",t.term.length<this.minimumInputLength?this.trigger("results:message",{message:"inputTooShort",args:{minimum:this.minimumInputLength,input:t.term,params:t}}):e.call(this,t,n)},e}),u.define("select2/data/maximumInputLength",[],function(){function e(e,t,n){this.maximumInputLength=n.get("maximumInputLength"),e.call(this,t,n)}return e.prototype.query=function(e,t,n){t.term=t.term||"",0<this.maximumInputLength&&t.term.length>this.maximumInputLength?this.trigger("results:message",{message:"inputTooLong",args:{maximum:this.maximumInputLength,input:t.term,params:t}}):e.call(this,t,n)},e}),u.define("select2/data/maximumSelectionLength",[],function(){function e(e,t,n){this.maximumSelectionLength=n.get("maximumSelectionLength"),e.call(this,t,n)}return e.prototype.bind=function(e,t,n){var s=this;e.call(this,t,n),t.on("select",function(){s._checkIfMaximumSelected()})},e.prototype.query=function(e,t,n){var s=this;this._checkIfMaximumSelected(function(){e.call(s,t,n)})},e.prototype._checkIfMaximumSelected=function(e,t){var n=this;this.current(function(e){e=null!=e?e.length:0;0<n.maximumSelectionLength&&e>=n.maximumSelectionLength?n.trigger("results:message",{message:"maximumSelected",args:{maximum:n.maximumSelectionLength}}):t&&t()})},e}),u.define("select2/dropdown",["jquery","./utils"],function(t,e){function n(e,t){this.$element=e,this.options=t,n.__super__.constructor.call(this)}return e.Extend(n,e.Observable),n.prototype.render=function(){var e=t('<span class="select2-dropdown"><span class="select2-results"></span></span>');return e.attr("dir",this.options.get("dir")),this.$dropdown=e},n.prototype.bind=function(){},n.prototype.position=function(e,t){},n.prototype.destroy=function(){this.$dropdown.remove()},n}),u.define("select2/dropdown/search",["jquery"],function(r){function e(){}return e.prototype.render=function(e){var t=e.call(this),n=this.options.get("translations").get("search"),e=r('<span class="select2-search select2-search--dropdown"><input class="select2-search__field" type="search" tabindex="-1" autocorrect="off" autocapitalize="none" spellcheck="false" role="searchbox" aria-autocomplete="list" /></span>');return this.$searchContainer=e,this.$search=e.find("input"),this.$search.prop("autocomplete",this.options.get("autocomplete")),this.$search.attr("aria-label",n()),t.prepend(e),t},e.prototype.bind=function(e,t,n){var s=this,i=t.id+"-results";e.call(this,t,n),this.$search.on("keydown",function(e){s.trigger("keypress",e),s._keyUpPrevented=e.isDefaultPrevented()}),this.$search.on("input",function(e){r(this).off("keyup")}),this.$search.on("keyup input",function(e){s.handleSearch(e)}),t.on("open",function(){s.$search.attr("tabindex",0),s.$search.attr("aria-controls",i),s.$search.trigger("focus"),window.setTimeout(function(){s.$search.trigger("focus")},0)}),t.on("close",function(){s.$search.attr("tabindex",-1),s.$search.removeAttr("aria-controls"),s.$search.removeAttr("aria-activedescendant"),s.$search.val(""),s.$search.trigger("blur")}),t.on("focus",function(){t.isOpen()||s.$search.trigger("focus")}),t.on("results:all",function(e){null!=e.query.term&&""!==e.query.term||(s.showSearch(e)?s.$searchContainer[0].classList.remove("select2-search--hide"):s.$searchContainer[0].classList.add("select2-search--hide"))}),t.on("results:focus",function(e){e.data._resultId?s.$search.attr("aria-activedescendant",e.data._resultId):s.$search.removeAttr("aria-activedescendant")})},e.prototype.handleSearch=function(e){var t;this._keyUpPrevented||(t=this.$search.val(),this.trigger("query",{term:t})),this._keyUpPrevented=!1},e.prototype.showSearch=function(e,t){return!0},e}),u.define("select2/dropdown/hidePlaceholder",[],function(){function e(e,t,n,s){this.placeholder=this.normalizePlaceholder(n.get("placeholder")),e.call(this,t,n,s)}return e.prototype.append=function(e,t){t.results=this.removePlaceholder(t.results),e.call(this,t)},e.prototype.normalizePlaceholder=function(e,t){return t="string"==typeof t?{id:"",text:t}:t},e.prototype.removePlaceholder=function(e,t){for(var n=t.slice(0),s=t.length-1;0<=s;s--){var i=t[s];this.placeholder.id===i.id&&n.splice(s,1)}return n},e}),u.define("select2/dropdown/infiniteScroll",["jquery"],function(n){function e(e,t,n,s){this.lastParams={},e.call(this,t,n,s),this.$loadingMore=this.createLoadingMore(),this.loading=!1}return e.prototype.append=function(e,t){this.$loadingMore.remove(),this.loading=!1,e.call(this,t),this.showLoadingMore(t)&&(this.$results.append(this.$loadingMore),this.loadMoreIfNeeded())},e.prototype.bind=function(e,t,n){var s=this;e.call(this,t,n),t.on("query",function(e){s.lastParams=e,s.loading=!0}),t.on("query:append",function(e){s.lastParams=e,s.loading=!0}),this.$results.on("scroll",this.loadMoreIfNeeded.bind(this))},e.prototype.loadMoreIfNeeded=function(){var e=n.contains(document.documentElement,this.$loadingMore[0]);!this.loading&&e&&(e=this.$results.offset().top+this.$results.outerHeight(!1),this.$loadingMore.offset().top+this.$loadingMore.outerHeight(!1)<=e+50&&this.loadMore())},e.prototype.loadMore=function(){this.loading=!0;var e=n.extend({},{page:1},this.lastParams);e.page++,this.trigger("query:append",e)},e.prototype.showLoadingMore=function(e,t){return t.pagination&&t.pagination.more},e.prototype.createLoadingMore=function(){var e=n('<li class="select2-results__option select2-results__option--load-more"role="option" aria-disabled="true"></li>'),t=this.options.get("translations").get("loadingMore");return e.html(t(this.lastParams)),e},e}),u.define("select2/dropdown/attachBody",["jquery","../utils"],function(u,o){function e(e,t,n){this.$dropdownParent=u(n.get("dropdownParent")||document.body),e.call(this,t,n)}return e.prototype.bind=function(e,t,n){var s=this;e.call(this,t,n),t.on("open",function(){s._showDropdown(),s._attachPositioningHandler(t),s._bindContainerResultHandlers(t)}),t.on("close",function(){s._hideDropdown(),s._detachPositioningHandler(t)}),this.$dropdownContainer.on("mousedown",function(e){e.stopPropagation()})},e.prototype.destroy=function(e){e.call(this),this.$dropdownContainer.remove()},e.prototype.position=function(e,t,n){t.attr("class",n.attr("class")),t[0].classList.remove("select2"),t[0].classList.add("select2-container--open"),t.css({position:"absolute",top:-999999}),this.$container=n},e.prototype.render=function(e){var t=u("<span></span>"),e=e.call(this);return t.append(e),this.$dropdownContainer=t},e.prototype._hideDropdown=function(e){this.$dropdownContainer.detach()},e.prototype._bindContainerResultHandlers=function(e,t){var n;this._containerResultsHandlersBound||(n=this,t.on("results:all",function(){n._positionDropdown(),n._resizeDropdown()}),t.on("results:append",function(){n._positionDropdown(),n._resizeDropdown()}),t.on("results:message",function(){n._positionDropdown(),n._resizeDropdown()}),t.on("select",function(){n._positionDropdown(),n._resizeDropdown()}),t.on("unselect",function(){n._positionDropdown(),n._resizeDropdown()}),this._containerResultsHandlersBound=!0)},e.prototype._attachPositioningHandler=function(e,t){var n=this,s="scroll.select2."+t.id,i="resize.select2."+t.id,r="orientationchange.select2."+t.id,t=this.$container.parents().filter(o.hasScroll);t.each(function(){o.StoreData(this,"select2-scroll-position",{x:u(this).scrollLeft(),y:u(this).scrollTop()})}),t.on(s,function(e){var t=o.GetData(this,"select2-scroll-position");u(this).scrollTop(t.y)}),u(window).on(s+" "+i+" "+r,function(e){n._positionDropdown(),n._resizeDropdown()})},e.prototype._detachPositioningHandler=function(e,t){var n="scroll.select2."+t.id,s="resize.select2."+t.id,t="orientationchange.select2."+t.id;this.$container.parents().filter(o.hasScroll).off(n),u(window).off(n+" "+s+" "+t)},e.prototype._positionDropdown=function(){var e=u(window),t=this.$dropdown[0].classList.contains("select2-dropdown--above"),n=this.$dropdown[0].classList.contains("select2-dropdown--below"),s=null,i=this.$container.offset();i.bottom=i.top+this.$container.outerHeight(!1);var r={height:this.$container.outerHeight(!1)};r.top=i.top,r.bottom=i.top+r.height;var o=this.$dropdown.outerHeight(!1),a=e.scrollTop(),l=e.scrollTop()+e.height(),c=a<i.top-o,e=l>i.bottom+o,a={left:i.left,top:r.bottom},l=this.$dropdownParent;"static"===l.css("position")&&(l=l.offsetParent());i={top:0,left:0};(u.contains(document.body,l[0])||l[0].isConnected)&&(i=l.offset()),a.top-=i.top,a.left-=i.left,t||n||(s="below"),e||!c||t?!c&&e&&t&&(s="below"):s="above",("above"==s||t&&"below"!==s)&&(a.top=r.top-i.top-o),null!=s&&(this.$dropdown[0].classList.remove("select2-dropdown--below"),this.$dropdown[0].classList.remove("select2-dropdown--above"),this.$dropdown[0].classList.add("select2-dropdown--"+s),this.$container[0].classList.remove("select2-container--below"),this.$container[0].classList.remove("select2-container--above"),this.$container[0].classList.add("select2-container--"+s)),this.$dropdownContainer.css(a)},e.prototype._resizeDropdown=function(){var e={width:this.$container.outerWidth(!1)+"px"};this.options.get("dropdownAutoWidth")&&(e.minWidth=e.width,e.position="relative",e.width="auto"),this.$dropdown.css(e)},e.prototype._showDropdown=function(e){this.$dropdownContainer.appendTo(this.$dropdownParent),this._positionDropdown(),this._resizeDropdown()},e}),u.define("select2/dropdown/minimumResultsForSearch",[],function(){function e(e,t,n,s){this.minimumResultsForSearch=n.get("minimumResultsForSearch"),this.minimumResultsForSearch<0&&(this.minimumResultsForSearch=1/0),e.call(this,t,n,s)}return e.prototype.showSearch=function(e,t){return!(function e(t){for(var n=0,s=0;s<t.length;s++){var i=t[s];i.children?n+=e(i.children):n++}return n}(t.data.results)<this.minimumResultsForSearch)&&e.call(this,t)},e}),u.define("select2/dropdown/selectOnClose",["../utils"],function(s){function e(){}return e.prototype.bind=function(e,t,n){var s=this;e.call(this,t,n),t.on("close",function(e){s._handleSelectOnClose(e)})},e.prototype._handleSelectOnClose=function(e,t){if(t&&null!=t.originalSelect2Event){var n=t.originalSelect2Event;if("select"===n._type||"unselect"===n._type)return}n=this.getHighlightedResults();n.length<1||(null!=(n=s.GetData(n[0],"data")).element&&n.element.selected||null==n.element&&n.selected||this.trigger("select",{data:n}))},e}),u.define("select2/dropdown/closeOnSelect",[],function(){function e(){}return e.prototype.bind=function(e,t,n){var s=this;e.call(this,t,n),t.on("select",function(e){s._selectTriggered(e)}),t.on("unselect",function(e){s._selectTriggered(e)})},e.prototype._selectTriggered=function(e,t){var n=t.originalEvent;n&&(n.ctrlKey||n.metaKey)||this.trigger("close",{originalEvent:n,originalSelect2Event:t})},e}),u.define("select2/dropdown/dropdownCss",["../utils"],function(n){function e(){}return e.prototype.render=function(e){var t=e.call(this),e=this.options.get("dropdownCssClass")||"";return-1!==e.indexOf(":all:")&&(e=e.replace(":all:",""),n.copyNonInternalCssClasses(t[0],this.$element[0])),t.addClass(e),t},e}),u.define("select2/dropdown/tagsSearchHighlight",["../utils"],function(s){function e(){}return e.prototype.highlightFirstItem=function(e){var t=this.$results.find(".select2-results__option--selectable:not(.select2-results__option--selected)");if(0<t.length){var n=t.first(),t=s.GetData(n[0],"data").element;if(t&&t.getAttribute&&"true"===t.getAttribute("data-select2-tag"))return void n.trigger("mouseenter")}e.call(this)},e}),u.define("select2/i18n/en",[],function(){return{errorLoading:function(){return"The results could not be loaded."},inputTooLong:function(e){var t=e.input.length-e.maximum,e="Please delete "+t+" character";return 1!=t&&(e+="s"),e},inputTooShort:function(e){return"Please enter "+(e.minimum-e.input.length)+" or more characters"},loadingMore:function(){return"Loading more results…"},maximumSelected:function(e){var t="You can only select "+e.maximum+" item";return 1!=e.maximum&&(t+="s"),t},noResults:function(){return"No results found"},searching:function(){return"Searching…"},removeAllItems:function(){return"Remove all items"},removeItem:function(){return"Remove item"},search:function(){return"Search"}}}),u.define("select2/defaults",["jquery","./results","./selection/single","./selection/multiple","./selection/placeholder","./selection/allowClear","./selection/search","./selection/selectionCss","./selection/eventRelay","./utils","./translation","./diacritics","./data/select","./data/array","./data/ajax","./data/tags","./data/tokenizer","./data/minimumInputLength","./data/maximumInputLength","./data/maximumSelectionLength","./dropdown","./dropdown/search","./dropdown/hidePlaceholder","./dropdown/infiniteScroll","./dropdown/attachBody","./dropdown/minimumResultsForSearch","./dropdown/selectOnClose","./dropdown/closeOnSelect","./dropdown/dropdownCss","./dropdown/tagsSearchHighlight","./i18n/en"],function(l,r,o,a,c,u,d,p,h,f,g,t,m,y,v,_,b,$,w,x,A,D,S,E,O,C,L,T,q,I,e){function n(){this.reset()}return n.prototype.apply=function(e){var t;null==(e=l.extend(!0,{},this.defaults,e)).dataAdapter&&(null!=e.ajax?e.dataAdapter=v:null!=e.data?e.dataAdapter=y:e.dataAdapter=m,0<e.minimumInputLength&&(e.dataAdapter=f.Decorate(e.dataAdapter,$)),0<e.maximumInputLength&&(e.dataAdapter=f.Decorate(e.dataAdapter,w)),0<e.maximumSelectionLength&&(e.dataAdapter=f.Decorate(e.dataAdapter,x)),e.tags&&(e.dataAdapter=f.Decorate(e.dataAdapter,_)),null==e.tokenSeparators&&null==e.tokenizer||(e.dataAdapter=f.Decorate(e.dataAdapter,b))),null==e.resultsAdapter&&(e.resultsAdapter=r,null!=e.ajax&&(e.resultsAdapter=f.Decorate(e.resultsAdapter,E)),null!=e.placeholder&&(e.resultsAdapter=f.Decorate(e.resultsAdapter,S)),e.selectOnClose&&(e.resultsAdapter=f.Decorate(e.resultsAdapter,L)),e.tags&&(e.resultsAdapter=f.Decorate(e.resultsAdapter,I))),null==e.dropdownAdapter&&(e.multiple?e.dropdownAdapter=A:(t=f.Decorate(A,D),e.dropdownAdapter=t),0!==e.minimumResultsForSearch&&(e.dropdownAdapter=f.Decorate(e.dropdownAdapter,C)),e.closeOnSelect&&(e.dropdownAdapter=f.Decorate(e.dropdownAdapter,T)),null!=e.dropdownCssClass&&(e.dropdownAdapter=f.Decorate(e.dropdownAdapter,q)),e.dropdownAdapter=f.Decorate(e.dropdownAdapter,O)),null==e.selectionAdapter&&(e.multiple?e.selectionAdapter=a:e.selectionAdapter=o,null!=e.placeholder&&(e.selectionAdapter=f.Decorate(e.selectionAdapter,c)),e.allowClear&&(e.selectionAdapter=f.Decorate(e.selectionAdapter,u)),e.multiple&&(e.selectionAdapter=f.Decorate(e.selectionAdapter,d)),null!=e.selectionCssClass&&(e.selectionAdapter=f.Decorate(e.selectionAdapter,p)),e.selectionAdapter=f.Decorate(e.selectionAdapter,h)),e.language=this._resolveLanguage(e.language),e.language.push("en");for(var n=[],s=0;s<e.language.length;s++){var i=e.language[s];-1===n.indexOf(i)&&n.push(i)}return e.language=n,e.translations=this._processTranslations(e.language,e.debug),e},n.prototype.reset=function(){function a(e){return e.replace(/[^\u0000-\u007E]/g,function(e){return t[e]||e})}this.defaults={amdLanguageBase:"./i18n/",autocomplete:"off",closeOnSelect:!0,debug:!1,dropdownAutoWidth:!1,escapeMarkup:f.escapeMarkup,language:{},matcher:function e(t,n){if(null==t.term||""===t.term.trim())return n;if(n.children&&0<n.children.length){for(var s=l.extend(!0,{},n),i=n.children.length-1;0<=i;i--)null==e(t,n.children[i])&&s.children.splice(i,1);return 0<s.children.length?s:e(t,s)}var r=a(n.text).toUpperCase(),o=a(t.term).toUpperCase();return-1<r.indexOf(o)?n:null},minimumInputLength:0,maximumInputLength:0,maximumSelectionLength:0,minimumResultsForSearch:0,selectOnClose:!1,scrollAfterSelect:!1,sorter:function(e){return e},templateResult:function(e){return e.text},templateSelection:function(e){return e.text},theme:"default",width:"resolve"}},n.prototype.applyFromElement=function(e,t){var n=e.language,s=this.defaults.language,i=t.prop("lang"),t=t.closest("[lang]").prop("lang"),t=Array.prototype.concat.call(this._resolveLanguage(i),this._resolveLanguage(n),this._resolveLanguage(s),this._resolveLanguage(t));return e.language=t,e},n.prototype._resolveLanguage=function(e){if(!e)return[];if(l.isEmptyObject(e))return[];if(l.isPlainObject(e))return[e];for(var t,n=Array.isArray(e)?e:[e],s=[],i=0;i<n.length;i++)s.push(n[i]),"string"==typeof n[i]&&0<n[i].indexOf("-")&&(t=n[i].split("-")[0],s.push(t));return s},n.prototype._processTranslations=function(e,t){for(var n=new g,s=0;s<e.length;s++){var i=new g,r=e[s];if("string"==typeof r)try{i=g.loadPath(r)}catch(e){try{r=this.defaults.amdLanguageBase+r,i=g.loadPath(r)}catch(e){t&&window.console&&console.warn&&console.warn('Select2: The language file for "'+r+'" could not be automatically loaded. A fallback will be used instead.')}}else i=l.isPlainObject(r)?new g(r):r;n.extend(i)}return n},n.prototype.set=function(e,t){var n={};n[l.camelCase(e)]=t;n=f._convertData(n);l.extend(!0,this.defaults,n)},new n}),u.define("select2/options",["jquery","./defaults","./utils"],function(c,n,u){function e(e,t){this.options=e,null!=t&&this.fromElement(t),null!=t&&(this.options=n.applyFromElement(this.options,t)),this.options=n.apply(this.options)}return e.prototype.fromElement=function(e){var t=["select2"];null==this.options.multiple&&(this.options.multiple=e.prop("multiple")),null==this.options.disabled&&(this.options.disabled=e.prop("disabled")),null==this.options.autocomplete&&e.prop("autocomplete")&&(this.options.autocomplete=e.prop("autocomplete")),null==this.options.dir&&(e.prop("dir")?this.options.dir=e.prop("dir"):e.closest("[dir]").prop("dir")?this.options.dir=e.closest("[dir]").prop("dir"):this.options.dir="ltr"),e.prop("disabled",this.options.disabled),e.prop("multiple",this.options.multiple),u.GetData(e[0],"select2Tags")&&(this.options.debug&&window.console&&console.warn&&console.warn('Select2: The `data-select2-tags` attribute has been changed to use the `data-data` and `data-tags="true"` attributes and will be removed in future versions of Select2.'),u.StoreData(e[0],"data",u.GetData(e[0],"select2Tags")),u.StoreData(e[0],"tags",!0)),u.GetData(e[0],"ajaxUrl")&&(this.options.debug&&window.console&&console.warn&&console.warn("Select2: The `data-ajax-url` attribute has been changed to `data-ajax--url` and support for the old attribute will be removed in future versions of Select2."),e.attr("ajax--url",u.GetData(e[0],"ajaxUrl")),u.StoreData(e[0],"ajax-Url",u.GetData(e[0],"ajaxUrl")));var n={};function s(e,t){return t.toUpperCase()}for(var i=0;i<e[0].attributes.length;i++){var r=e[0].attributes[i].name,o="data-";r.substr(0,o.length)==o&&(r=r.substring(o.length),o=u.GetData(e[0],r),n[r.replace(/-([a-z])/g,s)]=o)}c.fn.jquery&&"1."==c.fn.jquery.substr(0,2)&&e[0].dataset&&(n=c.extend(!0,{},e[0].dataset,n));var a,l=c.extend(!0,{},u.GetData(e[0]),n);for(a in l=u._convertData(l))-1<t.indexOf(a)||(c.isPlainObject(this.options[a])?c.extend(this.options[a],l[a]):this.options[a]=l[a]);return this},e.prototype.get=function(e){return this.options[e]},e.prototype.set=function(e,t){this.options[e]=t},e}),u.define("select2/core",["jquery","./options","./utils","./keys"],function(t,i,r,s){var o=function(e,t){null!=r.GetData(e[0],"select2")&&r.GetData(e[0],"select2").destroy(),this.$element=e,this.id=this._generateId(e),t=t||{},this.options=new i(t,e),o.__super__.constructor.call(this);var n=e.attr("tabindex")||0;r.StoreData(e[0],"old-tabindex",n),e.attr("tabindex","-1");t=this.options.get("dataAdapter");this.dataAdapter=new t(e,this.options);n=this.render();this._placeContainer(n);t=this.options.get("selectionAdapter");this.selection=new t(e,this.options),this.$selection=this.selection.render(),this.selection.position(this.$selection,n);t=this.options.get("dropdownAdapter");this.dropdown=new t(e,this.options),this.$dropdown=this.dropdown.render(),this.dropdown.position(this.$dropdown,n);n=this.options.get("resultsAdapter");this.results=new n(e,this.options,this.dataAdapter),this.$results=this.results.render(),this.results.position(this.$results,this.$dropdown);var s=this;this._bindAdapters(),this._registerDomEvents(),this._registerDataEvents(),this._registerSelectionEvents(),this._registerDropdownEvents(),this._registerResultsEvents(),this._registerEvents(),this.dataAdapter.current(function(e){s.trigger("selection:update",{data:e})}),e[0].classList.add("select2-hidden-accessible"),e.attr("aria-hidden","true"),this._syncAttributes(),r.StoreData(e[0],"select2",this),e.data("select2",this)};return r.Extend(o,r.Observable),o.prototype._generateId=function(e){return"select2-"+(null!=e.attr("id")?e.attr("id"):null!=e.attr("name")?e.attr("name")+"-"+r.generateChars(2):r.generateChars(4)).replace(/(:|\.|\[|\]|,)/g,"")},o.prototype._placeContainer=function(e){e.insertAfter(this.$element);var t=this._resolveWidth(this.$element,this.options.get("width"));null!=t&&e.css("width",t)},o.prototype._resolveWidth=function(e,t){var n=/^width:(([-+]?([0-9]*\.)?[0-9]+)(px|em|ex|%|in|cm|mm|pt|pc))/i;if("resolve"==t){var s=this._resolveWidth(e,"style");return null!=s?s:this._resolveWidth(e,"element")}if("element"==t){s=e.outerWidth(!1);return s<=0?"auto":s+"px"}if("style"!=t)return"computedstyle"!=t?t:window.getComputedStyle(e[0]).width;e=e.attr("style");if("string"!=typeof e)return null;for(var i=e.split(";"),r=0,o=i.length;r<o;r+=1){var a=i[r].replace(/\s/g,"").match(n);if(null!==a&&1<=a.length)return a[1]}return null},o.prototype._bindAdapters=function(){this.dataAdapter.bind(this,this.$container),this.selection.bind(this,this.$container),this.dropdown.bind(this,this.$container),this.results.bind(this,this.$container)},o.prototype._registerDomEvents=function(){var t=this;this.$element.on("change.select2",function(){t.dataAdapter.current(function(e){t.trigger("selection:update",{data:e})})}),this.$element.on("focus.select2",function(e){t.trigger("focus",e)}),this._syncA=r.bind(this._syncAttributes,this),this._syncS=r.bind(this._syncSubtree,this),this._observer=new window.MutationObserver(function(e){t._syncA(),t._syncS(e)}),this._observer.observe(this.$element[0],{attributes:!0,childList:!0,subtree:!1})},o.prototype._registerDataEvents=function(){var n=this;this.dataAdapter.on("*",function(e,t){n.trigger(e,t)})},o.prototype._registerSelectionEvents=function(){var n=this,s=["toggle","focus"];this.selection.on("toggle",function(){n.toggleDropdown()}),this.selection.on("focus",function(e){n.focus(e)}),this.selection.on("*",function(e,t){-1===s.indexOf(e)&&n.trigger(e,t)})},o.prototype._registerDropdownEvents=function(){var n=this;this.dropdown.on("*",function(e,t){n.trigger(e,t)})},o.prototype._registerResultsEvents=function(){var n=this;this.results.on("*",function(e,t){n.trigger(e,t)})},o.prototype._registerEvents=function(){var n=this;this.on("open",function(){n.$container[0].classList.add("select2-container--open")}),this.on("close",function(){n.$container[0].classList.remove("select2-container--open")}),this.on("enable",function(){n.$container[0].classList.remove("select2-container--disabled")}),this.on("disable",function(){n.$container[0].classList.add("select2-container--disabled")}),this.on("blur",function(){n.$container[0].classList.remove("select2-container--focus")}),this.on("query",function(t){n.isOpen()||n.trigger("open",{}),this.dataAdapter.query(t,function(e){n.trigger("results:all",{data:e,query:t})})}),this.on("query:append",function(t){this.dataAdapter.query(t,function(e){n.trigger("results:append",{data:e,query:t})})}),this.on("keypress",function(e){var t=e.which;n.isOpen()?t===s.ESC||t===s.UP&&e.altKey?(n.close(e),e.preventDefault()):t===s.ENTER||t===s.TAB?(n.trigger("results:select",{}),e.preventDefault()):t===s.SPACE&&e.ctrlKey?(n.trigger("results:toggle",{}),e.preventDefault()):t===s.UP?(n.trigger("results:previous",{}),e.preventDefault()):t===s.DOWN&&(n.trigger("results:next",{}),e.preventDefault()):(t===s.ENTER||t===s.SPACE||t===s.DOWN&&e.altKey)&&(n.open(),e.preventDefault())})},o.prototype._syncAttributes=function(){this.options.set("disabled",this.$element.prop("disabled")),this.isDisabled()?(this.isOpen()&&this.close(),this.trigger("disable",{})):this.trigger("enable",{})},o.prototype._isChangeMutation=function(e){var t=this;if(e.addedNodes&&0<e.addedNodes.length){for(var n=0;n<e.addedNodes.length;n++)if(e.addedNodes[n].selected)return!0}else{if(e.removedNodes&&0<e.removedNodes.length)return!0;if(Array.isArray(e))return e.some(function(e){return t._isChangeMutation(e)})}return!1},o.prototype._syncSubtree=function(e){var e=this._isChangeMutation(e),t=this;e&&this.dataAdapter.current(function(e){t.trigger("selection:update",{data:e})})},o.prototype.trigger=function(e,t){var n=o.__super__.trigger,s={open:"opening",close:"closing",select:"selecting",unselect:"unselecting",clear:"clearing"};if(void 0===t&&(t={}),e in s){var i=s[e],s={prevented:!1,name:e,args:t};if(n.call(this,i,s),s.prevented)return void(t.prevented=!0)}n.call(this,e,t)},o.prototype.toggleDropdown=function(){this.isDisabled()||(this.isOpen()?this.close():this.open())},o.prototype.open=function(){this.isOpen()||this.isDisabled()||this.trigger("query",{})},o.prototype.close=function(e){this.isOpen()&&this.trigger("close",{originalEvent:e})},o.prototype.isEnabled=function(){return!this.isDisabled()},o.prototype.isDisabled=function(){return this.options.get("disabled")},o.prototype.isOpen=function(){return this.$container[0].classList.contains("select2-container--open")},o.prototype.hasFocus=function(){return this.$container[0].classList.contains("select2-container--focus")},o.prototype.focus=function(e){this.hasFocus()||(this.$container[0].classList.add("select2-container--focus"),this.trigger("focus",{}))},o.prototype.enable=function(e){this.options.get("debug")&&window.console&&console.warn&&console.warn('Select2: The `select2("enable")` method has been deprecated and will be removed in later Select2 versions. Use $element.prop("disabled") instead.');e=!(e=null==e||0===e.length?[!0]:e)[0];this.$element.prop("disabled",e)},o.prototype.data=function(){this.options.get("debug")&&0<arguments.length&&window.console&&console.warn&&console.warn('Select2: Data can no longer be set using `select2("data")`. You should consider setting the value instead using `$element.val()`.');var t=[];return this.dataAdapter.current(function(e){t=e}),t},o.prototype.val=function(e){if(this.options.get("debug")&&window.console&&console.warn&&console.warn('Select2: The `select2("val")` method has been deprecated and will be removed in later Select2 versions. Use $element.val() instead.'),null==e||0===e.length)return this.$element.val();e=e[0];Array.isArray(e)&&(e=e.map(function(e){return e.toString()})),this.$element.val(e).trigger("input").trigger("change")},o.prototype.destroy=function(){r.RemoveData(this.$container[0]),this.$container.remove(),this._observer.disconnect(),this._observer=null,this._syncA=null,this._syncS=null,this.$element.off(".select2"),this.$element.attr("tabindex",r.GetData(this.$element[0],"old-tabindex")),this.$element[0].classList.remove("select2-hidden-accessible"),this.$element.attr("aria-hidden","false"),r.RemoveData(this.$element[0]),this.$element.removeData("select2"),this.dataAdapter.destroy(),this.selection.destroy(),this.dropdown.destroy(),this.results.destroy(),this.dataAdapter=null,this.selection=null,this.dropdown=null,this.results=null},o.prototype.render=function(){var e=t('<span class="select2 select2-container"><span class="selection"></span><span class="dropdown-wrapper" aria-hidden="true"></span></span>');return e.attr("dir",this.options.get("dir")),this.$container=e,this.$container[0].classList.add("select2-container--"+this.options.get("theme")),r.StoreData(e[0],"element",this.$element),e},o}),u.define("jquery-mousewheel",["jquery"],function(e){return e}),u.define("jquery.select2",["jquery","jquery-mousewheel","./select2/core","./select2/defaults","./select2/utils"],function(i,e,r,t,o){var a;return null==i.fn.select2&&(a=["open","close","destroy"],i.fn.select2=function(t){if("object"==typeof(t=t||{}))return this.each(function(){var e=i.extend(!0,{},t);new r(i(this),e)}),this;if("string"!=typeof t)throw new Error("Invalid arguments for Select2: "+t);var n,s=Array.prototype.slice.call(arguments,1);return this.each(function(){var e=o.GetData(this,"select2");null==e&&window.console&&console.error&&console.error("The select2('"+t+"') method was called on an element that is not using Select2."),n=e[t].apply(e,s)}),-1<a.indexOf(t)?this:n}),null==i.fn.select2.defaults&&(i.fn.select2.defaults=t),r}),{define:u.define,require:u.require});function b(e,t){return i.call(e,t)}function l(e,t){var n,s,i,r,o,a,l,c,u,d,p=t&&t.split("/"),h=y.map,f=h&&h["*"]||{};if(e){for(t=(e=e.split("/")).length-1,y.nodeIdCompat&&_.test(e[t])&&(e[t]=e[t].replace(_,"")),"."===e[0].charAt(0)&&p&&(e=p.slice(0,p.length-1).concat(e)),c=0;c<e.length;c++)"."===(d=e[c])?(e.splice(c,1),--c):".."===d&&(0===c||1===c&&".."===e[2]||".."===e[c-1]||0<c&&(e.splice(c-1,2),c-=2));e=e.join("/")}if((p||f)&&h){for(c=(n=e.split("/")).length;0<c;--c){if(s=n.slice(0,c).join("/"),p)for(u=p.length;0<u;--u)if(i=h[p.slice(0,u).join("/")],i=i&&i[s]){r=i,o=c;break}if(r)break;!a&&f&&f[s]&&(a=f[s],l=c)}!r&&a&&(r=a,o=l),r&&(n.splice(0,o,r),e=n.join("/"))}return e}function w(t,n){return function(){var e=a.call(arguments,0);return"string"!=typeof e[0]&&1===e.length&&e.push(null),o.apply(p,e.concat([t,n]))}}function x(e){var t;if(b(m,e)&&(t=m[e],delete m[e],v[e]=!0,r.apply(p,t)),!b(g,e)&&!b(v,e))throw new Error("No "+e);return g[e]}function c(e){var t,n=e?e.indexOf("!"):-1;return-1<n&&(t=e.substring(0,n),e=e.substring(n+1,e.length)),[t,e]}function A(e){return e?c(e):[]}var u=s.require("jquery.select2");return t.fn.select2.amd=s,u});
```
